# Pre-Diagnostic Cognitive and Functional Impairment in Multiple Sporadic Neurodegenerative Diseases

**DOI:** 10.1101/2022.04.05.22273468

**Authors:** Nol Swaddiwudhipong, David J. Whiteside, Frank H. Hezemans, Duncan Street, James B. Rowe, Timothy Rittman

**Affiliations:** Department of Clinical Neurosciences; Cambridge University Hospitals NHS Trust, University of Cambridge, UK; MRC Cognition and Brain Sciences Unit, University of Cambridge, UK

**Author notes:** **Corresponding author:** Dr Timothy Rittman, Department of Clinical Neurosciences, Herchel Smith Building, Robinson Way, Cambridge Biomedical Campus, United Kingdom, CB2 0SZ. **Funding information** This work has been funded by the MRC (SUAG/051 R101400) and Cambridge Centre for Parkinson-plus; and the NIHR Cambridge Biomedical Research Centre (BRC-1215-20014). The views expressed are those of the author(s) and not necessarily those of the NIHR or the Department of Health and Social Care. For the purpose of open access, the authors have applied a CC BY public copyright licence to any Author Accepted Manuscript version arising from this submission.

**Keywords:** Dementia, Neurodegenerative Disease, Sporadic, Pre-diagnostic, Cognition, Physical function, Alzheimer’s Disease, Parkinson’s Disease, Frontotemporal Dementia, Progressive Supranuclear Palsy, Multiple System Atrophy, Dementia with Lewy Bodies, UK Biobank

## Abstract

**INTRODUCTION:** The pathophysiological processes of neurodegenerative diseases begin years before diagnosis. However, pre-diagnostic changes in cognition and physical function are poorly understood, especially in sporadic neurodegenerative disease.

**METHODS:** UK Biobank data was extracted. Cognitive and functional measures in individuals who subsequently developed Alzheimer’s Disease, Parkinson’s Disease, Frontotemporal Dementia, Progressive Supranuclear Palsy, Dementia with Lewy Bodies, or Multiple System Atrophy, were compared against those without neurodegenerative diagnoses. The same measures were regressed against time to diagnosis, after adjusting for the effects of age.

**RESULTS:** There was evidence for pre-diagnostic cognitive impairment and decline with time, particularly in Alzheimer’s. Pre-diagnostic functional impairment and decline was observed in multiple diseases.

**DISCUSSION:** The scale and longitudinal follow-up of UK Biobank participants provides evidence for cognitive and functional decline years before symptoms become obvious in multiple neurodegenerative diseases. Identifying pre-diagnostic functional and cognitive changes could improve selection for preventive and early disease-modifying treatment trials.

**Research in Context:** *Systematic review:* Studies of genetic dementia cohorts provide evidence for pre-diagnostic changes in disease biomarkers and cognitive function in several genetic neurodegenerative diseases. The pre-diagnostic phase of sporadic neurodegenerative disease has been less well-studied. It is unclear whether early functional or cognitive changes are detectable in sporadic neurodegenerative disease.

*Interpretation:* We have established an approach to identify cognitive and functional pre-diagnostic markers of neurodegenerative disease years before diagnosis. We found disease-relevant patterns of pre-diagnostic cognitive and functional impairment, and observed a pre-diagnostic linear decline in a number of cognitive and functional measures.

*Future Directions:* Our approach can form the basis for pre-diagnostic cognitive and functional screening to recruit into trials of disease prevention and disease modifying therapies for neurodegenerative diseases. A screening panel based on cognition and function could be followed by disease-specific biomarkers to further improve risk stratification.

## 1. Introduction

Neurodegenerative diseases are associated with significant health, emotional and economic burden on individuals and society. Disease modifying therapies and effective preventive strategies are lacking [1]. However, treatment trials are typically conducted after symptoms have emerged, which may be too late in the disease process to alter its course [2, 3]. Understanding the earliest, pre-diagnostic, phase in neurodegenerative disease could provide a window of opportunity for more effective preventive and disease modifying treatment trials.

Studies of genetic dementia cohorts suggest that key disease biomarkers change in neurodegenerative diseases many years before symptoms are obvious and a diagnosis is made. In genetic frontotemporal dementia (FTD), structural brain changes are detectable 10 years before symptom onset [4–6], with pre-symptomatic alterations in functional brain network organisation [7] and microRNA expression [8]. In genetic Alzheimer’s disease (AD), cerebrospinal fluid and neuroimaging changes may be seen 15-25 years before symptom onset [9–11].

The pre-diagnostic phase of sporadic neurodegenerative disease is more challenging to assess. There is indirect evidence that amyloid-*β* neuropathology is present several years before symptom onset in sporadic AD and is associated with cognitive decline [12]. There is also evidence for a pre-symptomatic reduction in monoaminergic nuclei MRI signal [13].

These studies suggest early pathological changes, but it remains less certain whether this translates into cognitive change or impaired day-to-day function. There is evidence for pre-diagnostic accelerated forgetting in familial AD mutation carriers [14], whilst apathy and executive dysfunction appear early in those carrying mutations for FTD [5, 15]. However, global cognitive and behavioural function remain near normal if supported by reorganisation of the brain’s functional network [7, 16, 17]. It remains unclear whether similar cognitive changes in sporadic dementias are detectable before symptom onset, and how long before a diagnosis they are identifiable.

The UK Biobank [18] offers a rare opportunity to analyse pre-diagnostic changes across a wide range of sporadic neurodegenerative diseases. It includes over 500,000 individuals aged 40-69 recruited between 2006-2010 from the general population, from whom health-related data were collected during one to four assessment visits. This offers a rich dataset of prospective cognitive and functional data from a large pool of individuals, some of whom have gone on to develop a neurodegenerative disease. As a proof-of-concept, we recently published an analysis of pre-diagnostic data on the cohort of people in the UK Biobank who went on to develop Progressive Supranuclear Palsy [19].

In this study, we present an analysis of the data extracted from the UK Biobank, testing whether cognitive and functional changes are detectable in individuals who later went on to develop a neurodegenerative disease, the majority of which are sporadic. This provides a broad overview of the early manifestations of multiple rare and common neurodegenerative diseases.

## 2. Methods

### 2.1. Data Extraction from UK Biobank

Data extracted from the UK Biobank included participant demographics, diagnoses of neurodegenerative diseases (Table 1), and a set of cognitive and functional measures (Table 2). Where applicable for demographic data, pairwise comparisons were performed between each diagnostic group and controls using linear or multinomial logistic regression, with p-values adjusted for multiple comparison using the Benjamini-Hochberg procedure [20, 21] within each diagnosis.

**Table 1:** Demographics of individuals included from the UK Biobank from each diagnostic group. Participants with a diagnosis at baseline were excluded, so the groups here are those who converted during the study. Where appropriate the mean is shown with standard deviation in parentheses. Where the values differ from controls with an adjusted p-value <0.05, they are marked with an asterisk. Ctrl = Controls, AD = Alzheimer’s Disease, FTD = Frontotemporal Dementia, PD = Parkinson’s Disease, PSP = Progressive Supranuclear Palsy, DLB = Dementia with Lewy Bodies, MSA = Multiple System Atrophy.

|  | Ctrl | AD | FTD | PD | PSP | DLB | MSA |
| --- | --- | --- | --- | --- | --- | --- | --- |
| Number | 493,735 | 2778 | 211 | 2370 | 133 | 40 | 73 |
| Years to diagnosis<br>from baseline | - | 8.3<br>(3.0) | 7.7<br>(3.0) | 7.4<br>(3.2) | 7.6<br>(3.0) | 4.7<br>(2.1) | 6.5<br>(3.3) |
| Age | 56.4<br>(8.1) | 64.7*<br>(4.2) | 61.2*<br>(5.9) | 62.8*<br>(5.4) | 63.6*<br>(5.3) | 65.4*<br>(3.7) | 60.8*<br>(6.3) |
| BMI | 27.4<br>(4.8) | 27.4<br>(4.8) | 27.2<br>(5.0) | 27.8*<br>(4.5) | 29*<br>(4.8) | 27.5<br>(4.4) | 26.6<br>(4.2) |
| Sex |  |  |  |  |  |  |  |
| Male | 44.3% | 45.3% | 49.8% | 60.3%* | 60.9%* | 67.5%* | 39.7% |
| Female | 52.9% | 50.7% | 47.4% | 36.1%* | 35.3%* | 30%* | 58.9% |
| Handedness |  |  |  |  |  |  |  |
| Right | 88.7% | 89.3% | 91.5% | 88.4% | 90.2% | 92.5% | 90.4% |
| Left | 9.3% | 8.2% | 6.2% | 9.3% | 6% | 7.5% | 6.8% |
| Ambidextrous | 1.7% | 2.1% | 2.4% | 1.9% | 3% | 0% | 2.7% |
| Ethnicity |  |  |  |  |  |  |  |
| White | 93.9% | 95.5%* | 96.7% | 95.8%* | 96.2%* | 100%* | 93.2%* |
| Afro-Caribbean | 1.6% | 1.4% | 0.9% | 0.8%* | 0% | 0%* | 2.7% |
| Asian | 2.3% | 1.4%* | 0.9% | 1.8% | 1.5% | 0%* | 1.4% |
| Mixed | 0.5% | 0.3% | 0% | 0.3% | 0%* | 0% | 2.7% |
| Others | 0.9% | 0.4%* | 0.5% | 0.5%* | 0%* | 0%* | 0%* |
| Smoking Status |  |  |  |  |  |  |  |
| Never | 40% | 37%* | 34.1% | 39.3%* | 34.6%* | 32.5% | 46.6% |
| Previous | 34.3% | 41.9%* | 40.3% | 39.9%* | 48.1%* | 47.5% | 27.4% |
| Current | 10.6% | 9% | 11.4% | 6.7%* | 8.3% | 2.5% | 8.2% |

**Table 2:**
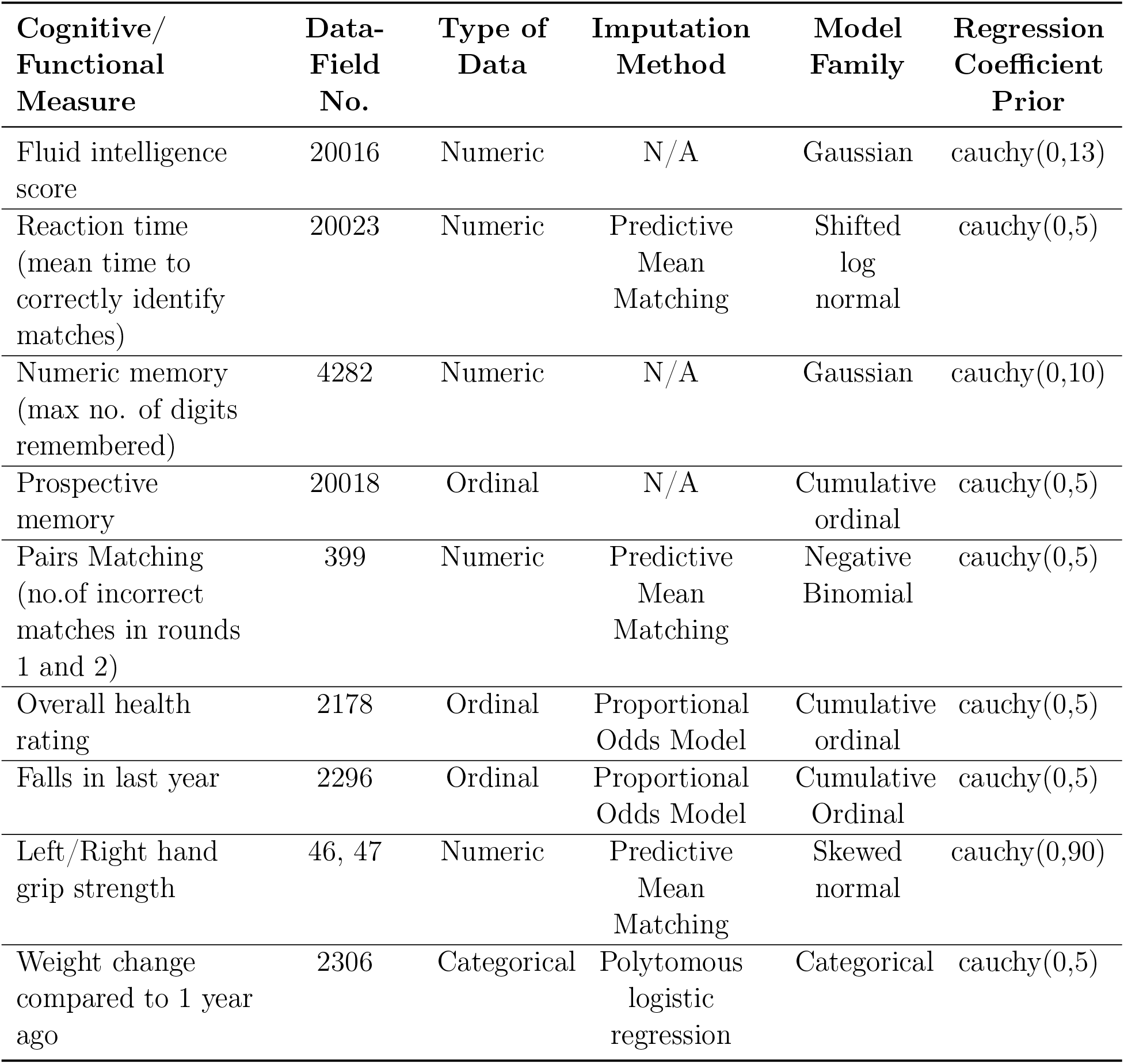
The cognitive and functional outcome measures used in this study and information relevant to the statistical analysis. Full descriptions are available by keying in the corresponding data-field number at biobank.ndph.ox.ac.uk/showcase/search.cgi. Where missing data was imputed we report the imputation method used. We also report the *BRMS* family used to specify the Bayesian model, and the prior specified for the regression coefficient.

| Cognitive/<br>Functional<br>Measure | Data-Field<br>No. | Type of<br>Data | Imputation<br>Method | Model<br>Family | Regression<br>Coefficient<br>Prior |
| --- | --- | --- | --- | --- | --- |
| Fluid intelligence<br>score | 20016 | Numeric | N/A | Gaussian | cauchy(0,13) |
| Reaction time<br>(mean time to<br>correctly identify<br>matches) | 20023 | Numeric | Predictive<br>Mean<br>Matching | Shifted<br>log<br>normal | cauchy(0,5) |
| Numeric memory<br>(max no. of digits<br>remembered) | 4282 | Numeric | N/A | Gaussian | cauchy(0,10) |
| Prospective<br>memory | 20018 | Ordinal | N/A | Cumulative<br>ordinal | cauchy(0,5) |
| Pairs Matching<br>(no.of incorrect<br>matches in rounds<br>1 and 2) | 399 | Numeric | Predictive<br>Mean<br>Matching | Negative<br>Binomial | cauchy(0,5) |
| Overall health<br>rating | 2178 | Ordinal | Proportional<br>Odds Model | Cumulative<br>ordinal | cauchy(0,5) |
| Falls in last year | 2296 | Ordinal | Proportional<br>Odds Model | Cumulative<br>Ordinal | cauchy(0,5) |
| Left/Right hand<br>grip strength | 46, 47 | Numeric | Predictive<br>Mean<br>Matching | Skewed<br>normal | cauchy(0,90) |
| Weight change<br>compared to 1 year<br>ago | 2306 | Categorical | Polytomous<br>logistic<br>regression | Categorical | cauchy(0,5) |

#### 2.1.1. Extraction of Diagnosis

The diagnoses and dates of diagnosis were compiled from hospital inpatient data, primary care data, death certificate data, and self-reported diagnoses. This data was extracted on 23 May 2021. Primary care data were available for approximately 45% of the UK Biobank cohort and covered up to the year 2017. Codes associated with the following diagnoses were searched for: Alzheimer’s Disease (AD), Frontotemporal Dementia (FTD), Parkinson’s Disease (PD), Progressive Supranuclear Palsy (PSP), Dementia with Lewy Bodies (DLB) and Multiple System Atrophy (MSA). Codes associated non-specifically with neurodegenerative disease or other named neurodegenerative diseases affecting cognition were also searched for and labelled as ‘Others’. The most recent diagnosis was used. Where multiple diagnoses were present on the most recent date, the rarer diagnosis was used as the most likely diagnosis (for example, it is common for people with PSP to first receive a diagnosis of PD). Where ambiguity remained the label “Multiple” was used. The earliest date the diagnosis was recorded was used as a proxy for the actual date of diagnosis.

To identify pre-diagnostic individuals only, any individuals with a diagnosis date pre-dating the baseline assessment visit were excluded. Individuals with a self-reported diagnosis of neurodegenerative disease at any time but without a formal diagnosis, and those whose diagnoses were labelled as “Multiple” or “Others” were excluded. The remaining individuals without a labelled diagnosis during follow-up were designated as controls.

### 2.2. Analysis of Baseline Assessment Data

A set of cognitive and functional outcome measures (Table 2) recorded at baseline assessment performed between 2006-2010, were compared between pre-diagnostic individuals and controls using Bayesian regression analysis.

#### 2.2.1. Multiple Imputation

We used multiple imputation, splitting the data into five sub-datasets to account for the 5% of cases with incomplete data among the imputed categories. [22, 23].

Imputation was carried out using the *mice* package in R (version 4.0.3) [24]. “Prospective memory”, “fluid intelligence score”, “numeric memory” and “smoking pack-years” were not imputed and excluded as predictors for imputation as a large proportion of data was missing. All other data fields including demographic data were used as predictor variables and imputed according to the default method for the respective data type as defined in *mice* (Table 2).

#### 2.2.2. Bayesian Regression Modelling

We chose Bayesian regression modelling to assess differences between diagnostic groups given the marked differences in group sizes, the ability to accept or reject the null and model hypothesis based on data precision (cf. ‘power’ for classical inference), and the ability to assess difference between groups based on effect size.

We used the *brms* package in R [25, 26]. Each cognitive or functional outcome measure was fitted to a model with diagnosis category and age as predictors, and also to a null model with age as the sole predictor. Handedness was included as an additional predictor when analysing hand grip strength. Model families were selected based on the characteristics of the data, and weakly informative Cauchy priors centered at zero were used for the regression coefficients (see Table 2). For each outcome measure, the regression model was initially fit separately to each of the five imputed sub-datasets. The posterior draws from the resulting sub-models were then aggregated to obtain a combined model with 50,000 post-warm-up iterations.

#### 2.2.3. Model Validation and Interpretation

All models converged with no divergent transitions or other diagnostic warnings. Diagnostic traceplots showed good mixing of sampling chains. 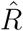 values were all approximately 1.00, and less than 1.05. Graphical posterior predictive checks demonstrated that simulated data drawn from each model’s posterior predictive distributions agreed well with the observed data across the diagnostic categories analysed (see supplementary material).

To assess the evidence for group differences, we obtained the 95% Credible Interval (CI) of the posterior distribution for the regression coefficient of each diagnostic group relative to controls, and compared it to a pre-defined Region Of Practical Equivalence (ROPE). For numerical data, the ROPE was defined as the values ranging between *±*0.1 of the standard deviation around the control mean, which has been suggested as a default [27]. Where logistic regression was used the ROPE was defined as *±*0.18, which is suggested as an equivalent default [28]. For pairs matching data, which was modelled using a negative binomial distribution, the ROPE was defined as a multiplicative effect 0.9-1.1 on the number of incorrect matches. It has been suggested that if the CI falls entirely outside the ROPE, there is strong evidence to reject the null hypothesis; if the CI falls entirely within the ROPE there is strong evidence for accepting the null hypothesis [27].

### 2.3. Regression Analysis Over Time Prior to Diagnosis

To look for linear changes in the years prior to diagnosis, we used classical linear regression. This analysis included a set of cognitive and functional measures (table 2) recorded during baseline assessment and any subsequent visits pre-dating diagnosis.

For numerical data, we first regressed out the data on age, then within each diagnosis regressed the residuals against years to diagnosis. In cases where more than five individuals had multiple data points, we used a linear mixed effects model with the individual as a random effect (using the *lmer* function). Otherwise, simple linear regression was used.

For categorical and ordinal data, we were unable to generate a valid random effects model which took into account individual variation due to the paucity of data points repeated from the same individual. Ordinal logistic regression was performed using the *polr* function from the *MASS* library. Data were first regressed on age, then within each diagnosis data were regressed against years to diagnosis, with age multiplied by the coefficient determined from the first regression model as an offset term. P-values were approximated by comparing the generated t-values against the standard normal distribution.

Multinomial logistic regression for categorical data was performed using the *multinom* function from the *nnet* library. Data were first regressed on age, then within each diagnosis data were regressed against years to diagnosis, with age multiplied by the coefficient determined from the first regression model as an offset term. P-values were calculated using two-tailed Wald z-tests.

P-values were grouped by diagnosis and further separated into those that applied to functional and cognitive measures. Within each of these twelve groups, the p-values were adjusted using the Benjamini-Hochberg method [20, 21]. We report both uncorrected and adjusted p-values.

### 2.4. Code Availability

Search queries and processing scripts are available on Gitlab gitlab.developers.cam.ac. uk/ns651/neurodegeneration.

## 3. Results

### 3.1. Baseline cognition in pre-diagnostic neurodegenerative disease

We assessed whether people who developed a range of neurodegenerative diagnoses demonstrated reduced cognitive function at their baseline assessment. The time between baseline assessment and diagnosis varied between 4.7 years for DLB to 8.3 years for AD (see Table 1). There was strong evidence of decreased fluid intelligence in the pre-AD (raw score difference = -0.96, 95% Credible Interval -0.80 to -1.13), FTD (−1.12, CI -0.53 to -1.71) and PSP groups (−1.17, CI -0.56 to -1.78); in these groups the credible interval lay outside the ROPE (Fig 1A). There was weaker evidence of a difference in DLB (−1.87, CI -3.75 to 0.00) where the mean lay outside the ROPE, but with an overlapping credible interval. There was strong evidence *against* reduced fluid intelligence in PD (−0.13, CI -0.29 to 0.03) and MSA (−0.06, CI -0.88 to 0.75), with the mean and majority of the credible interval overlapping the ROPE in both cases.

**Figure 1:**
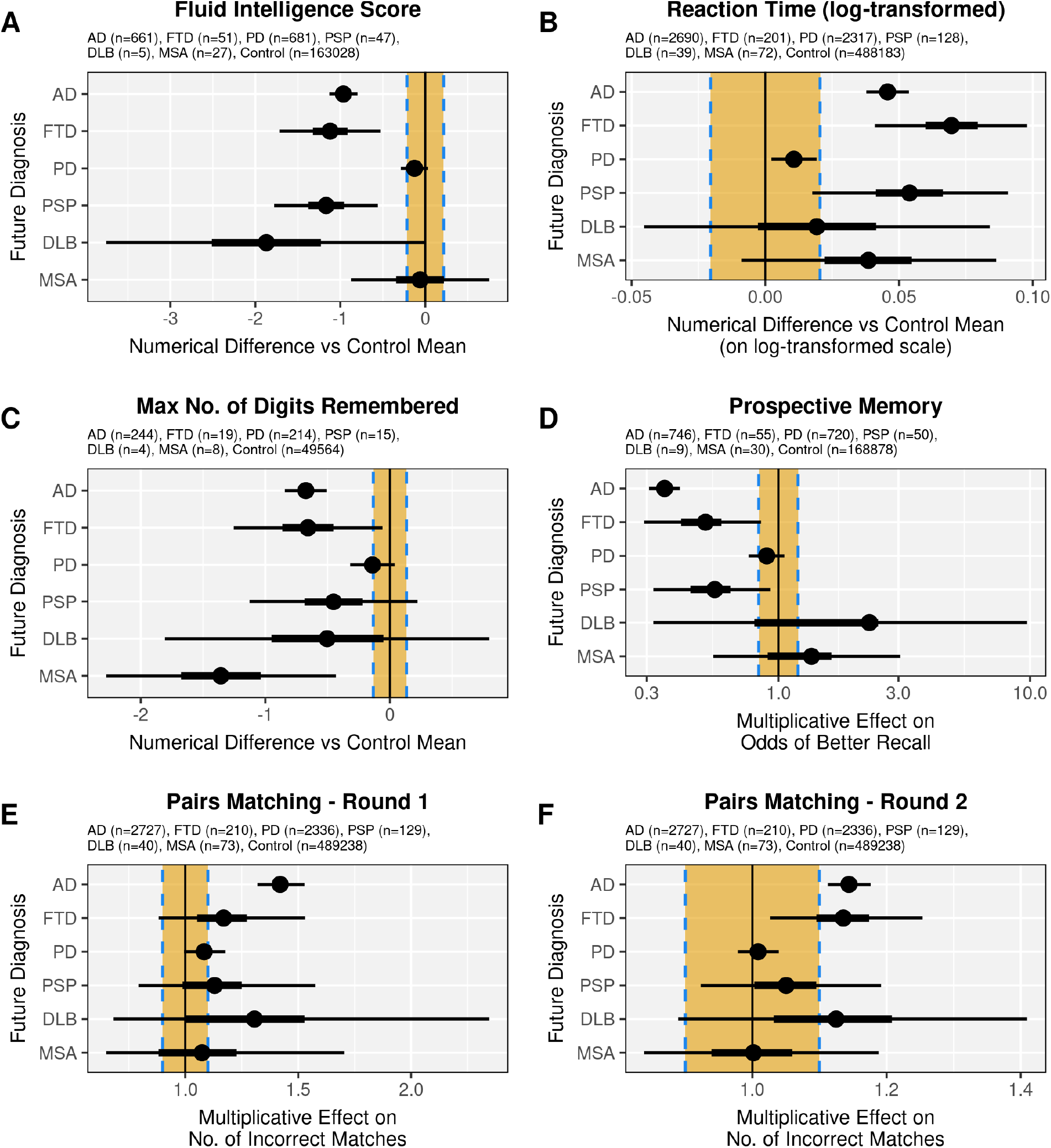
Baseline cognitive measures in UK Biobank participants who go on to develop neurodegenerative disease. The Bayesian posterior probability distributions of the difference relative to controls are plotted, with the mean the shaded circle, 50% credible interval the thicker black line and 95% credible interval the thinner black line. The yellow rectangle represents the region of practical equivalence (ROPE), with a vertical black line denoting the point of zero difference from the control mean or odds depending on the data type, as indicated on the x-axis. Sample sizes shown indicate the number of available raw data points prior to imputation.

There was strong evidence of slower reaction time in pre-AD (difference on log-transformed scale = 0.046, CI 0.038 to 0.054) and FTD individuals (0.070, CI 0.041 to 0.098) (Fig 1B). There was moderate evidence for slower reaction time in PSP (0.054, CI 0.018 to 0.091) and MSA (0.039, CI -0.009 to 0.086). There was strong evidence of *no difference* in reaction time in PD (0.011, CI 0.002 to 0.019), and indeterminate evidence in DLB (0.019, CI -0.045 to 0.084).

Poorer numeric memory was observed in pre-AD individuals (difference in digits remembered = -0.67, CI -0.84 to -0.51) and MSA (−1.36, CI -2.28 to -0.43) (Fig 1C), with some evidence of a difference in FTD (−0.66, CI -1.25 to -0.06). The evidence in PD (−0.14, CI -0.32 to 0.04), PSP (−0.45, CI -1.13 to 0.22) and DLB (−0.50, CI -1.81 to 0.80) was indeterminate.

Poorer prospective memory was observed in pre-AD individuals (multiplicative effect on odds of better recall = 0.35, CI 0.31 to 0.41) (Fig 1D). There was weaker evidence to support differences in FTD (0.51, CI 0.29 to 0.85) and PSP (0.56, CI 0.32 to 0.93). There was moderate evidence that prospective memory was *not impaired* in pre-PD individuals (0.90, CI 0.76 to 1.06), and indeterminate evidence in the DLB (2.30, CI 0.32 to 9.74) and MSA (1.35, CI 0.55 to 3.04) groups.

An increase in incorrectly matched pairs was observed in pre-AD individuals in rounds 1 (multiplicative effect = 1.42, CI 1.32 to 1.53) (Fig 1E) and 2 (1.14, CI 1.11 to 1.18) (Fig 1F). In FTD the evidence was indeterminate for round 1 (1.17, CI 0.88 to 1.53) but there was some evidence of worse performance in round 2 (1.14, CI 1.03 to 1.25). In PD, there was weak evidence in round 1 (1.08, CI 0.99 to 1.18) and strong evidence in round 2 (1.01, CI 0.98 to 1.04) that there was *no difference* in performance. In PSP, the evidence was indeterminate for round 1 (1.13, CI 0.79 to 1.58) but there was some evidence in round 2 that there was *no difference* in performance (1.05, CI 0.92 to 1.19). The evidence was indeterminate in DLB (round 1: 1.31, CI 0.68 to 2.35; round 2: 1.12, CI 0.89 to 1.41) and MSA (round 1: 1.07, CI 0.65 to 1.70; round 2: 1.00, CI 0.84 to 1.19).

### 3.2. Functional impairment at baseline in pre-diagnostic idiopathic neurodegeneration

We went on to assess whether there was evidence of early impaired day-to-day function. Worse overall health was reported relative to controls in pre-AD (multiplicative effect on odds of better rating = 0.64, CI 0.59 to 0.69), FTD (0.65, CI 0.49 to 0.84), PD (0.71, CI 0.66 to 0.77) and PSP individuals (0.57, CI = 0.41 to 0.77) (Fig 2A). The evidence was indeterminate for DLB (0.83, CI 0.45 to 1.42) and MSA (0.75, CI 0.46 to 1.13).

**Figure 2:**
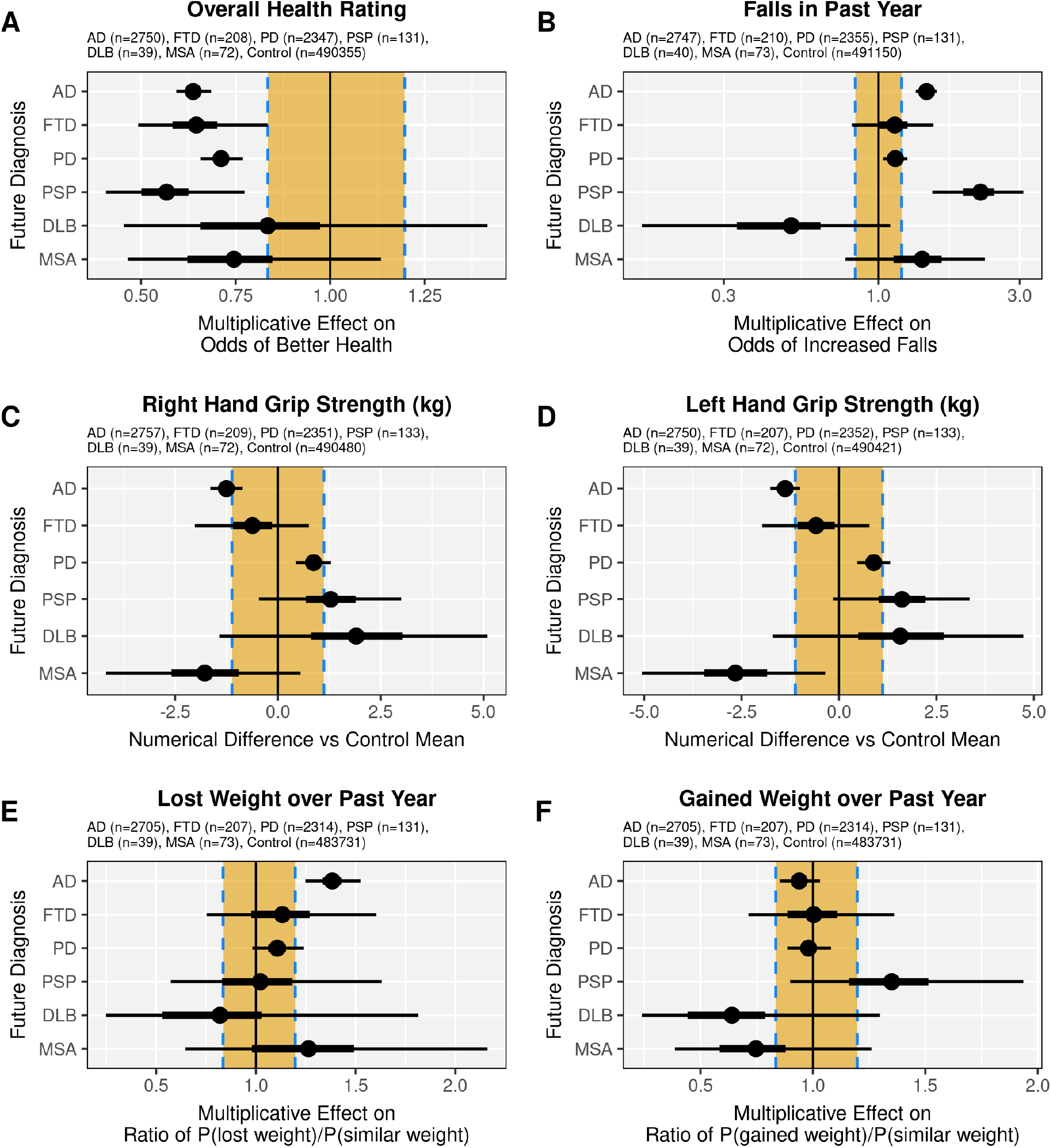
Physical measures in pre-diagnosis individuals. Posterior probability distributions of the difference relative to controls are plotted, with the mean the shaded circle, 50% credible interval the thicker black line, and 95% credible interval the thinner black line. The yellow rectangle represents the region of practical equivalence (ROPE), with a vertical black line denoting the point of zero difference from the control mean or odds depending on the data type, as indicated on the x-axis. Sample sizes shown indicate the number of available raw data points prior to imputation.

An increased number of falls was observed in pre-AD and (multiplicative effect on odds of more falls = 1.45, CI 1.34 to 1.58) and PSP individuals (2.21, CI 1.52 to 3.09) (Fig 2B). There was some evidence to suggest *no difference* in the risk of falls for FTD (1.14, CI 0.81 to 1.53) and PD (1.14, CI 1.04 to 1.25). The evidence was indeterminate for DLB (0.51, CI 0.16 to 1.10) and MSA (1.40, CI 0.77 to 2.29).

There was weak evidence for decreased grip strength in pre-AD (right hand -1.25kg, CI -1.64 to -0.87; left hand -1.38kg, CI -1.76 to -1.00) and MSA individuals (right -1.78kg, CI -4.18 to 0.55; left -2.66kg, CI -5.06 to -0.35) (Fig 2C,D). There was weak evidence to suggest *no difference* in grip strength in FTD (right -0.62kg, CI -2.02 to 0.75; left -0.59kg, CI -1.97 to 0.78) and PD (right 0.87kg, CI 0.44 to 1.29; left 0.89kg, CI 0.47 to 1.31). The evidence was indeterminate for PSP (right 1.28kg, CI -0.46 to 3.00; left 1.61kg, CI -0.16 to 3.35) and DLB (right 1.90kg, CI -1.43 to 5.10; left 1.57kg, CI -1.70 to 4.73).

Differences in weight change over the past year are plotted as the multiplicative effect of each diagnosis on the ratios 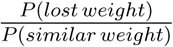 and 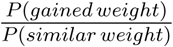. Pre-AD individuals exhibit an increased tendency towards weight loss over the past year (multiplicative effect of 1.38, CI 1.25 to 1.53) (Fig 2E), but no difference in tendency towards weight gain (0.94, CI 0.85 to 1.03) (Fig 2F). In FTD, there was weak evidence to suggest no difference in the tendency towards weight loss (1.13, CI 0.75 to 1.60) or weight gain (1.00, CI 0.71 to 1.36). In PD, there was weak evidence for no difference in weight loss (1.11, CI 0.98 to 1.24) and strong evidence for no difference in weight gain (0.98, CI 0.89 to 1.08). There was some evidence for no difference in weight loss in PSP (1.02, CI 0.57 to 1.63), and indeterminate evidence for weight gain (1.35, CI 0.90 to 1.94). There evidence was indeterminate for DLB (weight loss 0.82, CI 0.25 to 1.81) (weight gain 0.64, CI 0.24 to 1.30) and MSA (weight loss 1.26, CI 0.64 to 2.16) (weight gain 0.75, CI 0.39 to 1.26).

### 3.3. Progressive cognitive and functional decline leading up to diagnosis

A decline was observed for several cognitive and functional measures in the time leading up to diagnosis (Table 3).

**Table 3:** Data from regression of cognitive and functional measures against years to diagnosis, after adjusting for the effect of age. “Prospective memory” and “overall health rating” were analysed using ordinal regression, and log-odds of better outcomes were regressed. “Falls in past year” was analysed with ordinal regression, and log-odds of a greater number of falls were regressed. Weight loss was analysed as the log of the ratio Probability(weight loss)/Probability(weight unchanged), and weight gain was similarly analysed. n = number of data points, Coef = Regression coefficients, SE = standard error, Adj P = P-values adjusted using the Benjamini-Hochberg procedure, AD = Alzheimer’s Disease, FTD = Frontotemporal Dementia, PD = Parkinson’s Disease, PSP = Progressive Supranuclear Palsy, DLB = Dementia with Lewy Bodies, MSA = Multiple System Atrophy.

| Diag | Measure | n | Coef | SE | P-Value | Adj P |
| --- | --- | --- | --- | --- | --- | --- |
| **AD** | Fluid Intelligence Score | 729 | -0.036 | 0.0086 | 2.8e-05 | 5.6e-05 |
|  | Reaction Time (ms) | 2766 | 2.9 | 0.45 | 1.8e-10 | 1.1e-09 |
|  | Max No. of Digits Remembered | 246 | -0.00056 | 0.015 | 0.97 | 0.97 |
|  | Prospective Memory | 824 | -0.1 | 0.024 | 1.6e-05 | 4.7e-05 |
|  | Pairs Matching Round 1 Errors | 2742 | 0.0056 | 0.0069 | 0.42 | 0.51 |
|  | Pairs Matching Round 2 Errors | 2791 | 0.056 | 0.022 | 0.0093 | 0.014 |
|  | Overall Health Rating | 2833 | -0.014 | 0.012 | 0.24 | 0.48 |
|  | Falls in Past Year | 2830 | 0.012 | 0.013 | 0.36 | 0.51 |
|  | Right Hand Grip Strength (kg) | 2844 | -0.044 | 0.016 | 0.0069 | 0.021 |
|  | Left Hand Grip Strength (kg) | 2837 | -0.045 | 0.015 | 0.0032 | 0.019 |
|  | Weight Loss over Past Year | 2789 | 0.013 | 0.016 | 0.42 | 0.51 |
|  | Weight Gain Over Past Year | 2789 | 0.0075 | 0.015 | 0.63 | 0.63 |
| **FTD** | Fluid Intelligence Score | 56 | -0.0079 | 0.035 | 0.82 | 0.82 |
|  | Reaction Time (ms) | 206 | 1 | 1.6 | 0.53 | 0.64 |
|  | Max No. of Digits Remembered | 21 | 0.056 | 0.049 | 0.27 | 0.64 |
|  | Prospective Memory | 60 | -0.073 | 0.083 | 0.38 | 0.64 |
|  | Pairs Matching Round 1 Errors | 213 | -0.019 | 0.019 | 0.32 | 0.64 |
|  | Pairs Matching Round 2 Errors | 212 | 0.052 | 0.07 | 0.45 | 0.64 |
|  | Overall Health Rating | 213 | 0.0099 | 0.043 | 0.82 | 0.92 |
|  | Falls in Past Year | 215 | 0.018 | 0.053 | 0.74 | 0.92 |
|  | Right Hand Grip Strength (kg) | 214 | -0.059 | 0.062 | 0.34 | 0.92 |
|  | Left Hand Grip Strength (kg) | 210 | -0.029 | 0.058 | 0.61 | 0.92 |
|  | Weight Loss over Past Year | 212 | -0.14 | 0.07 | 0.04 | 0.24 |
|  | Weight Gain Over Past Year | 212 | -0.0054 | 0.054 | 0.92 | 0.92 |
| Continued on next page |  |  |  |  |  |  |

Table 3 Continued from previous page
| **Diag** | **Measure** | **n** | **Coef** | **SE** | **P-Value** | **Adj P** |
| --- | --- | --- | --- | --- | --- | --- |
| **PD** | Fluid Intelligence Score | 754 | -0.003 | 0.0085 | 0.73 | 0.74 |
|  | Reaction Time (ms) | 2395 | 0.52 | 0.38 | 0.17 | 0.52 |
|  | Max No. of Digits Remembered | 220 | -0.022 | 0.012 | 0.066 | 0.4 |
|  | Prospective Memory | 801 | -0.02 | 0.026 | 0.44 | 0.74 |
|  | Pairs Matching Round 1 Errors | 2359 | -0.0023 | 0.0057 | 0.7 | 0.74 |
|  | Pairs Matching Round 2 Errors | 2395 | -0.0059 | 0.018 | 0.74 | 0.74 |
|  | Overall Health Rating | 2432 | -0.024 | 0.012 | 0.046 | 0.068 |
|  | Falls in Past Year | 2439 | 0.03 | 0.015 | 0.043 | 0.068 |
|  | Right Hand Grip Strength (kg) | 2432 | -0.089 | 0.017 | 3.2e-07 | 9.7e-07 |
|  | Left Hand Grip Strength (kg) | 2433 | -0.086 | 0.016 | 1.1e-07 | 6.4e-07 |
|  | Weight Loss over Past Year | 2395 | 0.019 | 0.018 | 0.27 | 0.33 |
|  | Weight Gain Over Past Year | 2395 | 0.0058 | 0.015 | 0.7 | 0.7 |
| **PSP** | Fluid Intelligence Score | 49 | -0.039 | 0.033 | 0.25 | 0.25 |
|  | Reaction Time (ms) | 132 | 7.4 | 1.9 | 0.00018 | 0.0011 |
|  | Max No. of Digits Remembered | 16 | -0.085 | 0.048 | 0.1 | 0.2 |
|  | Prospective Memory | 54 | -0.14 | 0.1 | 0.16 | 0.24 |
|  | Pairs Matching Round 1 Errors | 131 | 0.032 | 0.027 | 0.25 | 0.25 |
|  | Pairs Matching Round 2 Errors | 131 | 0.21 | 0.09 | 0.019 | 0.058 |
|  | Overall Health Rating | 135 | -0.16 | 0.06 | 0.0089 | 0.027 |
|  | Falls in Past Year | 135 | 0.17 | 0.062 | 0.0063 | 0.027 |
|  | Right Hand Grip Strength (kg) | 137 | 0.037 | 0.08 | 0.64 | 0.94 |
|  | Left Hand Grip Strength (kg) | 137 | -0.02 | 0.074 | 0.78 | 0.94 |
|  | Weight Loss over Past Year | 135 | -0.0069 | 0.092 | 0.94 | 0.94 |
|  | Weight Gain Over Past Year | 135 | 0.14 | 0.065 | 0.034 | 0.069 |
| Continued on next page |  |  |  |  |  |  |

Table 3: Data from regression of cognitive and functional measures against years to diagnosis, after adjusting for the effect of age.
| Diag | Measure | n | Coef | SE | P-Value | Adj P |
| --- | --- | --- | --- | --- | --- | --- |
| DLB | Fluid Intelligence Score | 5 | -0.13 | 0.15 | 0.47 | 0.8 |
|  | Reaction Time (ms) | 39 | -5.6 | 5.9 | 0.35 | 0.8 |
|  | Max No. of Digits Remembered | 4 | -0.008 | 0.014 | 0.64 | 0.8 |
|  | Prospective Memory | 9 | -0.15 | 0.35 | 0.67 | 0.8 |
|  | Pairs Matching Round 1 Errors | 40 | 0.0053 | 0.093 | 0.95 | 0.95 |
|  | Pairs Matching Round 2 Errors | 40 | -0.2 | 0.25 | 0.42 | 0.8 |
|  | Overall Health Rating | 39 | 0.098 | 0.19 | 0.61 | 0.91 |
|  | Falls in Past Year | 40 | -0.16 | 0.26 | 0.54 | 0.91 |
|  | Right Hand Grip Strength (kg) | 39 | 0.0077 | 0.18 | 0.97 | 0.97 |
|  | Left Hand Grip Strength (kg) | 39 | -0.02 | 0.16 | 0.9 | 0.97 |
|  | Weight Loss over Past Year | 39 | 0.5 | 0.26 | 0.059 | 0.36 |
|  | Weight Gain Over Past Year | 39 | -0.24 | 0.25 | 0.34 | 0.91 |
| MSA | Fluid Intelligence Score | 29 | -0.042 | 0.037 | 0.26 | 0.59 |
|  | Reaction Time (ms) | 74 | 2.1 | 1.8 | 0.23 | 0.59 |
|  | Max No. of Digits Remembered | 8 | -0.07 | 0.099 | 0.51 | 0.76 |
|  | Prospective Memory | 32 | 0.049 | 0.14 | 0.73 | 0.88 |
|  | Pairs Matching Round 1 Errors | 74 | -0.00033 | 0.026 | 0.99 | 0.99 |
|  | Pairs Matching Round 2 Errors | 74 | 0.096 | 0.091 | 0.29 | 0.59 |
|  | Overall Health Rating | 74 | -0.28 | 0.077 | 0.00028 | 0.0017 |
|  | Falls in Past Year | 75 | 0.24 | 0.091 | 0.0096 | 0.029 |
|  | Right Hand Grip Strength (kg) | 74 | -0.17 | 0.089 | 0.061 | 0.091 |
|  | Left Hand Grip Strength (kg) | 74 | -0.11 | 0.084 | 0.21 | 0.25 |
|  | Weight Loss over Past Year | 75 | 0.2 | 0.099 | 0.046 | 0.091 |
|  | Weight Gain Over Past Year | 75 | 0.071 | 0.093 | 0.44 | 0.44 |

Pre-AD individuals demonstrated worsening fluid intelligence (−0.036/year, p = 5.6 *×* 10^−5^) (Fig 3A), reaction time (2.9ms/year, p = 1.1 *×* 10^−9^) (Fig 3B), prospective memory (x1.1/year odds of worse recall, p = 4.7 *×* 10^−5^) (Fig 3D) and pairs matching results (0.056 incorrect matches/year in round 2, p = 0.014) (Fig 3E). Reaction time also worsened (7.4ms/year, p = 0.0011) in pre-PSP individuals (Fig 3C).

**Figure 3:**
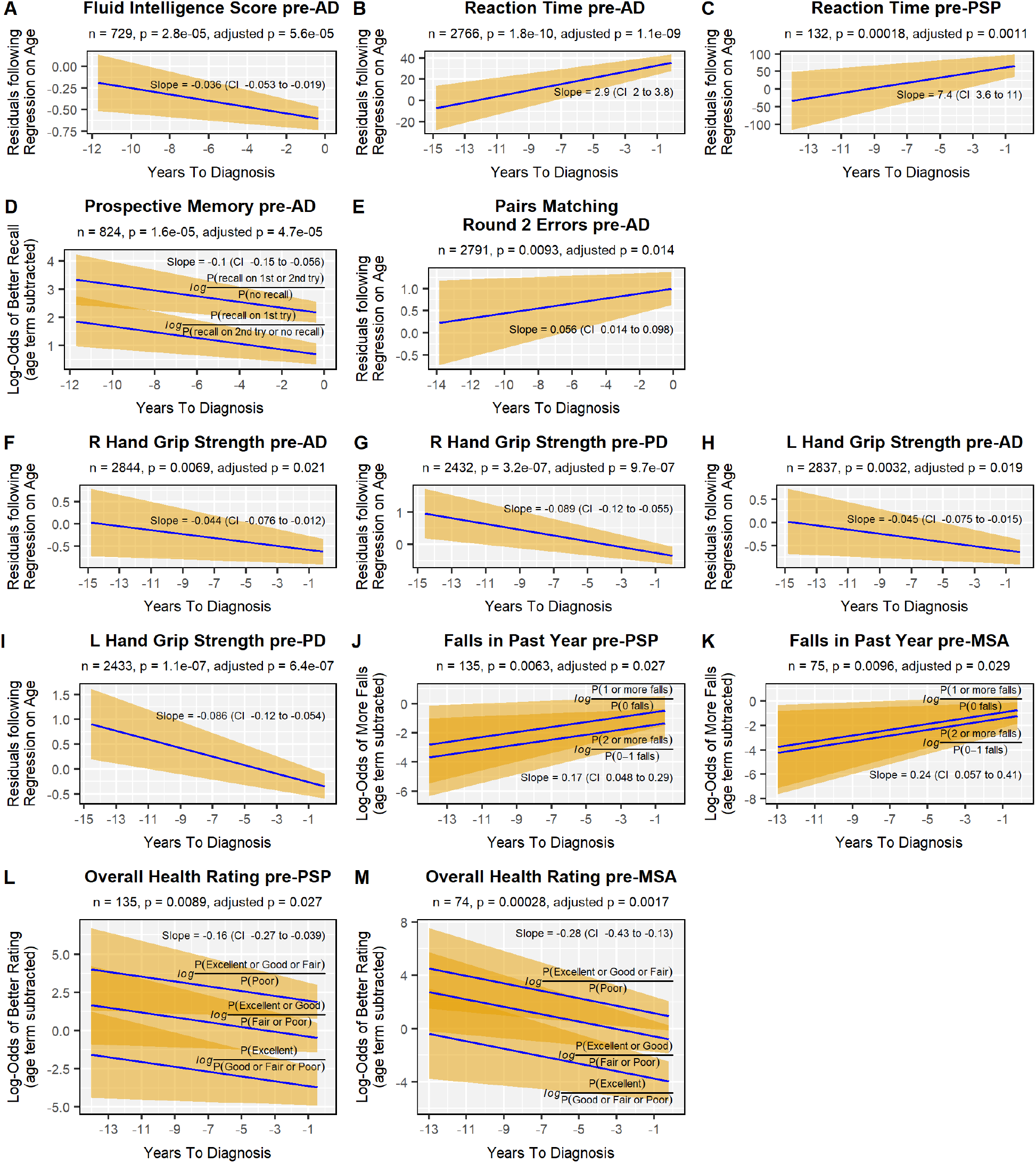
Cognitive and functional measures regressed on years to diagnosis after removing the effect of age. Blue lines indicate the estimated regression coefficient, the yellow shaded area represents the 95% confidence interval. Adjusted p-values were calculated using the Benjamini-Hochberg method. n = number of data points, CI = 95% confidence interval, AD = Alzheimer’s Disease, PD = Parkinson’s Disease, PSP = Progressive Supranuclear Palsy, MSA = Multiple System Atrophy.

Pre-PD individuals exhibited weakening right (−0.089kg/year, p = 9.7 *×* 10^−7^) and left (−0.086kg/year, p = 6.4 *×* 10^−7^) hand grip strength (Fig 3G,I) in the years leading up to diagnosis. Pre-PSP individuals exhibited a decline in overall health self-rating (x1.2/year odds of worse rating, p = 0.027) (Fig 3L), and an increased falls risk (x1.2/year odds of increased falls, p = 0.027) (Fig 3J). Pre-AD individuals demonstrated a significant decrease over time in right (−0.044kg/year, p = 0.021) and left (−0.045kg/year, p = 0.019) hand grip strength (Fig 3F,H). Pre-MSA individuals demonstrated a worsening overall health self-rating (x1.3/year odds of worse rating, p = 0.0017) (Fig 3M) and increased falls risk (x1.3/year odds of increased falls, p = 0.029) (Fig 3K).

## 4. Discussion

We demonstrate cognitive and functional antecedents of several idiopathic neurodegenerative syndromes in the years prior to diagnosis. In line with findings of pre-symptomatic cognitive decline in familial mutation carriers of AD and FTD [5, 14], these changes were identified at a baseline assessment averaging 5-9 years before diagnosis, extending the ‘symptomatic’ phase of sporadic neurodegenerative disease earlier than previously supposed. The pre-diagnostic linear decline in a number of measures supports our supposition that these changes represent early progressive neurodegeneration rather than a low cognitive or functional baseline.

We found patterns of cognitive and functional change that were disease-specific. Extensive differences in all cognitive assessments and some physical measures were observed in pre-AD individuals. This is consistent with: 1) genetic cohorts highlighting a long prodromal phase of AD identified using neuroimaging [29], 2) the concept of Mild Cognitive Impairment as a precursor to AD, and 3) the finding of early visual memory deficits 10 years prior to AD symptoms [30].

We also identified syndrome-relevant changes in other diseases. Pre-PSP individuals demonstrated an increased falls risk and reduced fluid intelligence score, reflecting the typical Richardson syndrome of PSP of early falls and a dysexecutive cognitive impairment [31]. The poorer numeric memory in pre-MSA patients is noteworthy as cognitive impairment is not a dominant symptom in MSA. Nevertheless, cognitive impairment has been consistently identified in at least a portion of MSA patients [32, 33], with frontal-executive dysfunction being the most common cognitive feature [34]. In addition, the pre-MSA and pre-PSP groups exhibited a rapid functional decline in falls risk and overall health rating leading up to the time of diagnosis.

Conversely, we were able to demonstrate that pre-PD individuals have preserved pre-symptomatic cognition and good evidence of preserved outcomes on some functional measures; in particular, the data for reaction time, pairs matching round 2 and weight gain in the past year demonstrate convincingly that there is no difference in these domains between pre-PD individuals and controls. Our study focused on cognition and function, so we did not capture the well recognised early systemic features seen in a proportion of people with PD [35].

Identifying early subtle changes in cognition and function could enable stratification into prevention trials targeting known risk factors [36], or early disease modification. Studies of prevention are ongoing with some evidence that treating blood pressure in middle age reduces cerebral white matter disease [37, 38], and that a multidomain preventive approach may reduce the risk of cognitive decline in a population aged 60-77 years of age [39]. However, most lifestyle factors are targeted at the vascular risk factors associated with AD, whereas other pathologies such as tau and *α*-synuclein are not associated with such risk.

In these other pathologies, early disease modifying treatments are being actively pursued. Given the toxicity of many such treatments [40], one would need to be confident in identifying specific pathologies, and initiating treatment at a time that maximises the risk-benefit ratio. The most common changes we identified were in fluid intelligence, memory and reporting of overall health. We speculate that screening of these domains could lead to additional pathology-specific biomarker assessments such as PET scans, CSF or blood-based biomarkers [41–48].

There are several limitations to our study. Analysis of the FTD (n=211), PSP (n=133), DLB (n=40) and MSA (n=73) groups was limited by the smaller sample size available, which resulted in wide uncertainty over the posterior distribution. In DLB this partly reflected the recording of the diagnosis in the UK Biobank, as it was only possible to search for it within the primary care dataset. Whilst the UK Biobank is a population-based study, it is biased towards a population with a lower risk of disease in general [49], and is not representative of ethnic and socioeconomic diversity in the UK. This may limit the generalizability of our results. Furthermore, the study can only demonstrate correlation and elucidation of the underlying pathophysiology is outside its scope.

In conclusion, our study identifies early functional and cognitive differences in the pre-diagnostic stage of multiple neurodegenerative diseases. Better characterisation of the pre-diagnostic stage will enable better risk stratification for prevention and disease-modifying studies[50].

## Supporting information

Graphs of diagnostics for BRMS models

Posterior predictive checks for BRMS models

## Data Availability

Data is available from the UK Biobank through the dataset managers.

https://www.ukbiobank.ac.uk/

## Acknowledgements

This research has been conducted using data from UK Biobank, a major biomedical database (http://www.ukbiobank.ac.uk/). This work has been funded by the MRC (SUAG/051 R101400) and Cambridge Centre for Parkinson-plus; and the NIHR Cambridge Biomedical Research Centre (BRC-1215-20014). The views expressed are those of the author(s) and not necessarily those of the NIHR or the Department of Health and Social Care. For the purpose of open access, the authors have applied a CC BY public copyright licence to any Author Accepted Manuscript version arising from this submission.

## Conflicts of Interest

The authors report no conflicts of interest related to the current work. JBR has received fees for consultancy from Astex, Asceneuron, SV Health, ICG, Curasen, UCB, and Alzheimer Research UK, and received research grants from Janssen, Lilly, AstraZeneca.

## Notes

### Author Declarations

The UK biobank

