## Supplementary material for "Pre-Diagnostic Cognitive and Functional Impairment in Multiple Sporadic Neurodegenerative Diseases": Graphs of diagnostics for BRMS models

### Slide 1
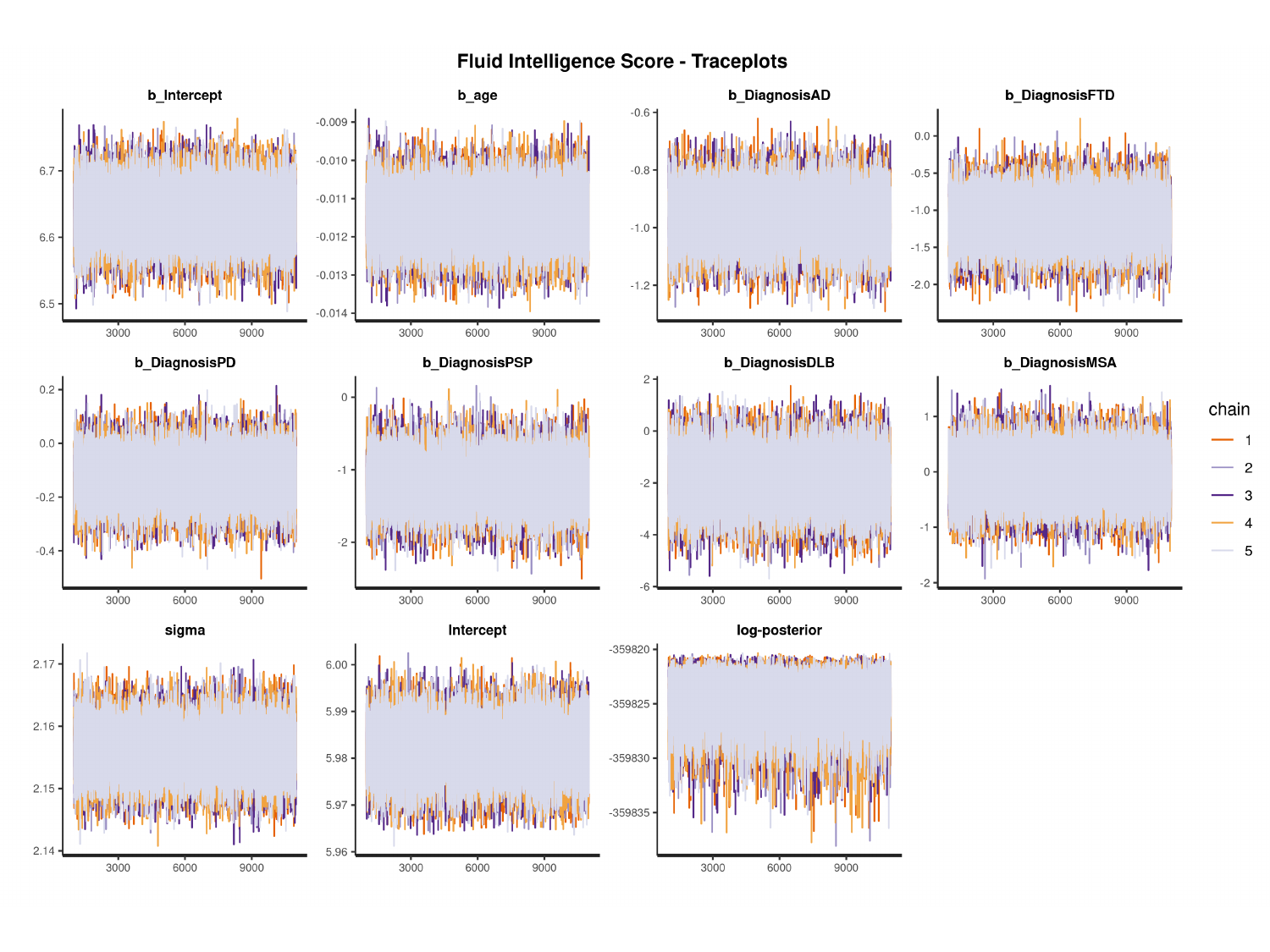

#

### Slide 2
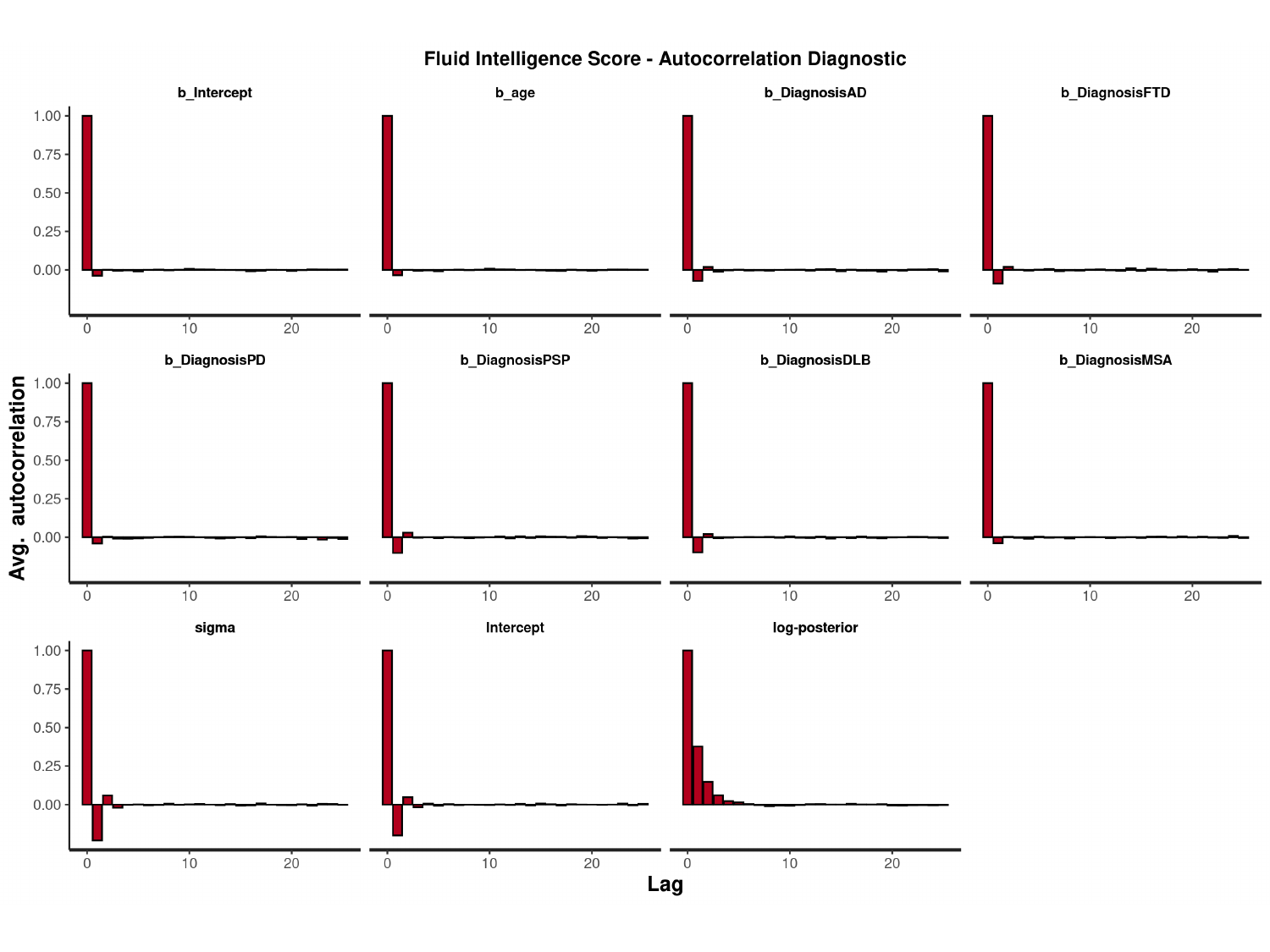

#

### Slide 3
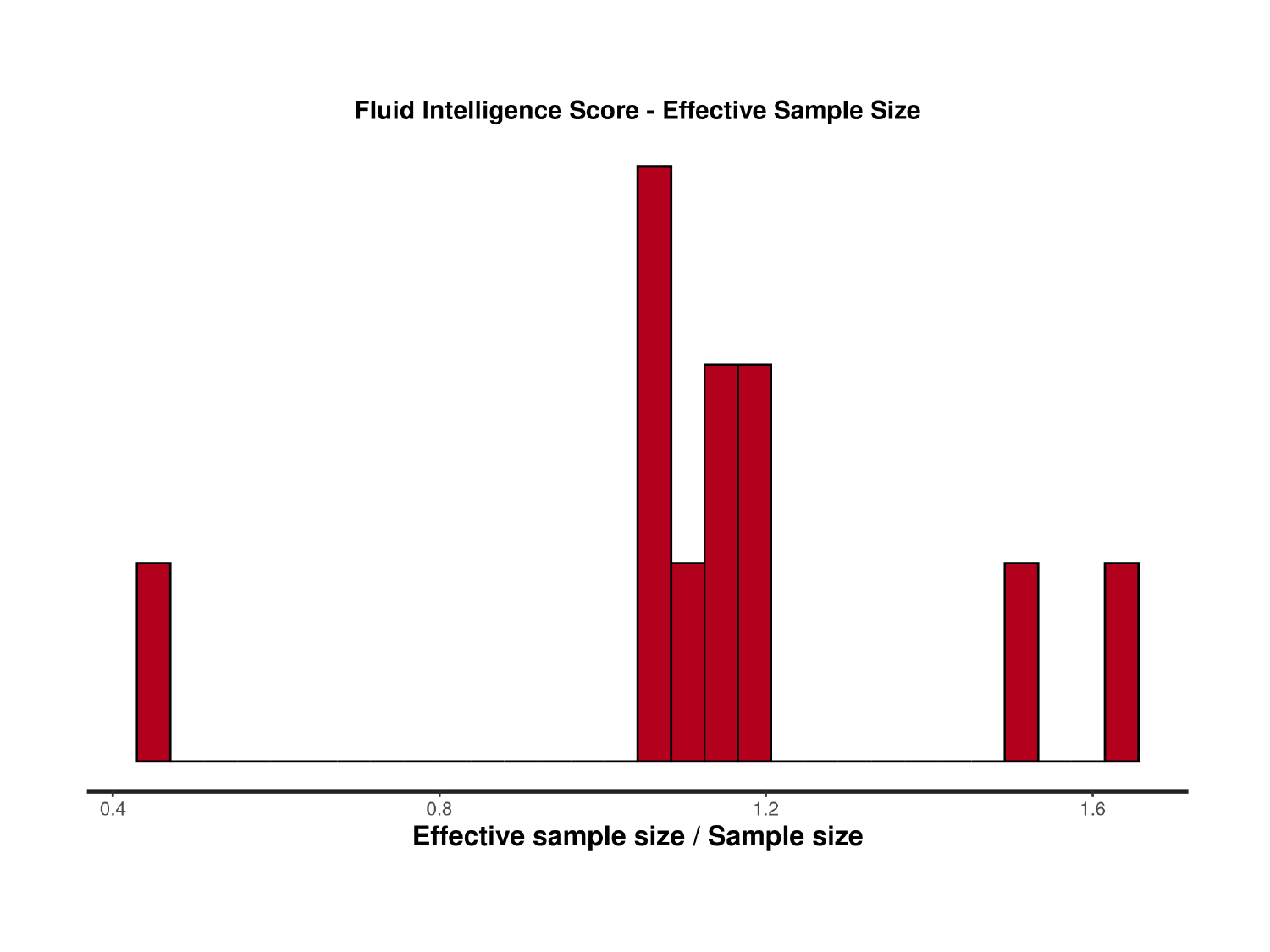

### Slide 4
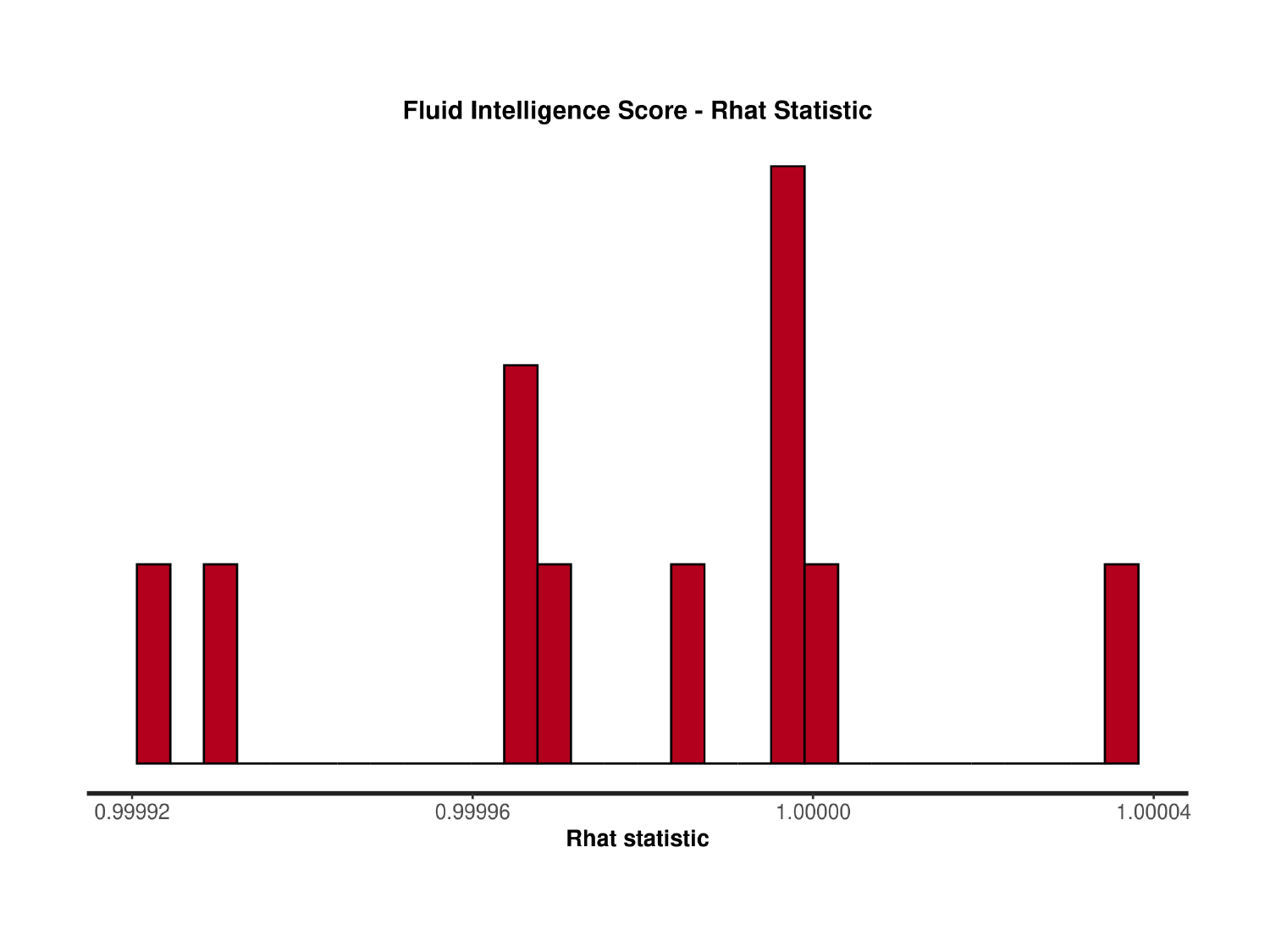

### Slide 5
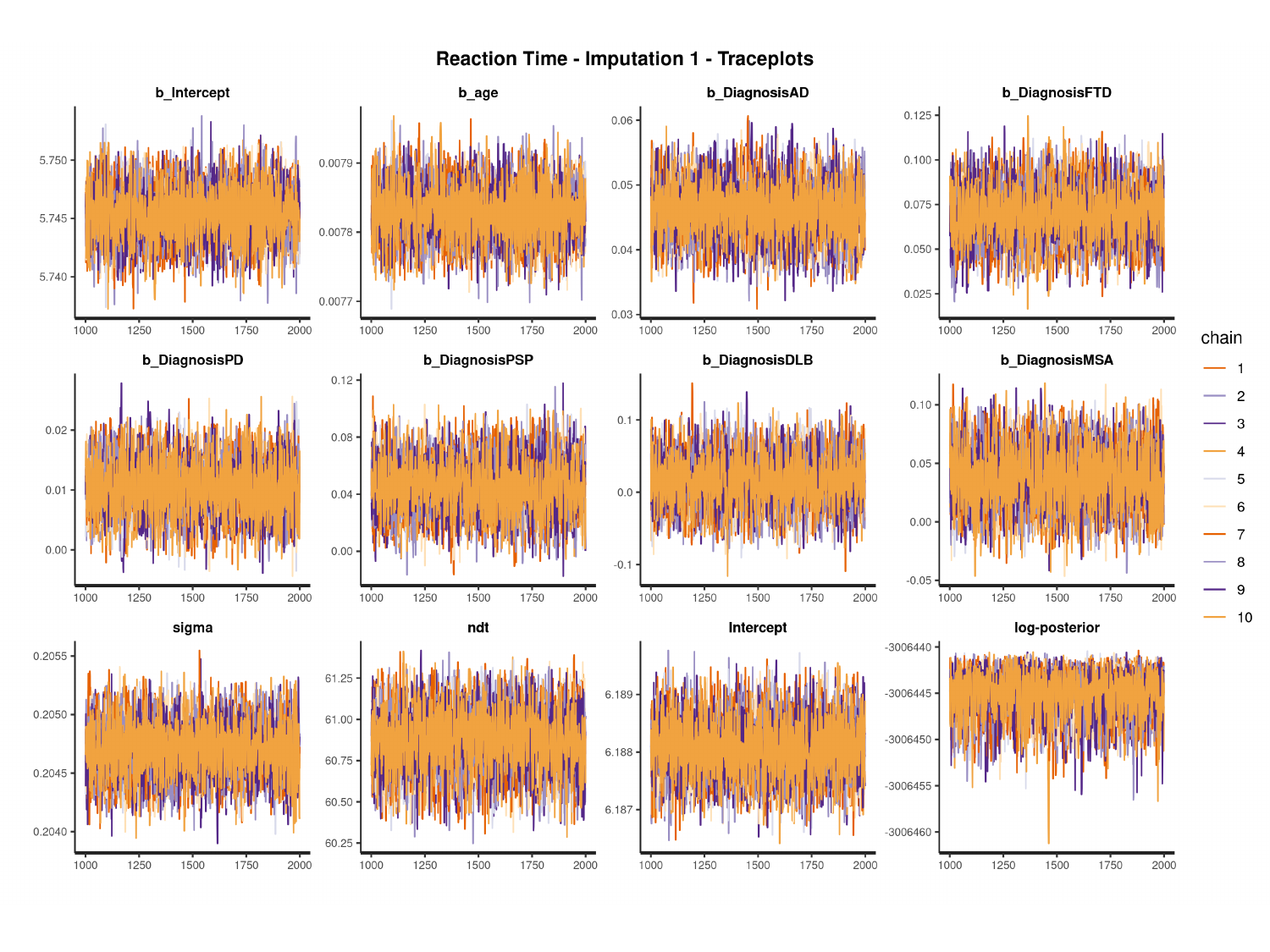

#

### Slide 6
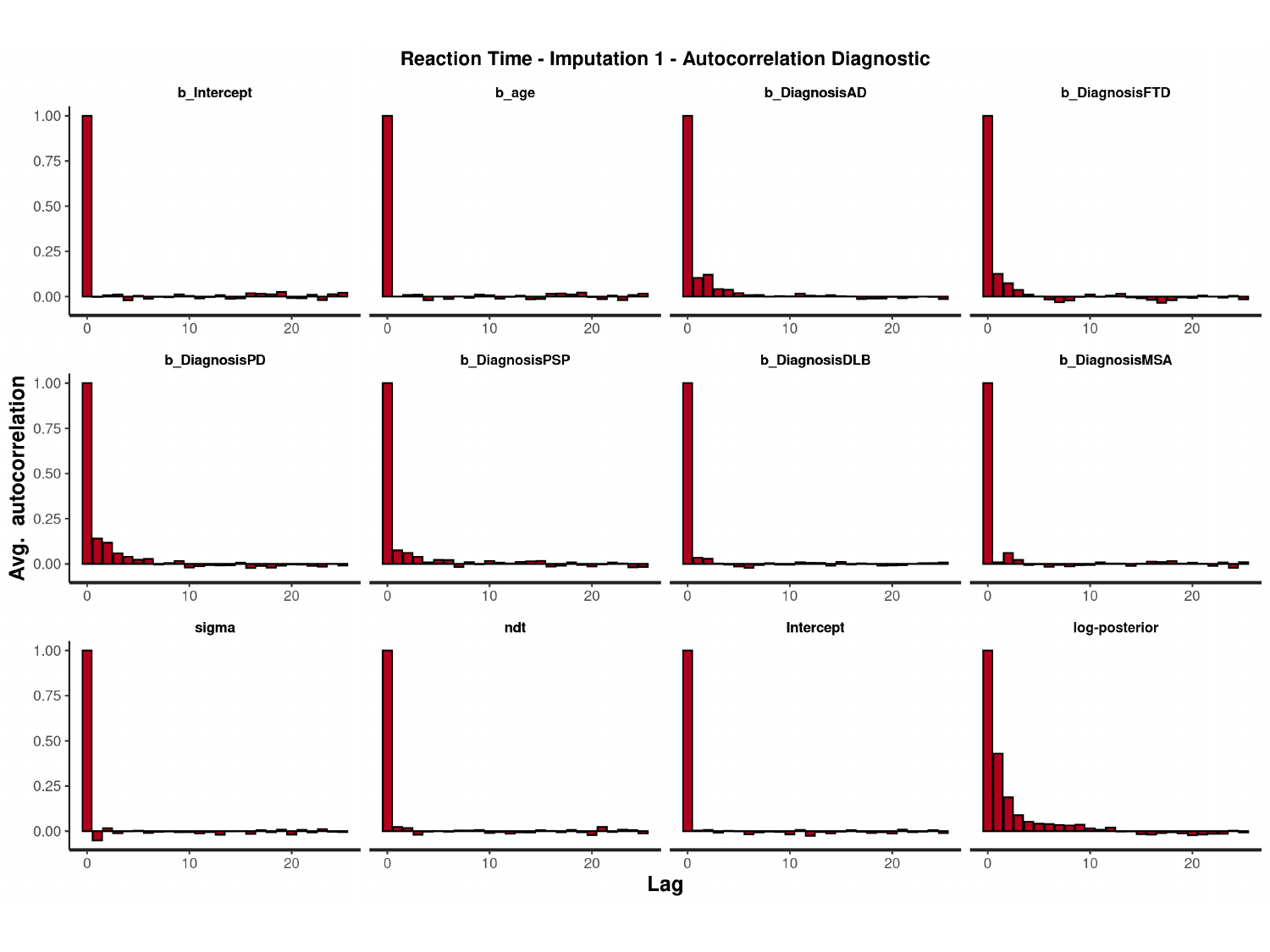

#

### Slide 7
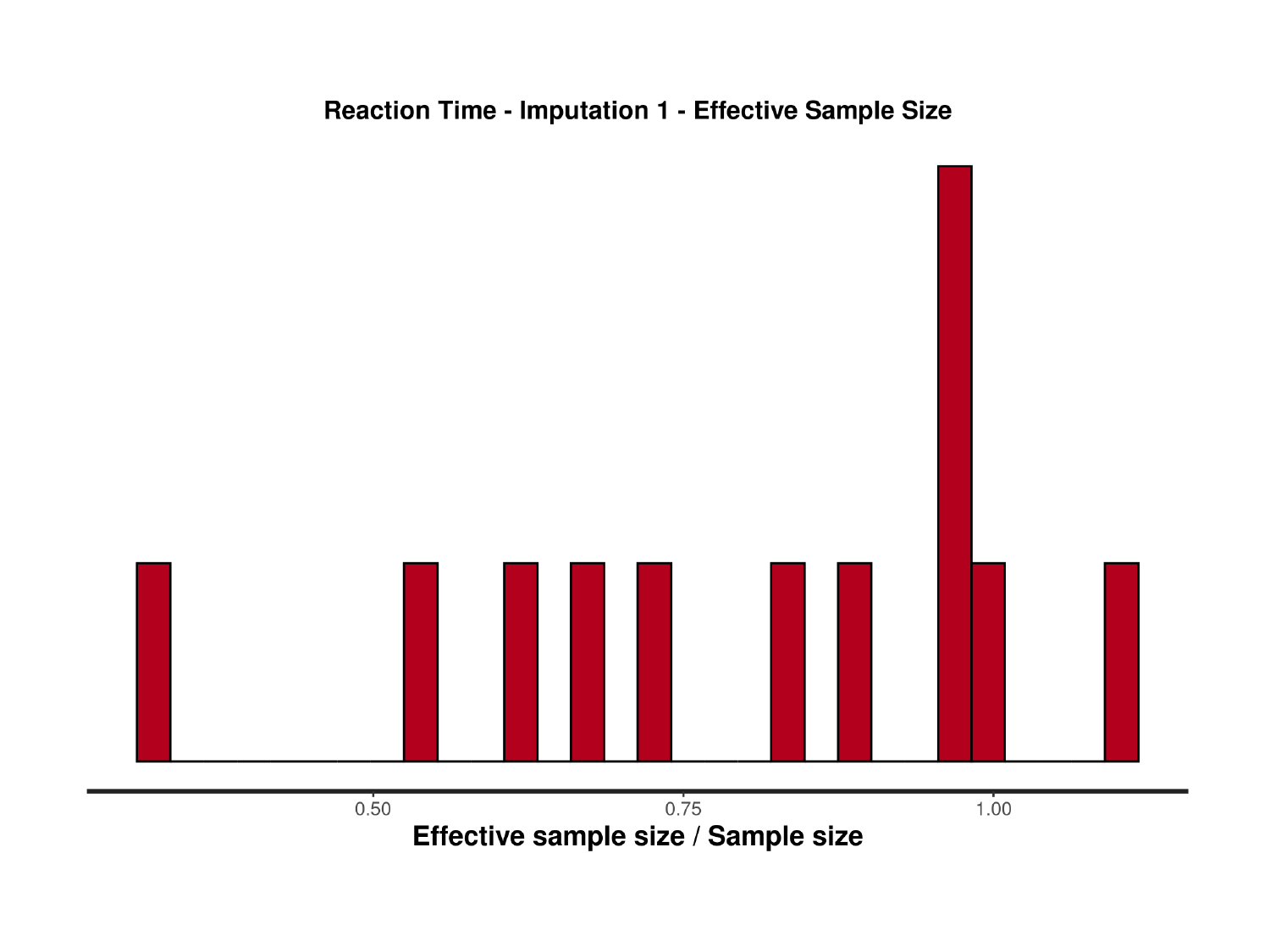

### Slide 8
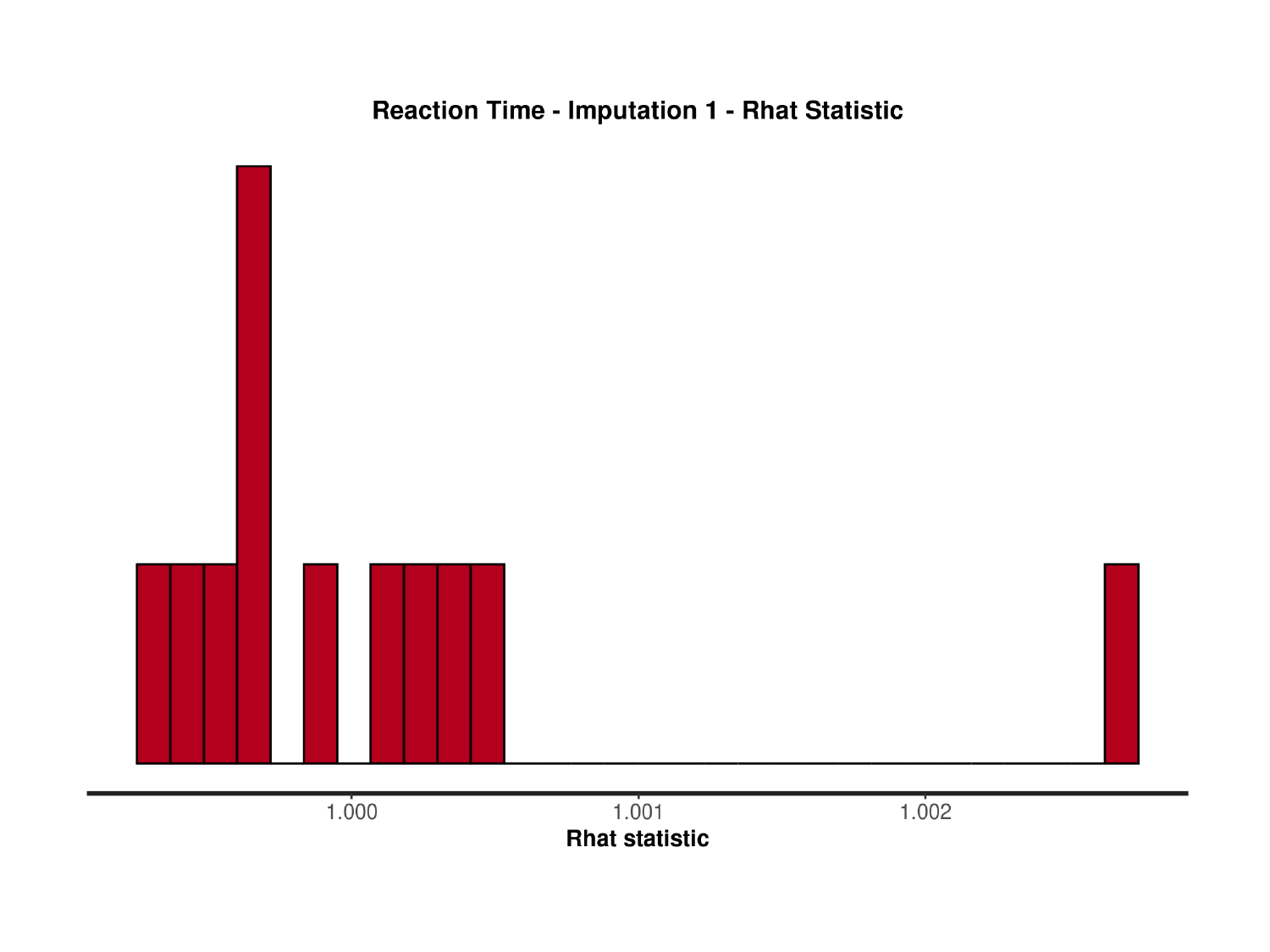

### Slide 9
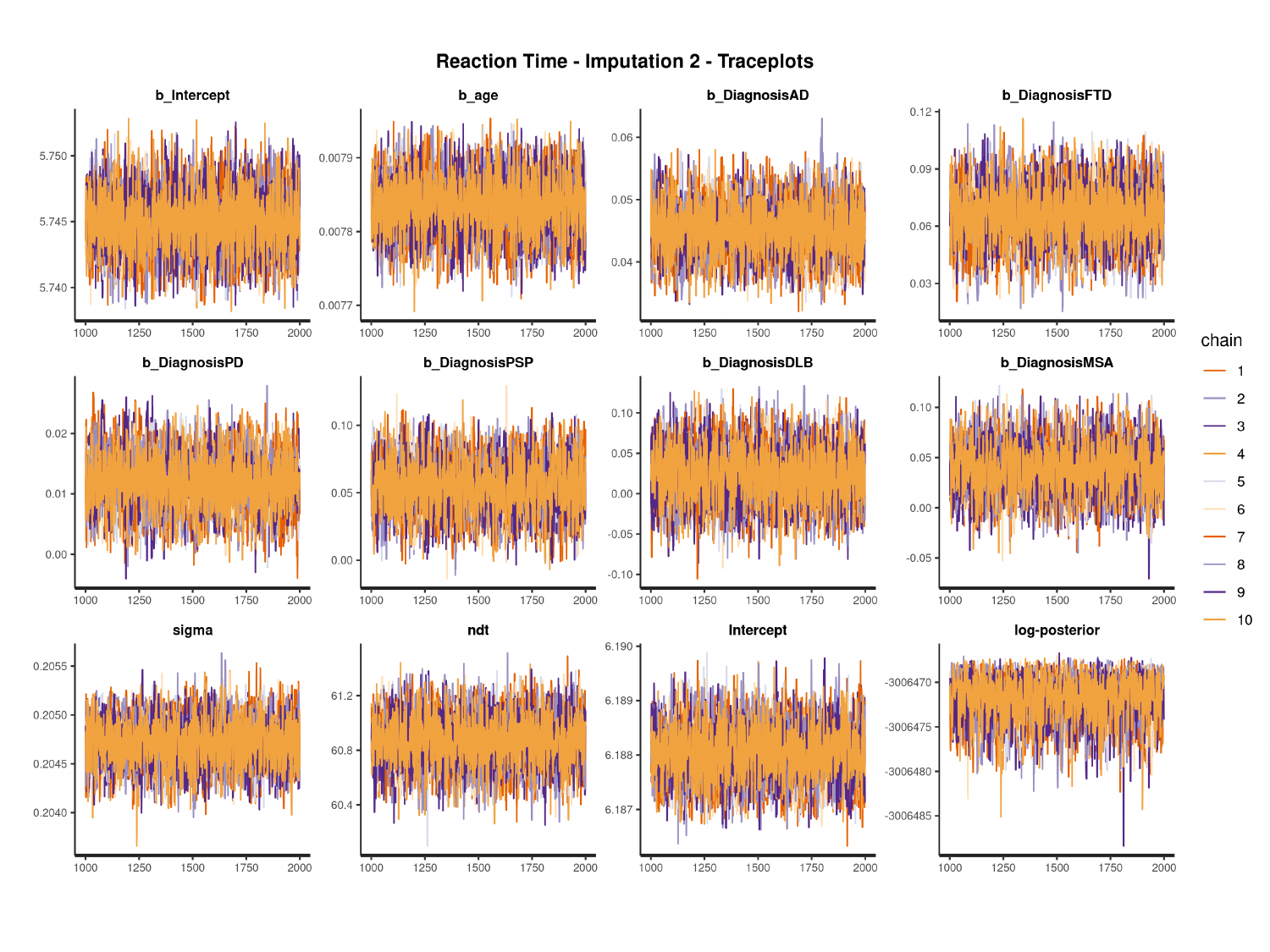

#

### Slide 10
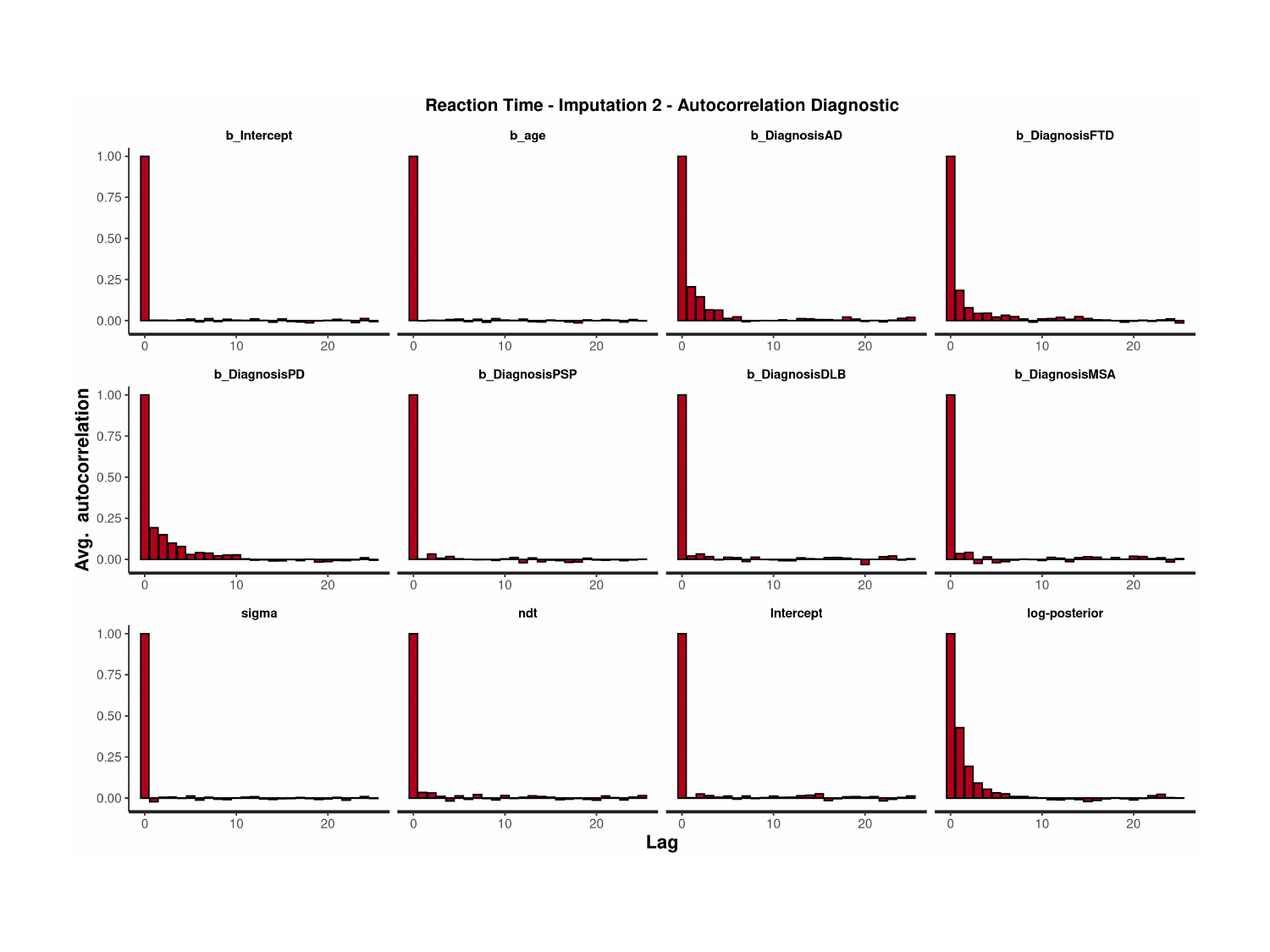

### Slide 11
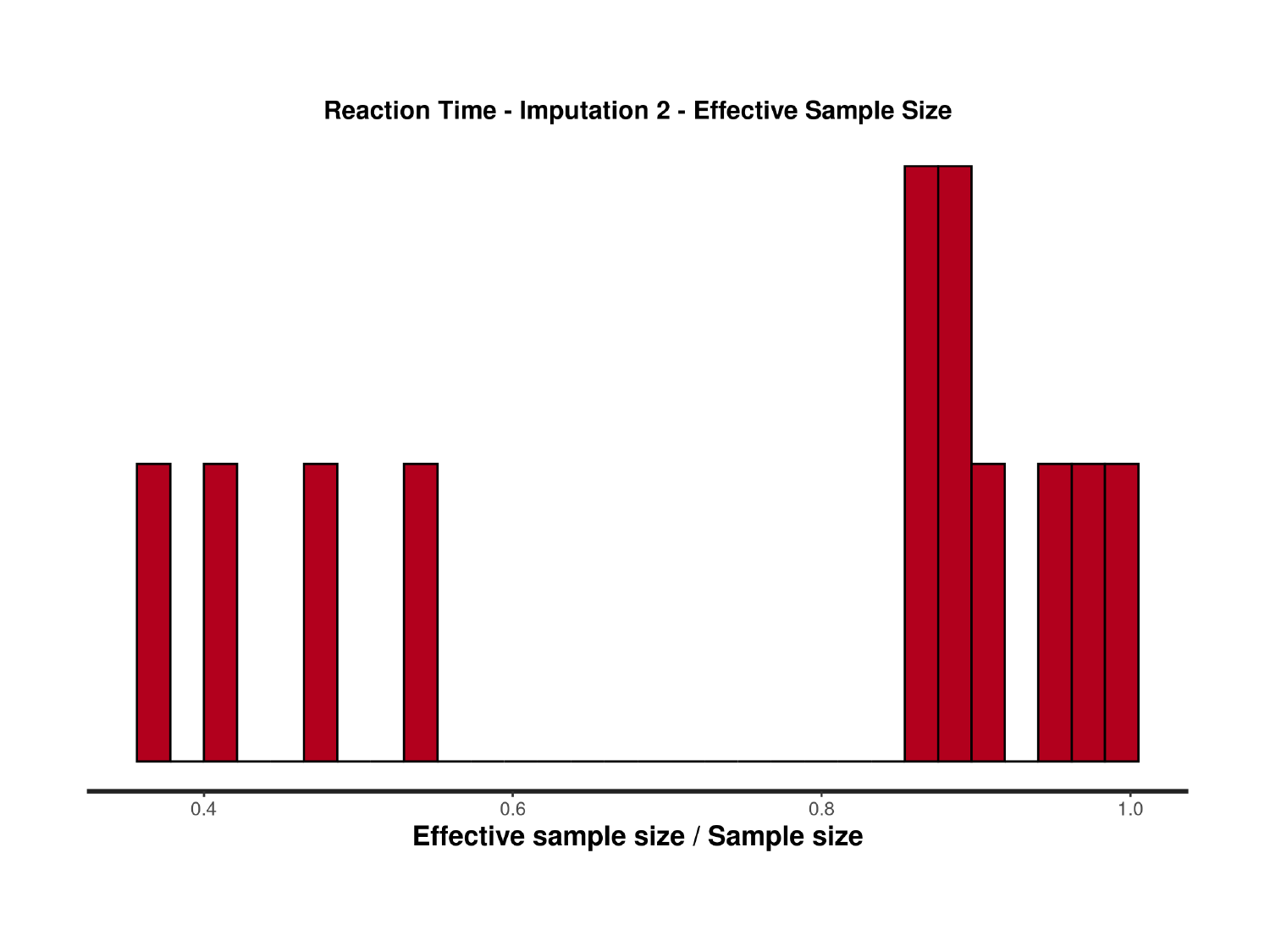

### Slide 12
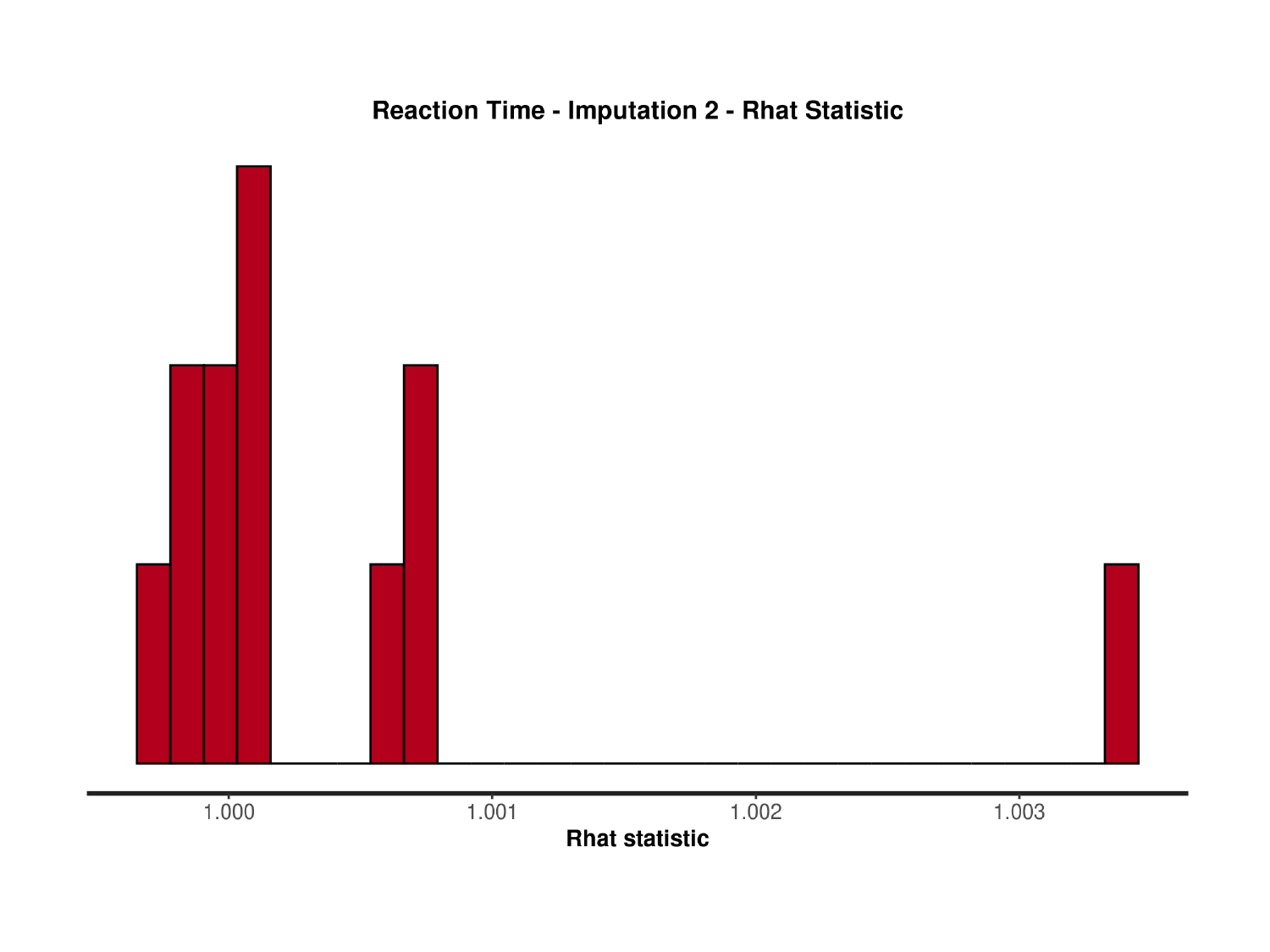

### Slide 13
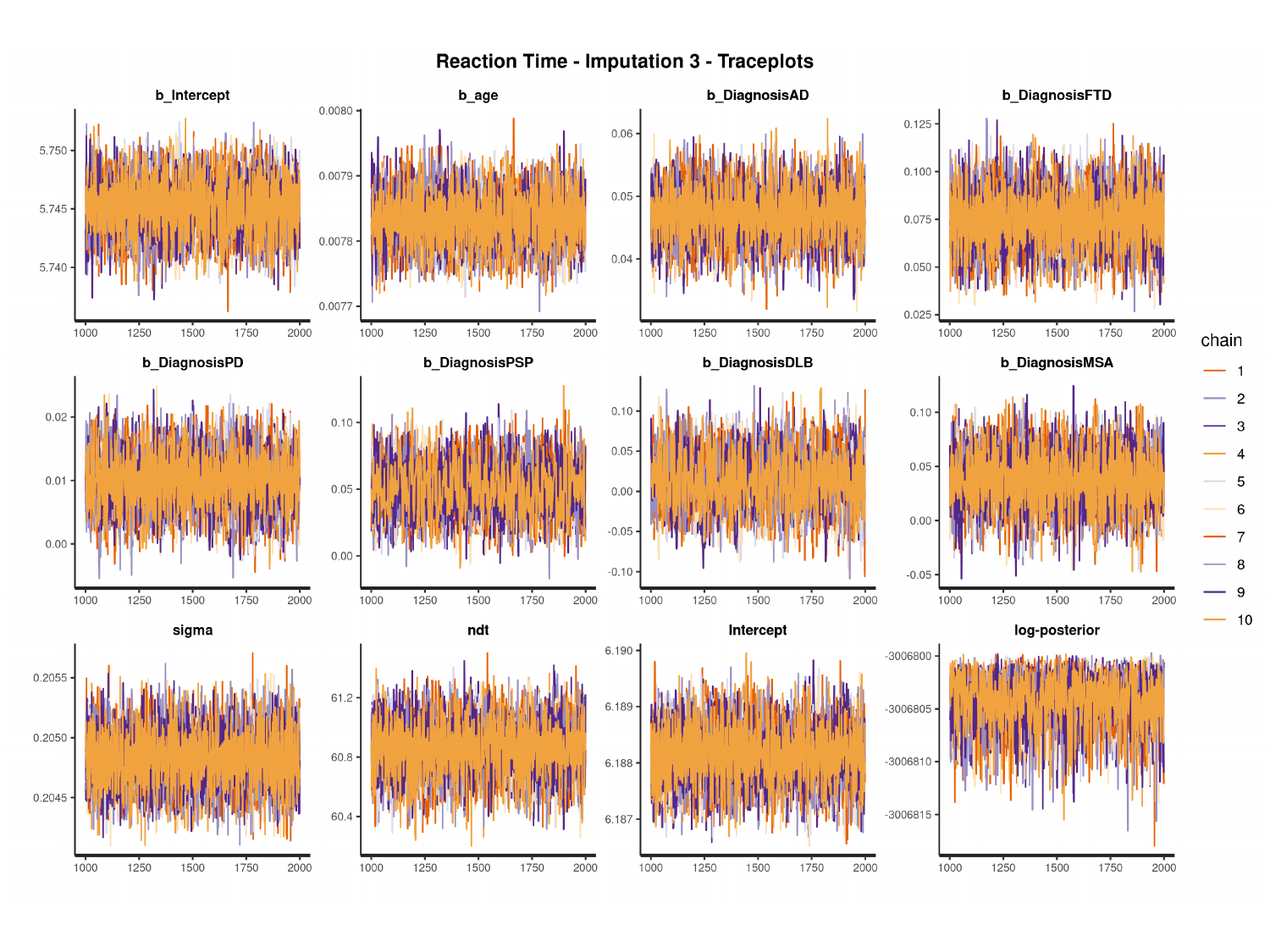

#

### Slide 14
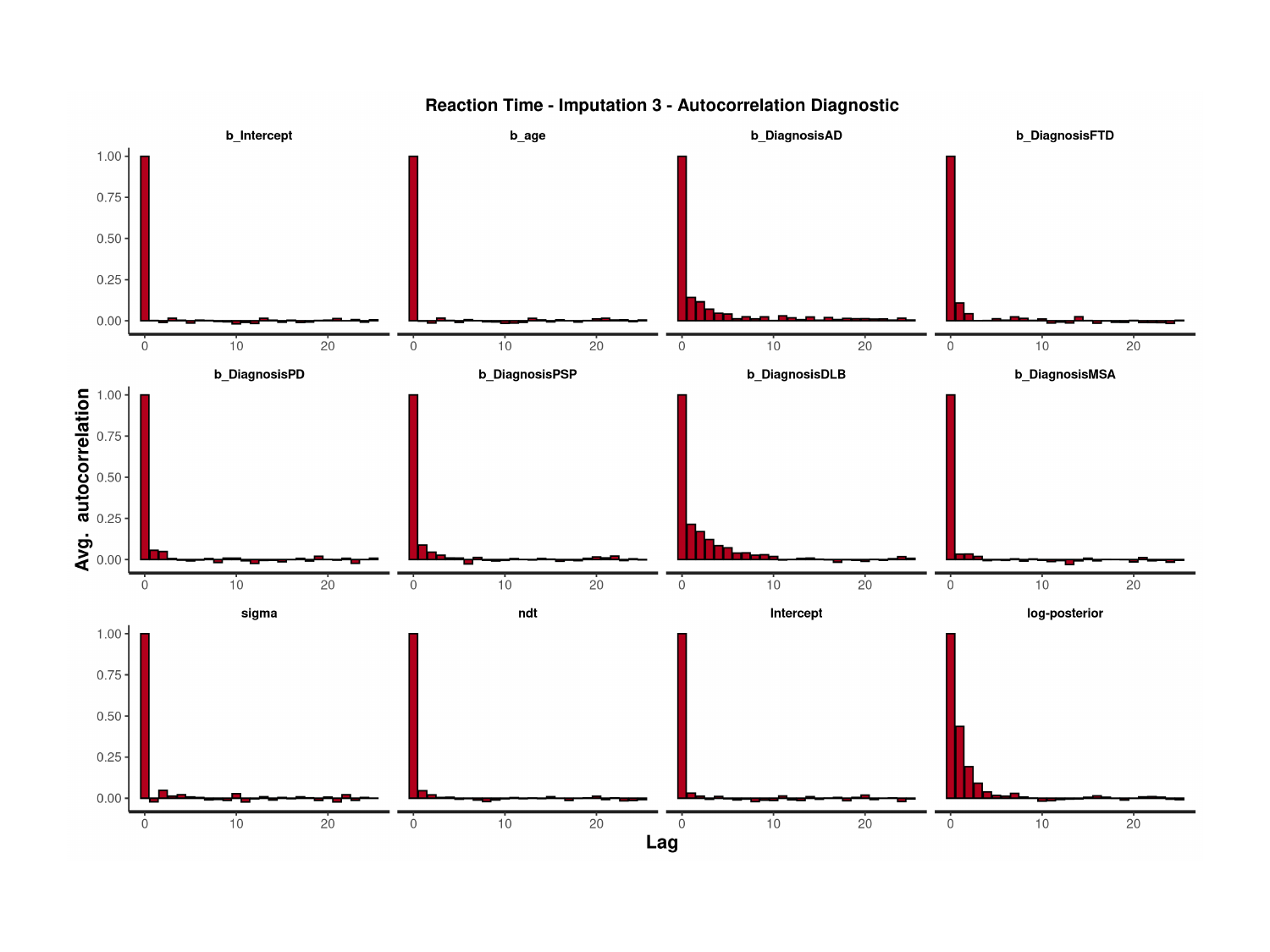

### Slide 15
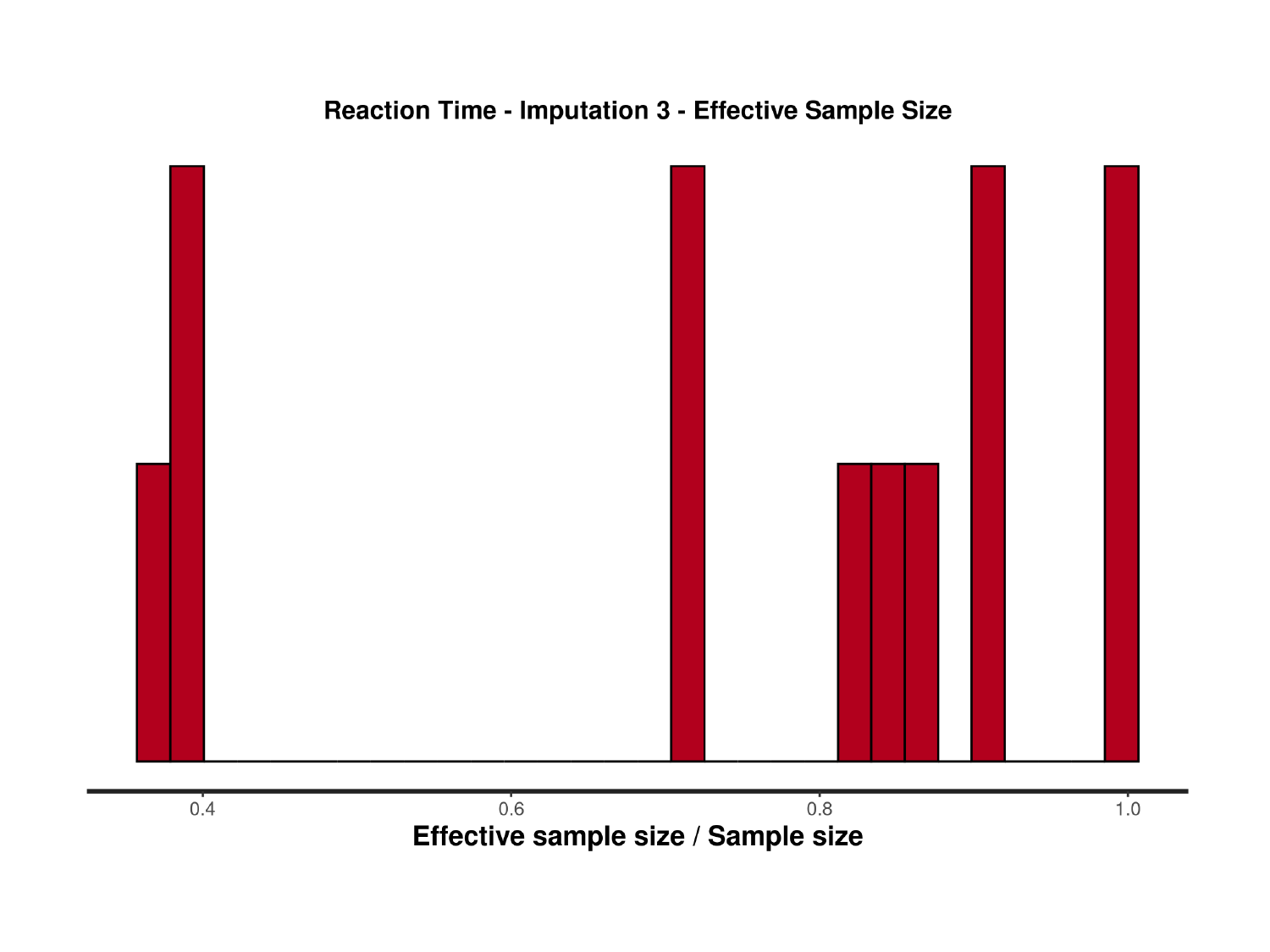

### Slide 16
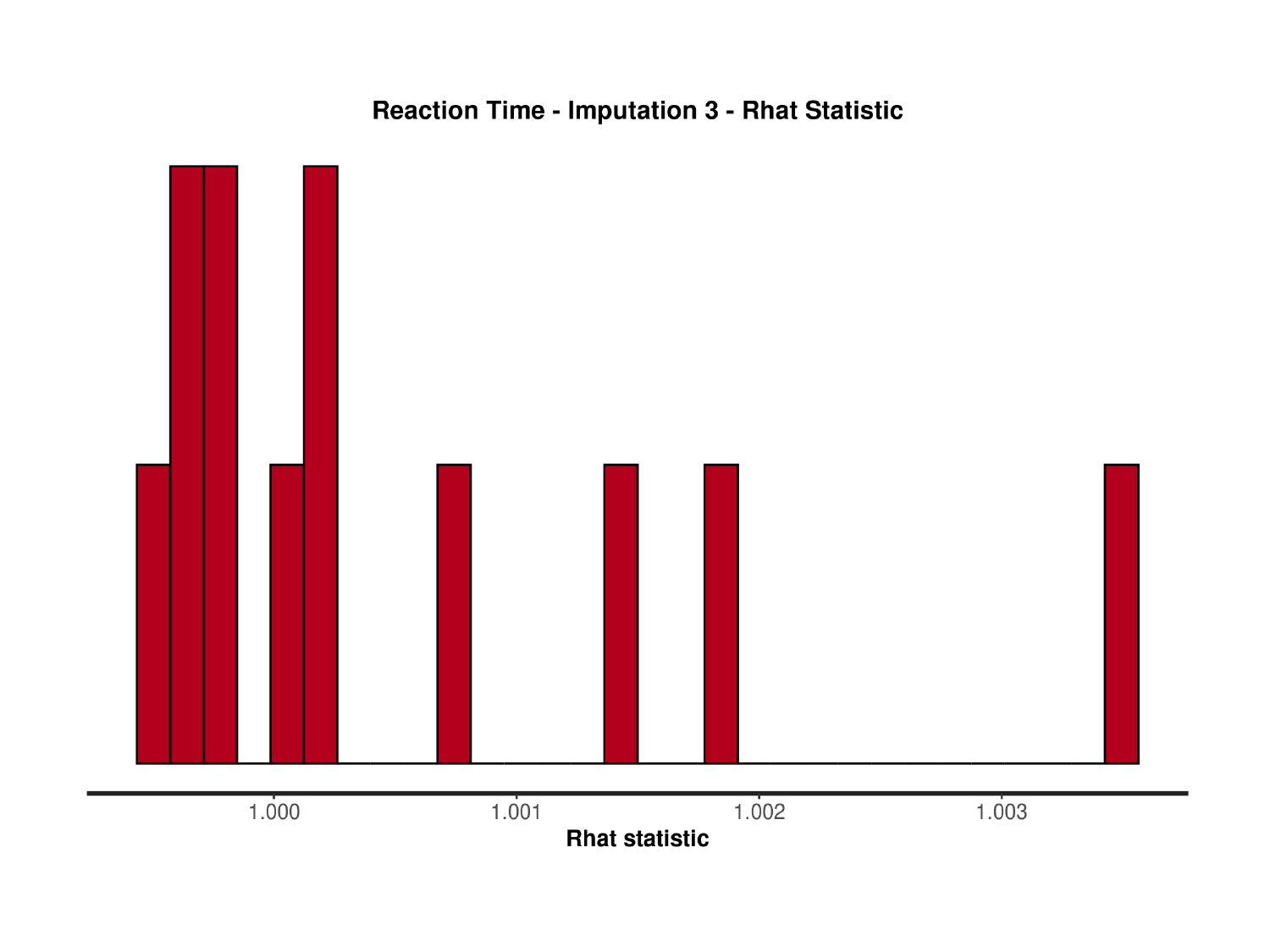

### Slide 17
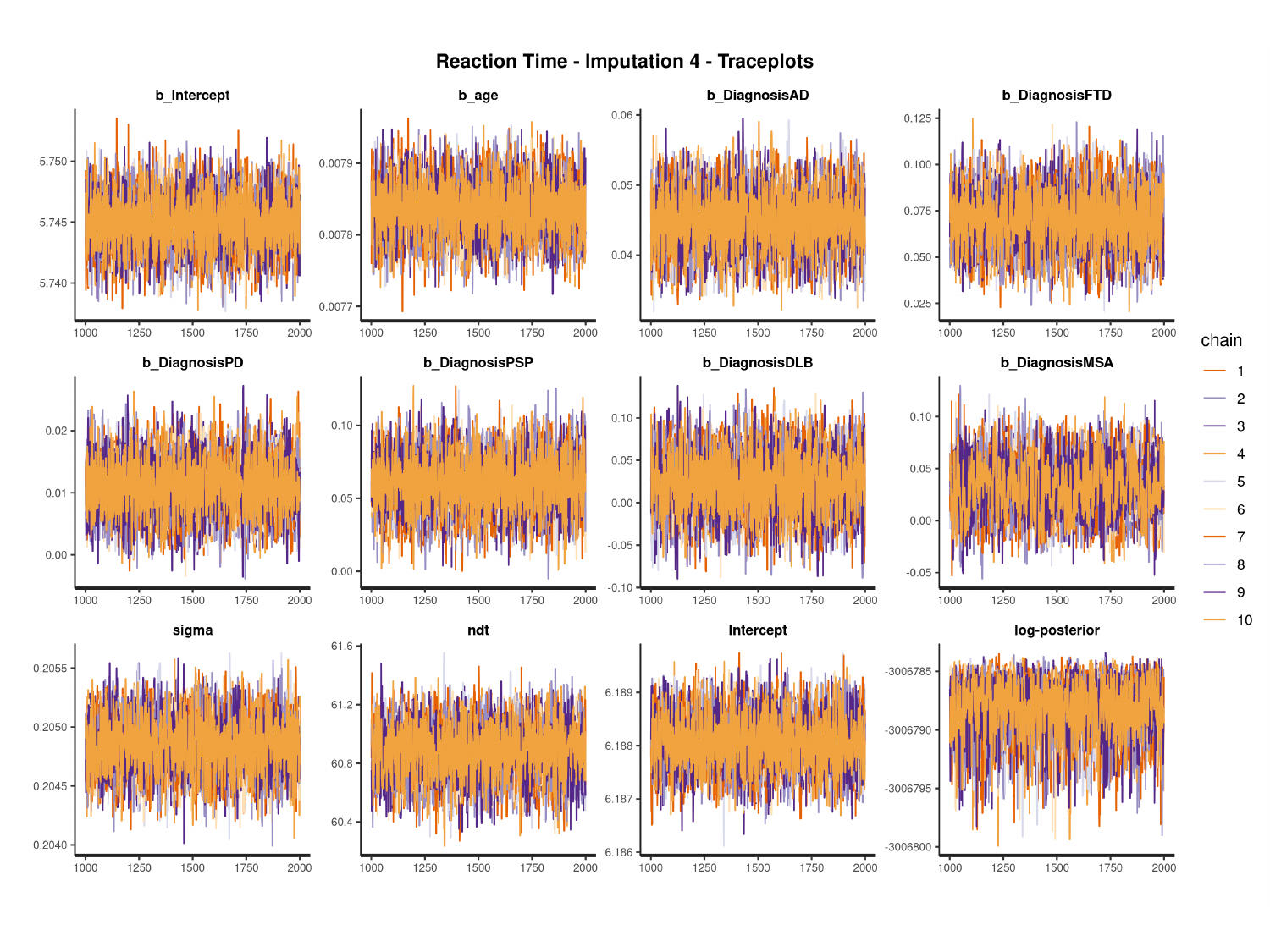

#

### Slide 18
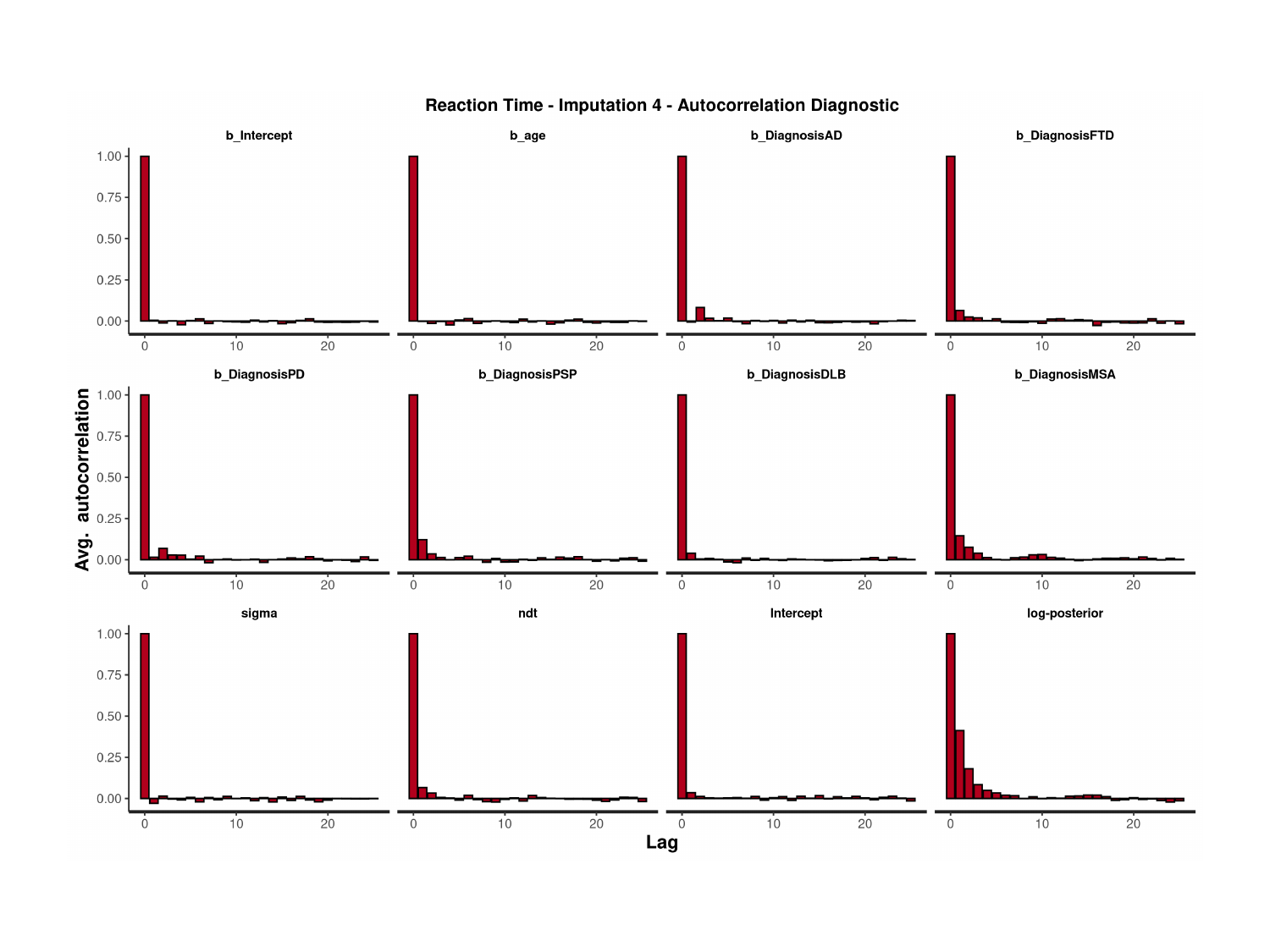

### Slide 19
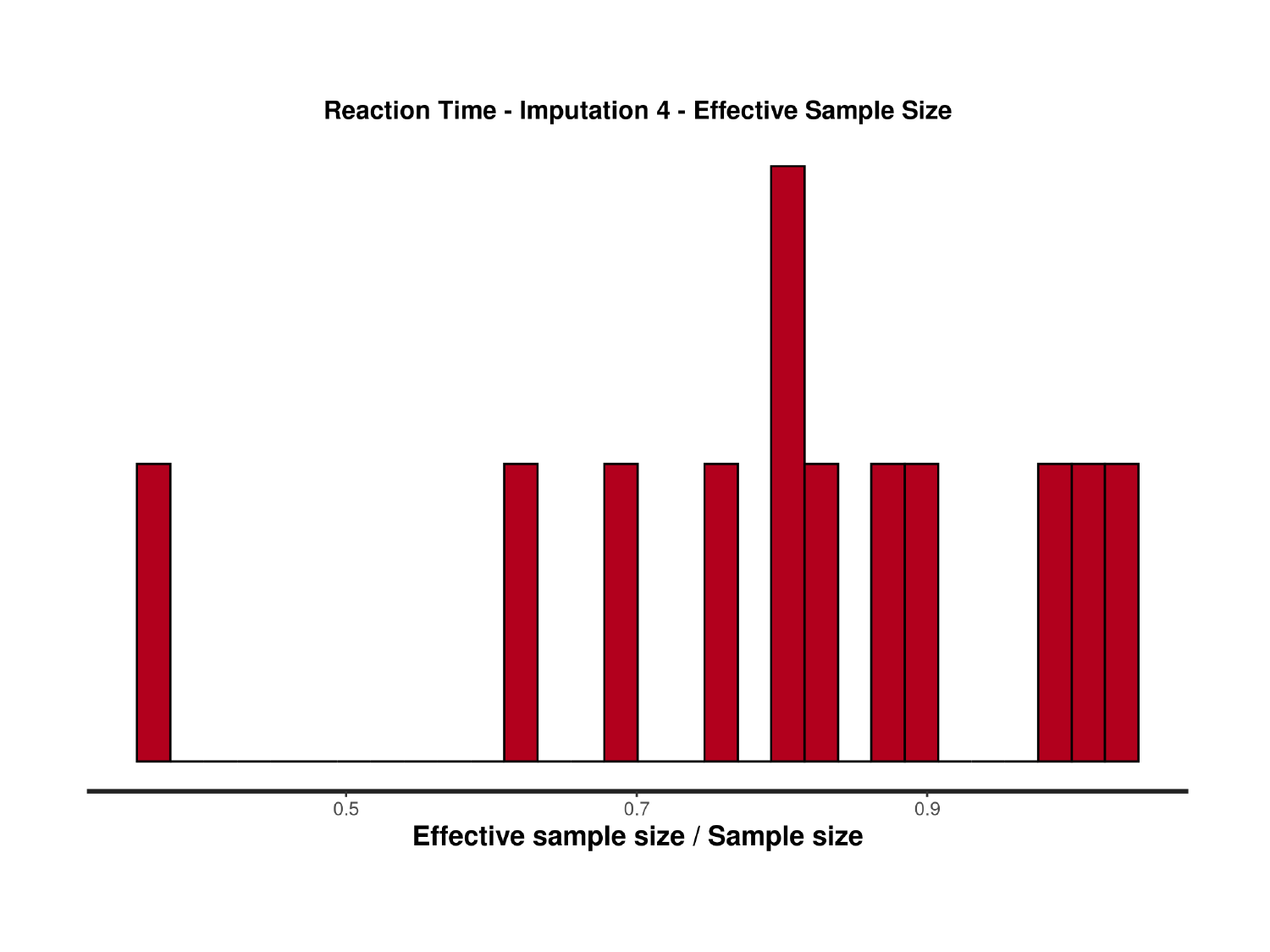

### Slide 20
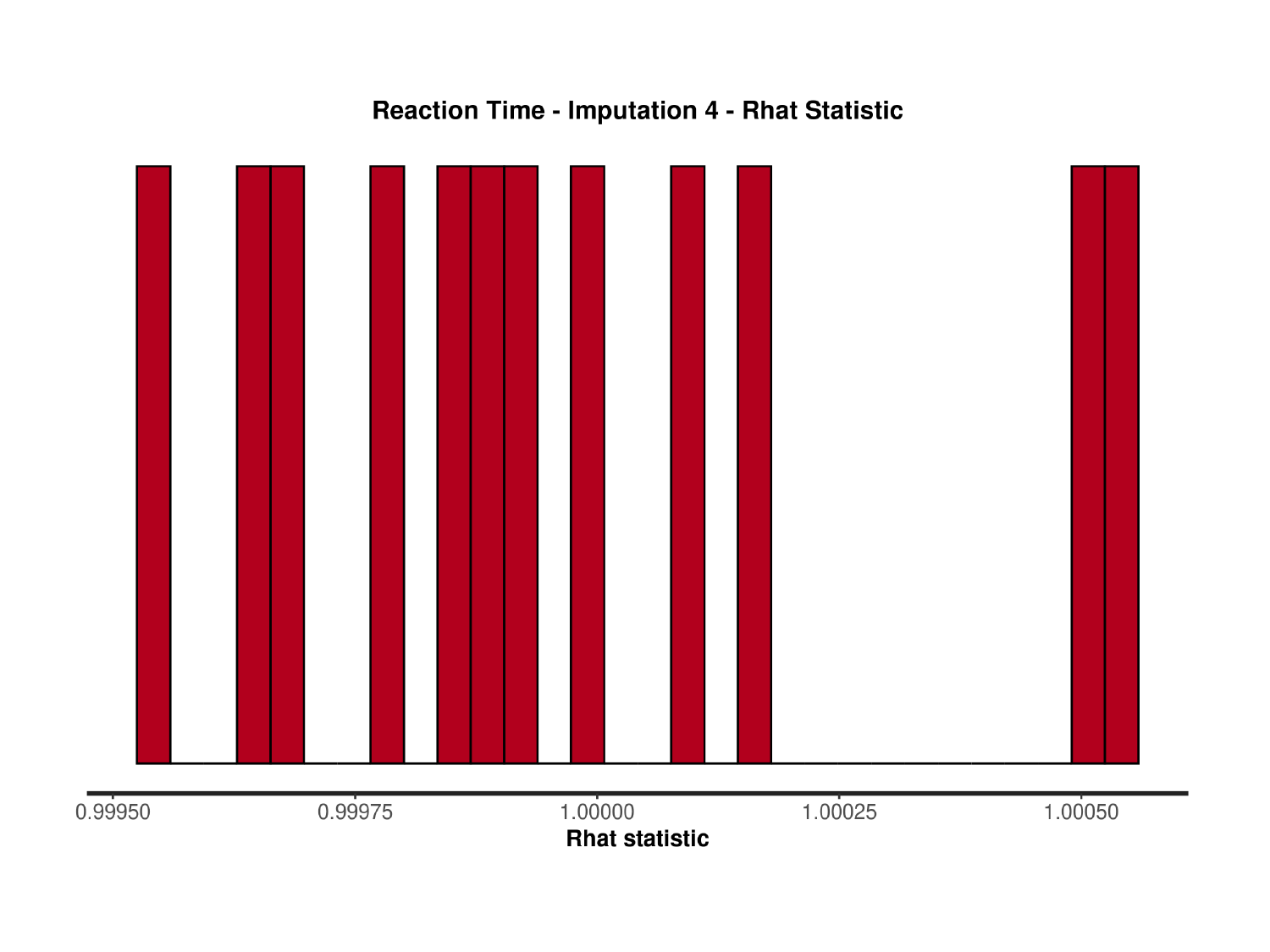

### Slide 21
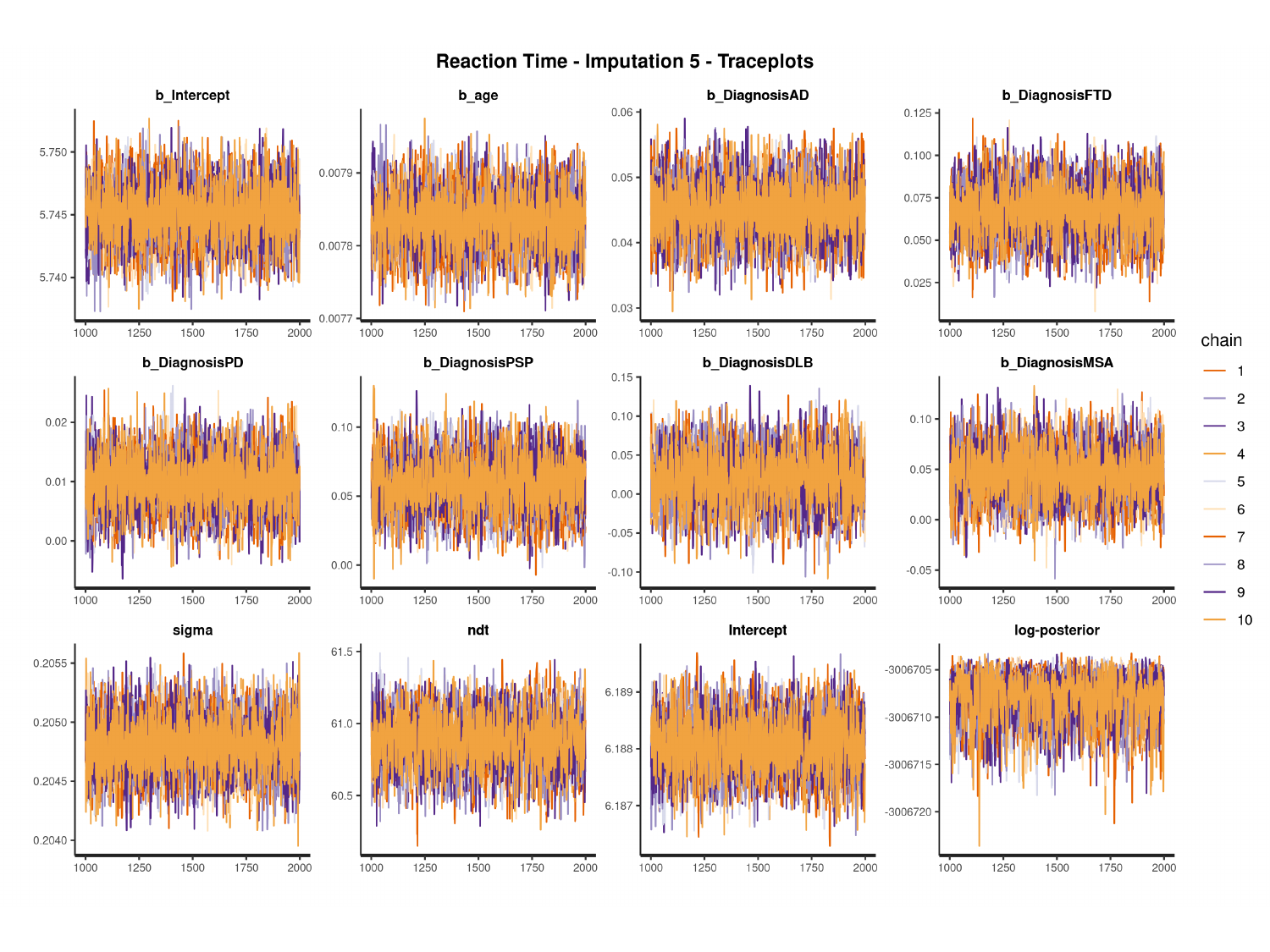

#

### Slide 22
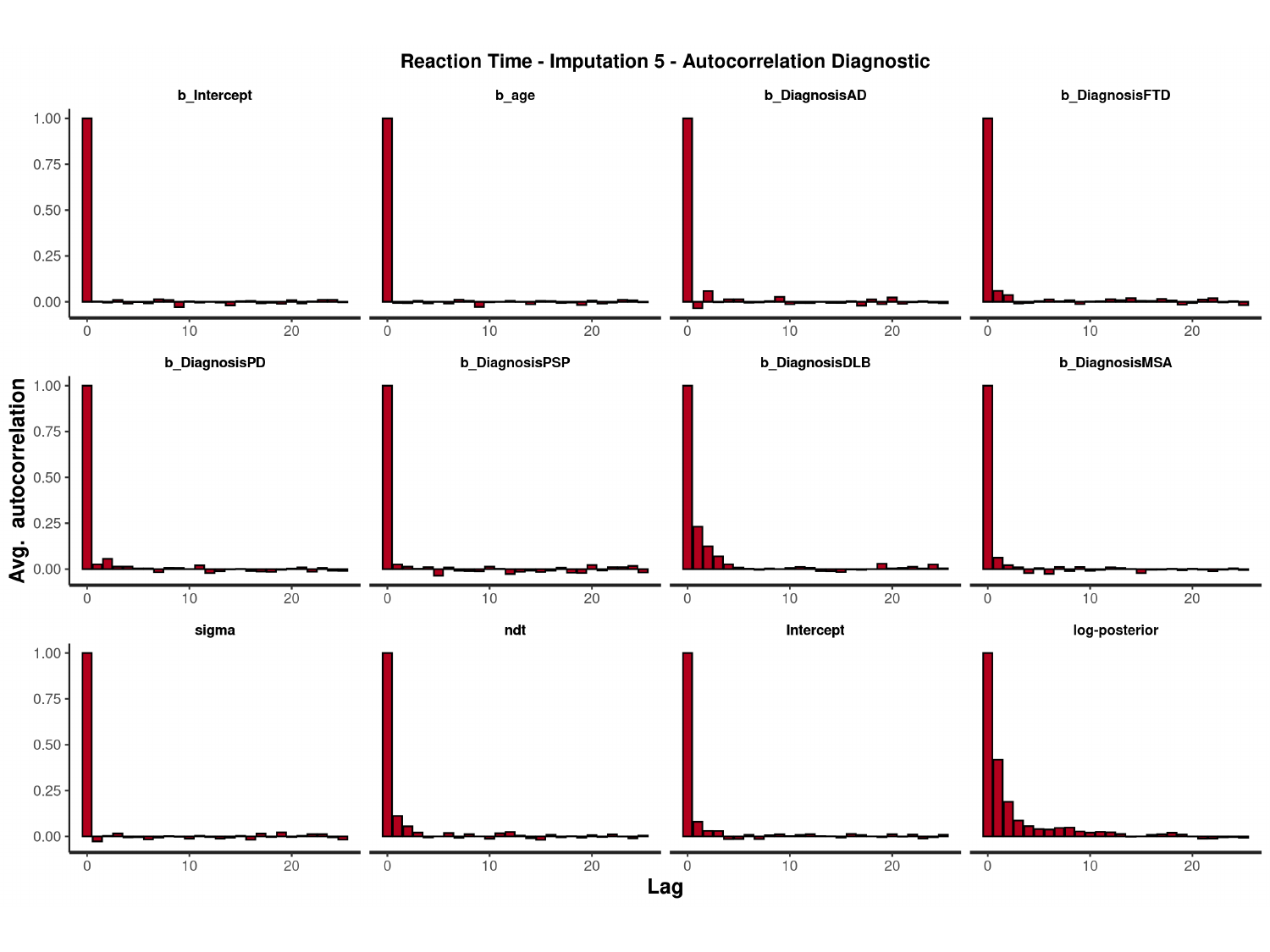

#

### Slide 23
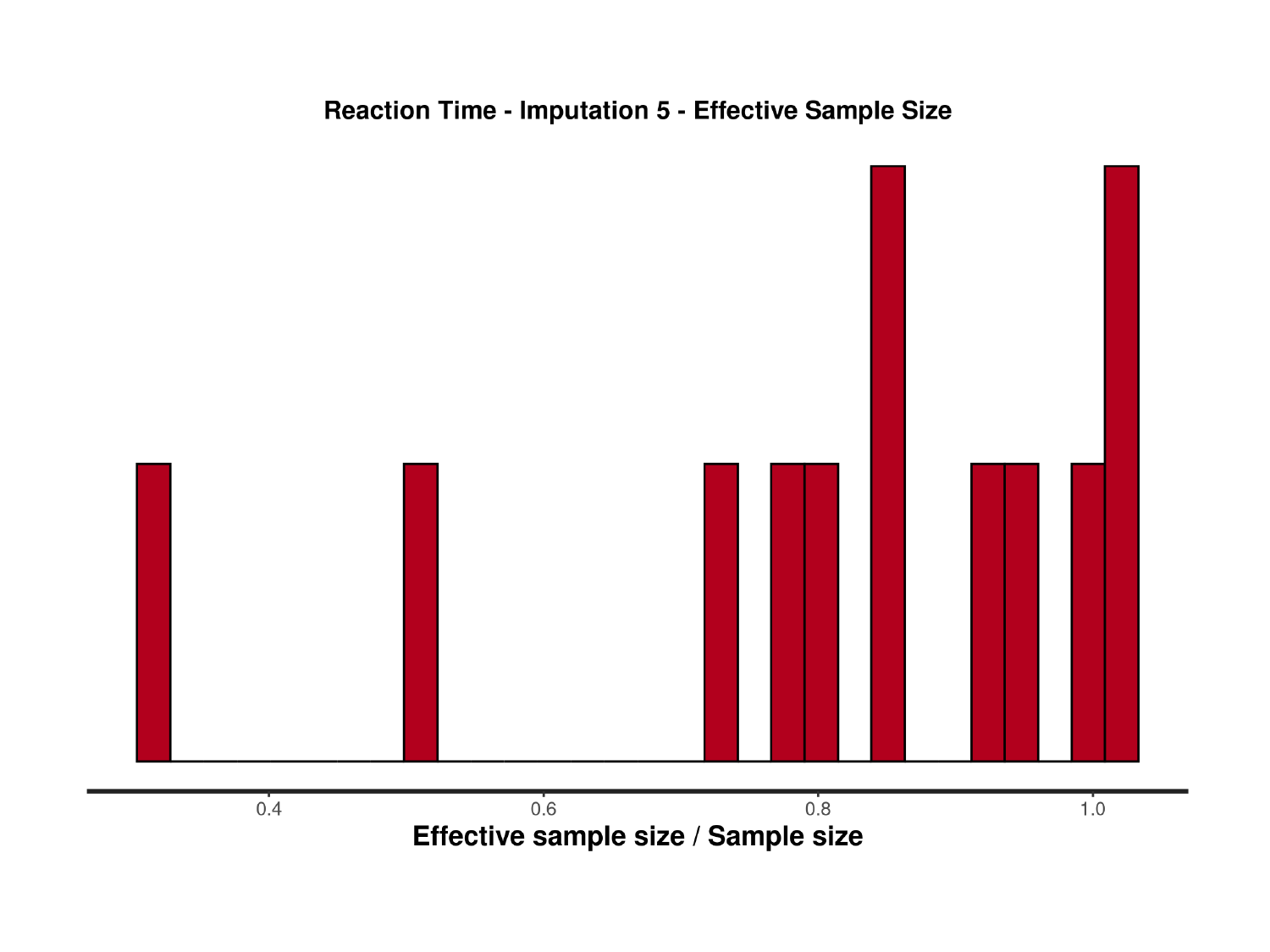

### Slide 24
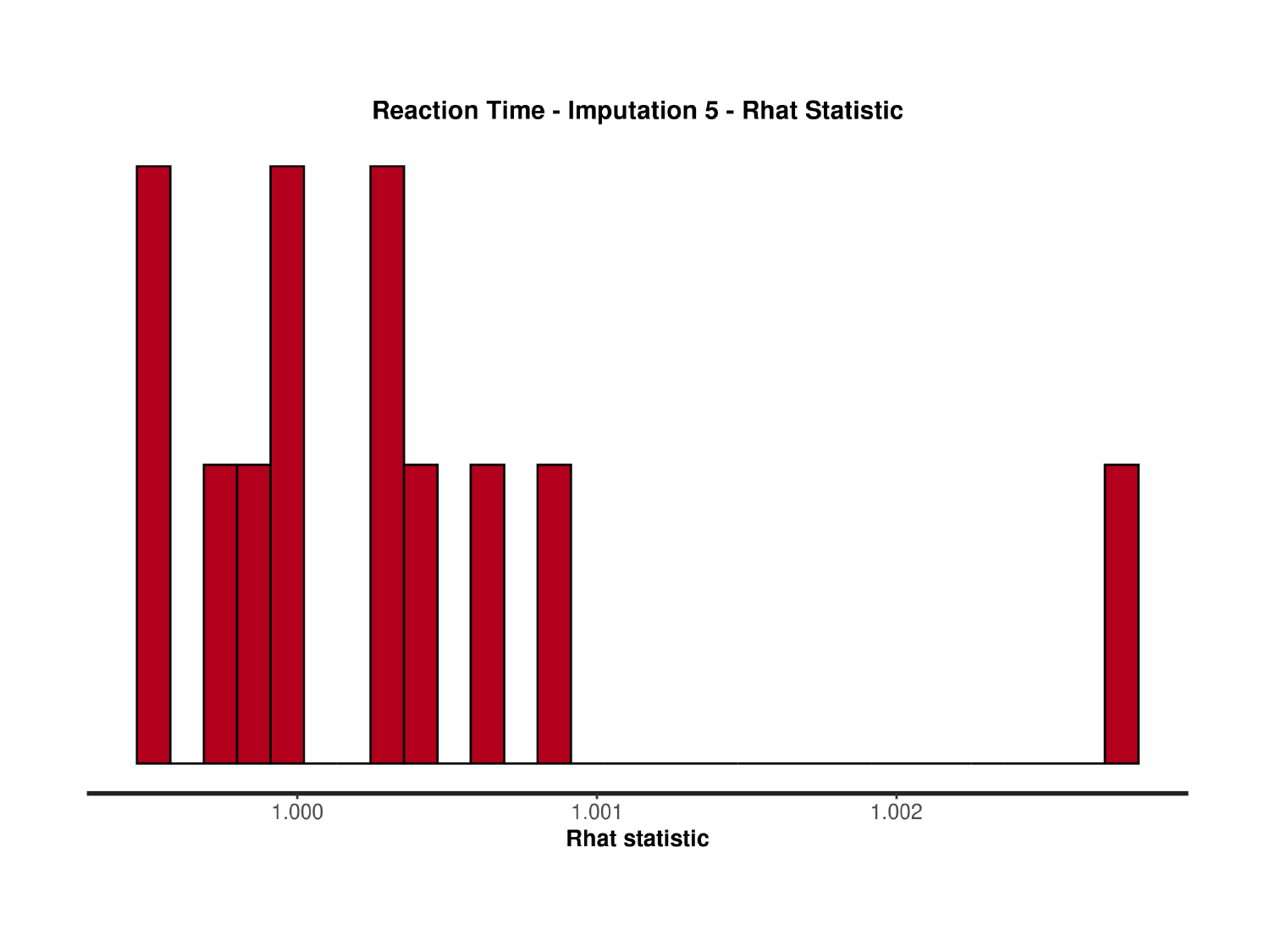

### Slide 25
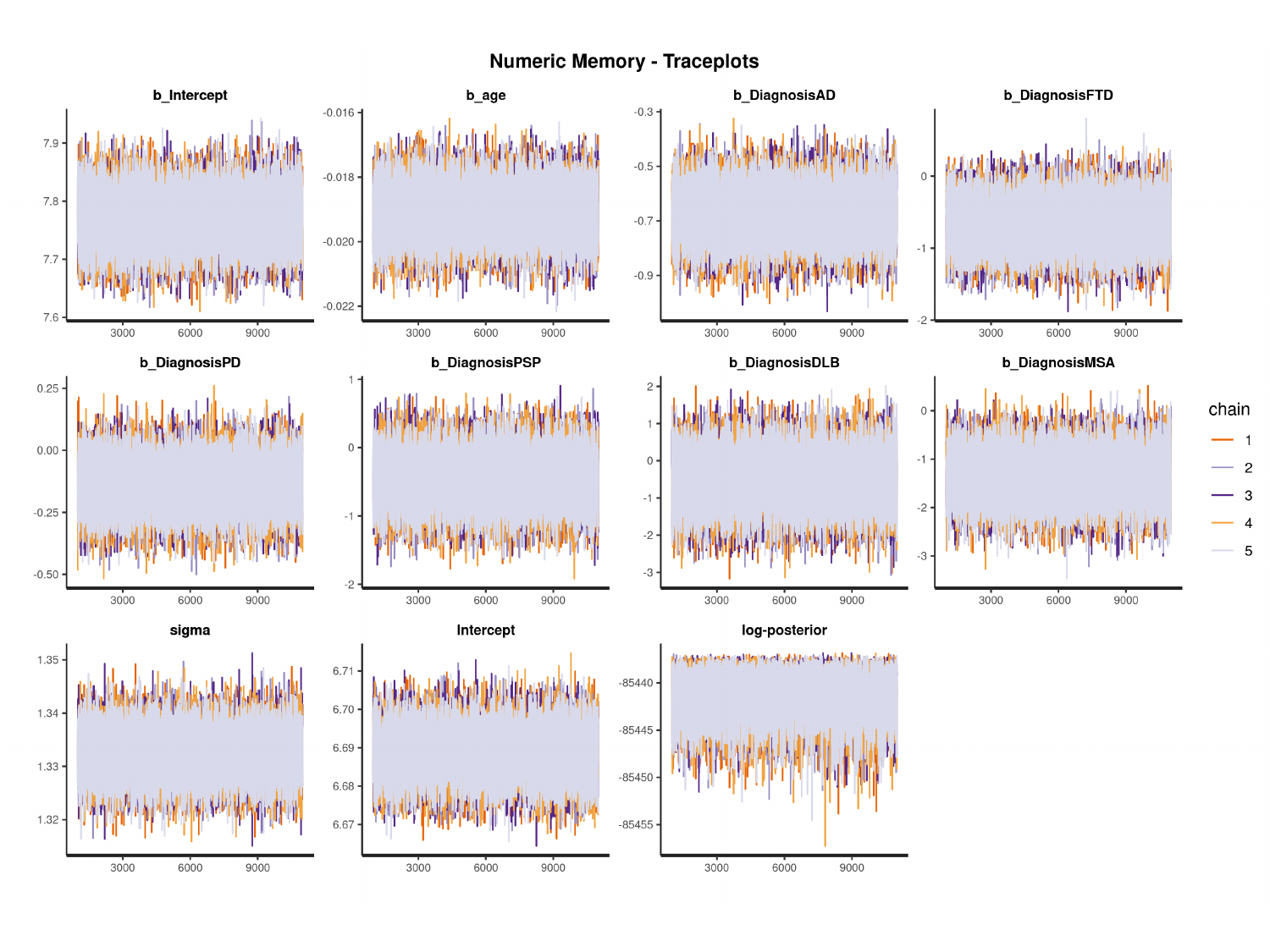

#

### Slide 26
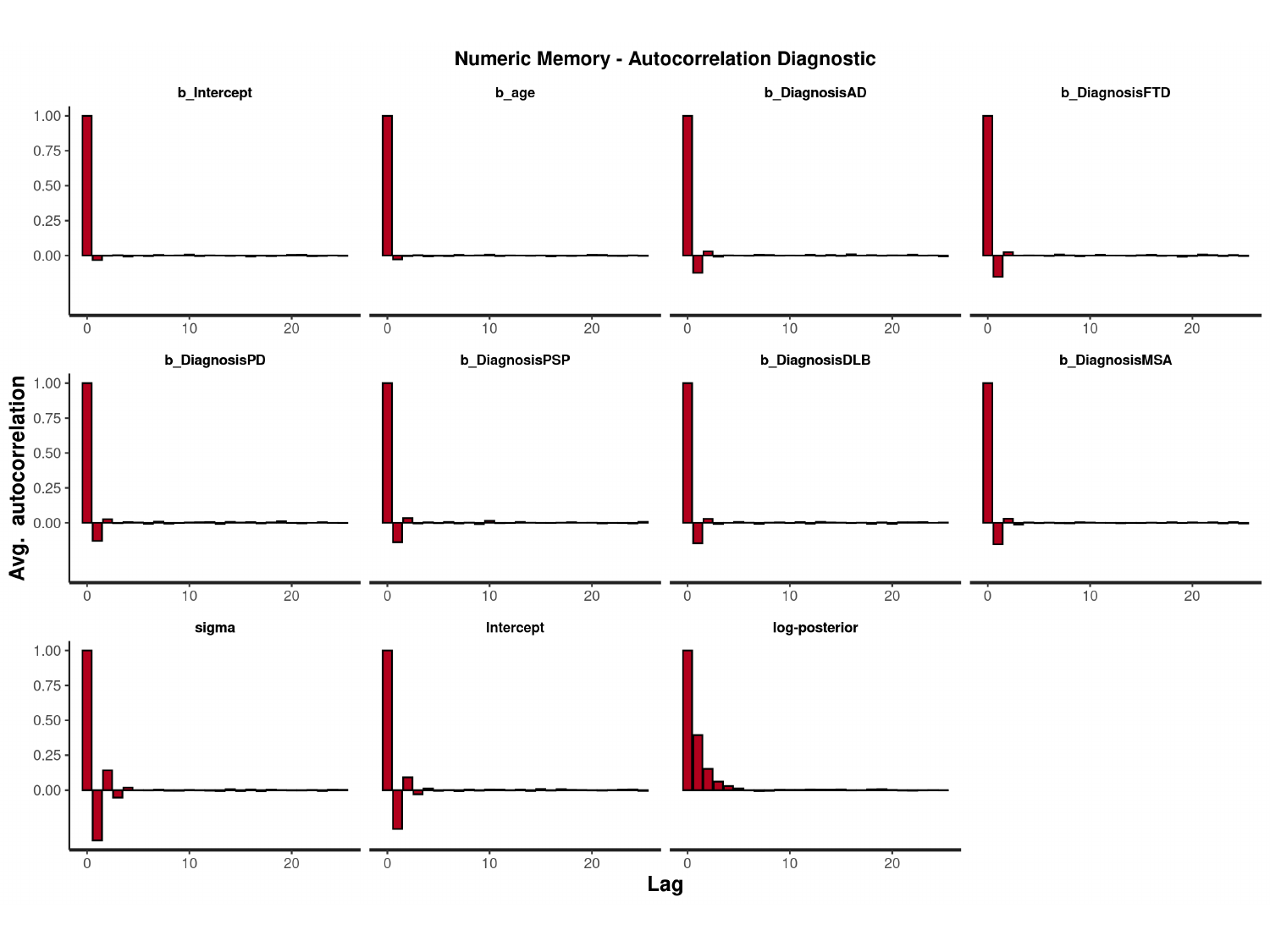

#

### Slide 27
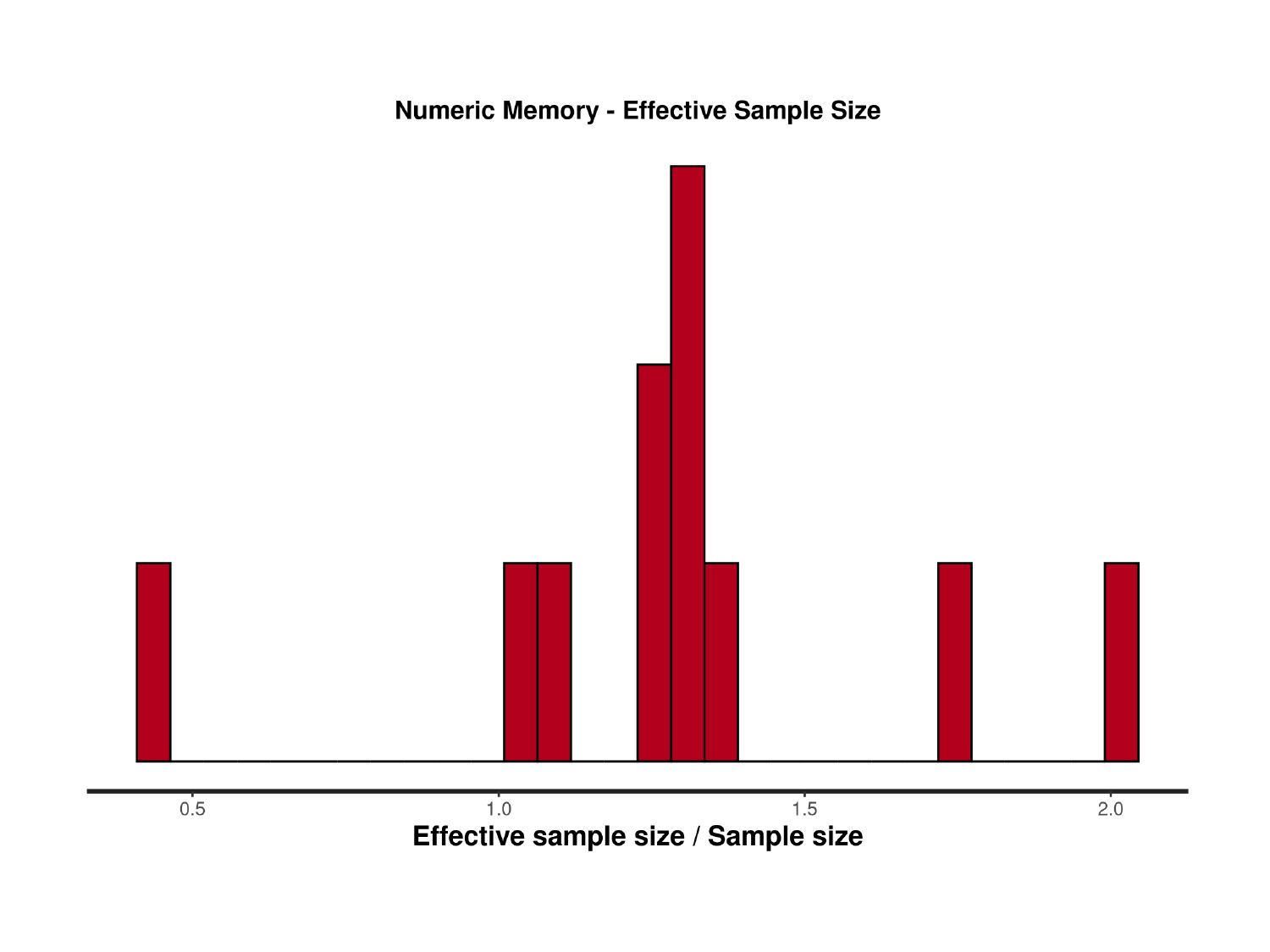

### Slide 28
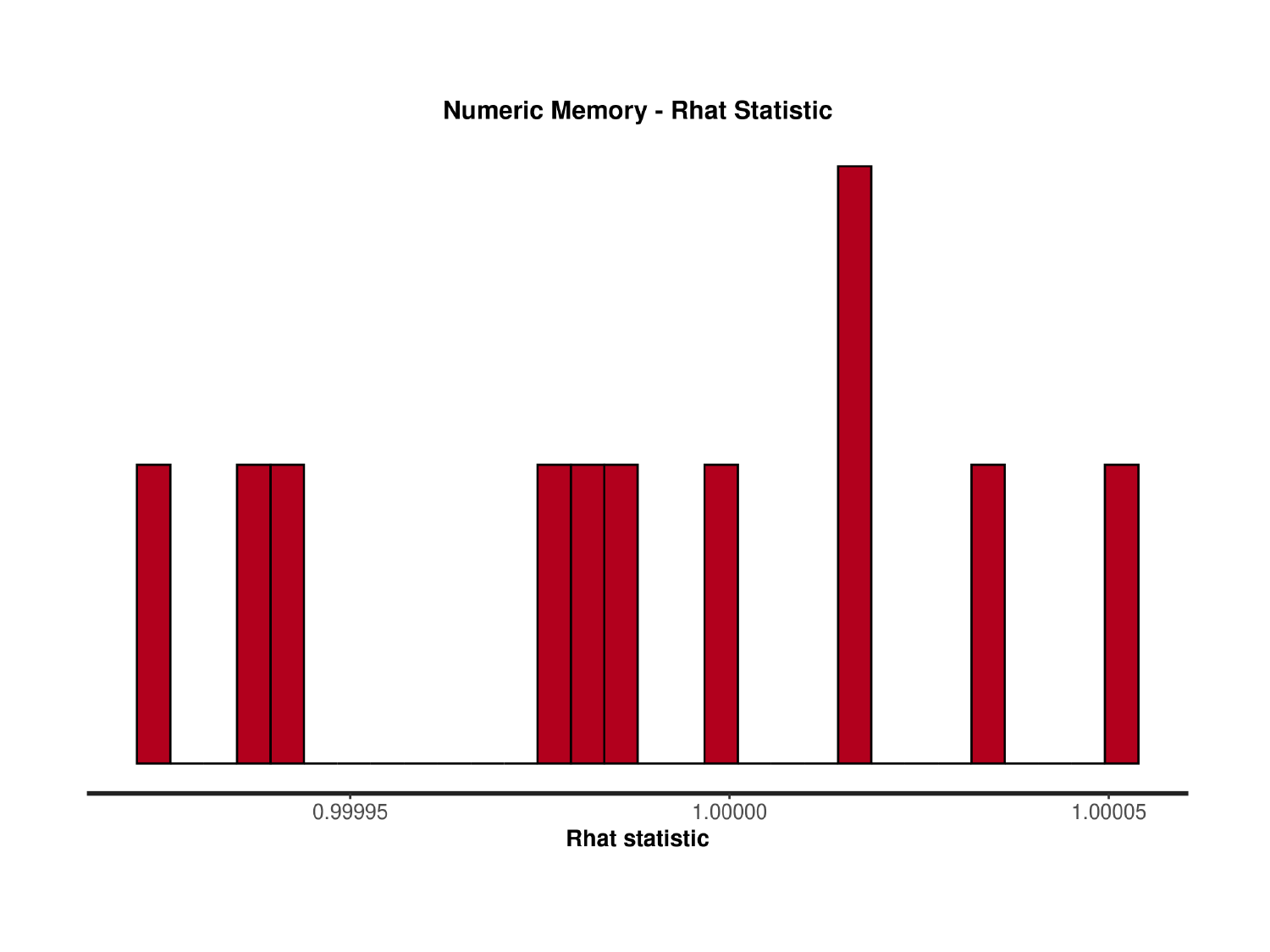

### Slide 29
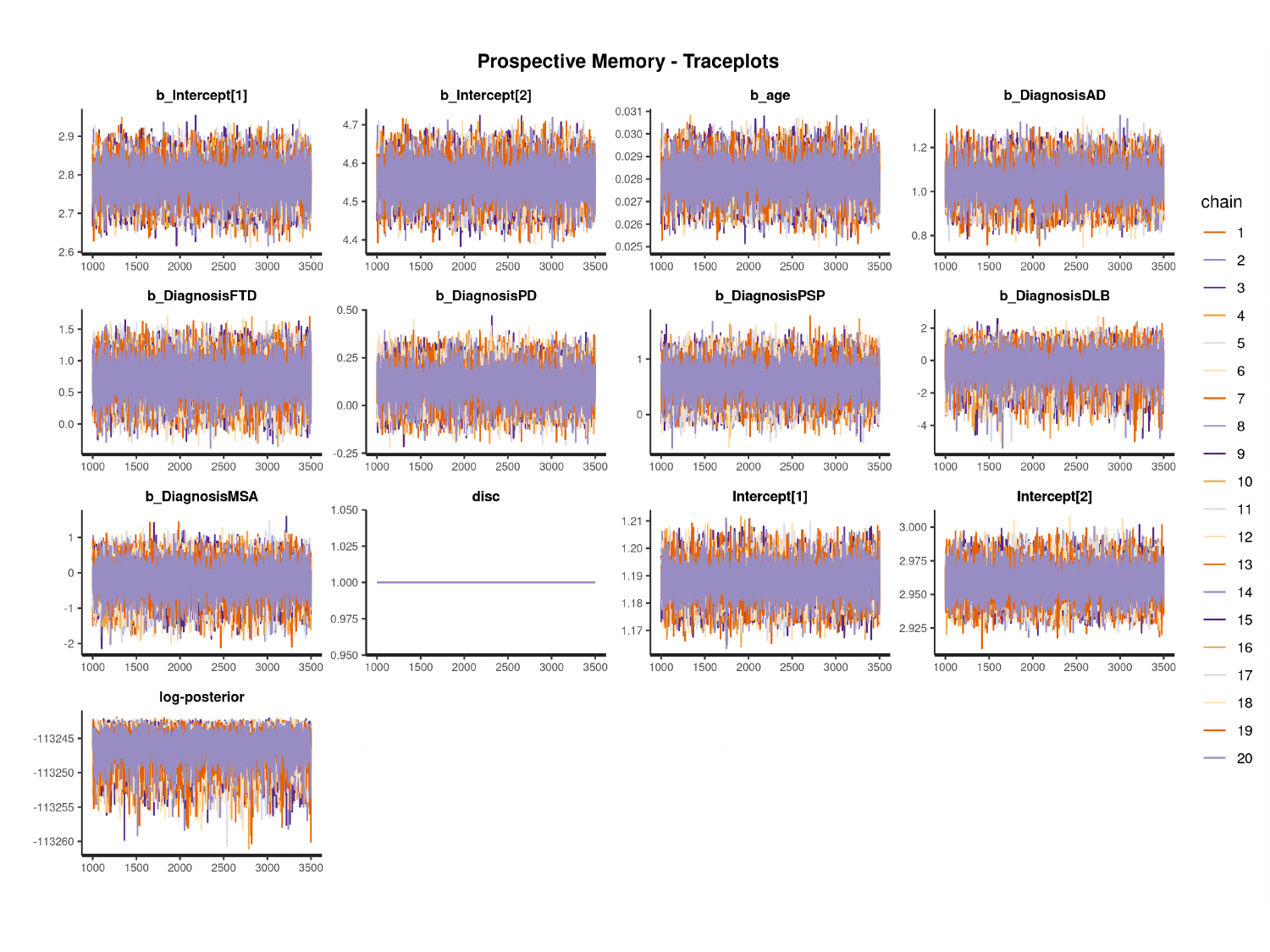

#

### Slide 30
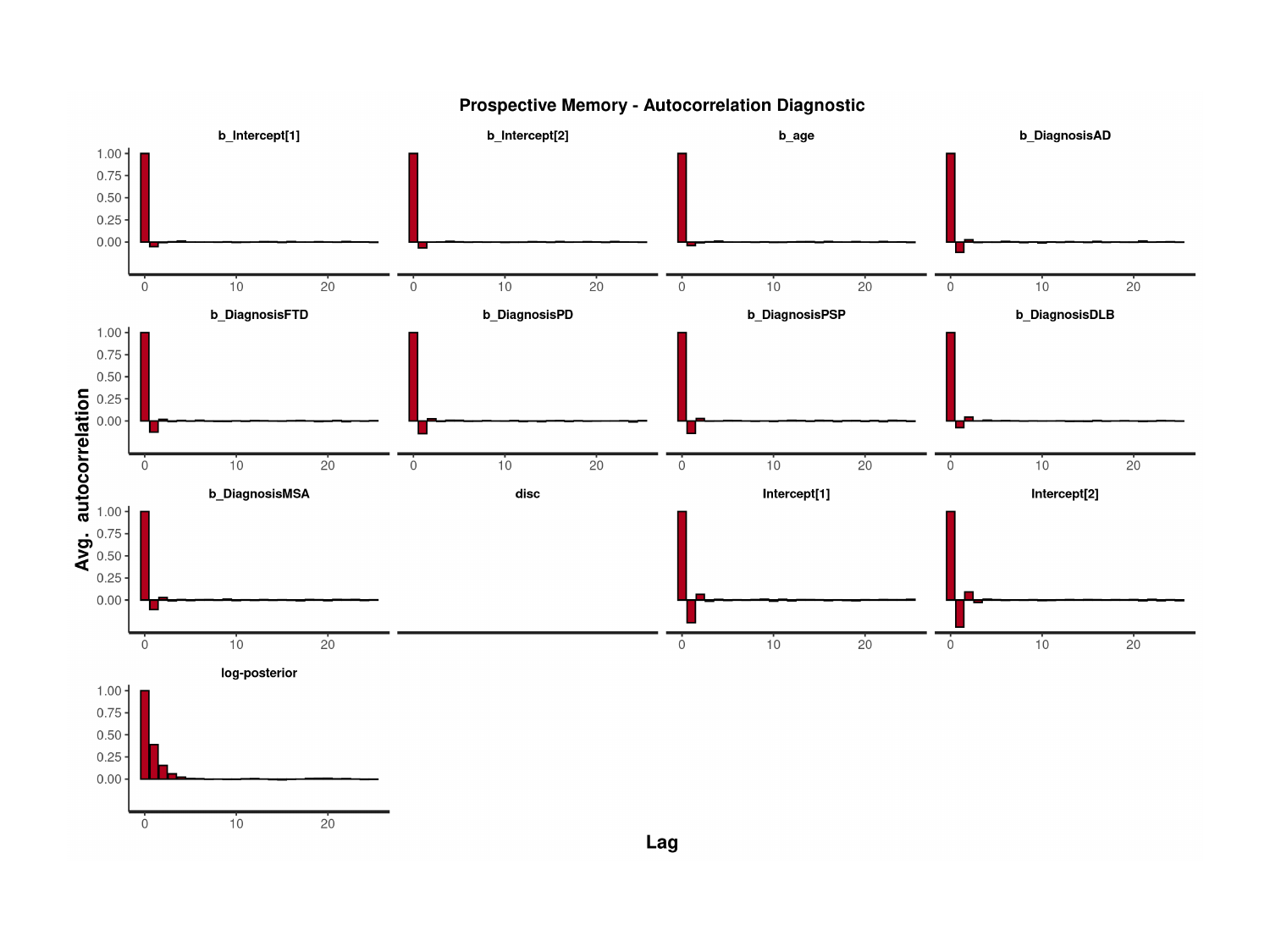

### Slide 31
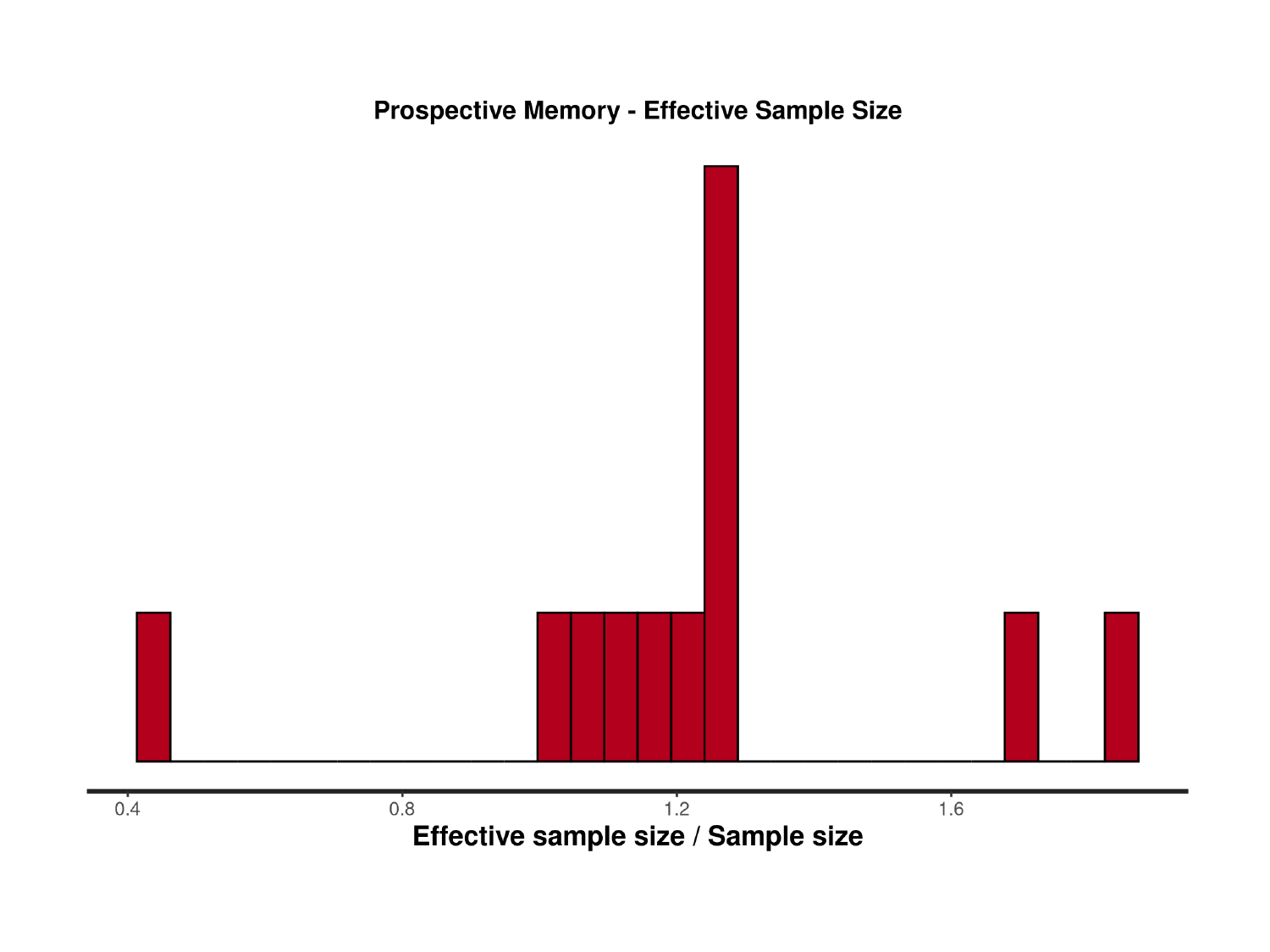

### Slide 32
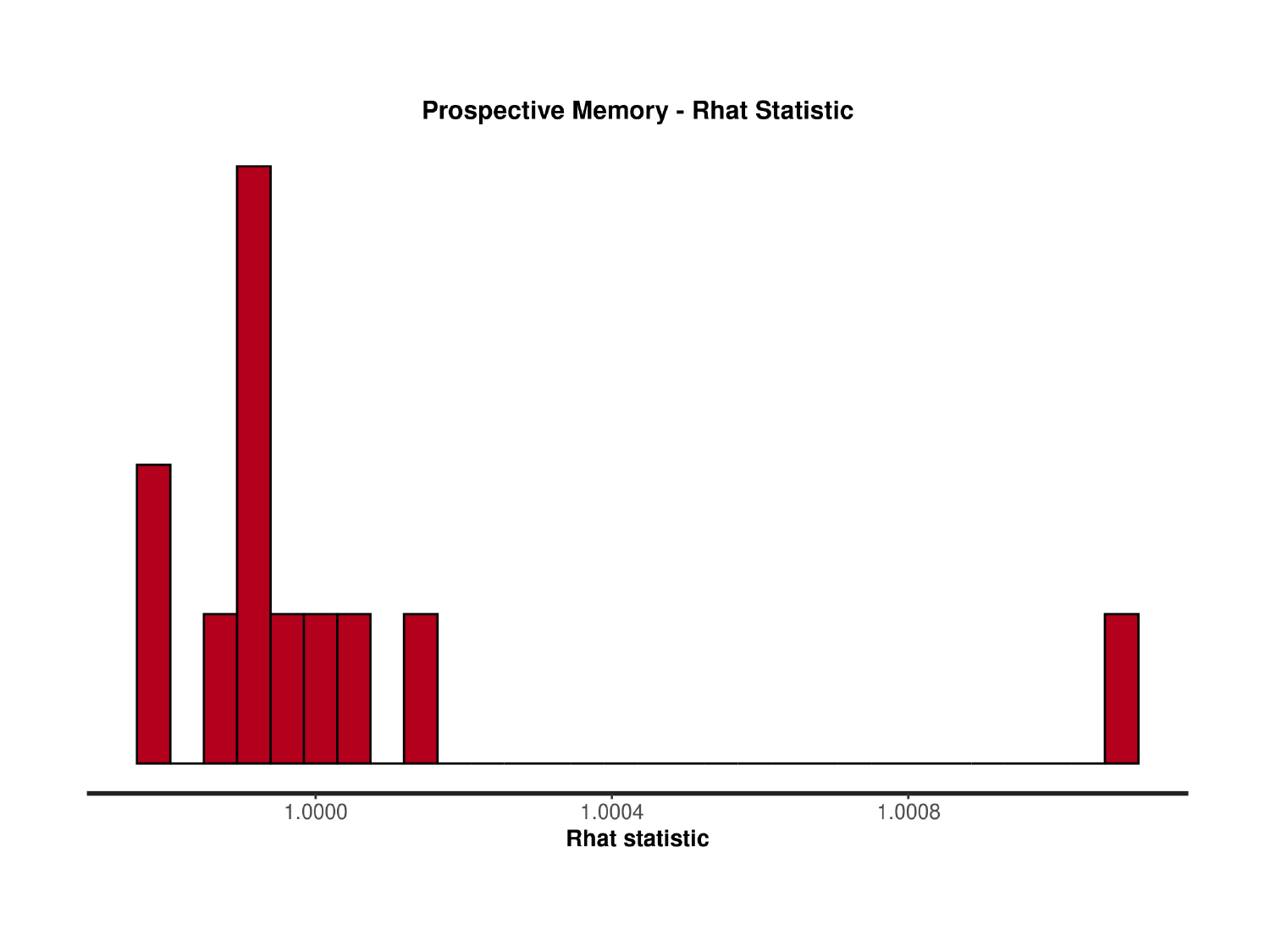

### Slide 33
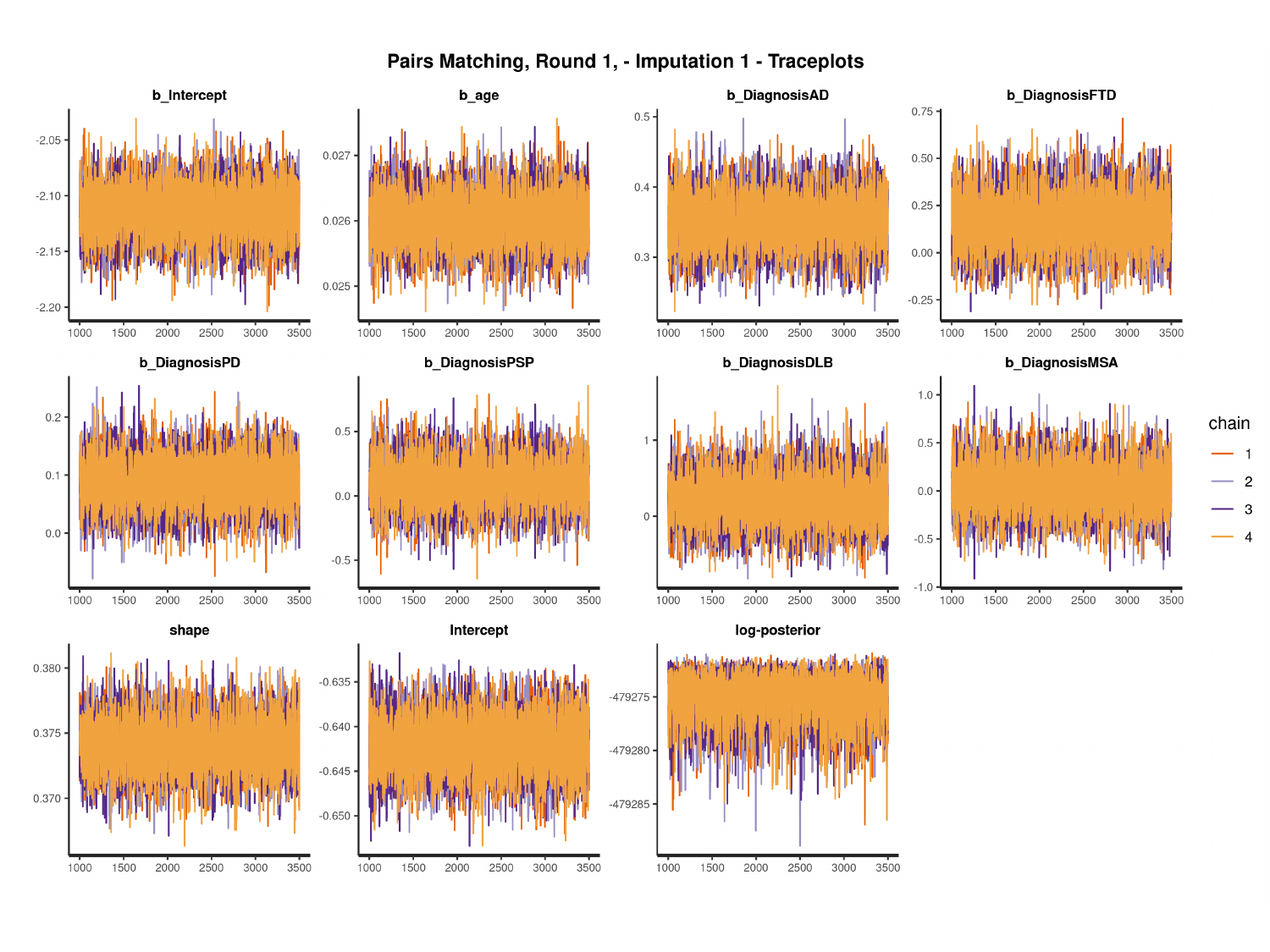

#

### Slide 34
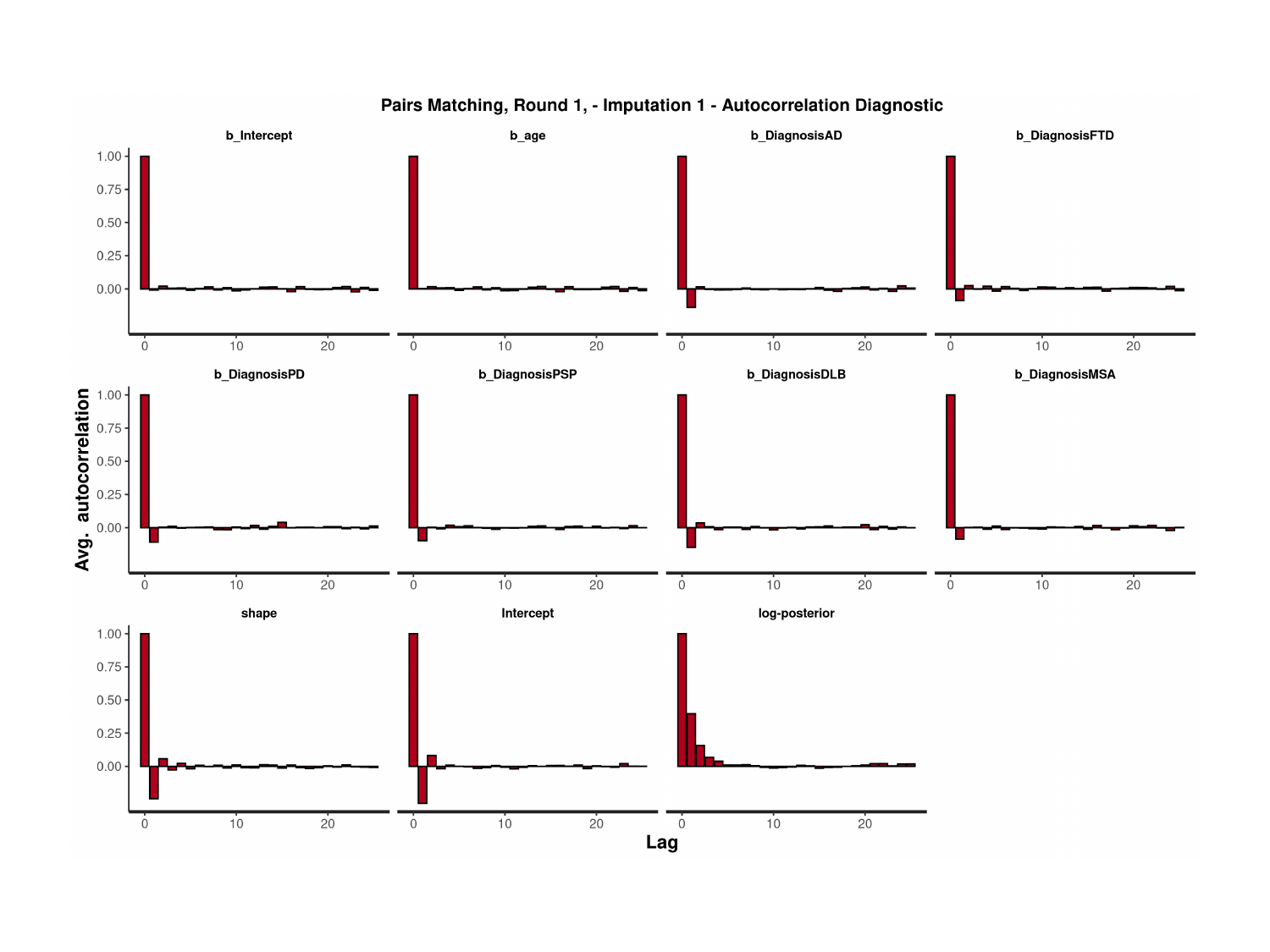

### Slide 35
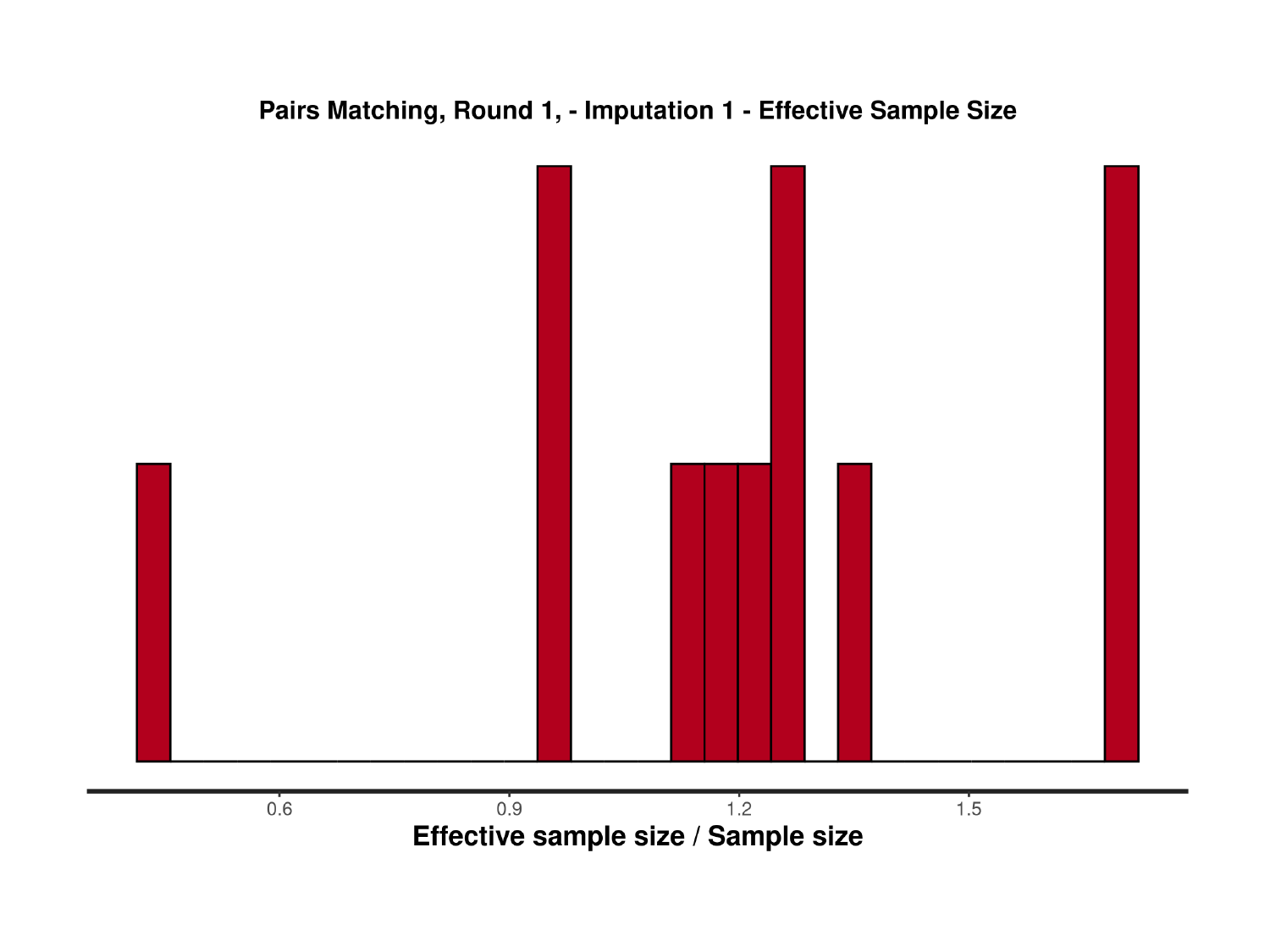

### Slide 36
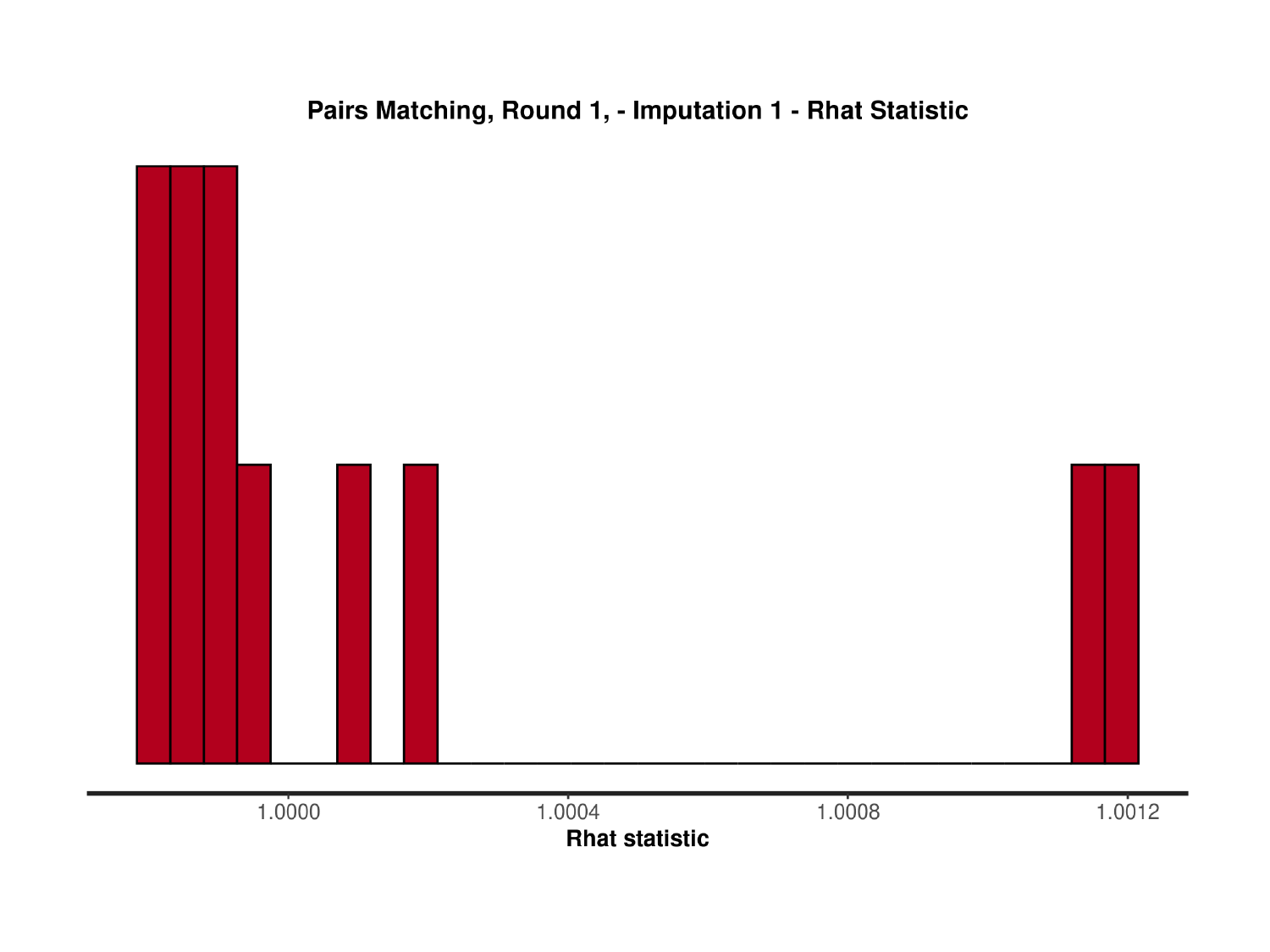

### Slide 37
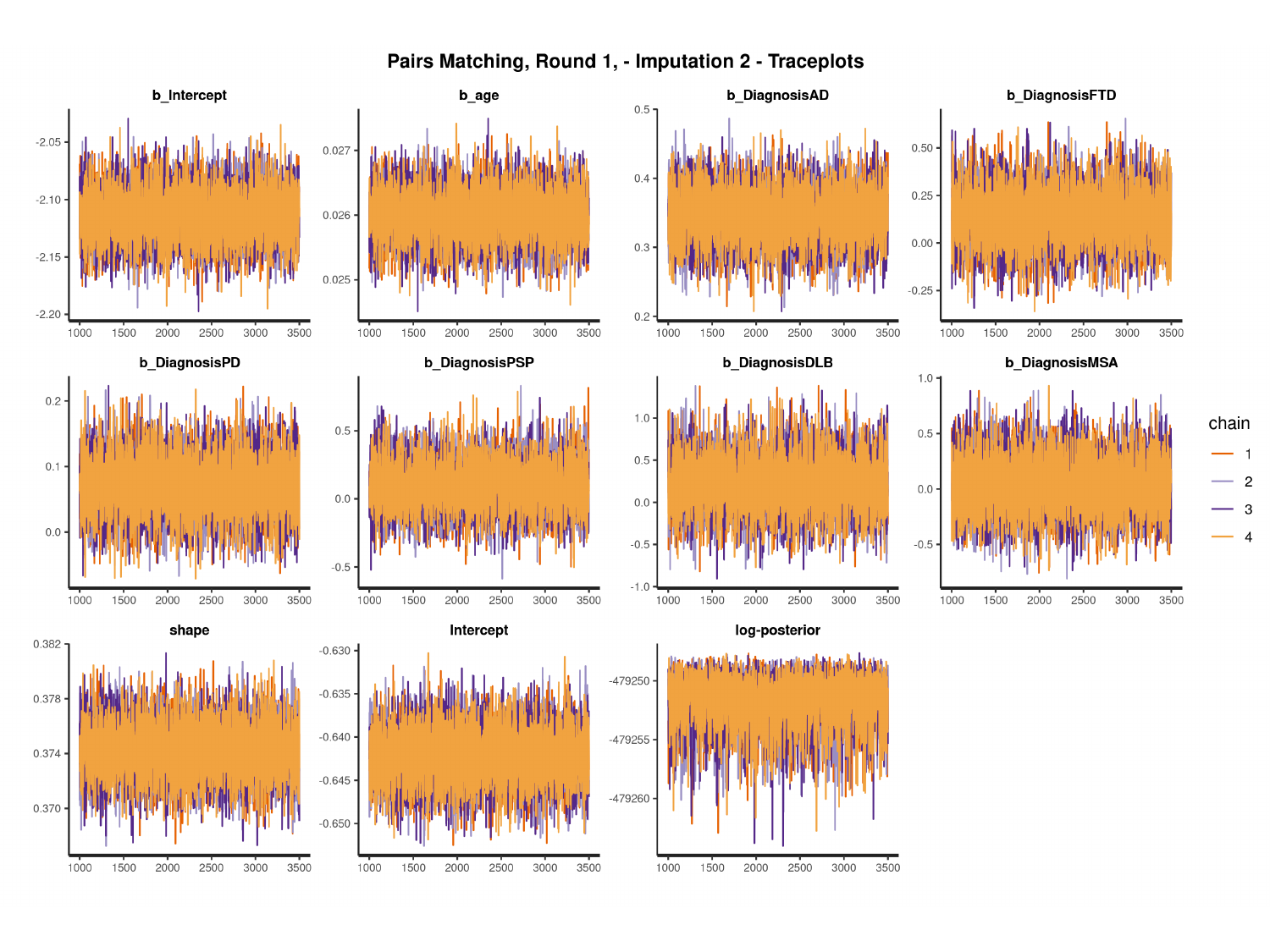

#

### Slide 38
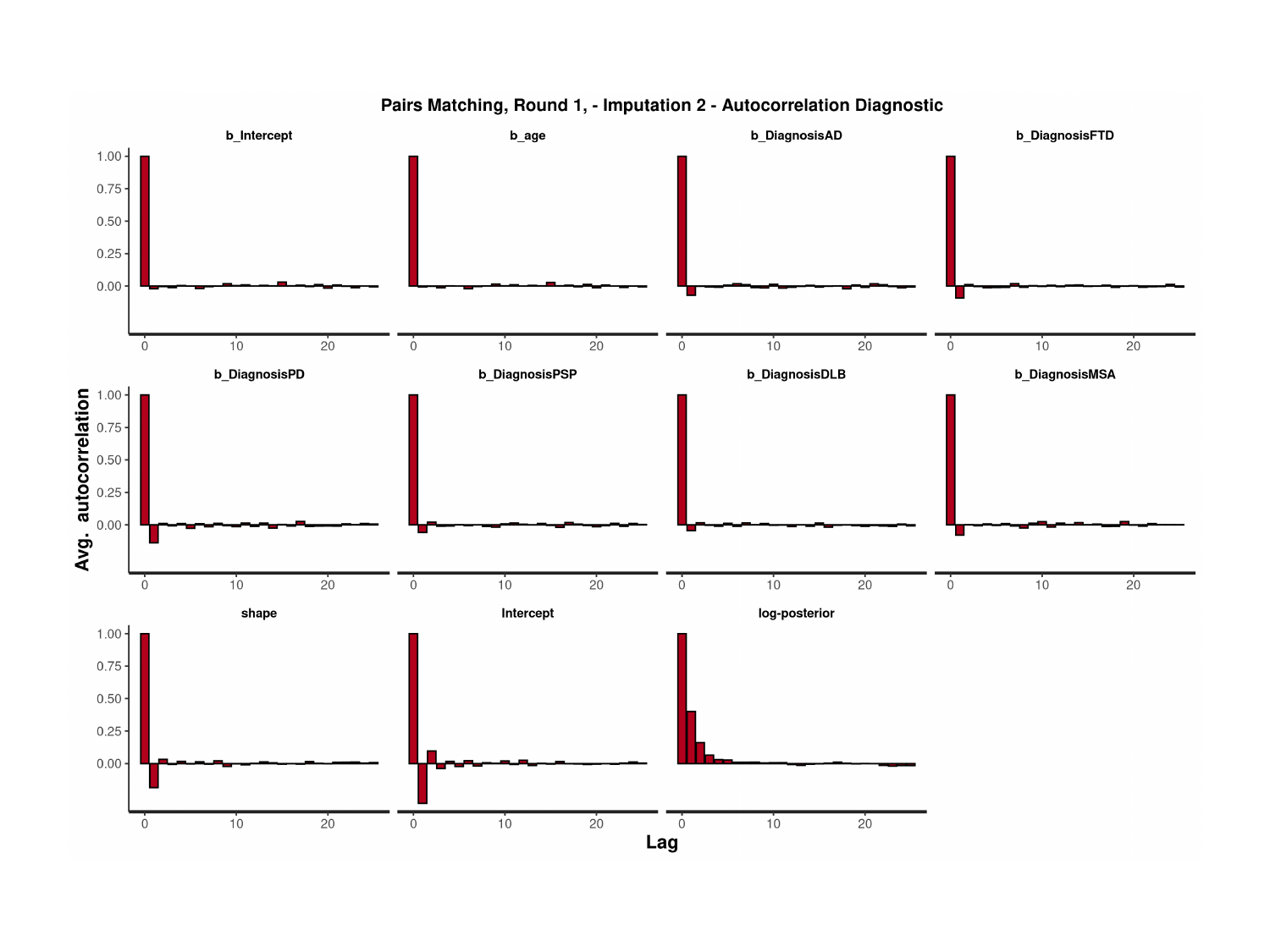

### Slide 39
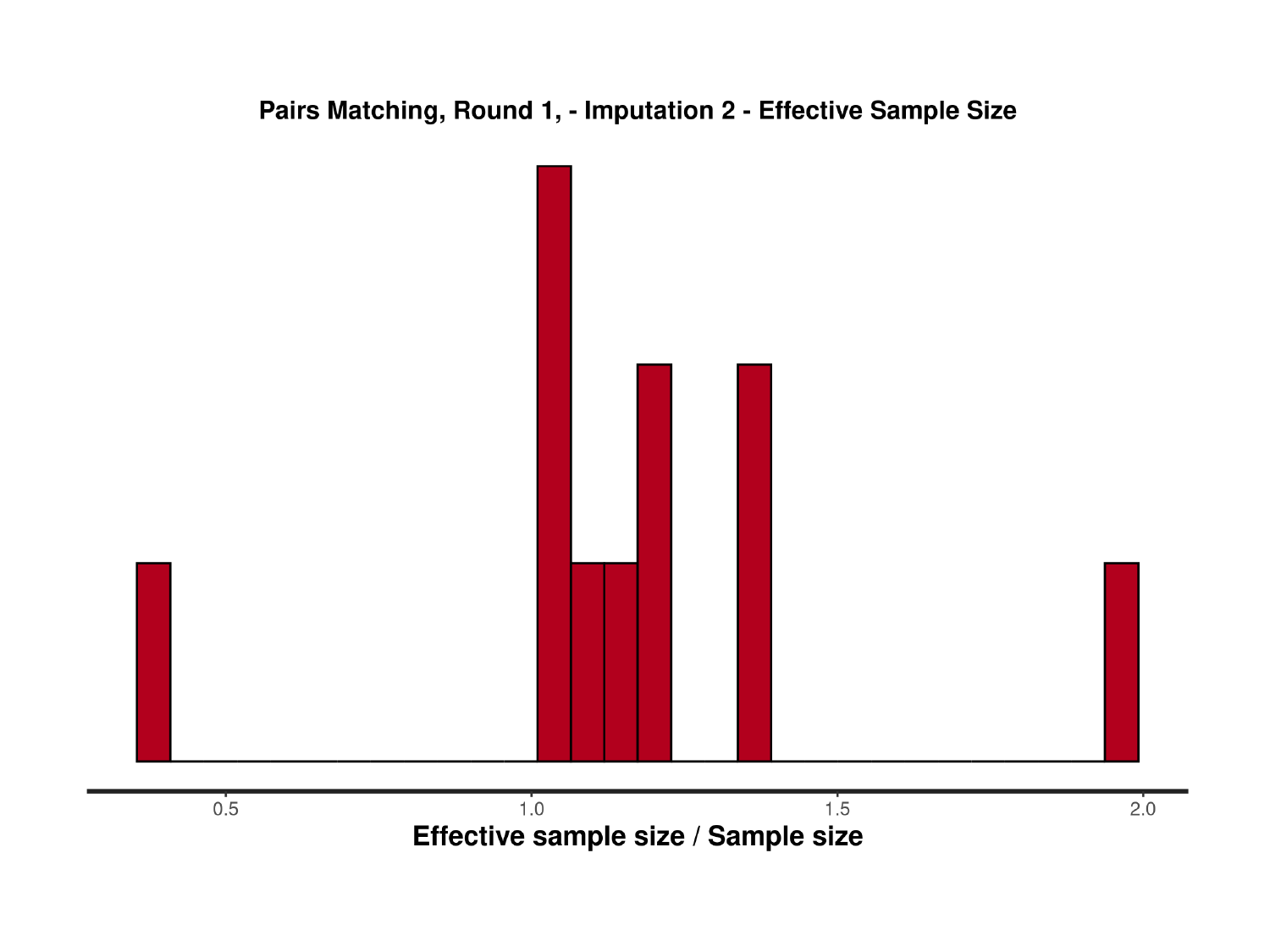

### Slide 40
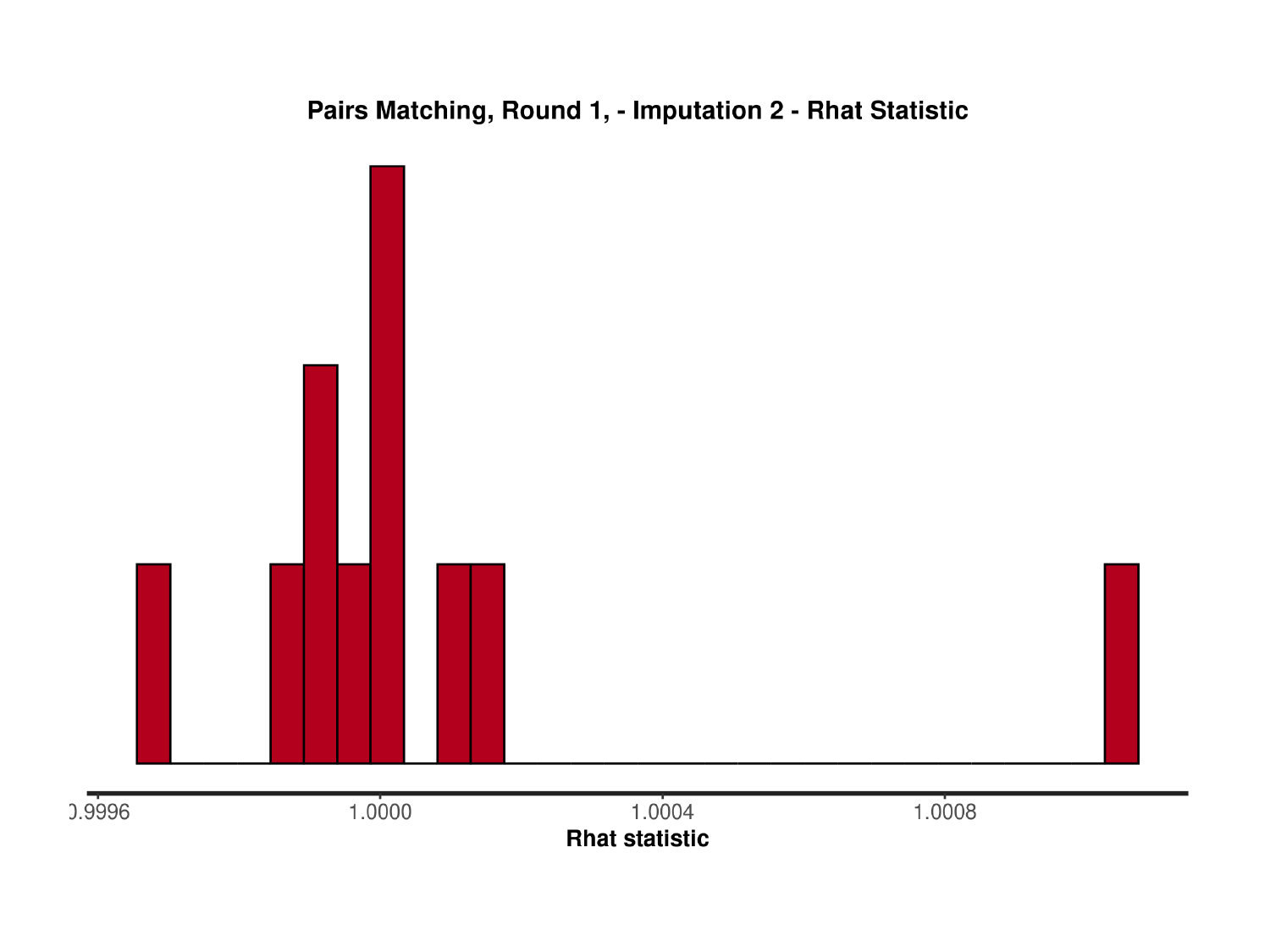

### Slide 41
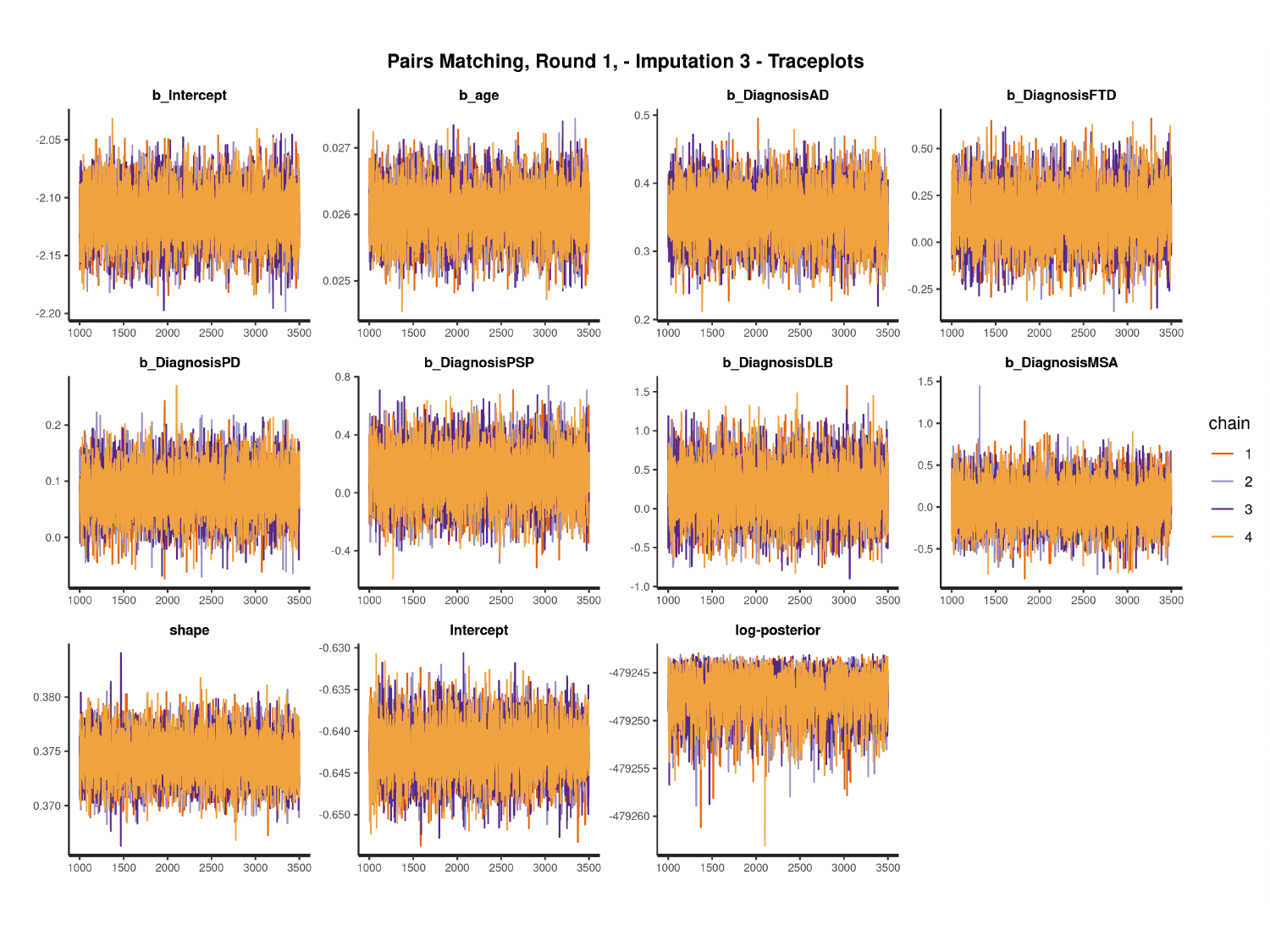

#

### Slide 42
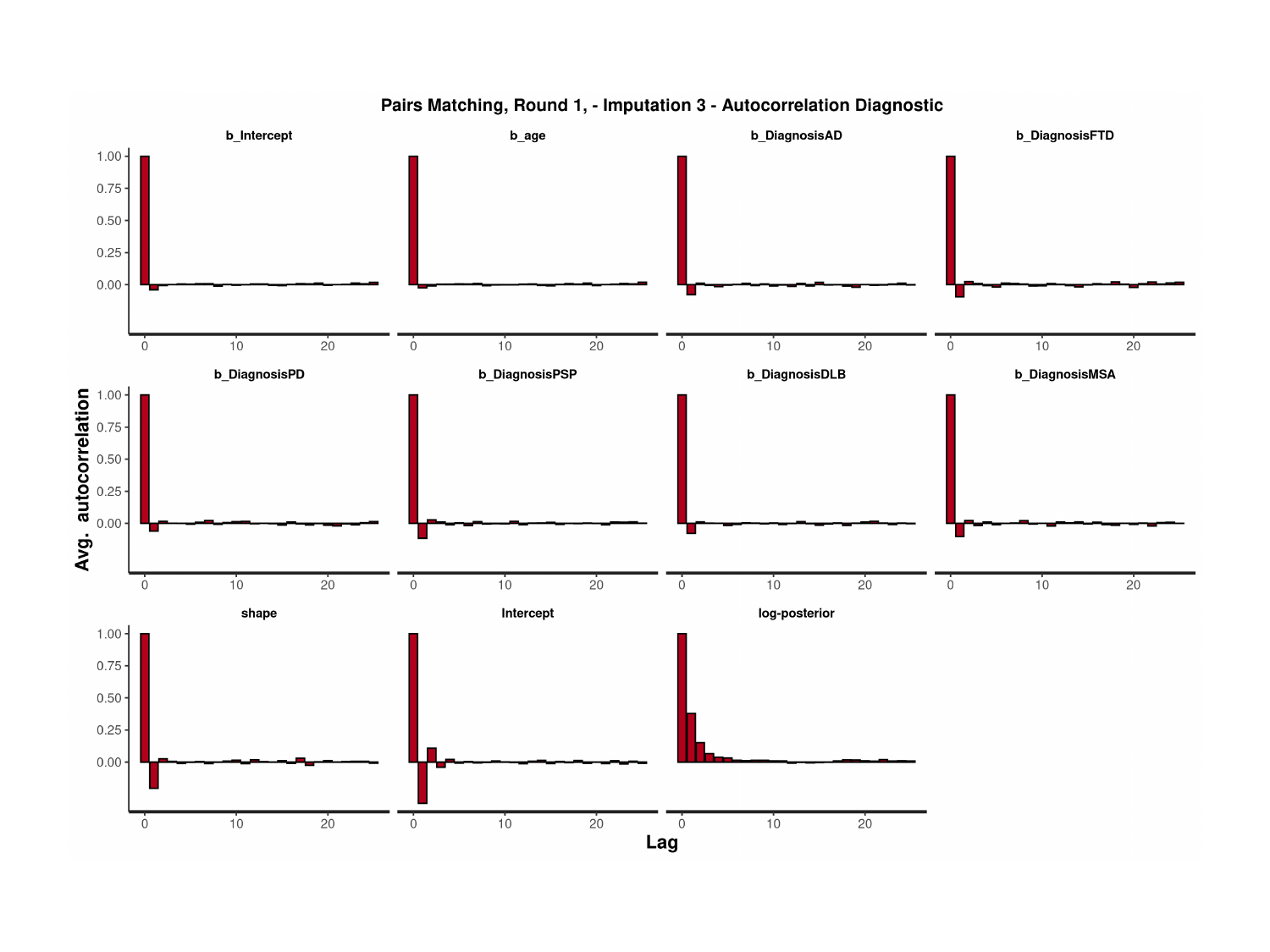

### Slide 43
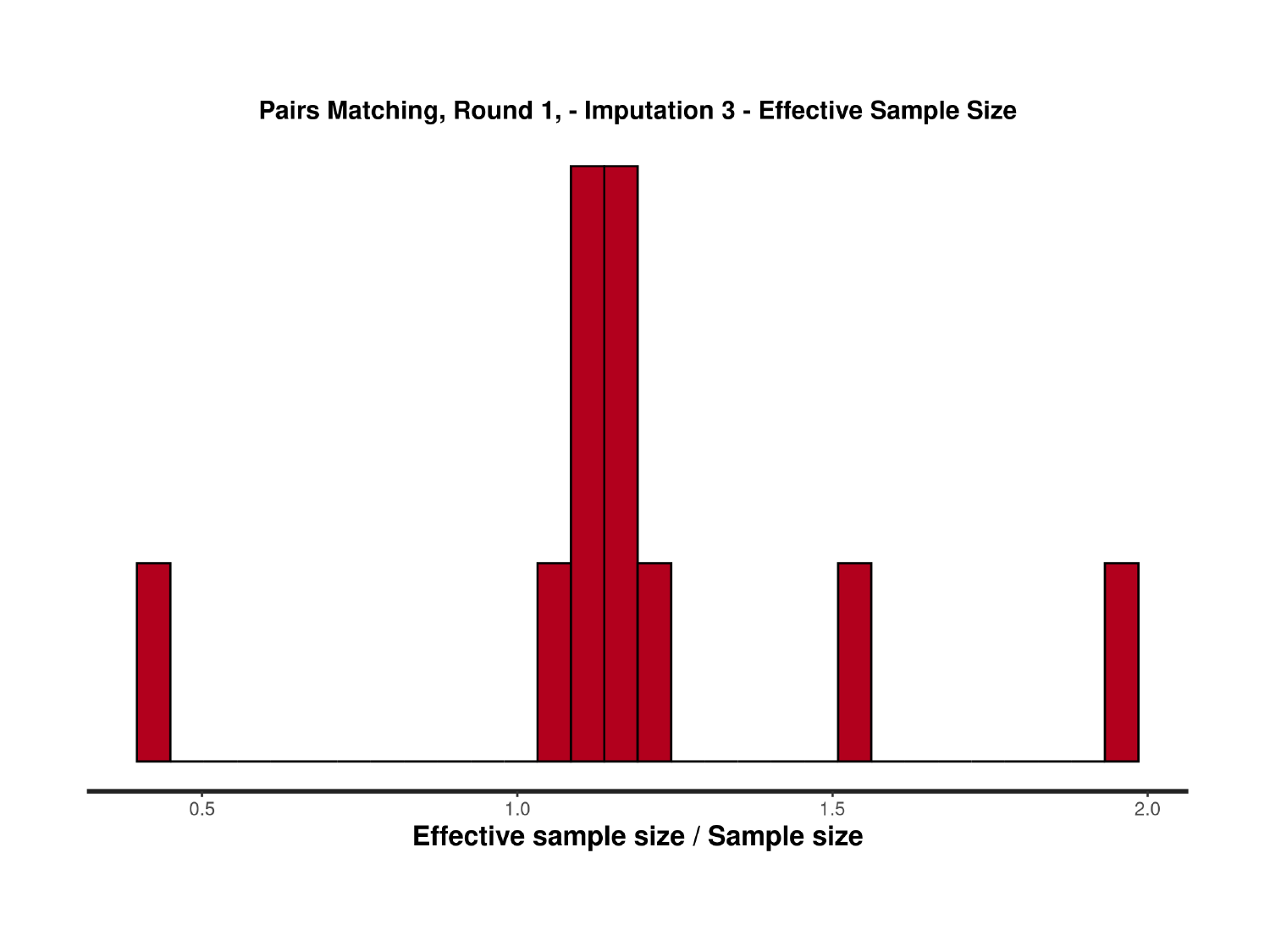

### Slide 44
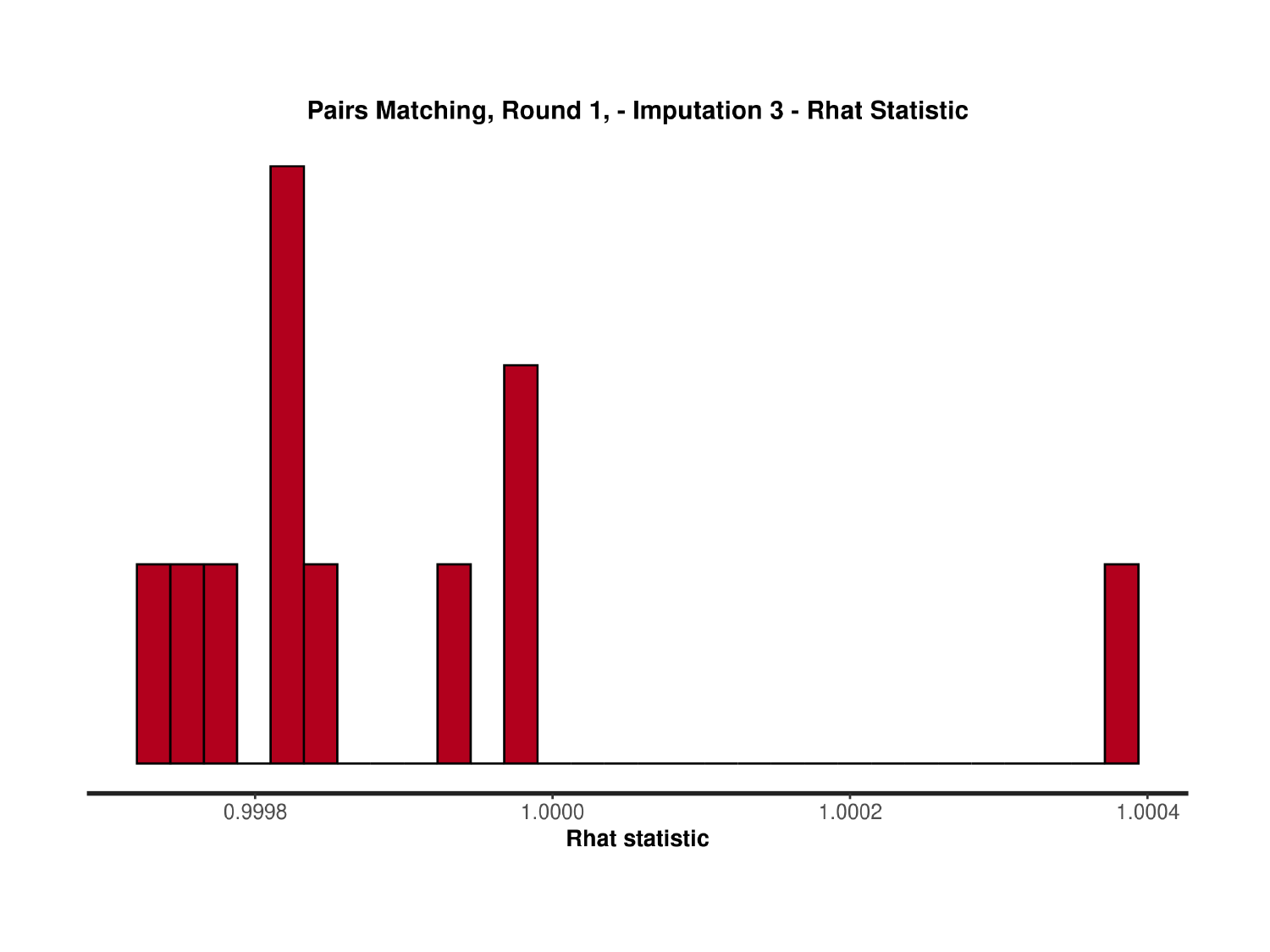

### Slide 45
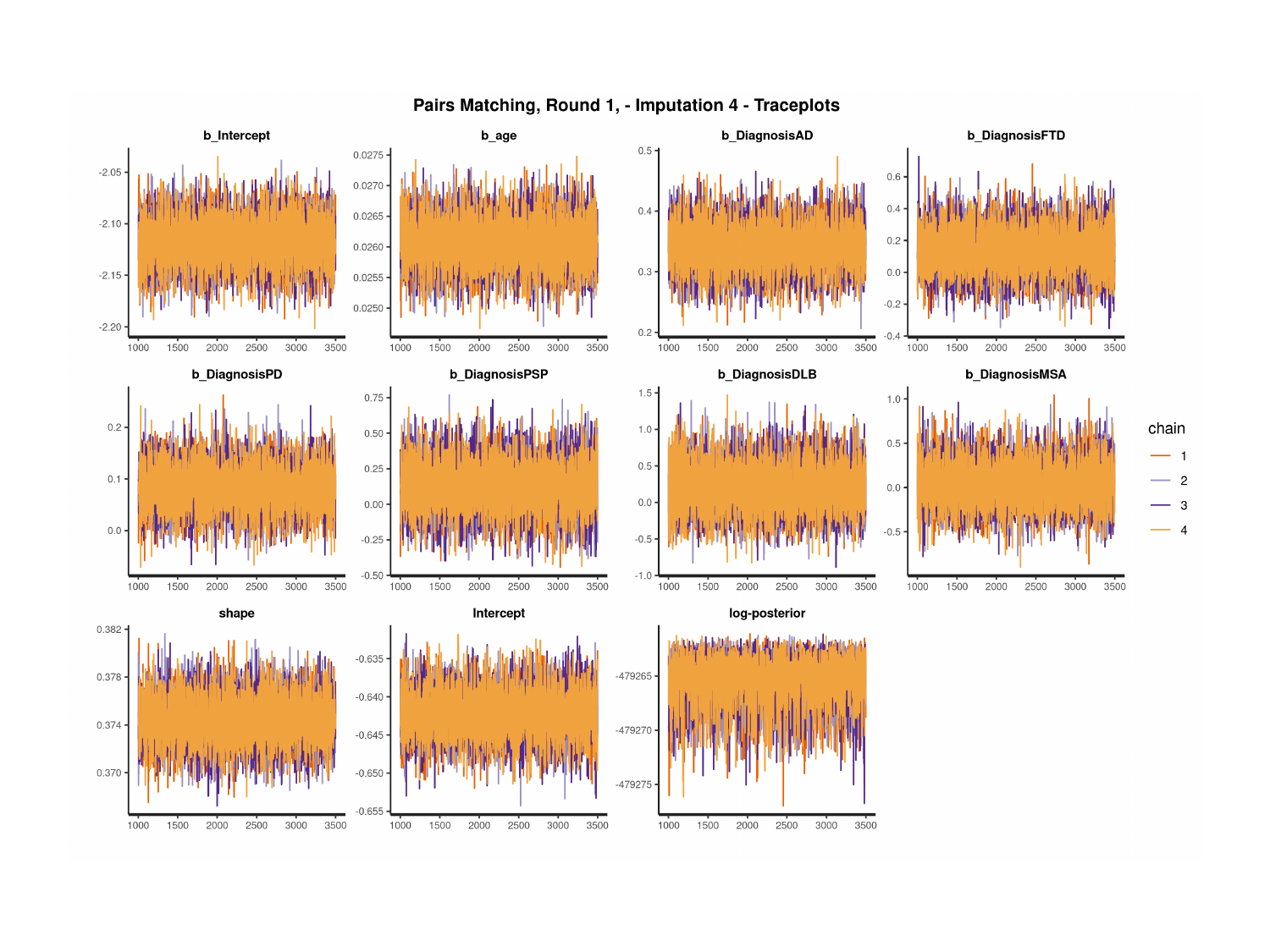

### Slide 46
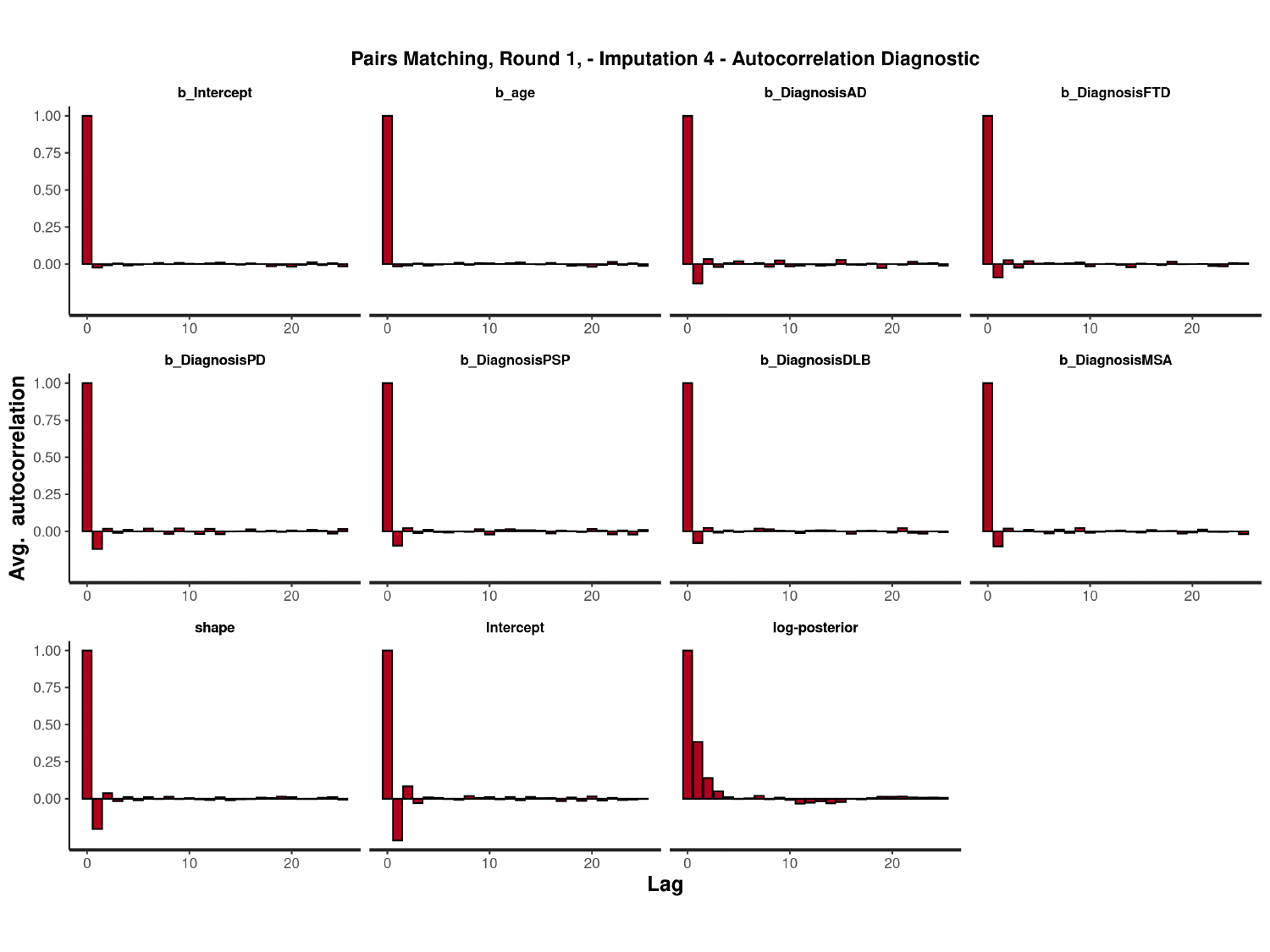

#

### Slide 47
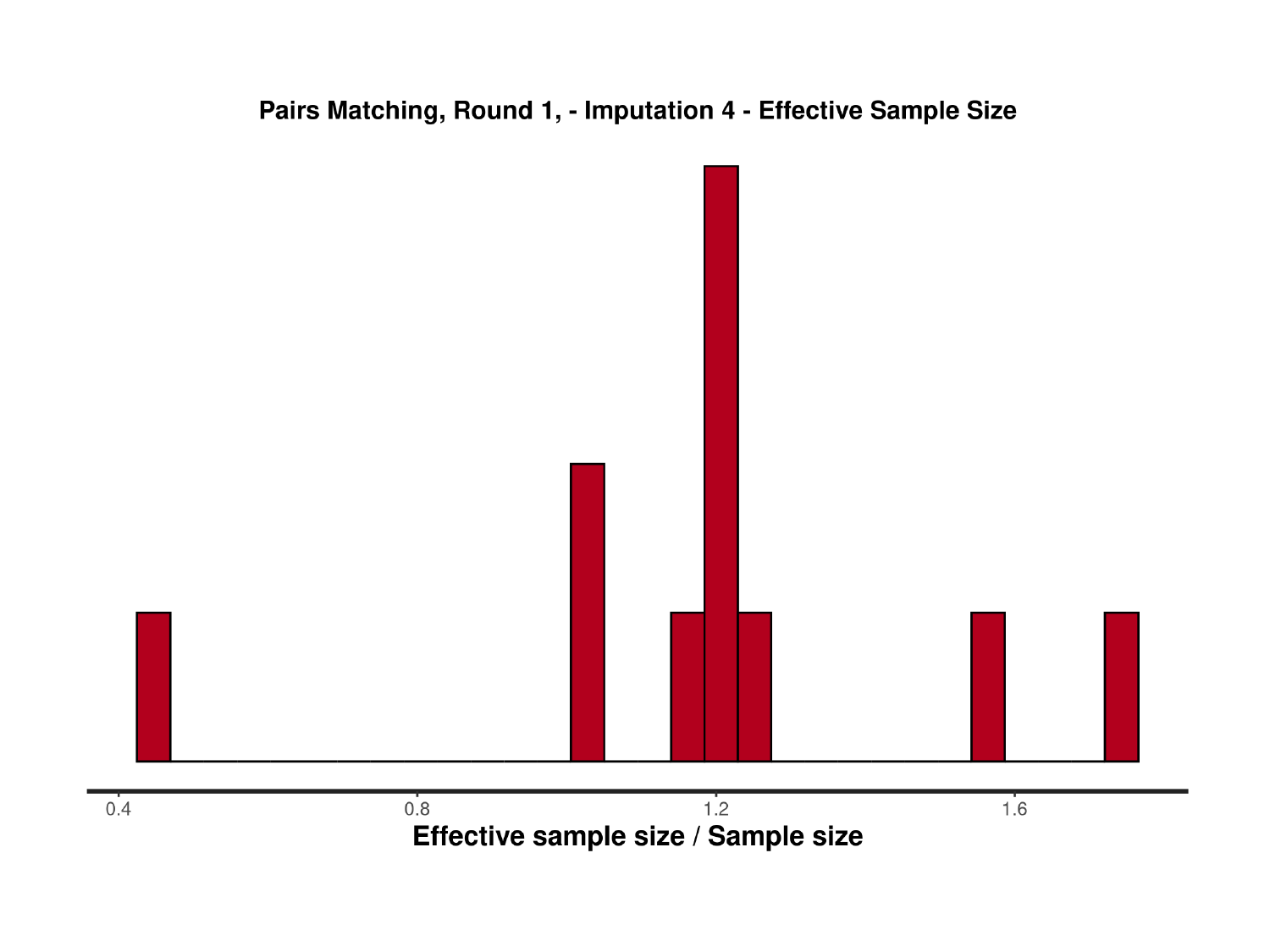

### Slide 48
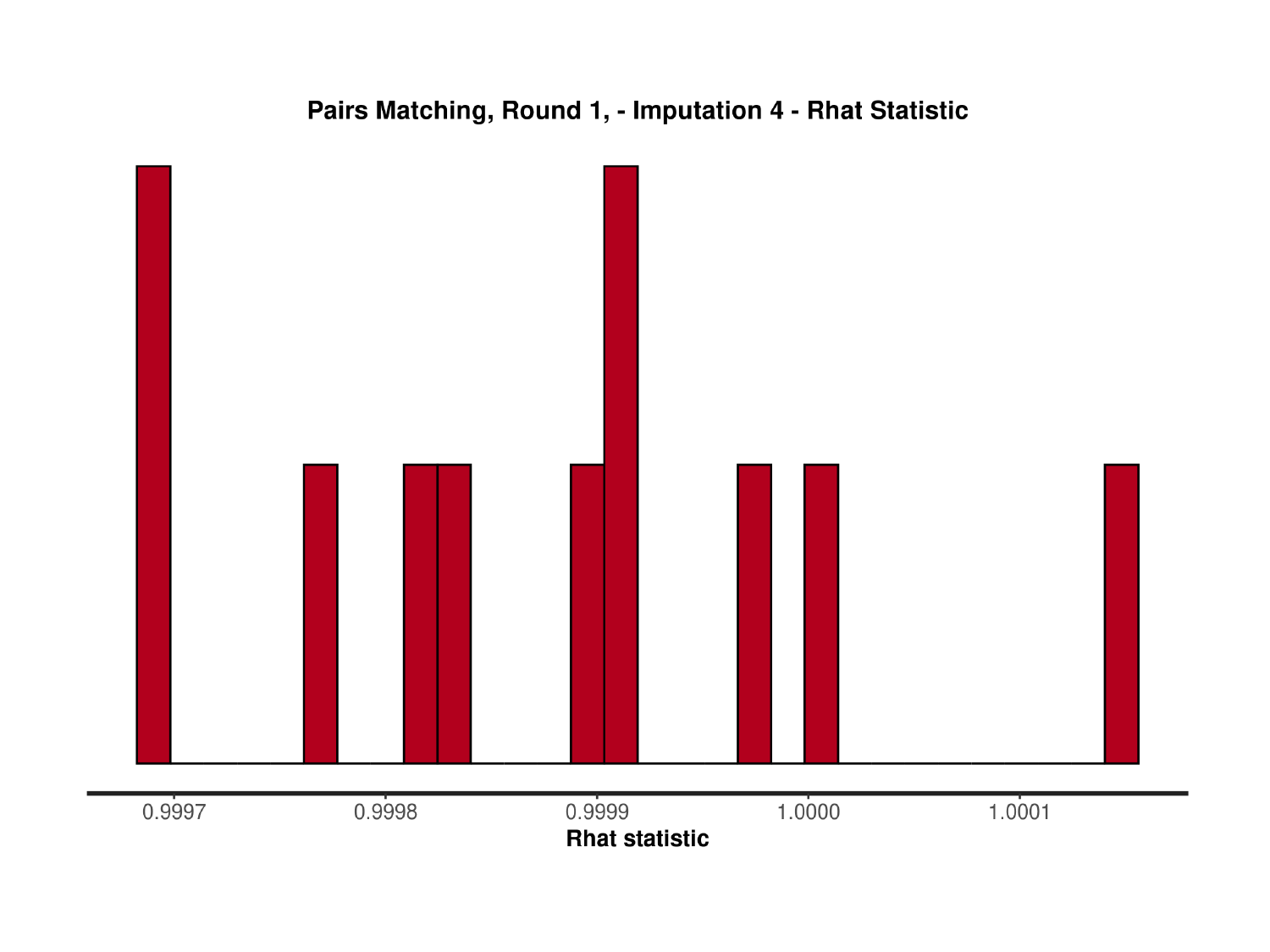

### Slide 49
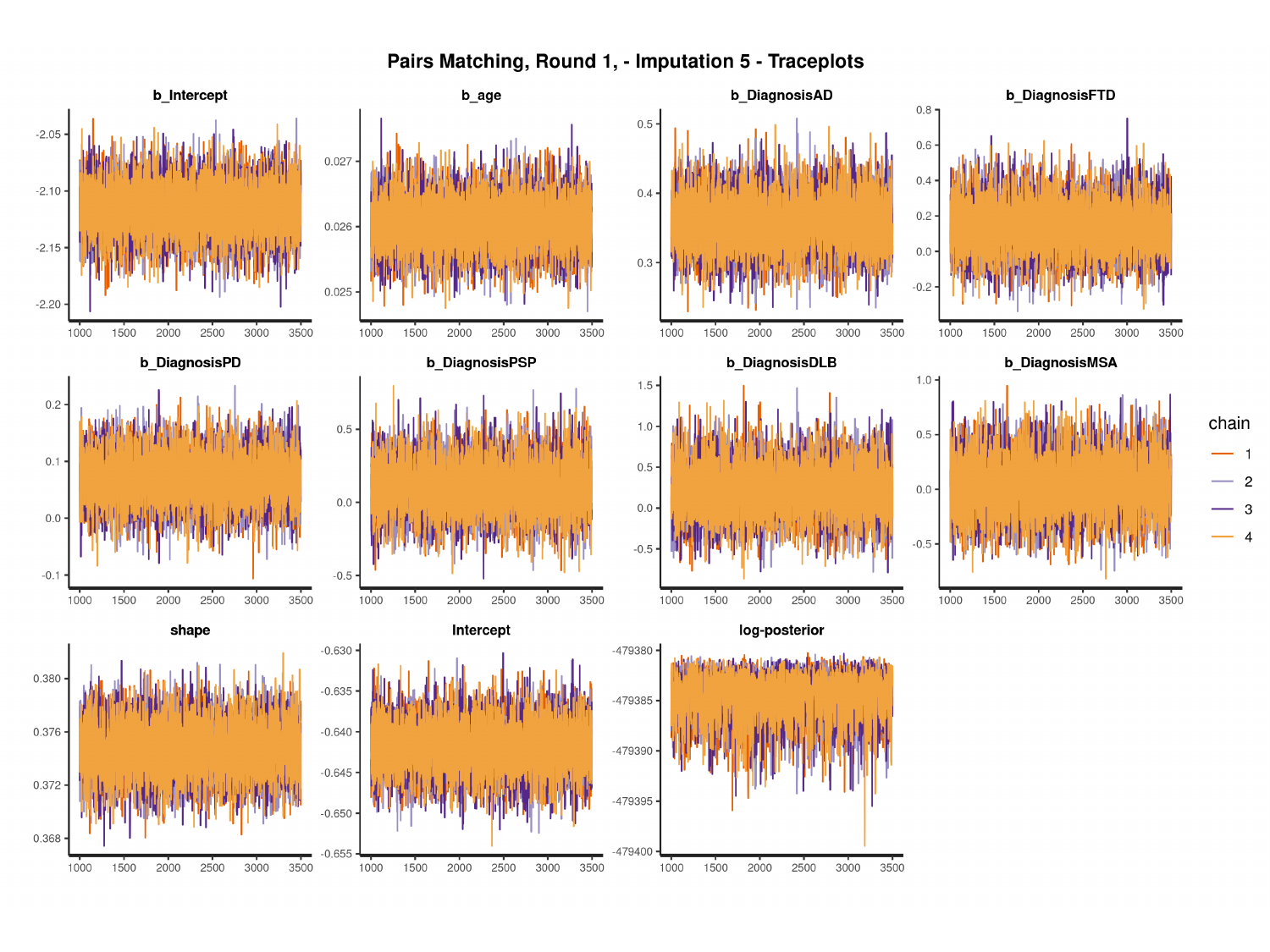

#

### Slide 50
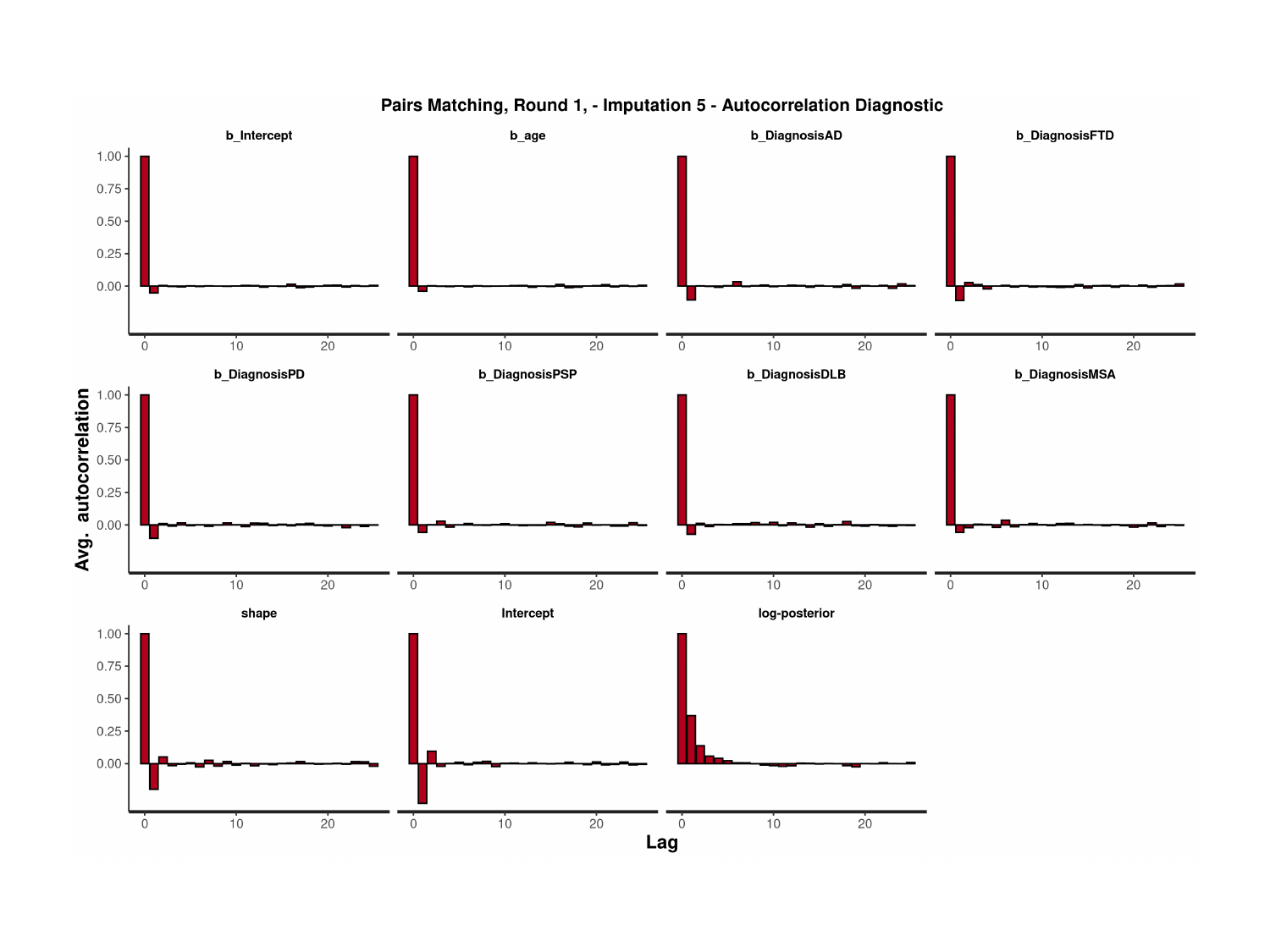

### Slide 51
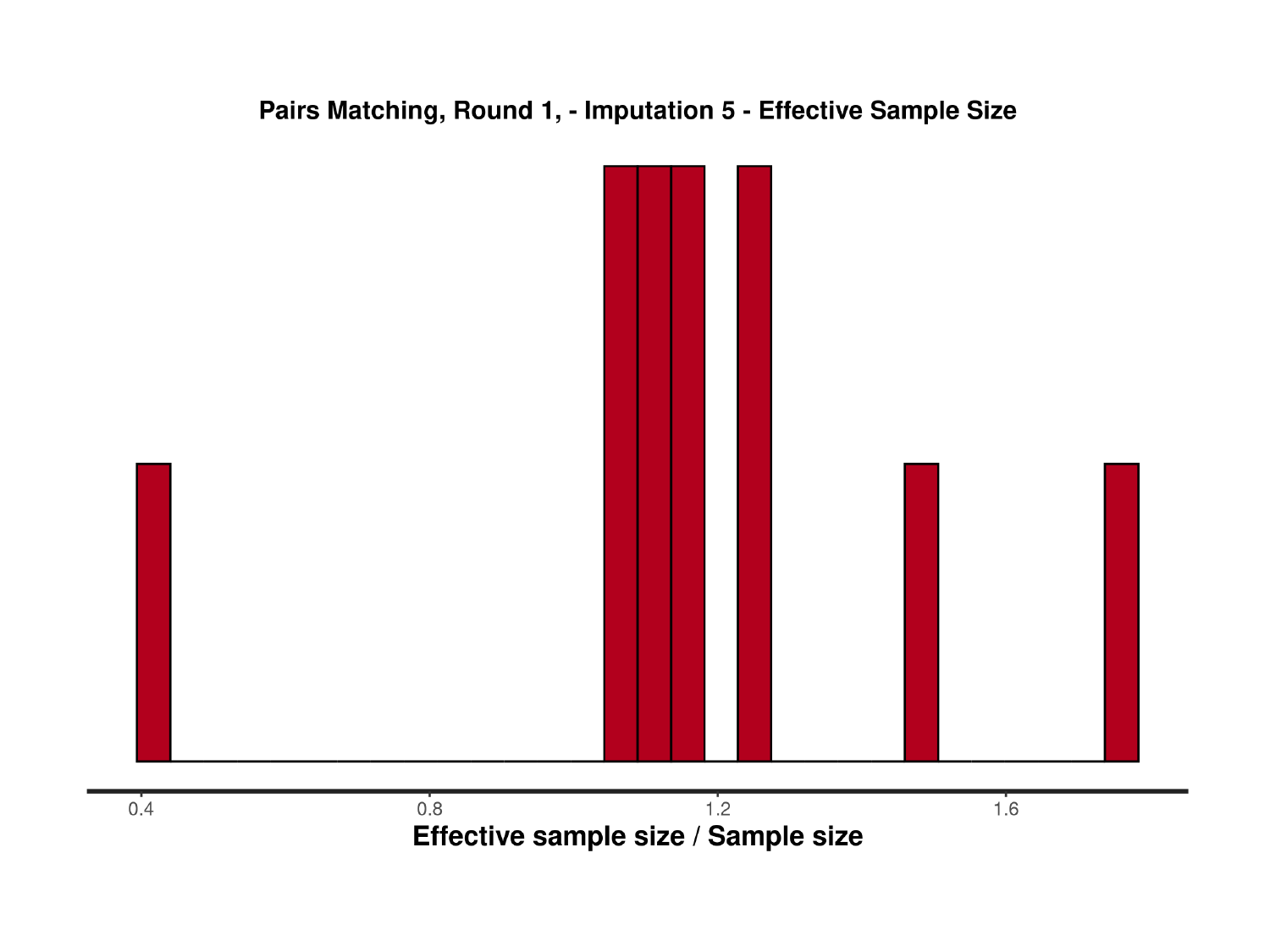

### Slide 52
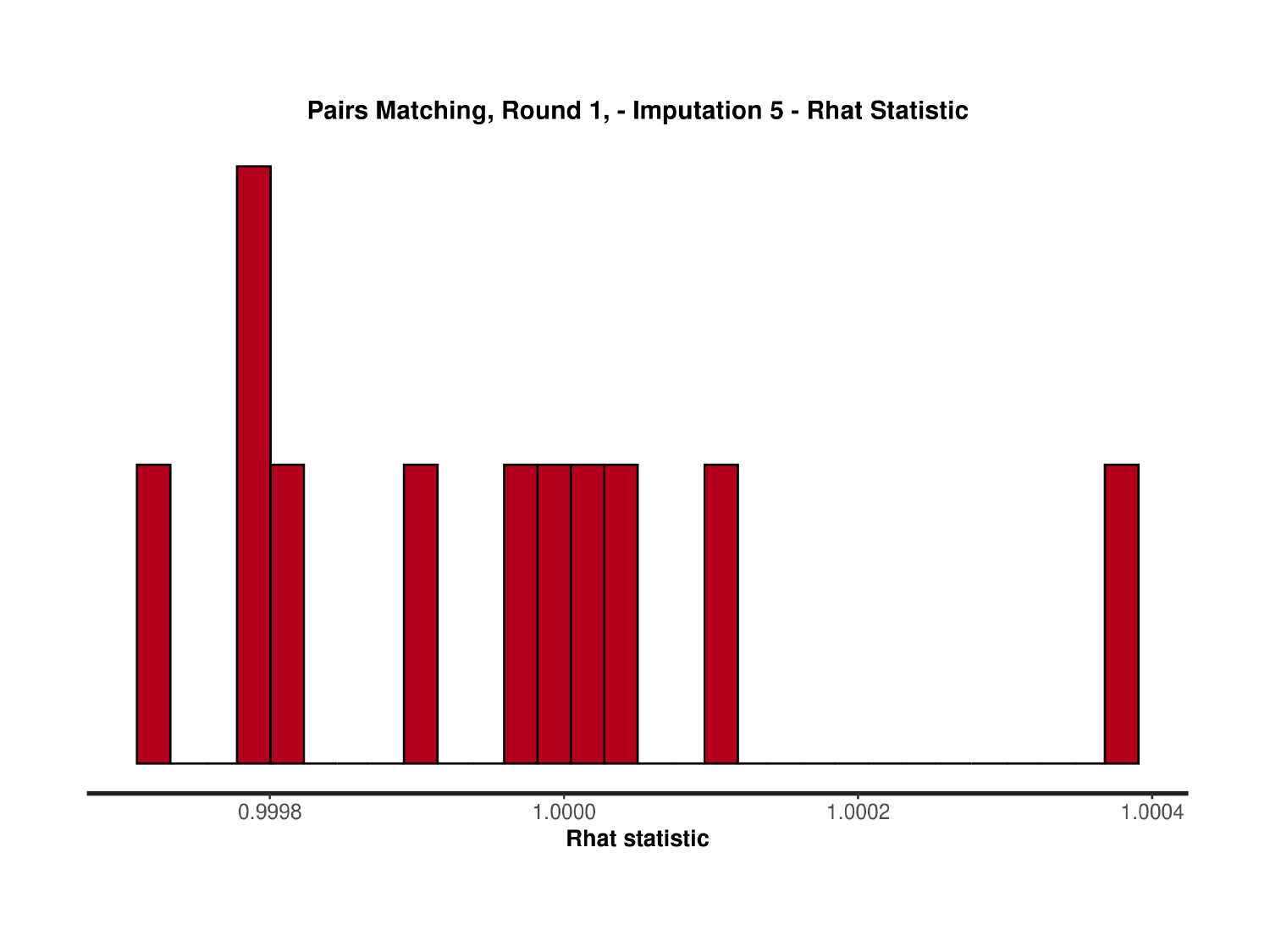

### Slide 53
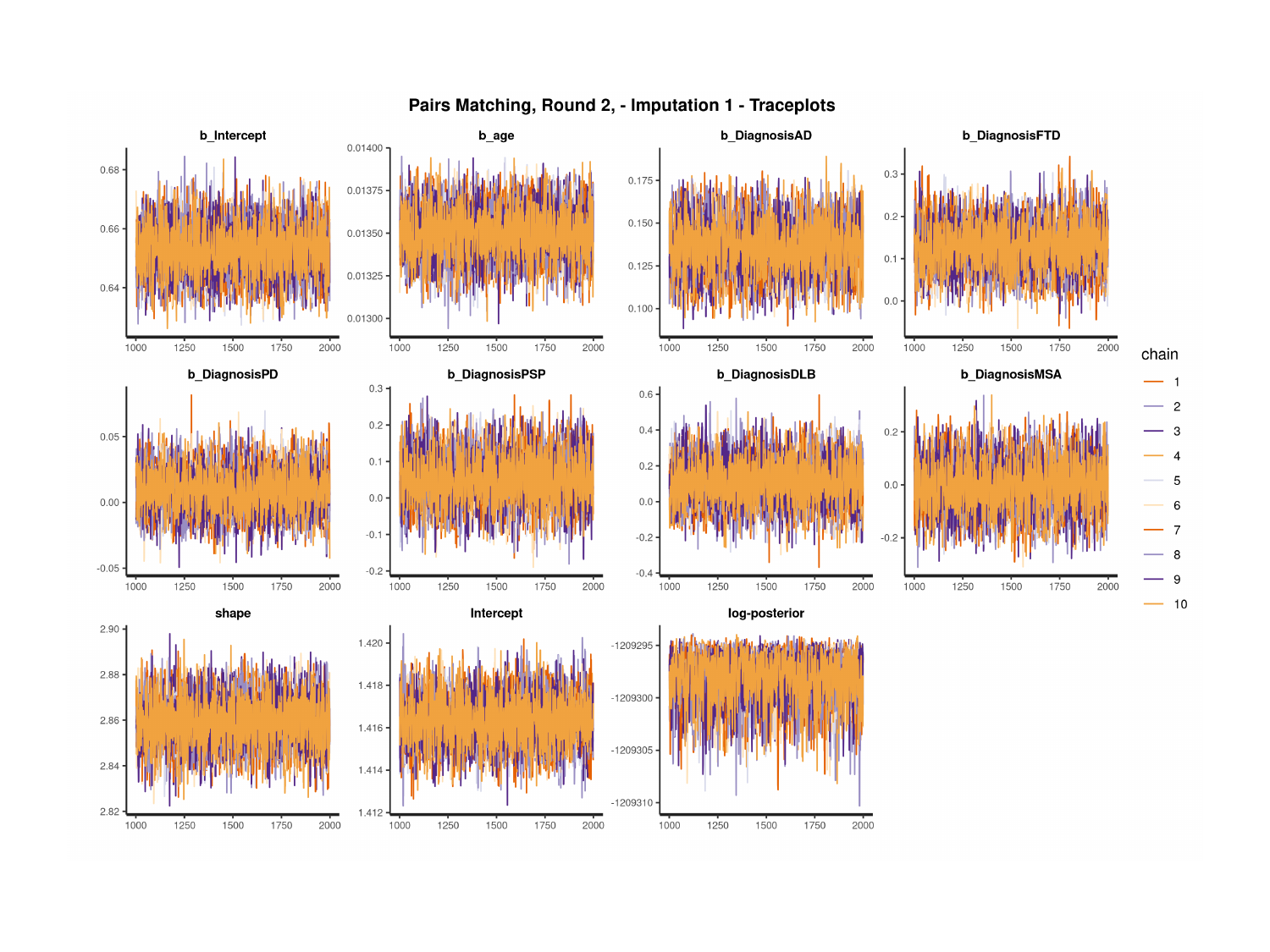

### Slide 54
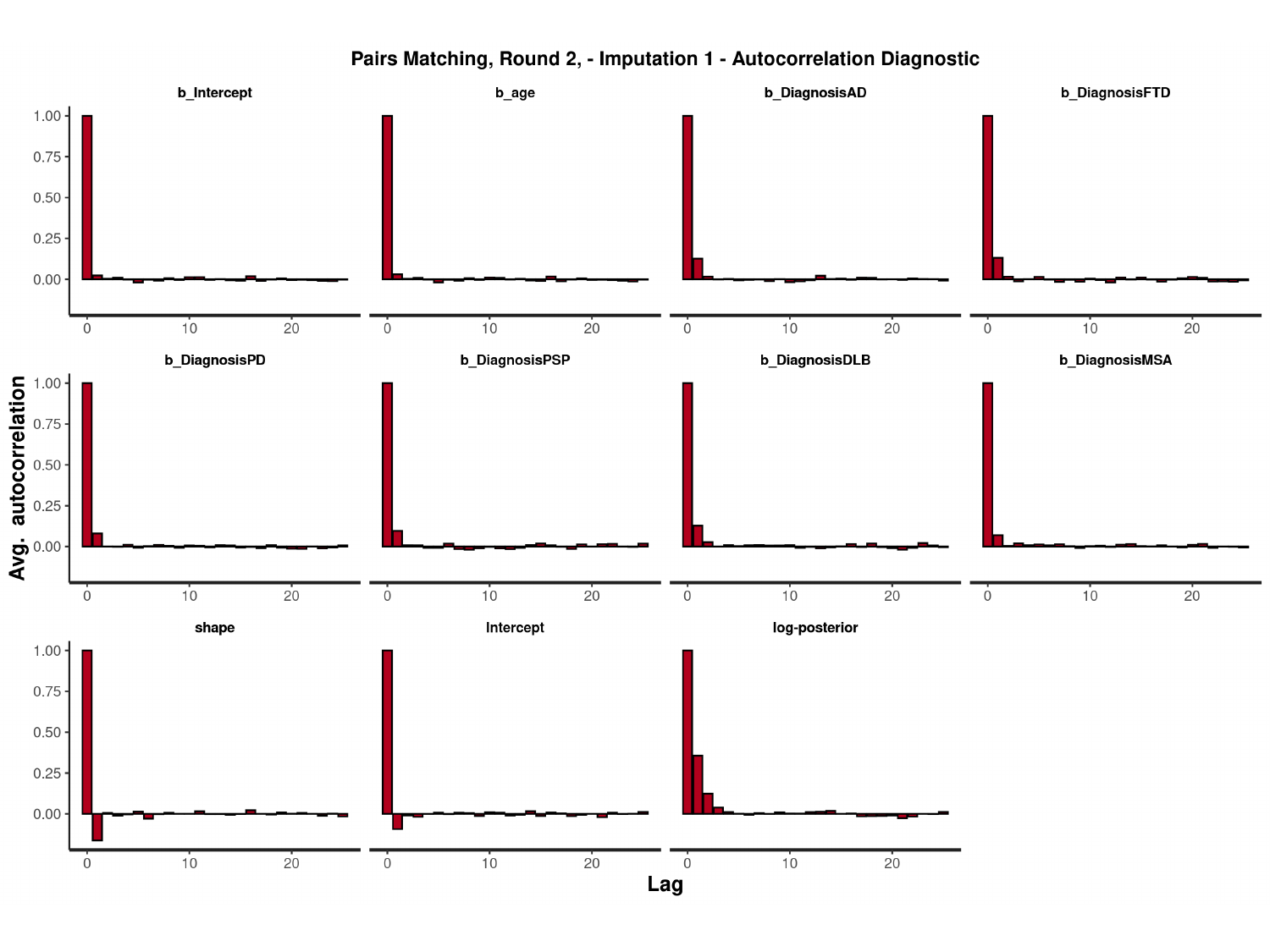

#

### Slide 55
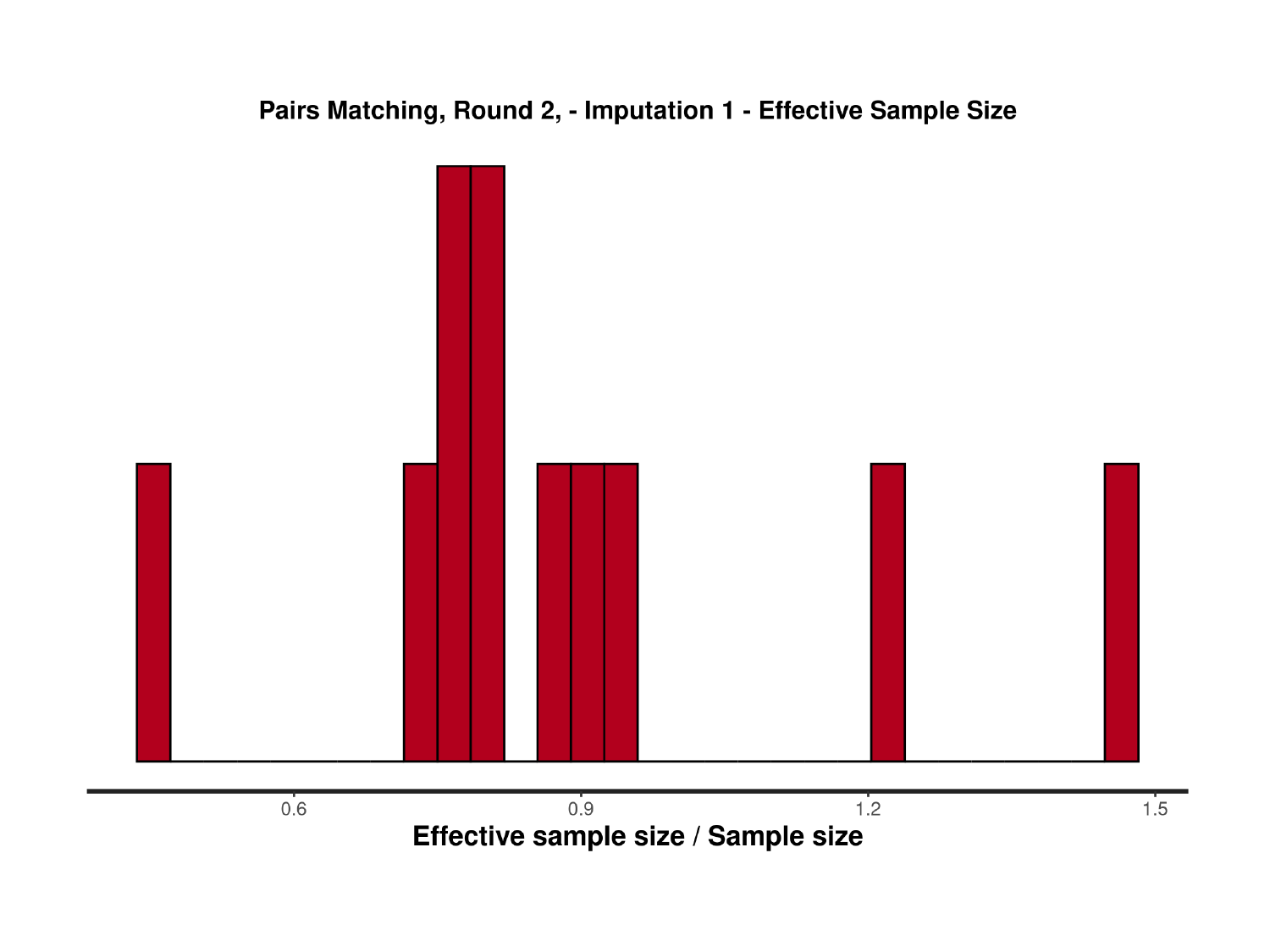

### Slide 56
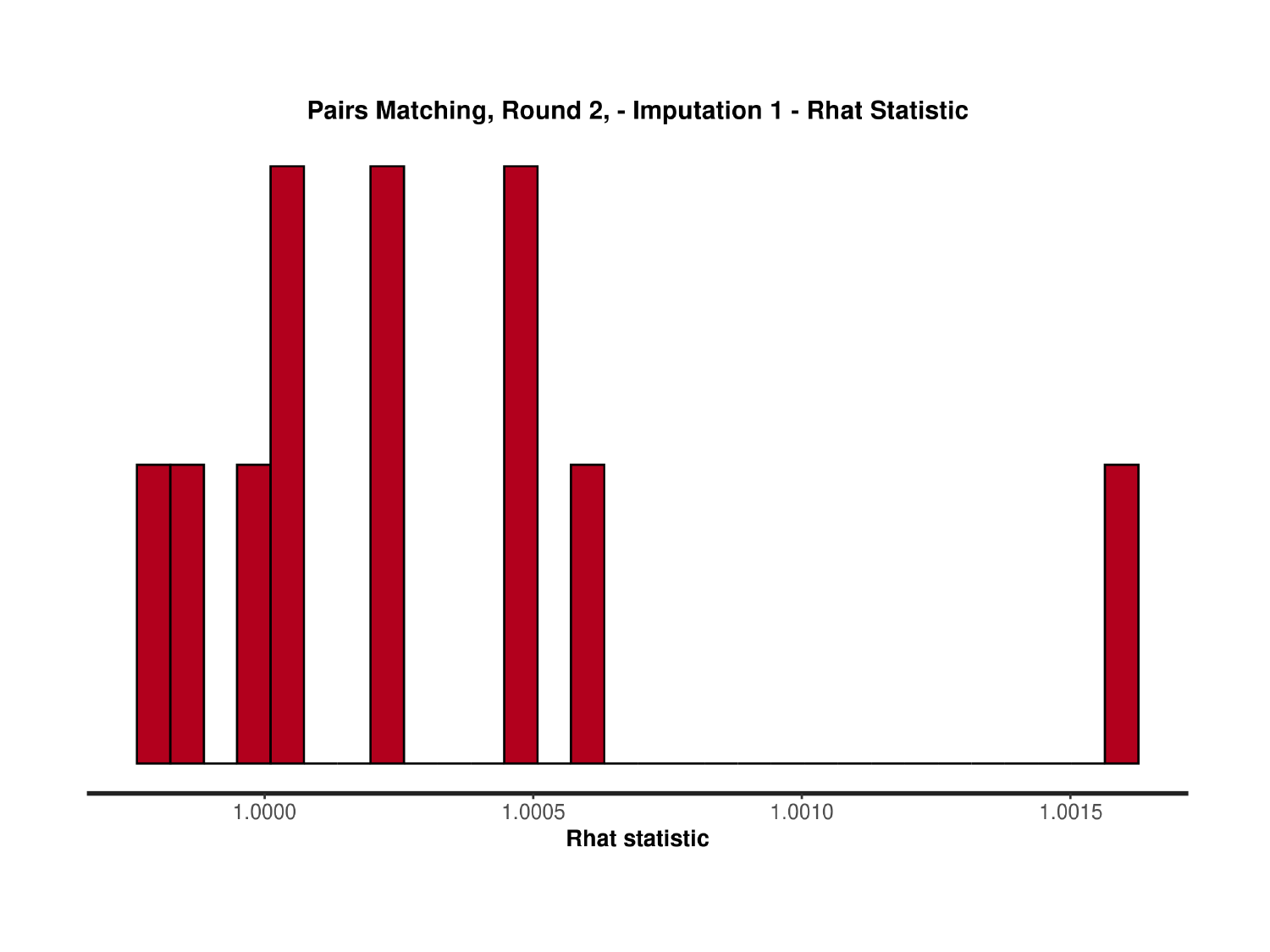

### Slide 57
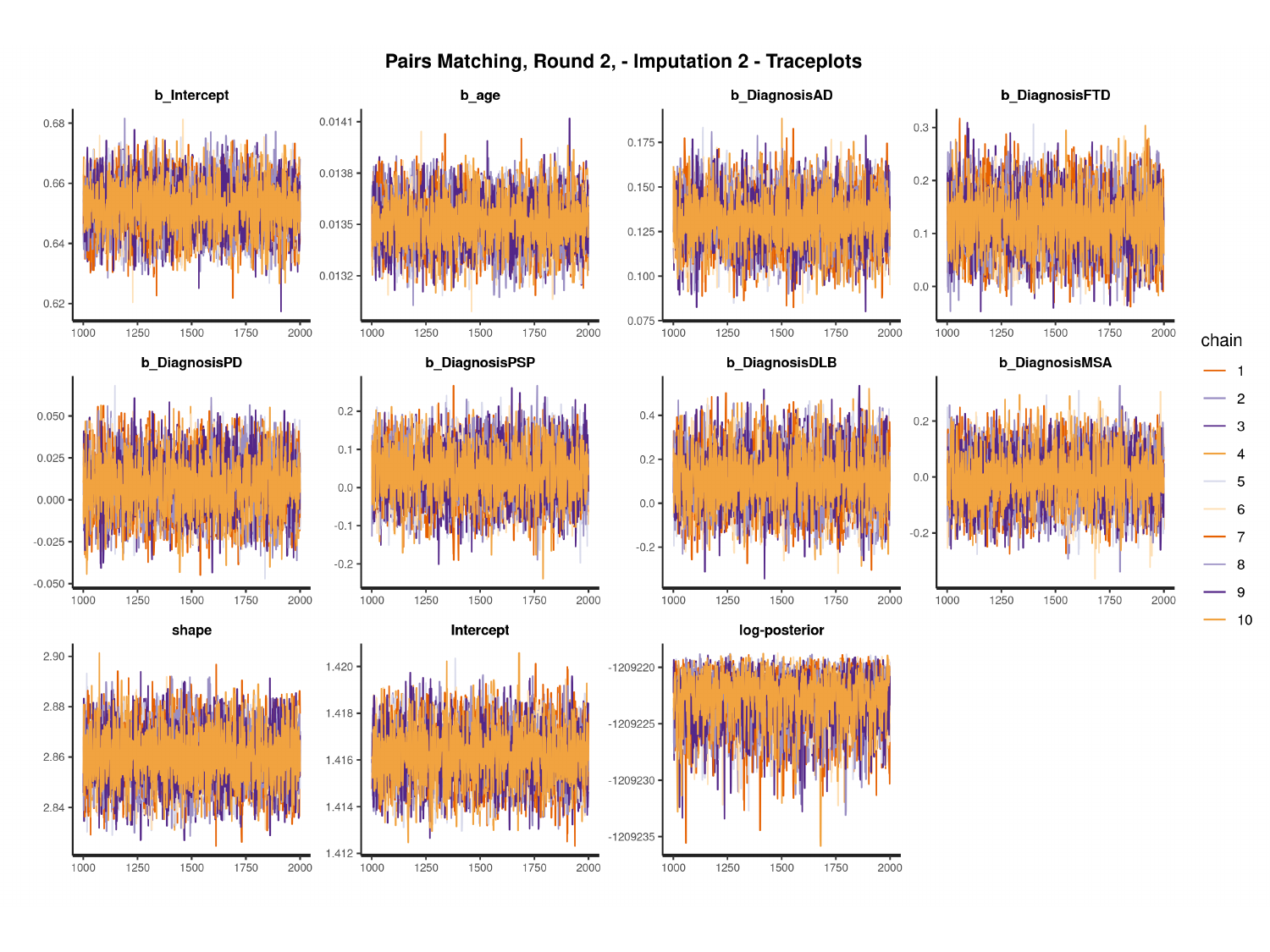

#

### Slide 58
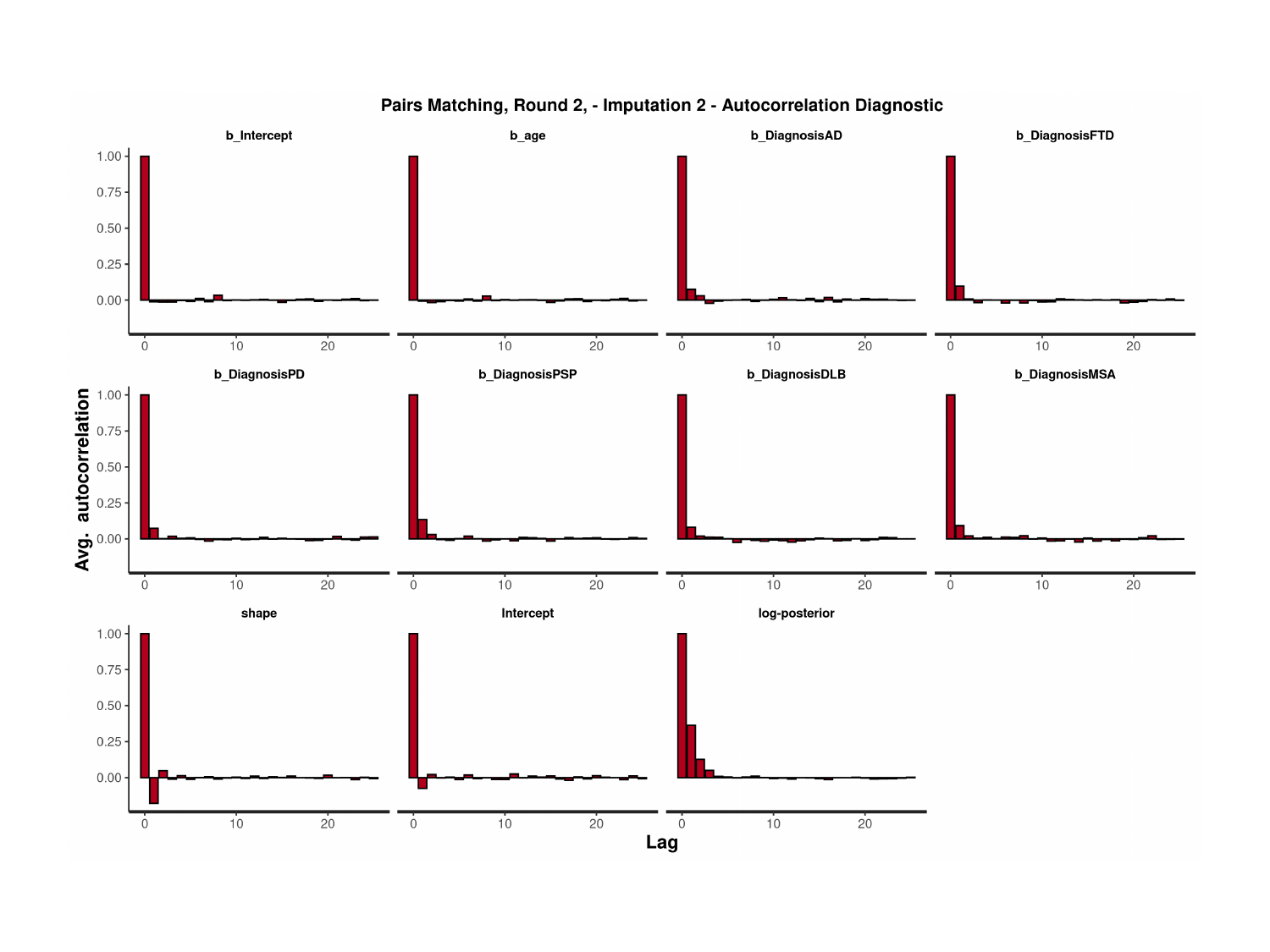

### Slide 59
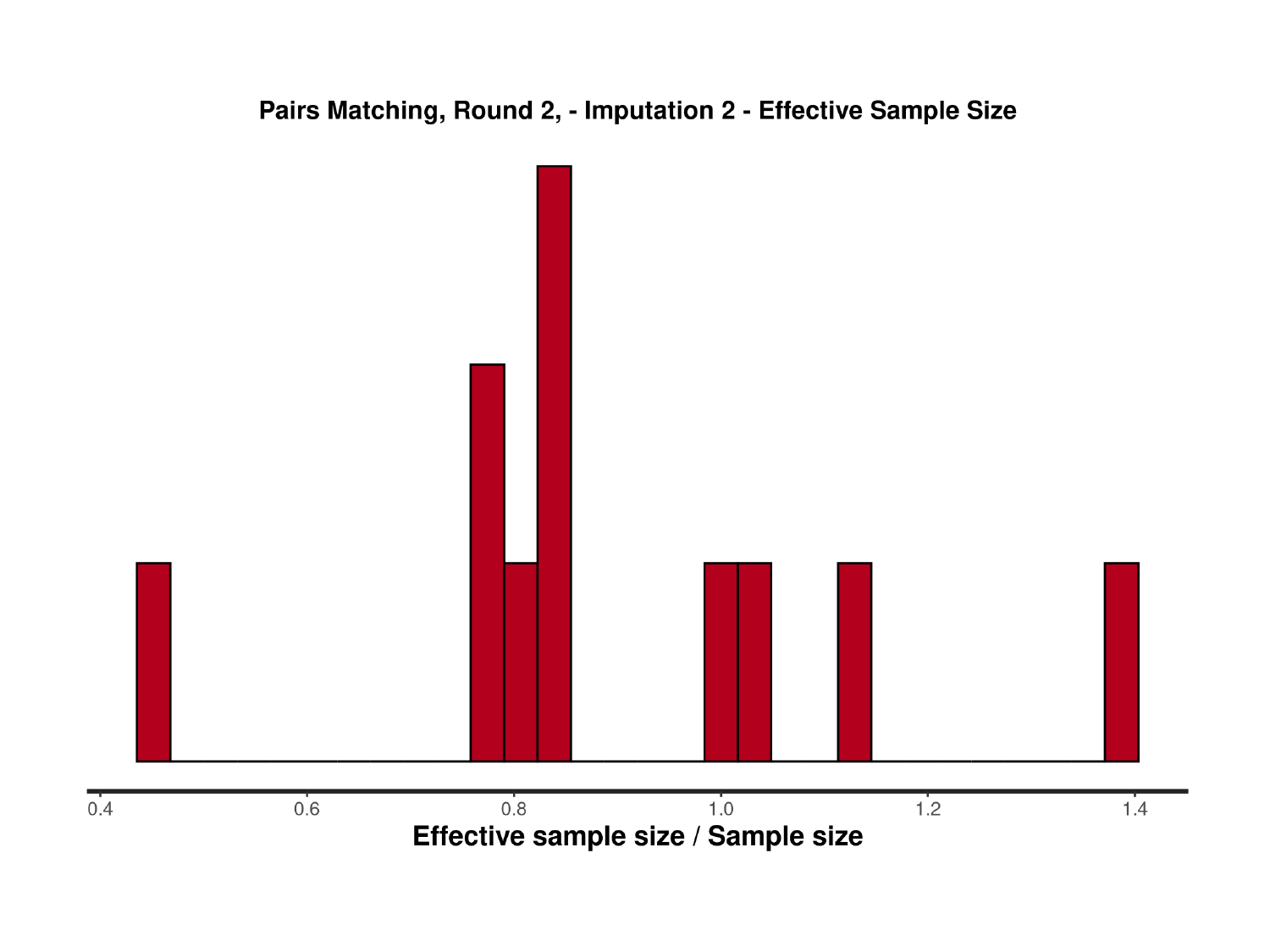

### Slide 60
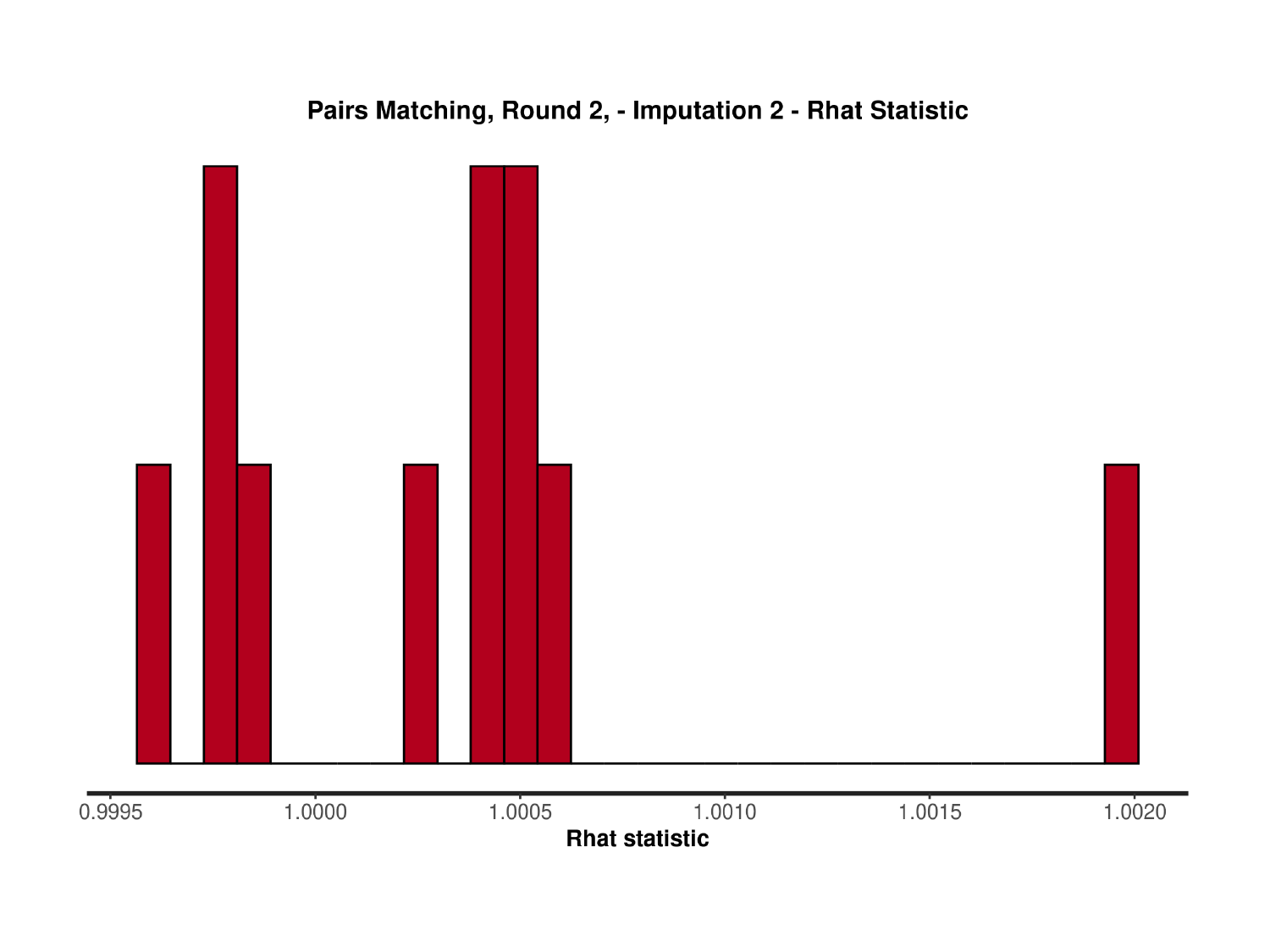

### Slide 61
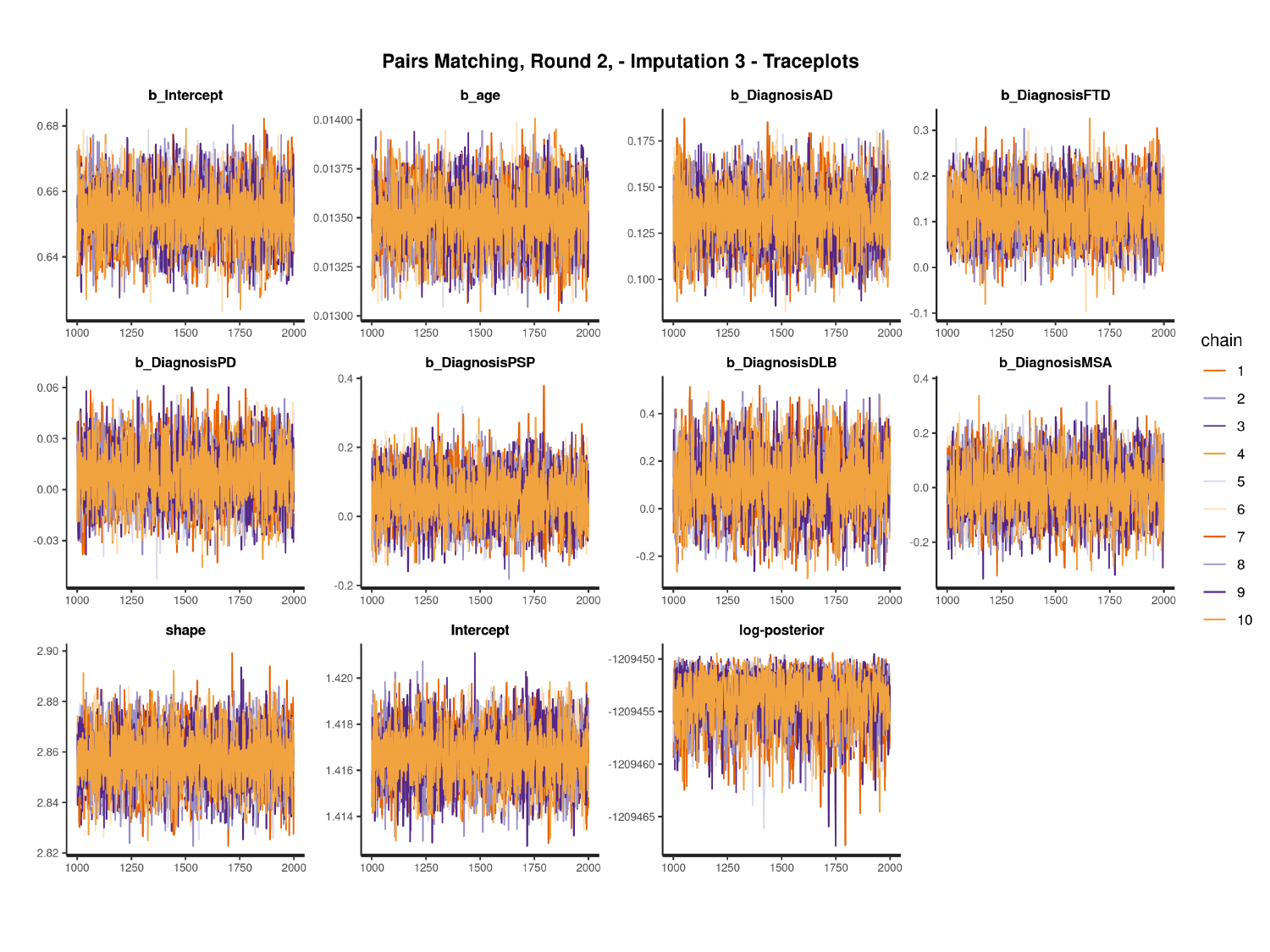

#

### Slide 62
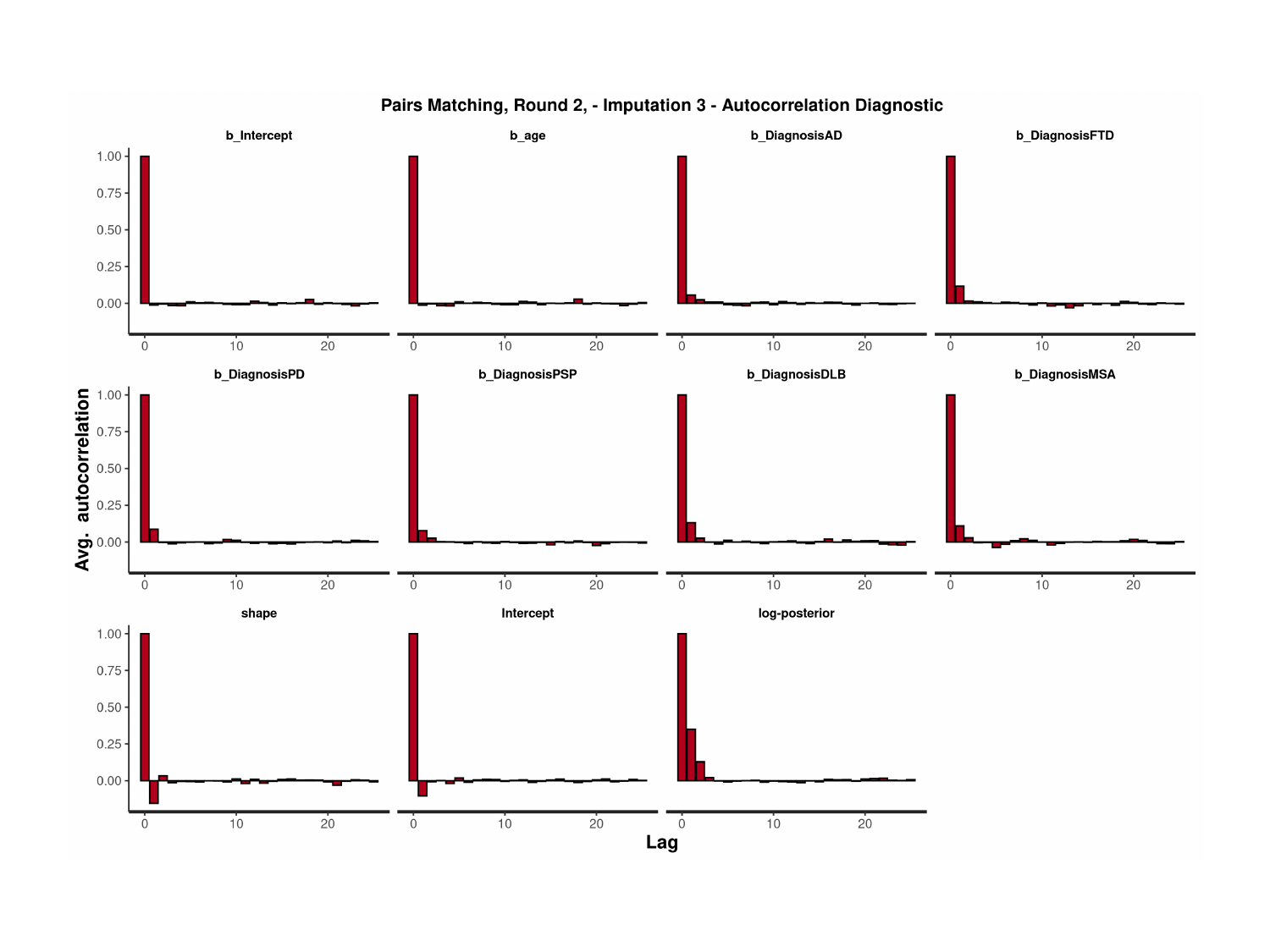

### Slide 63
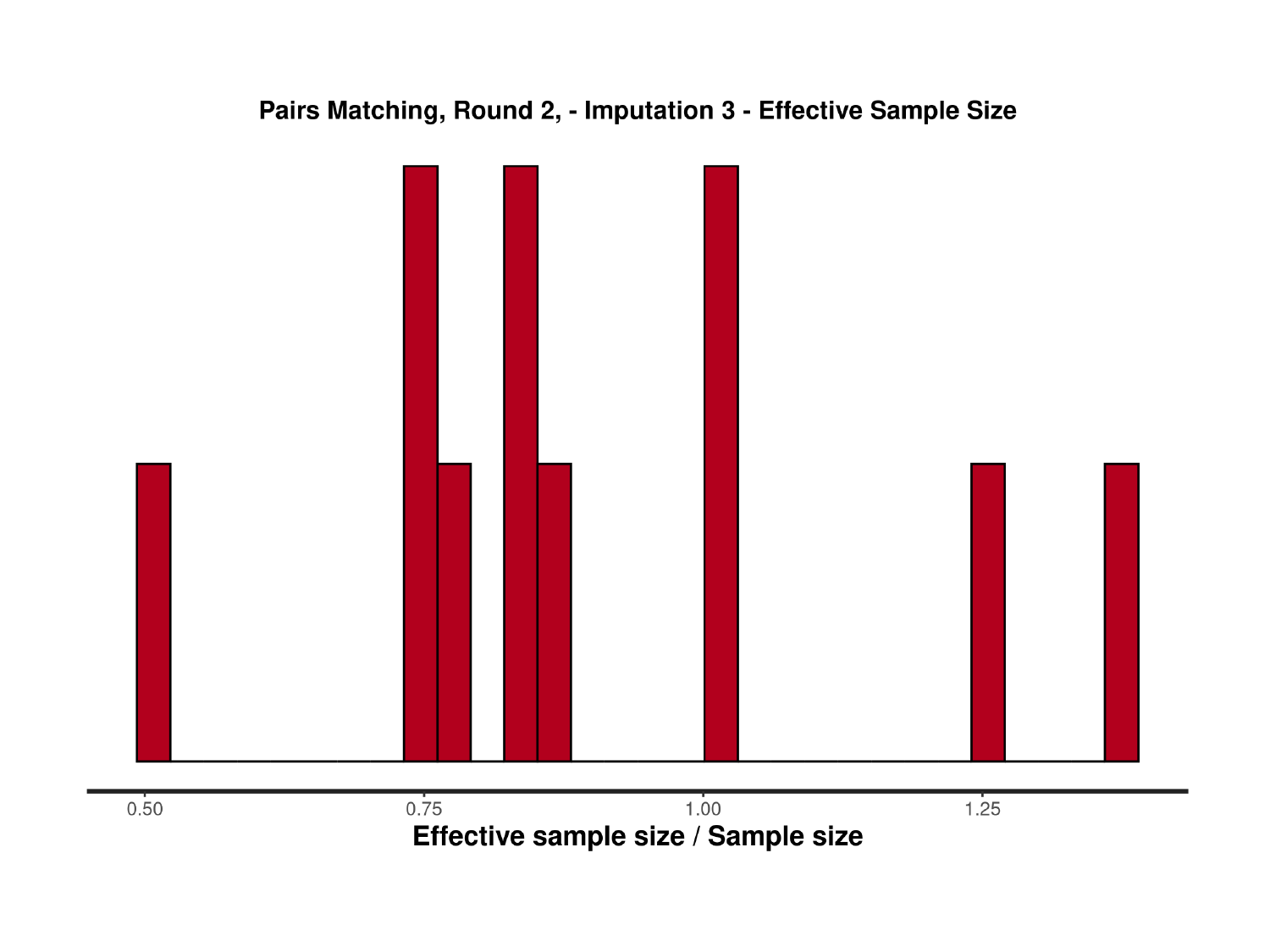

### Slide 64
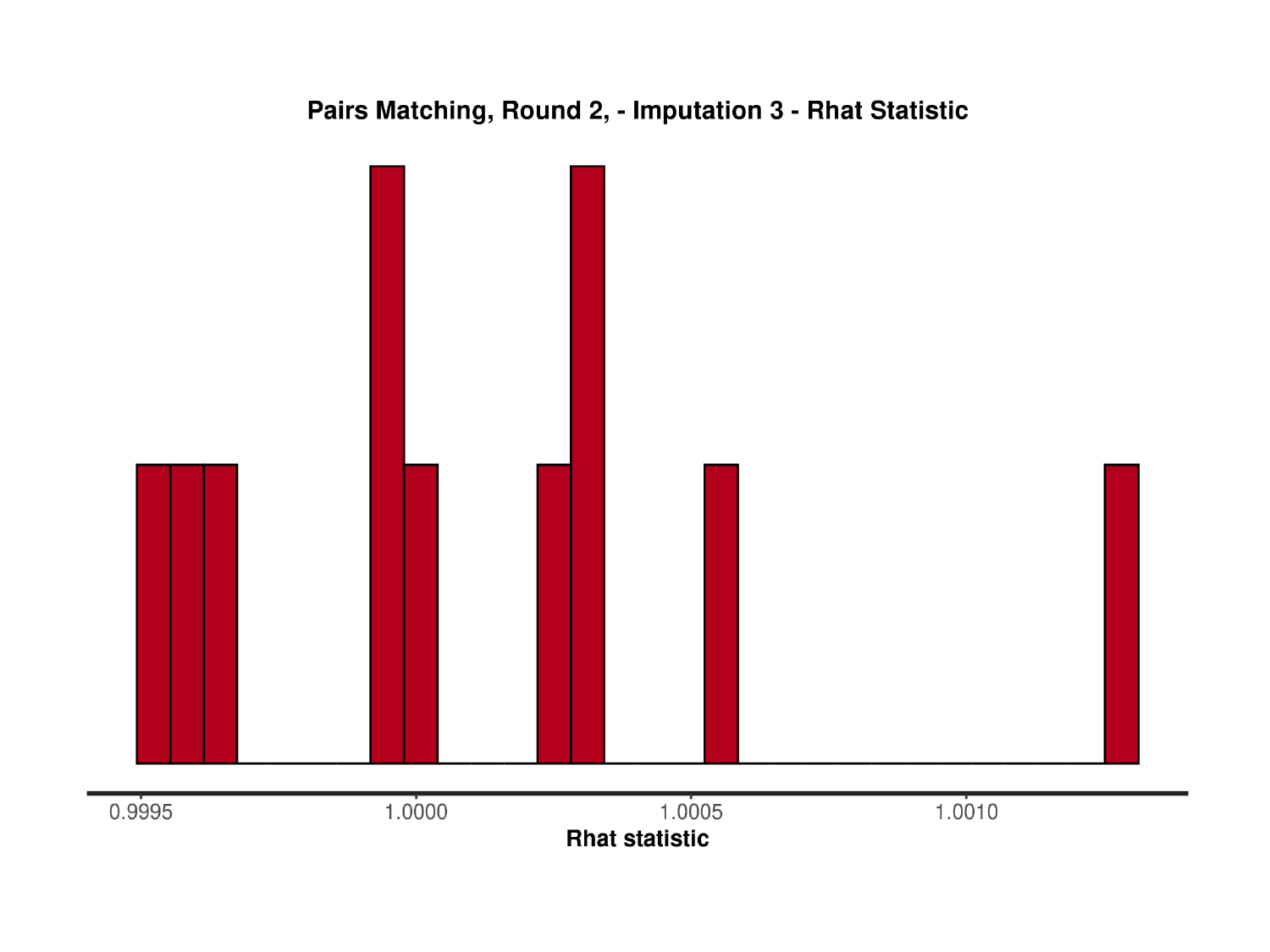

### Slide 65
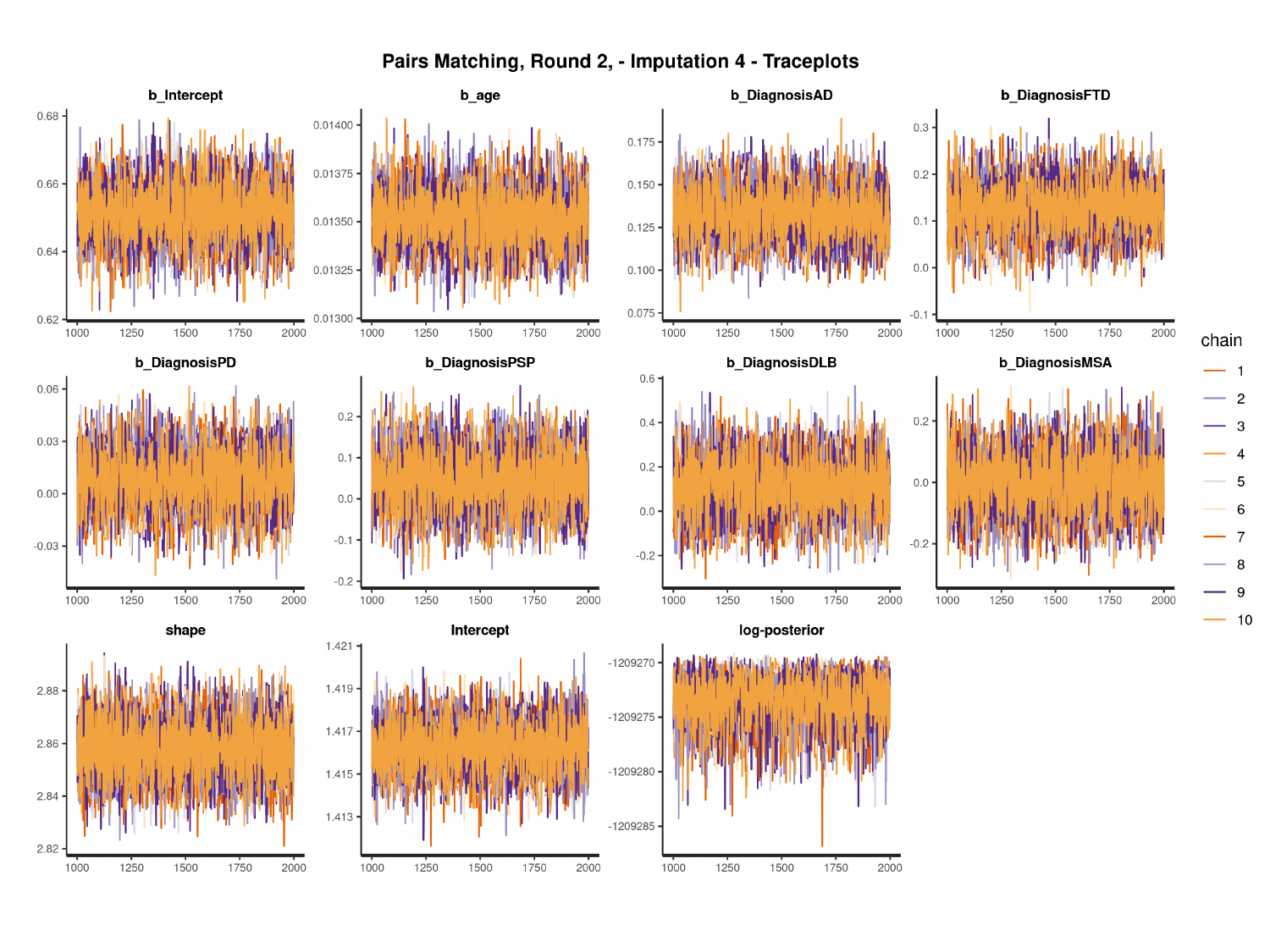

#

### Slide 66
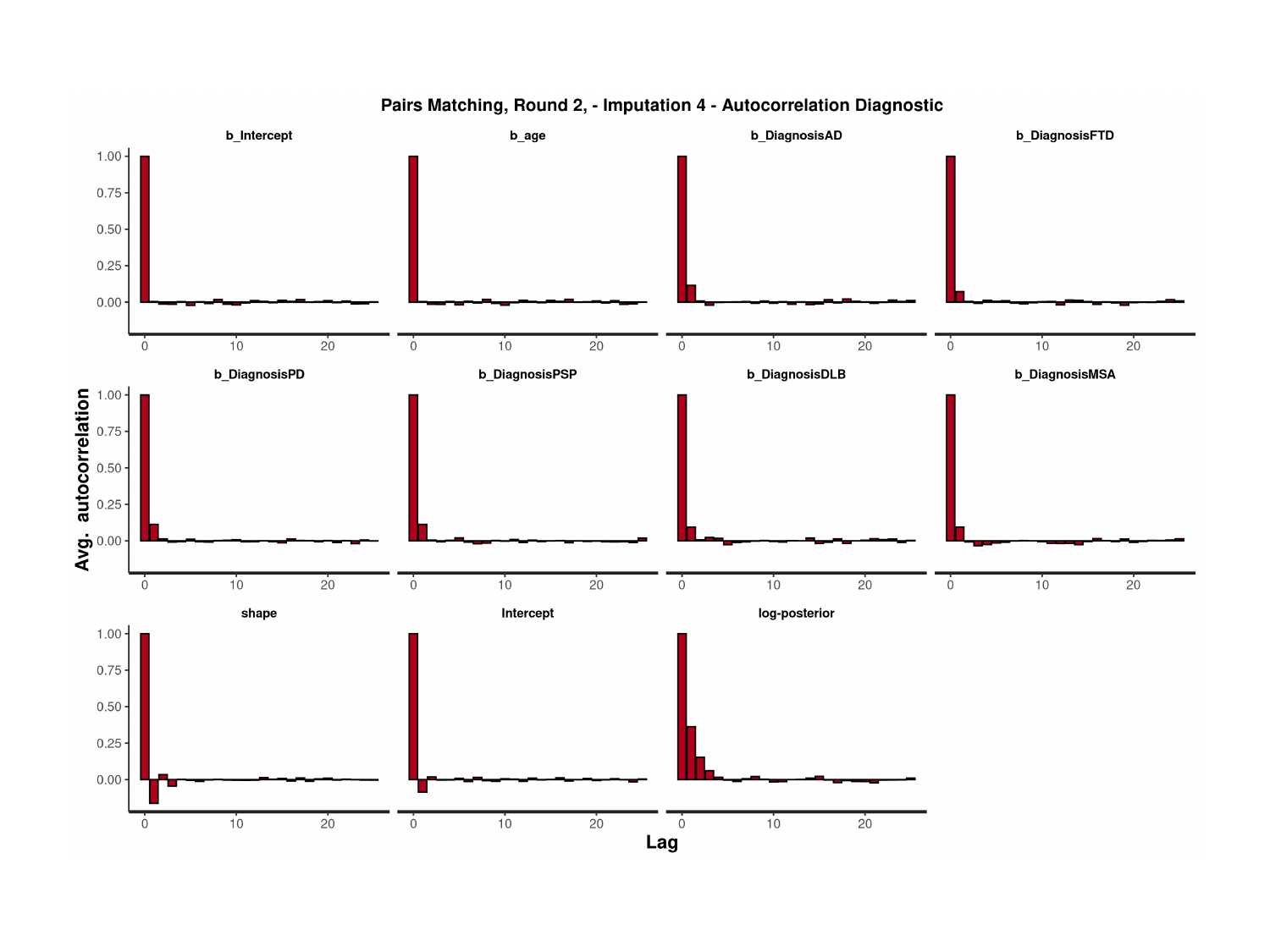

### Slide 67
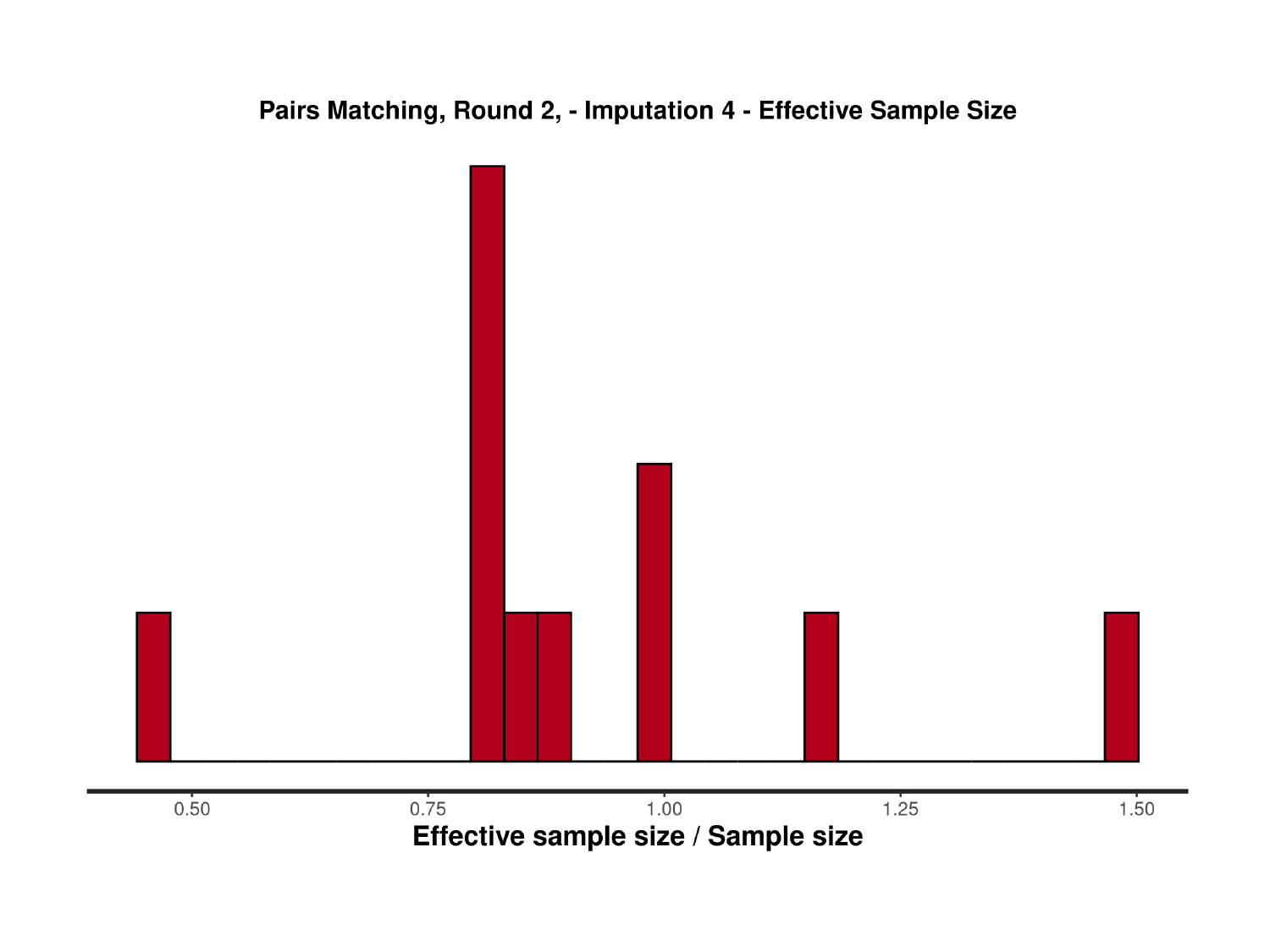

### Slide 68
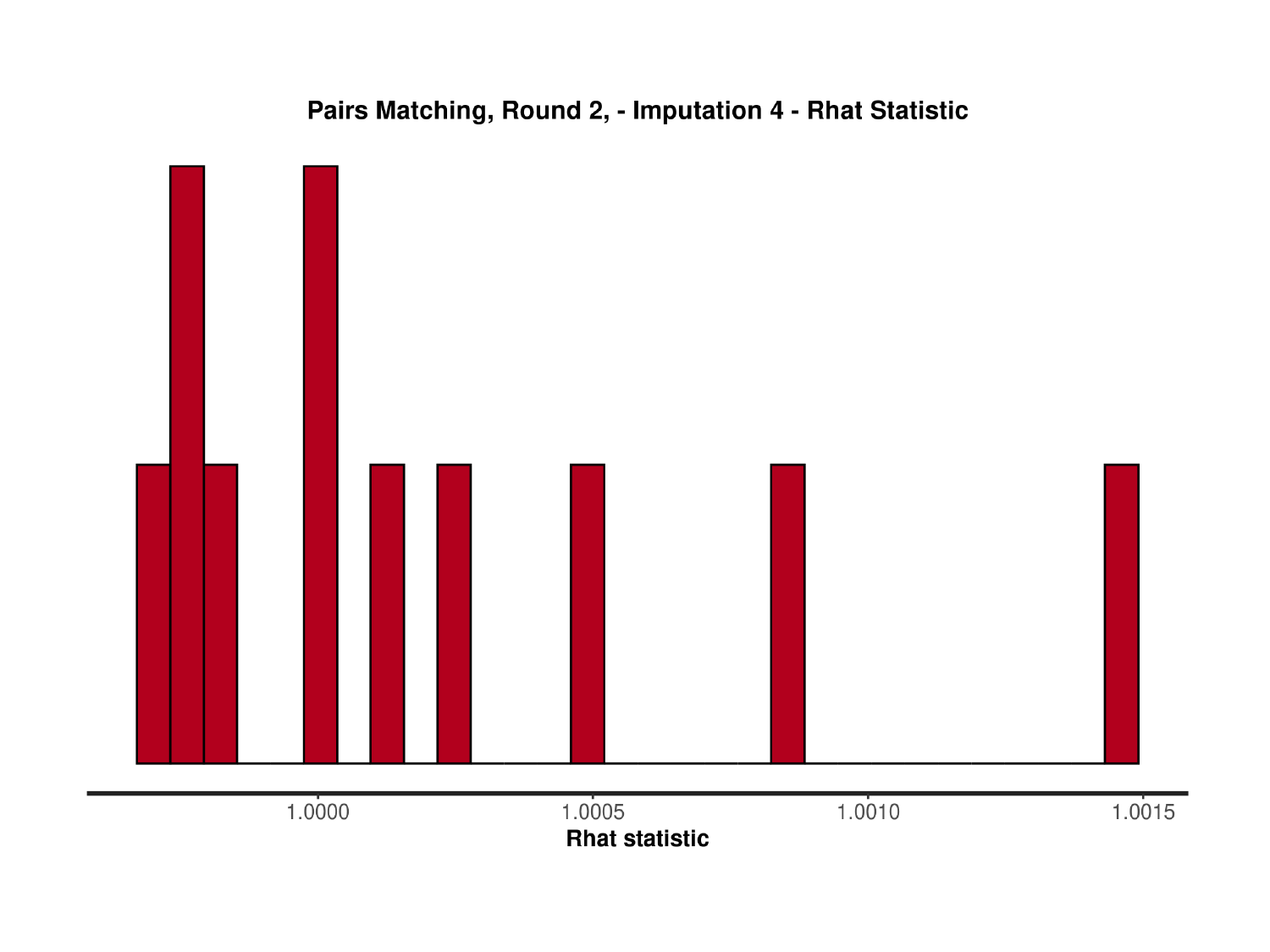

### Slide 69
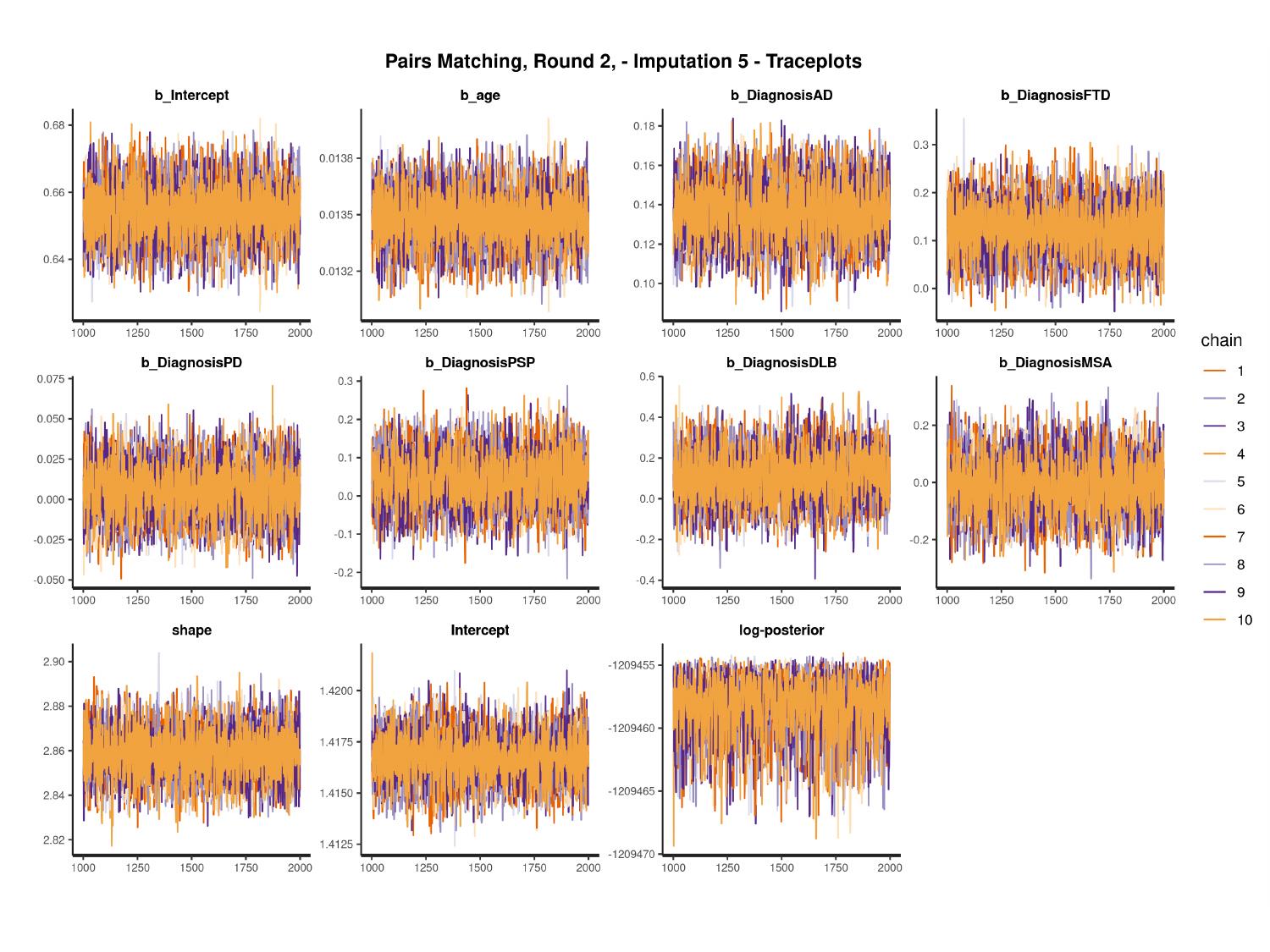

#

### Slide 70
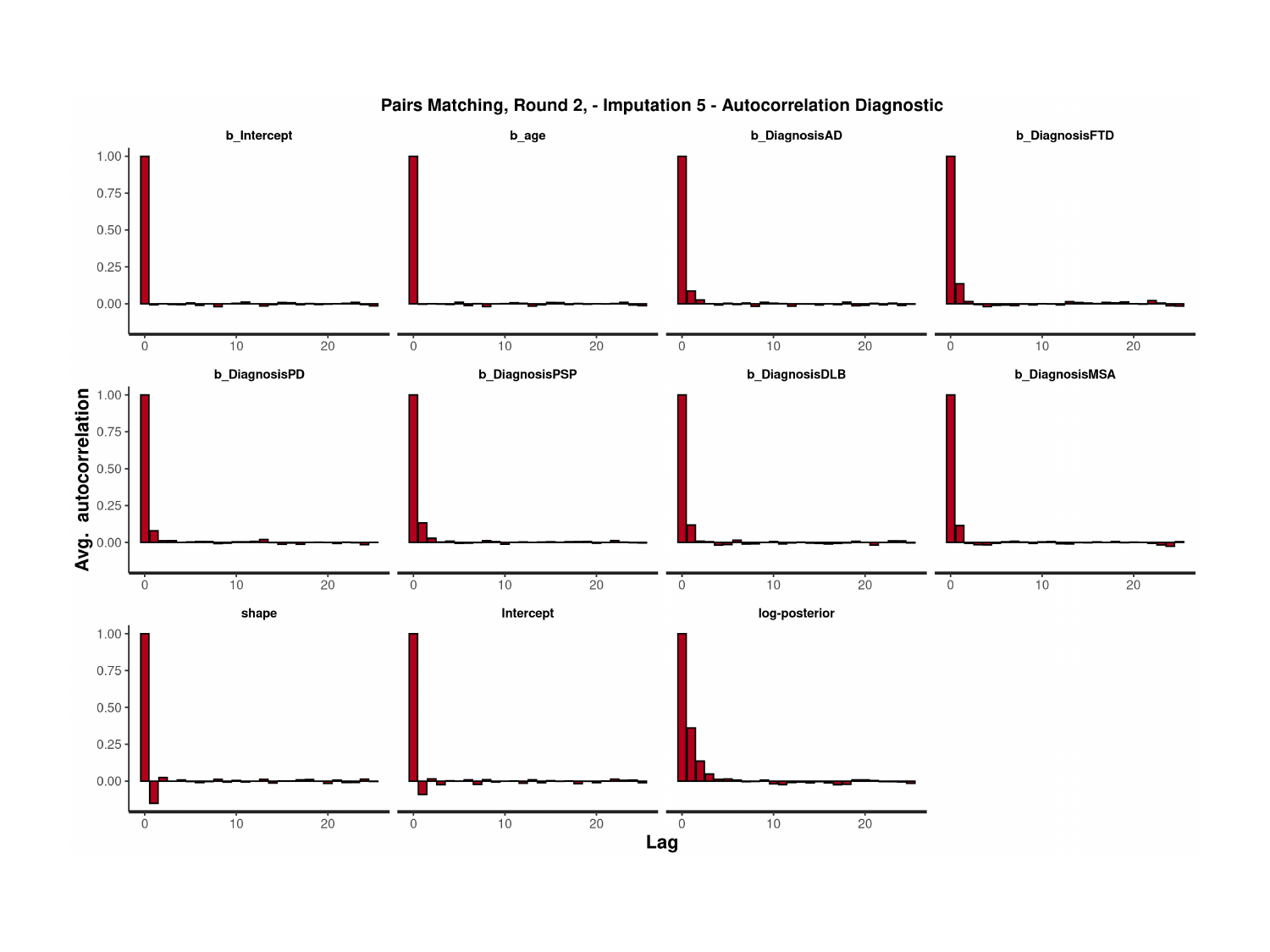

### Slide 71
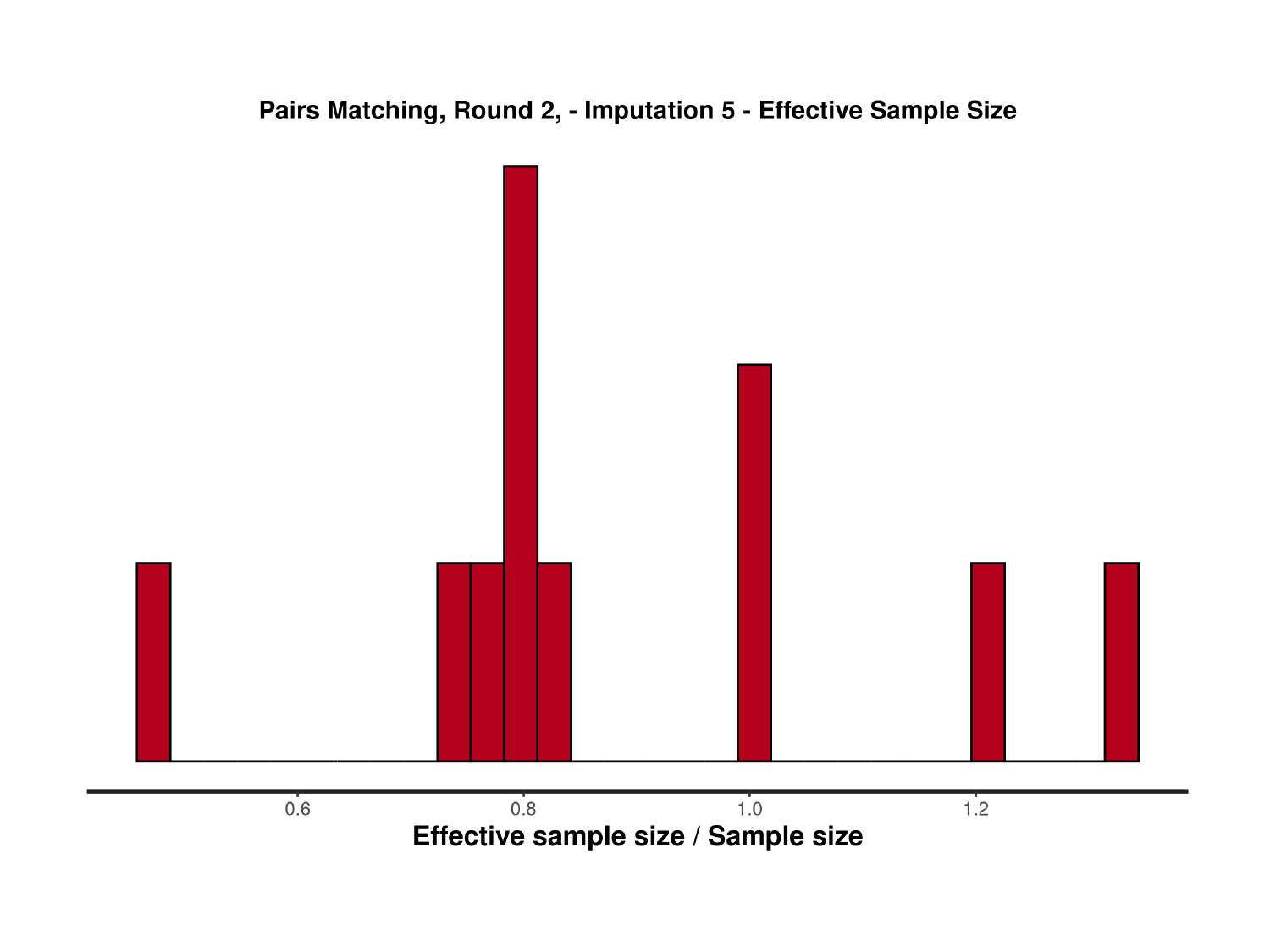

### Slide 72
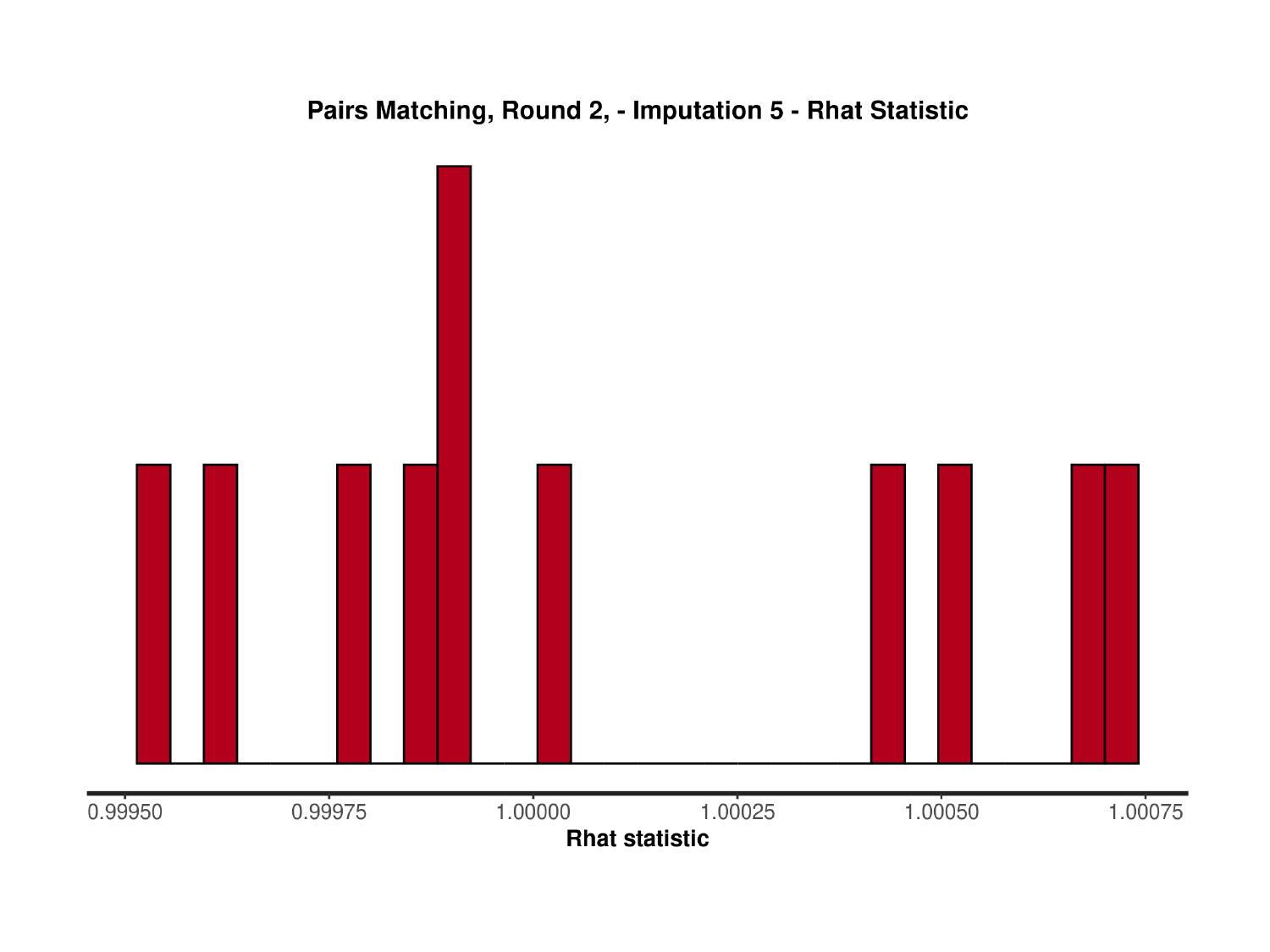

### Slide 73
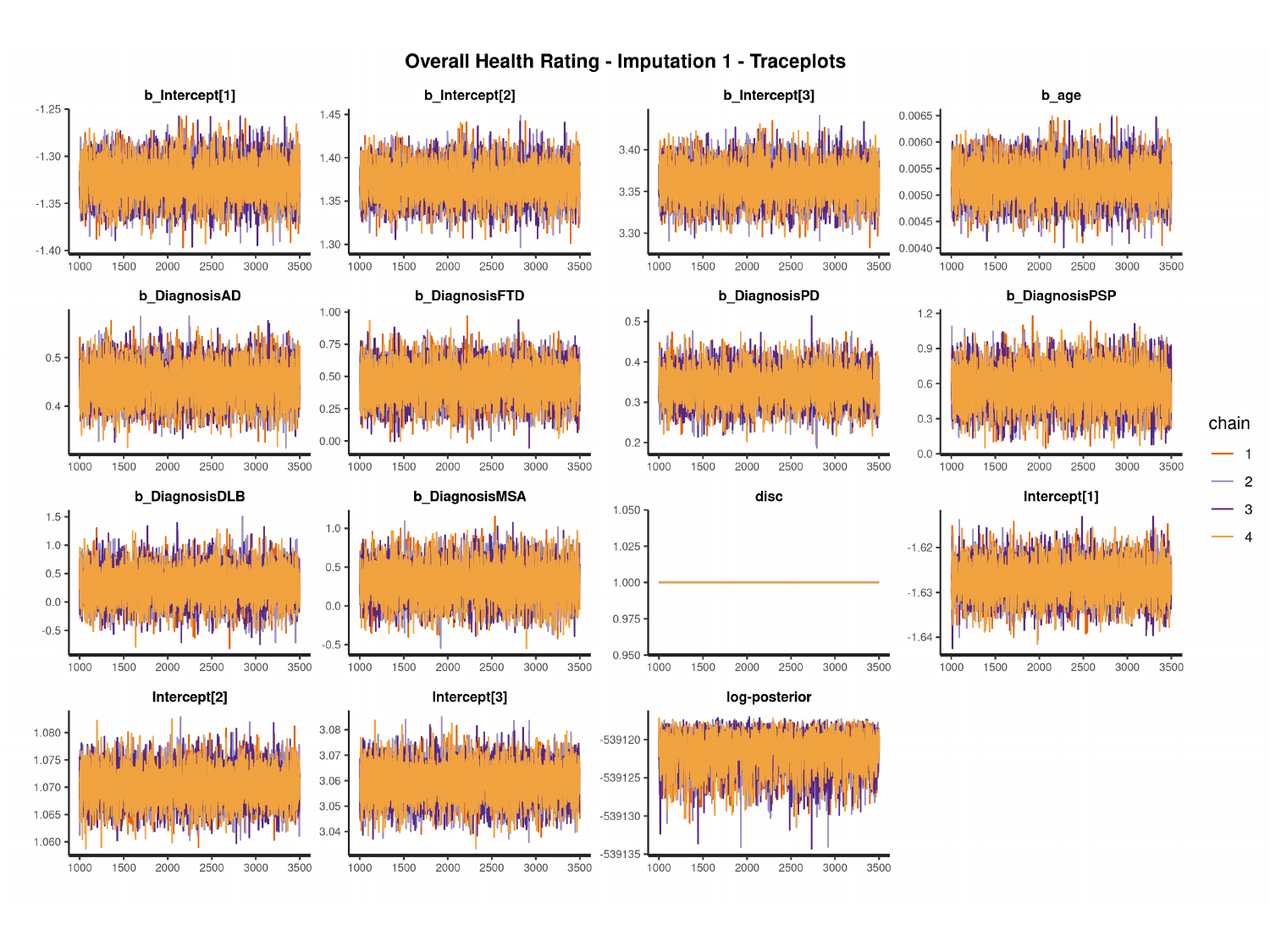

#

### Slide 74
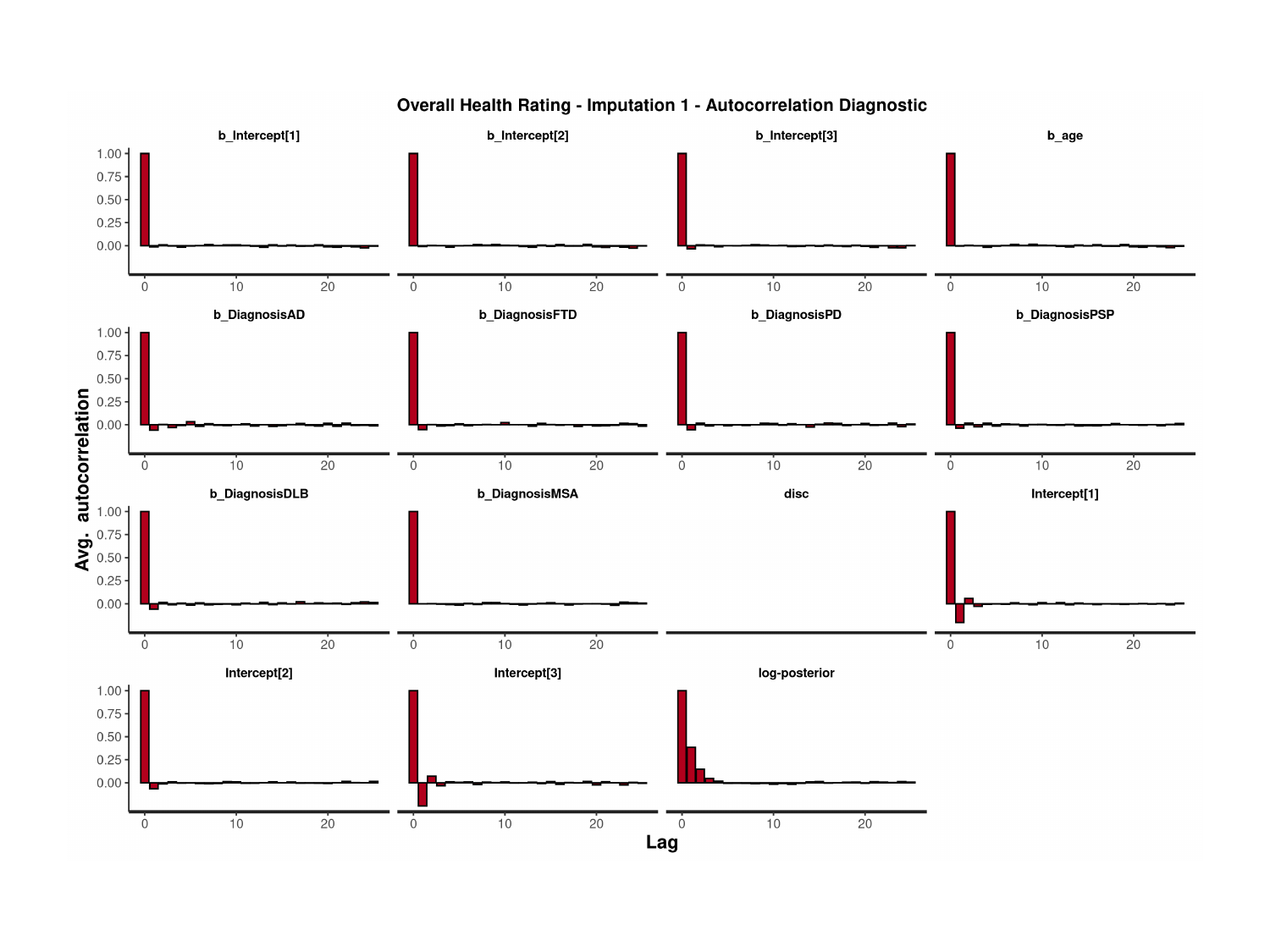

### Slide 75
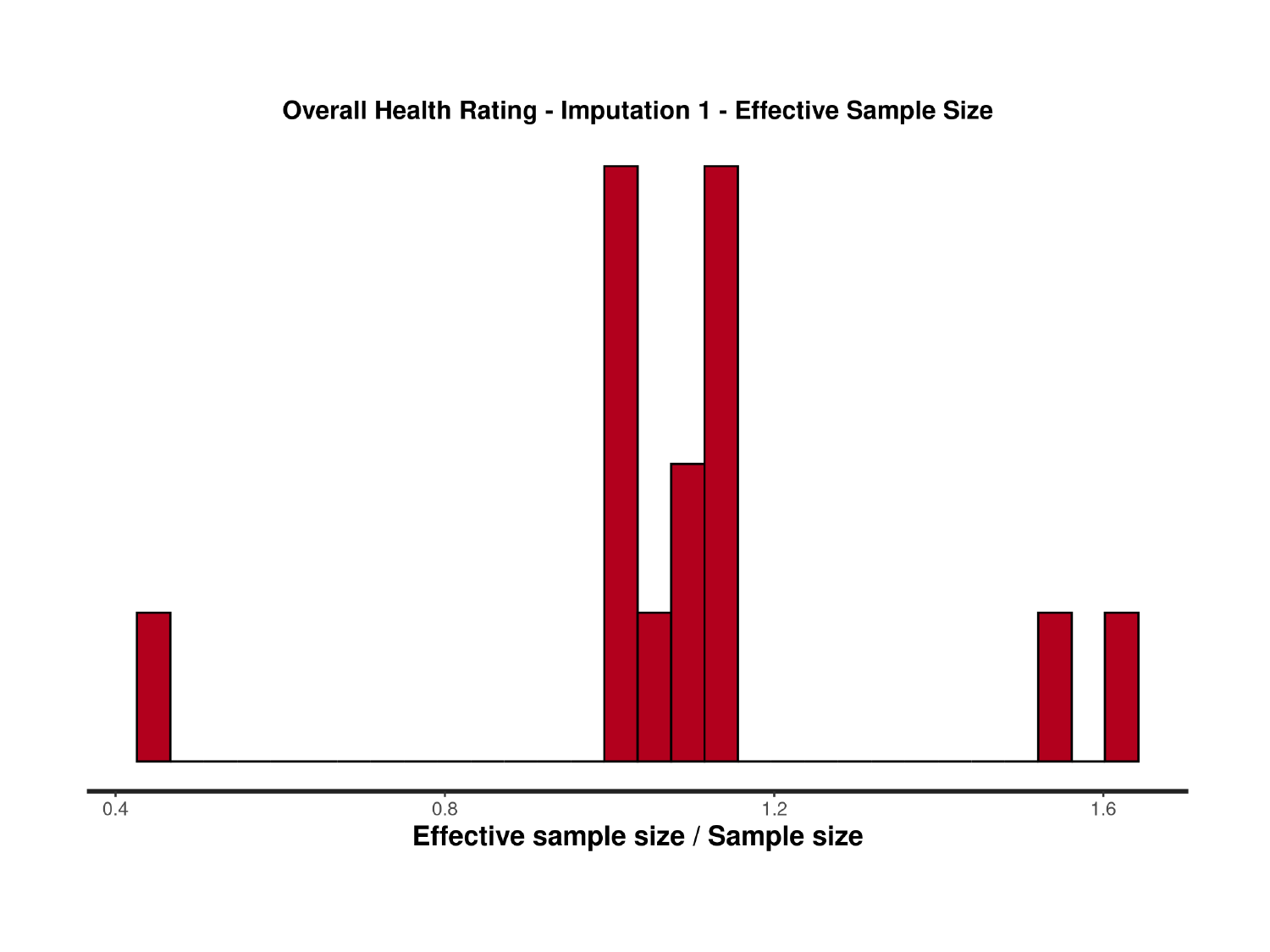

### Slide 76
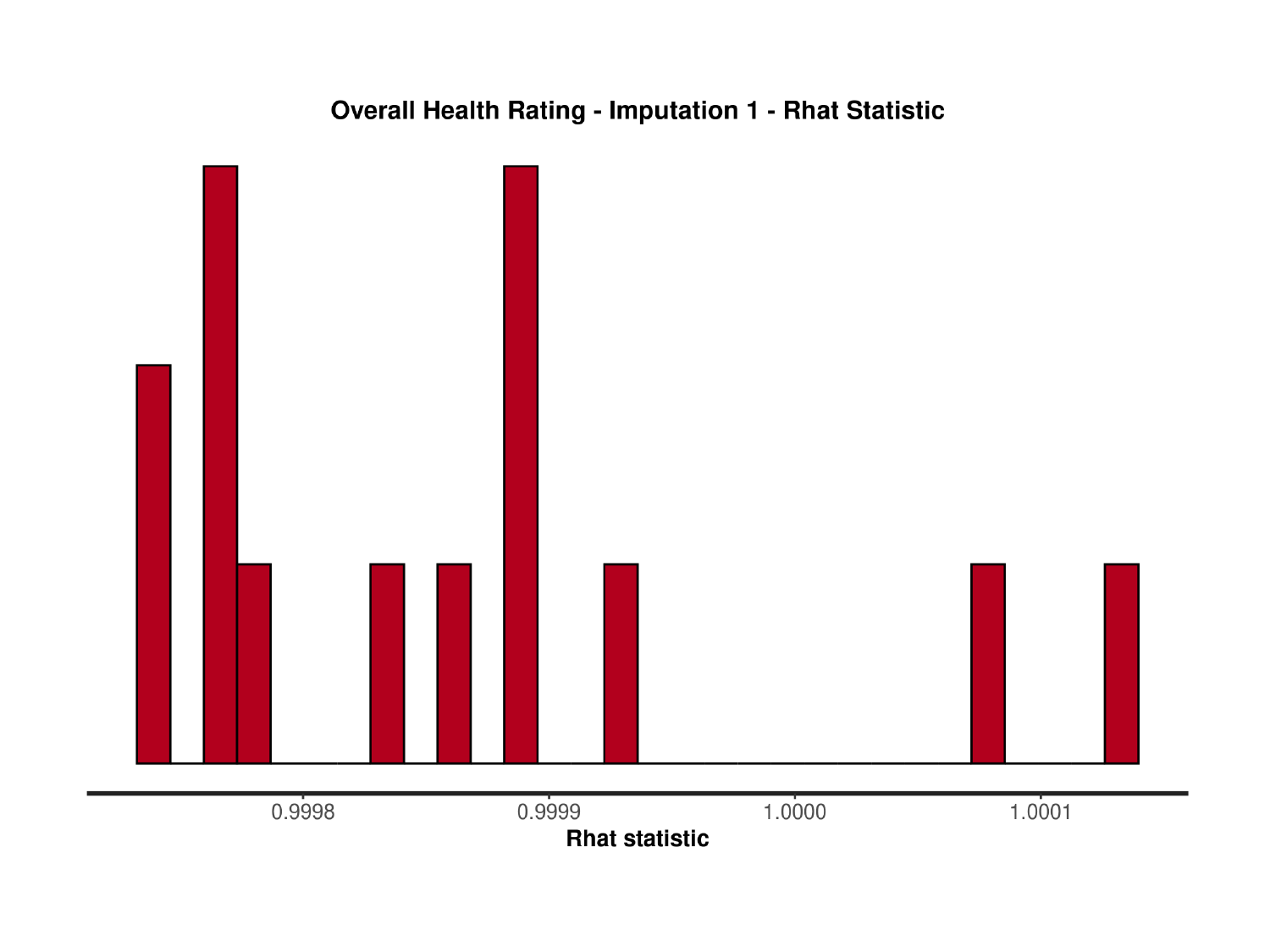

### Slide 77
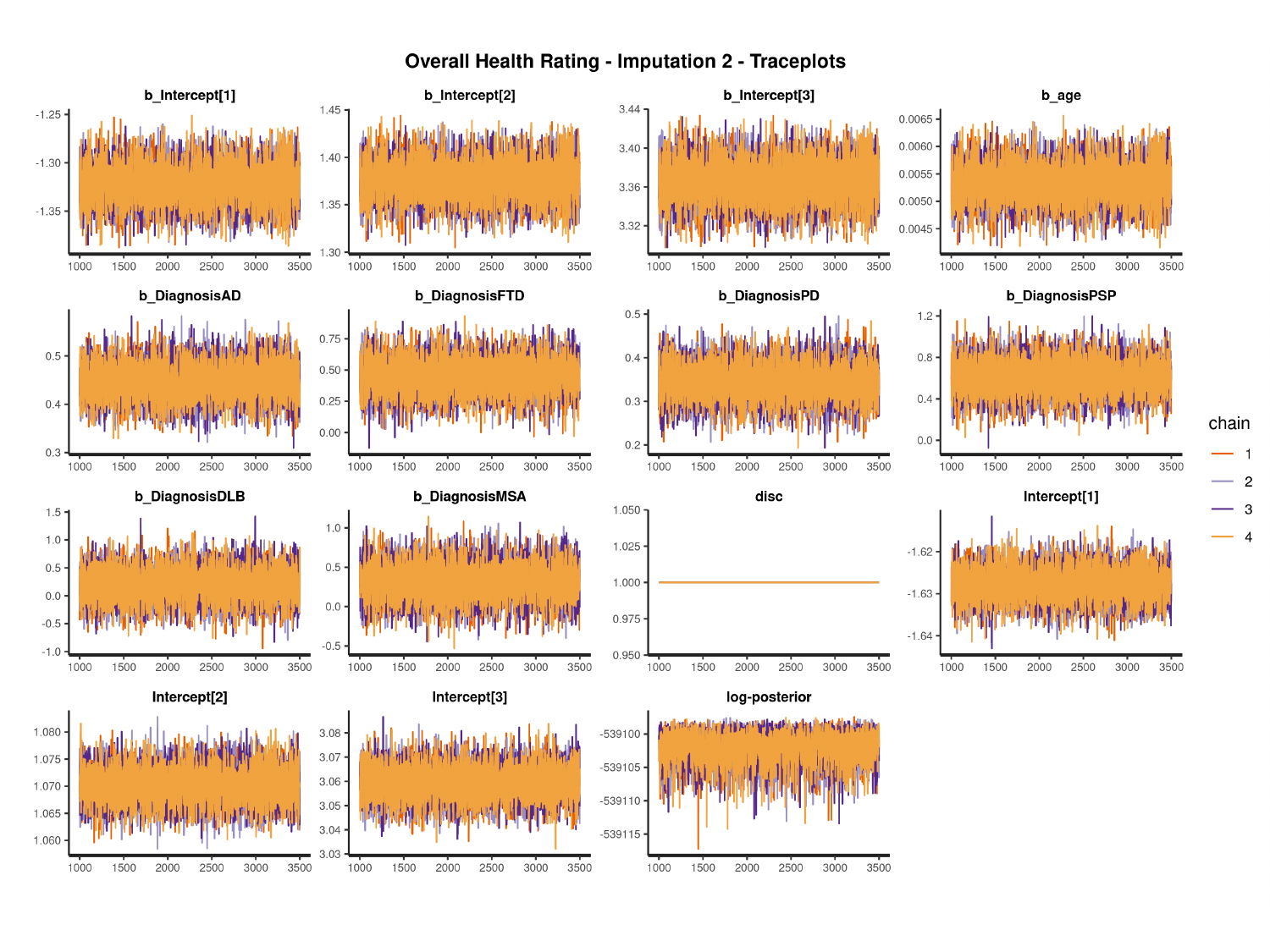

#

### Slide 78
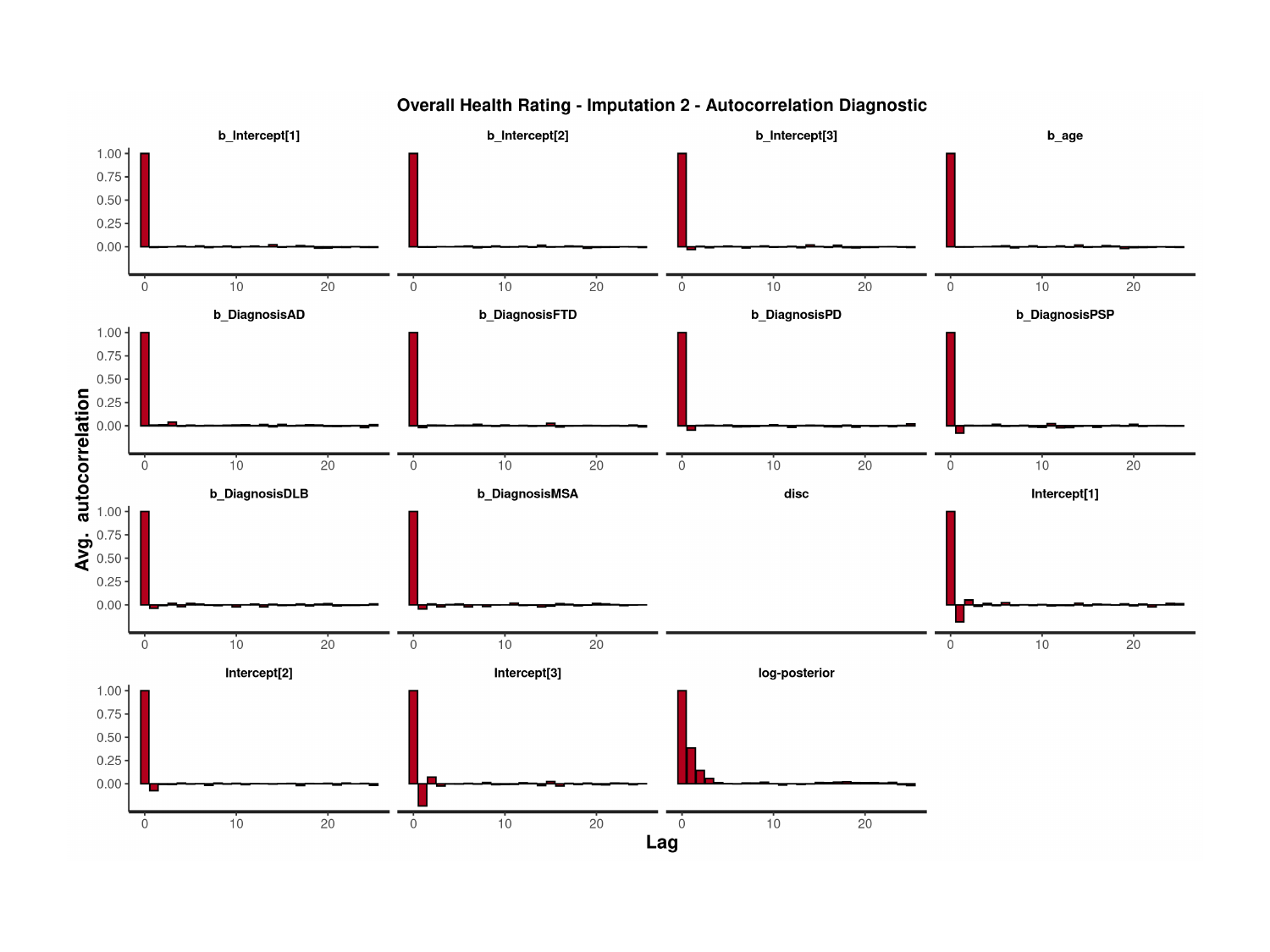

### Slide 79
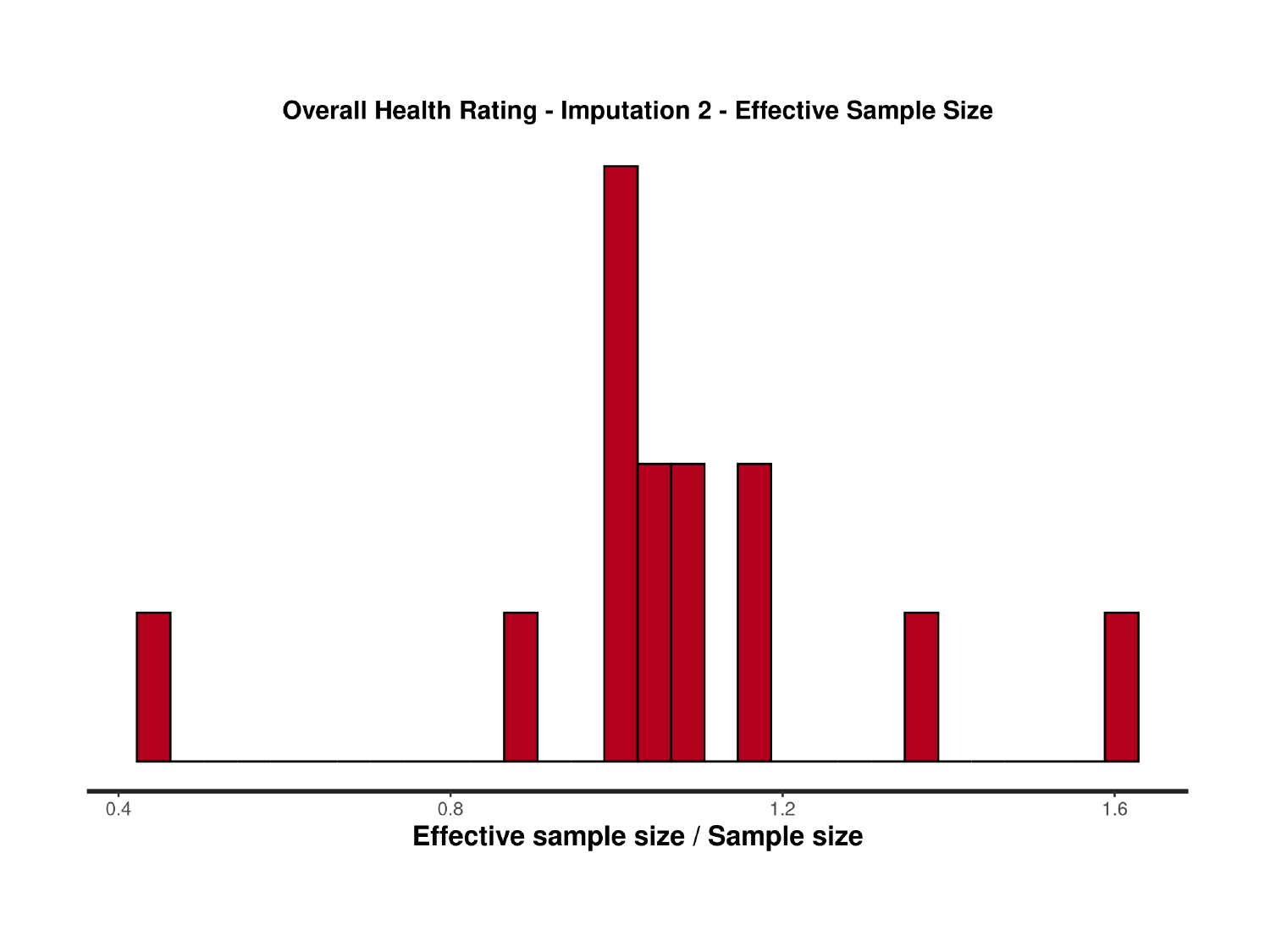

### Slide 80
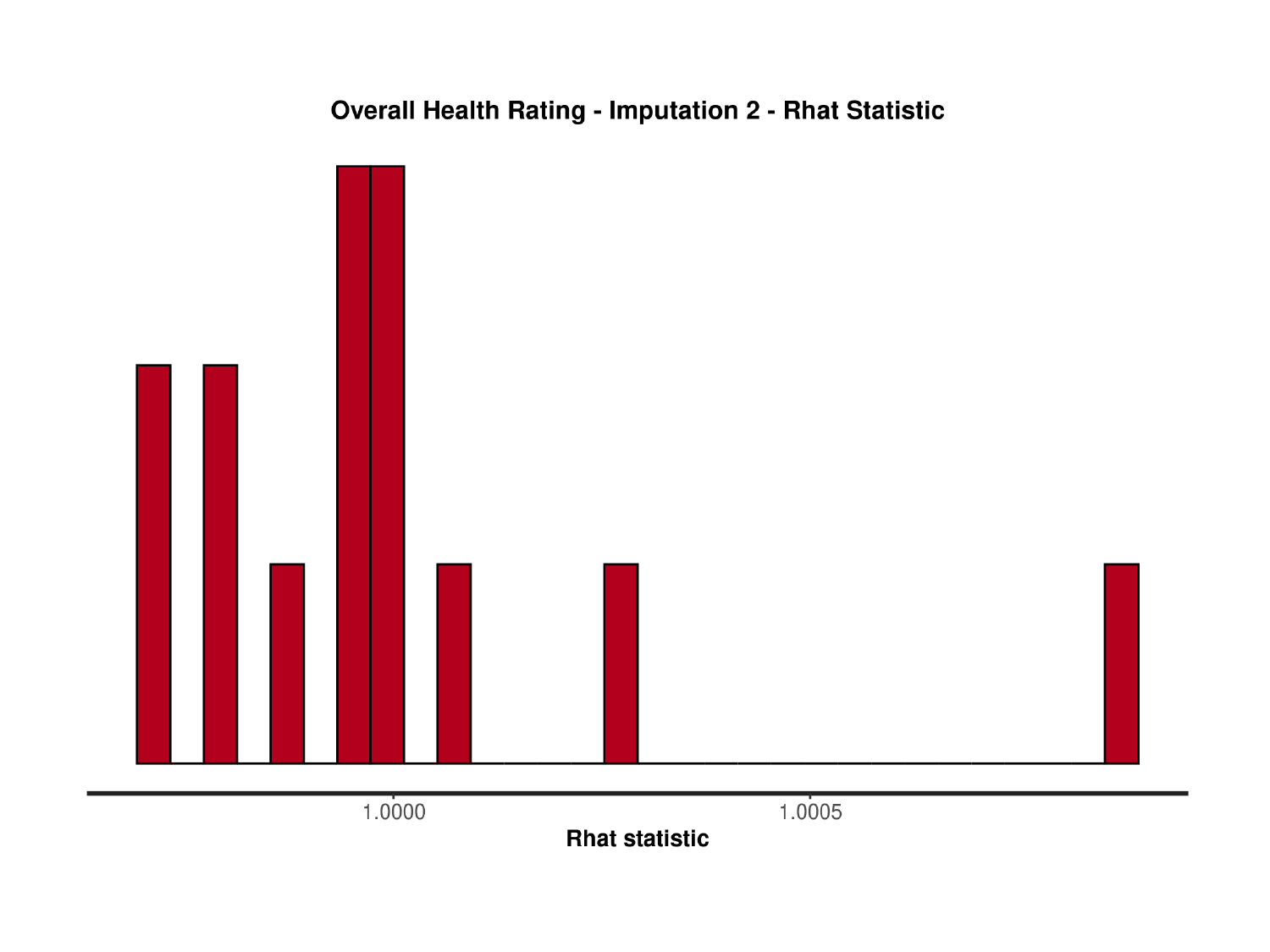

### Slide 81
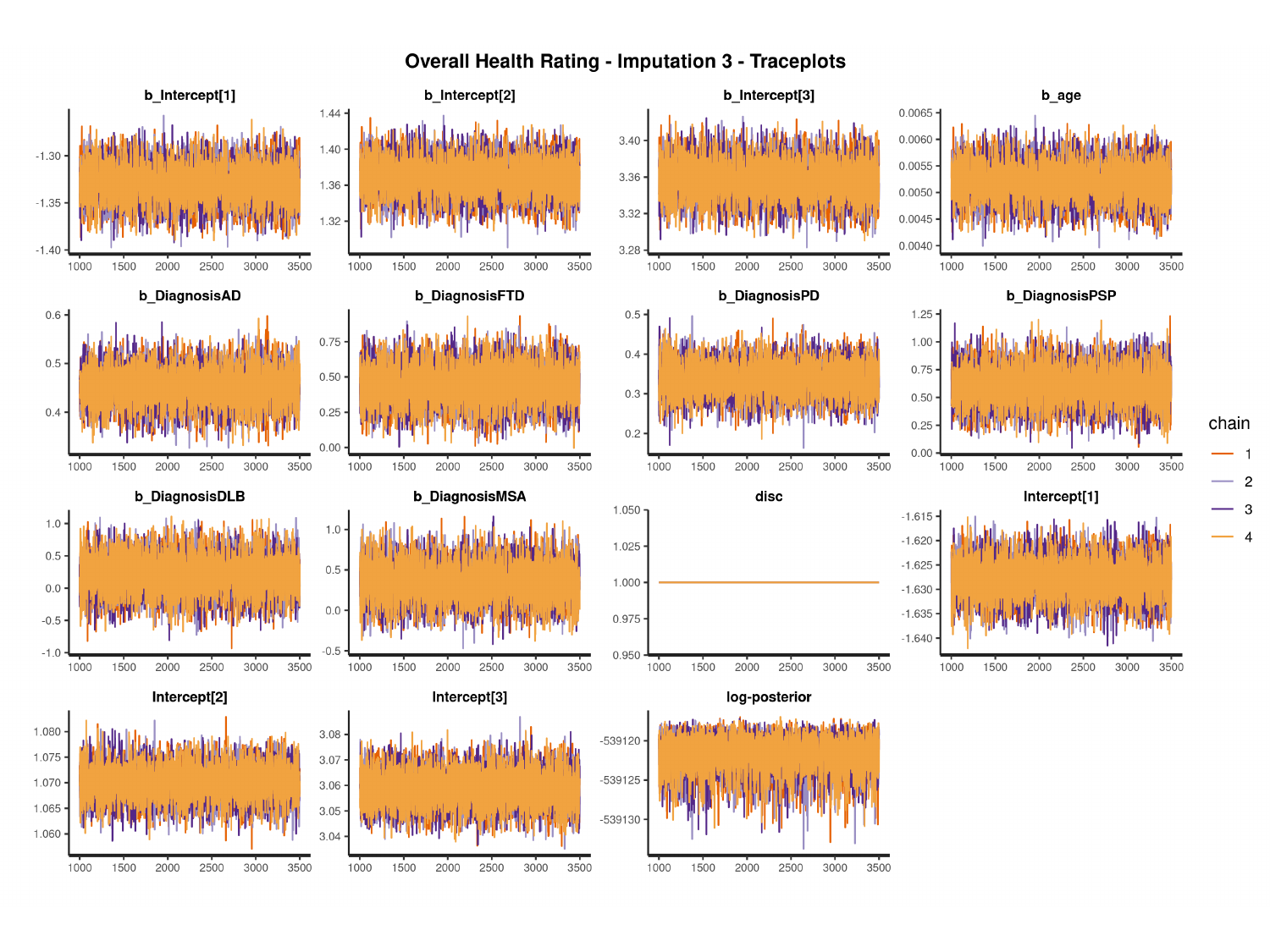

#

### Slide 82
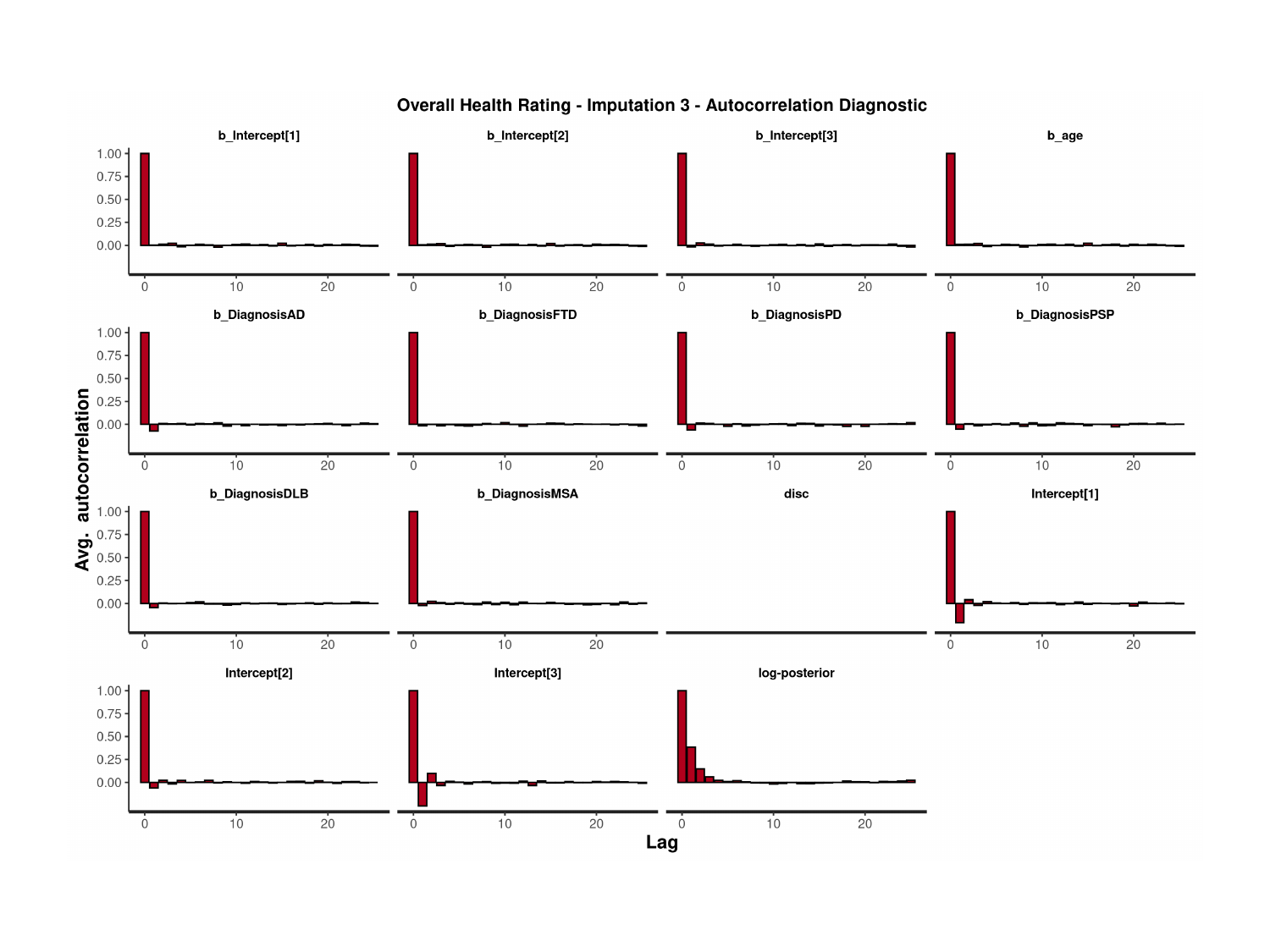

### Slide 83
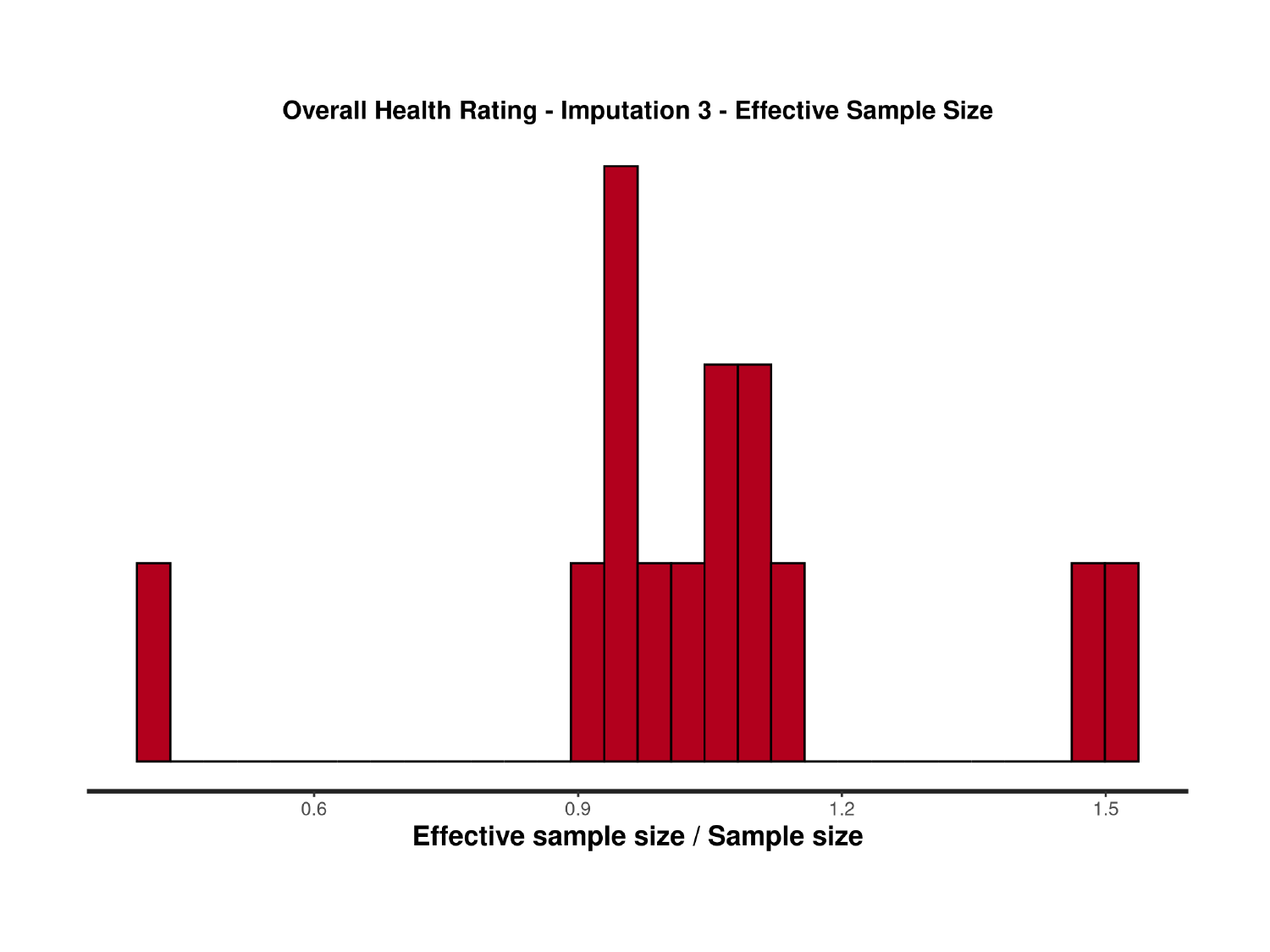

### Slide 84
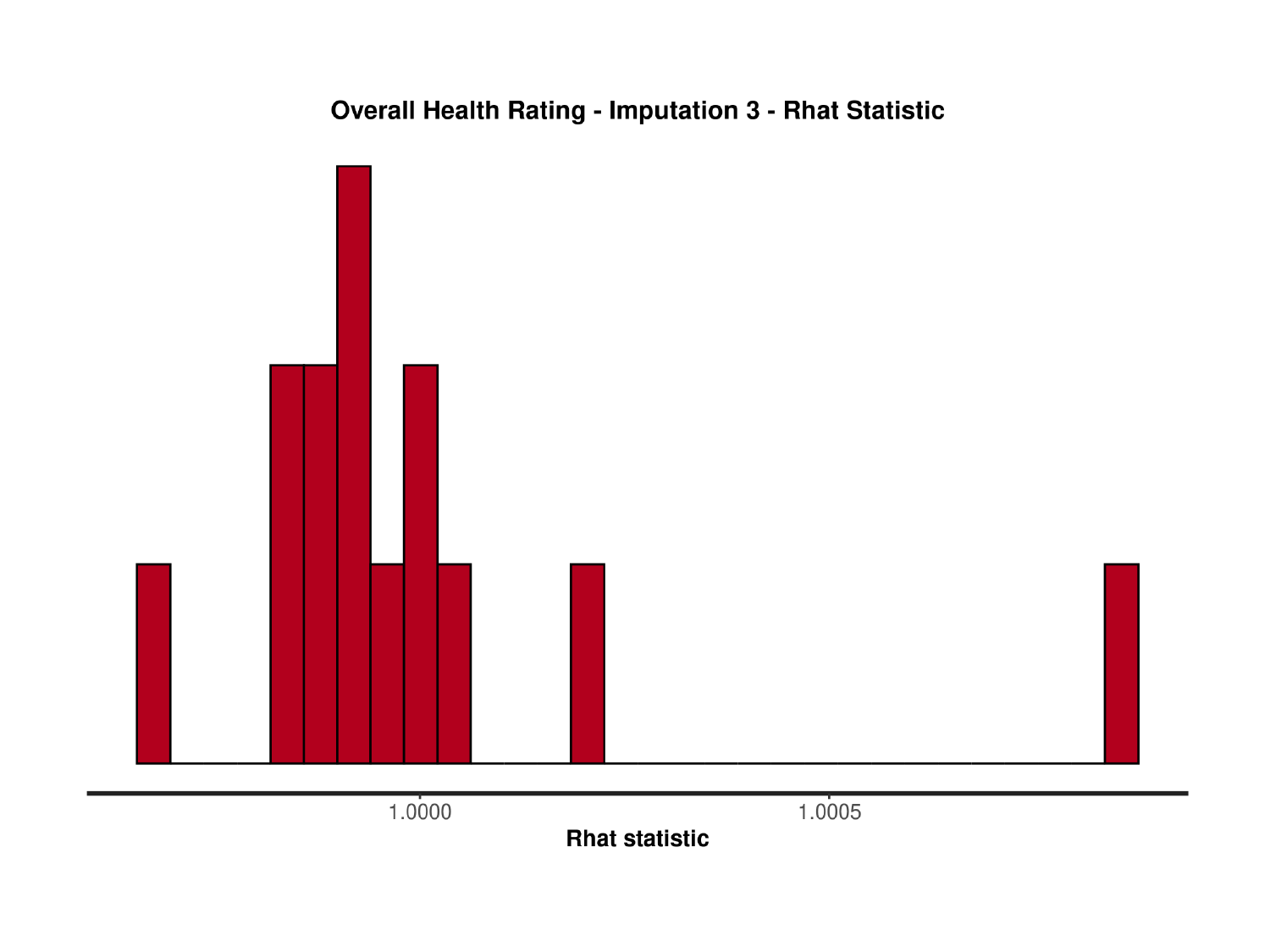

### Slide 85
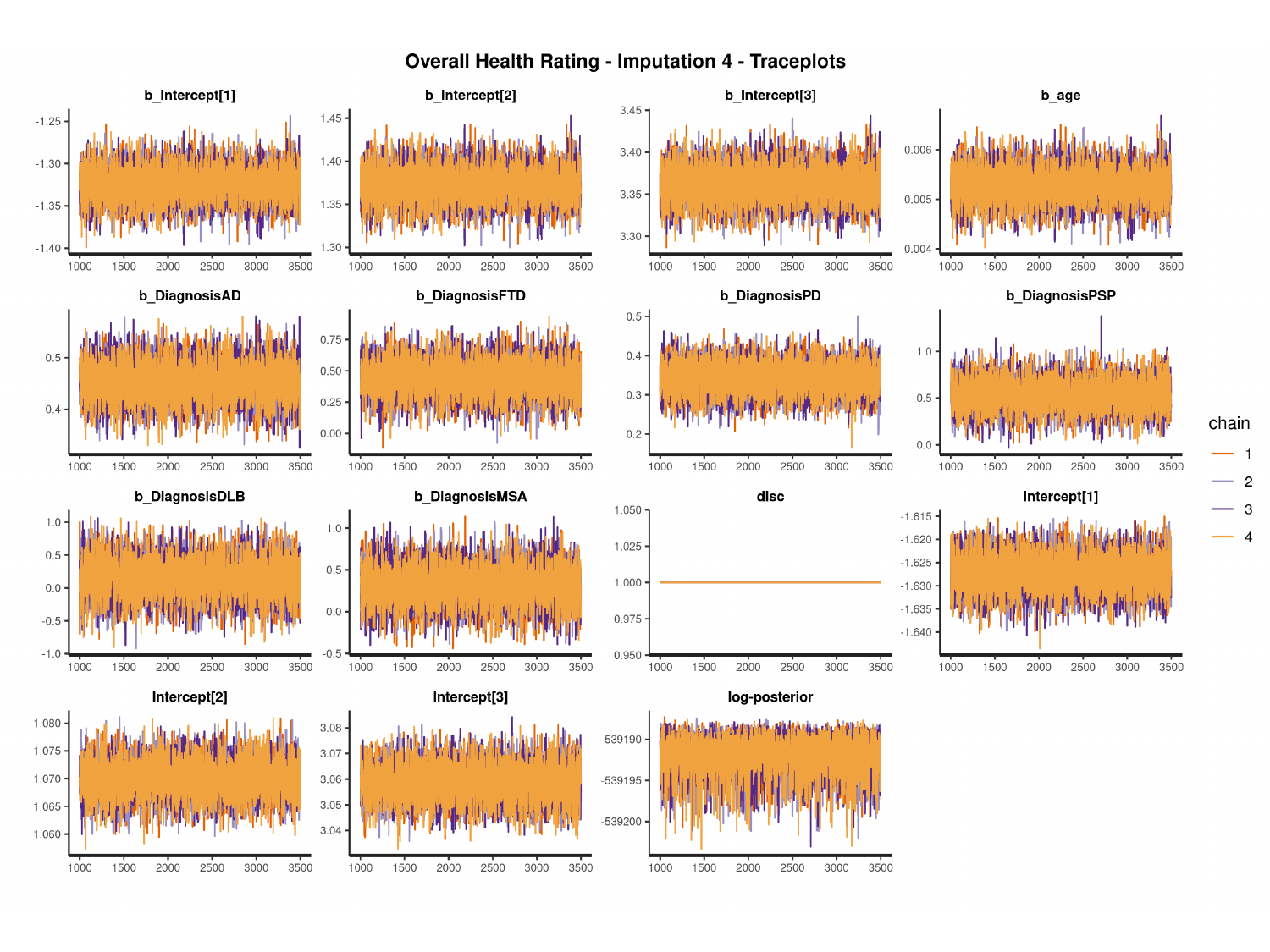

#

### Slide 86
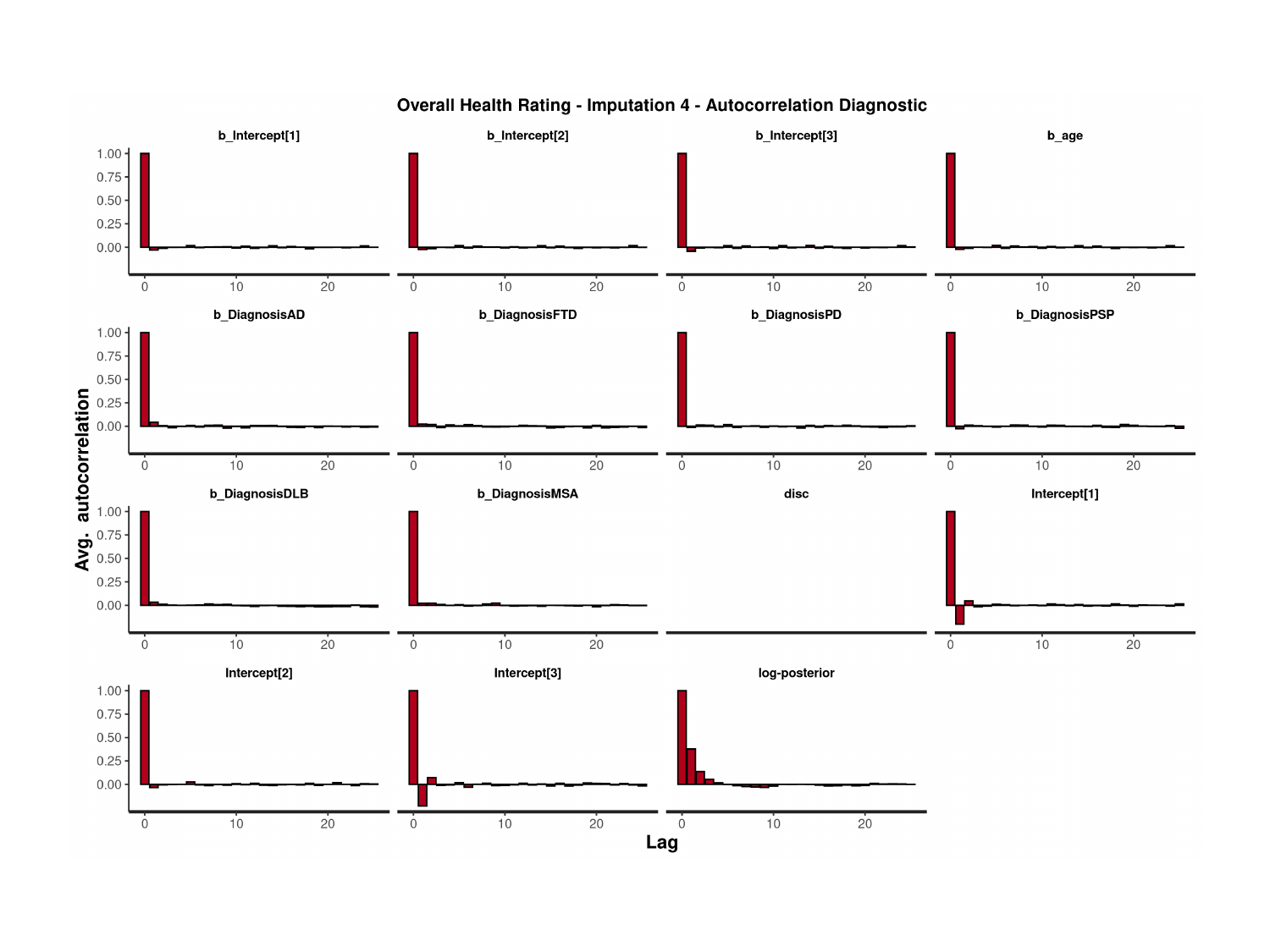

### Slide 87
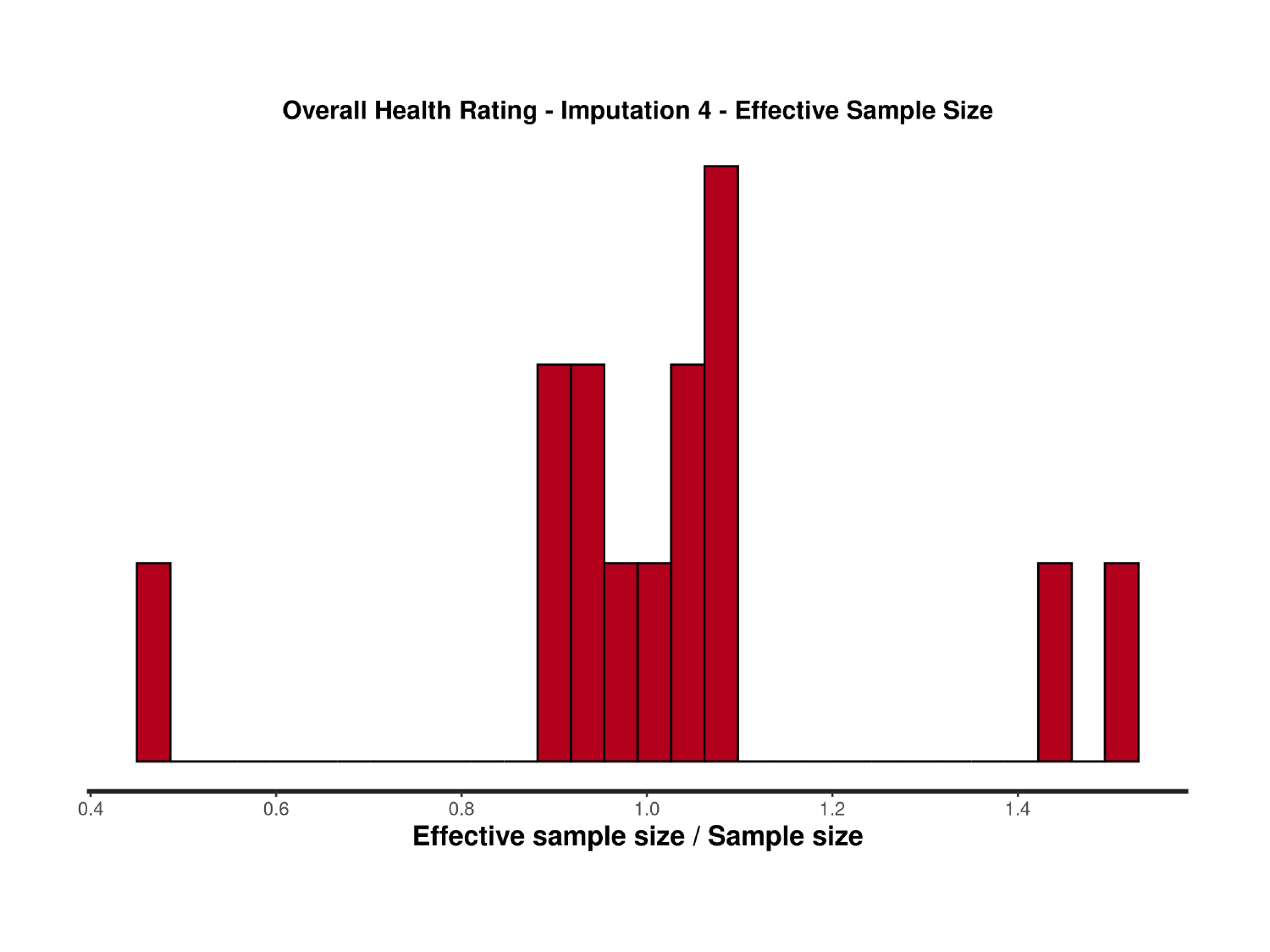

### Slide 88
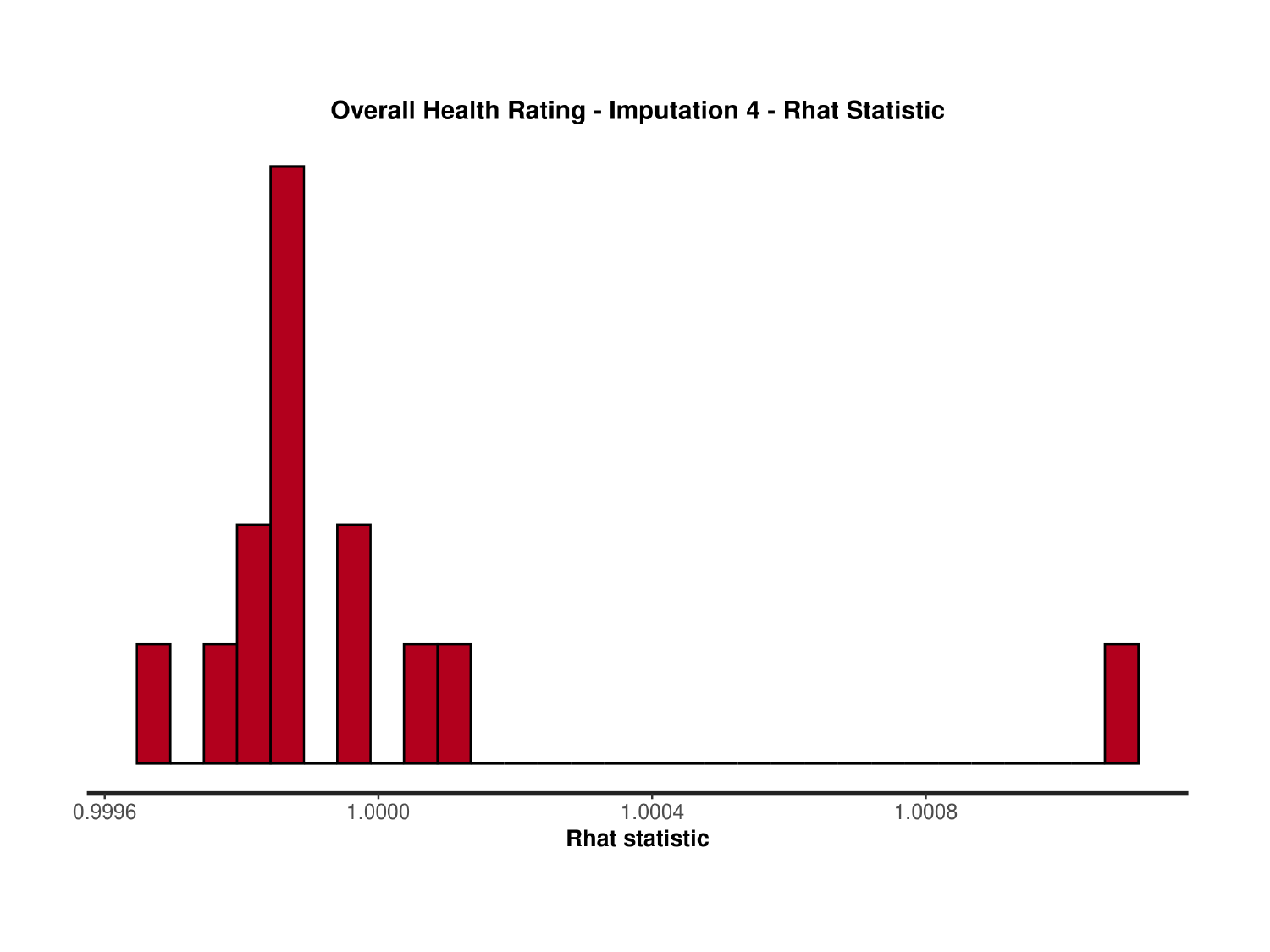

### Slide 89
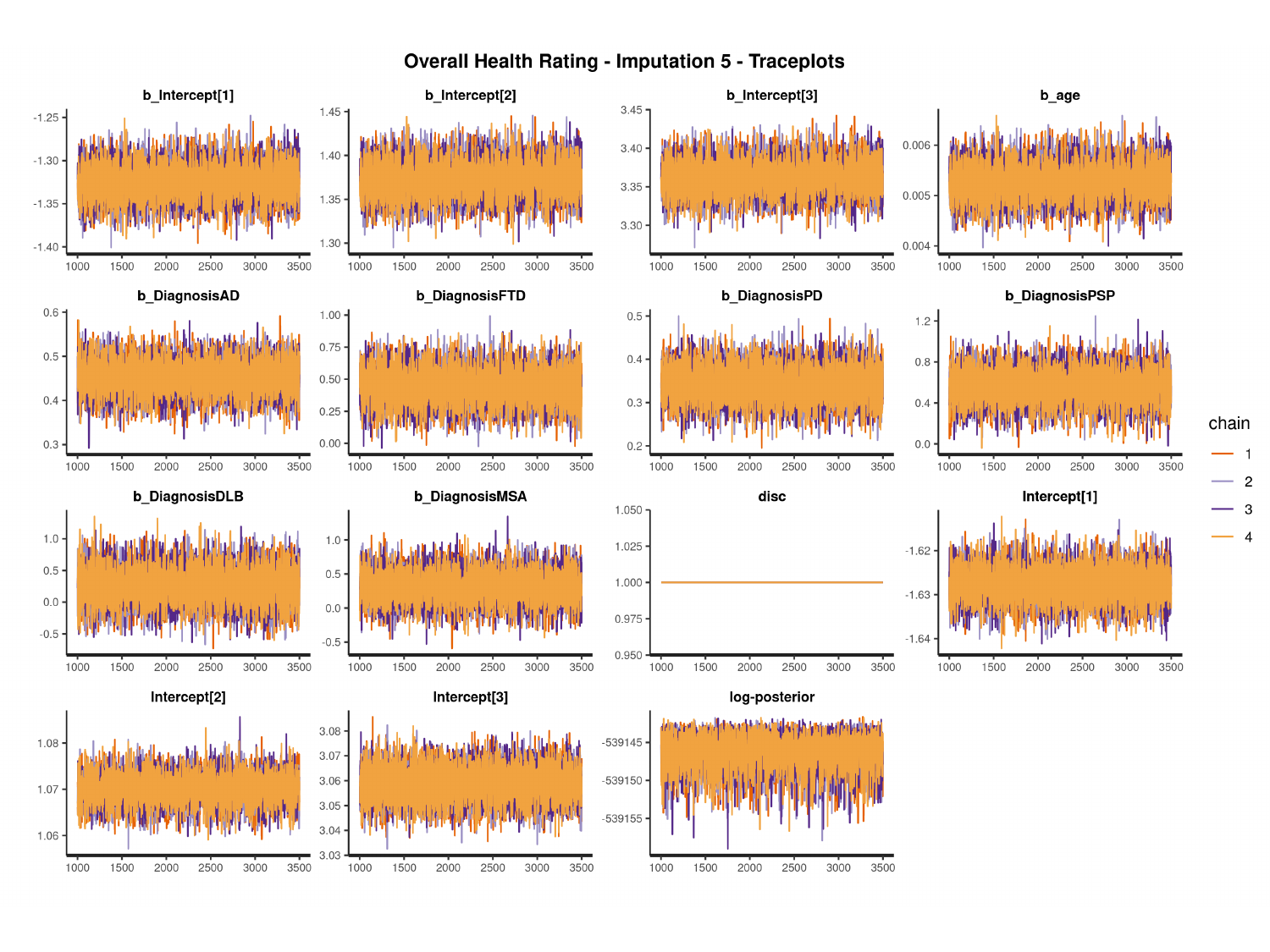

#

### Slide 90
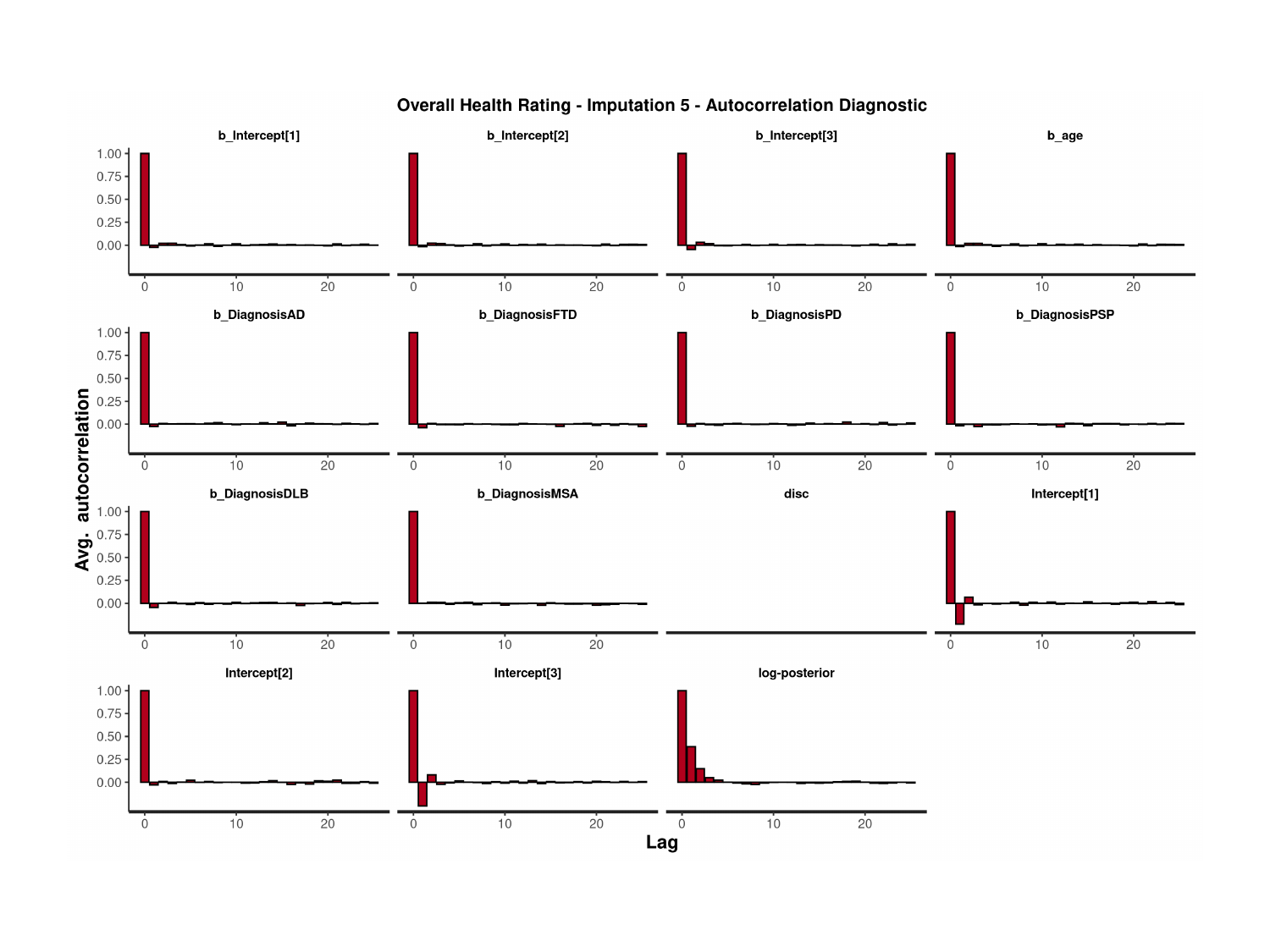

### Slide 91
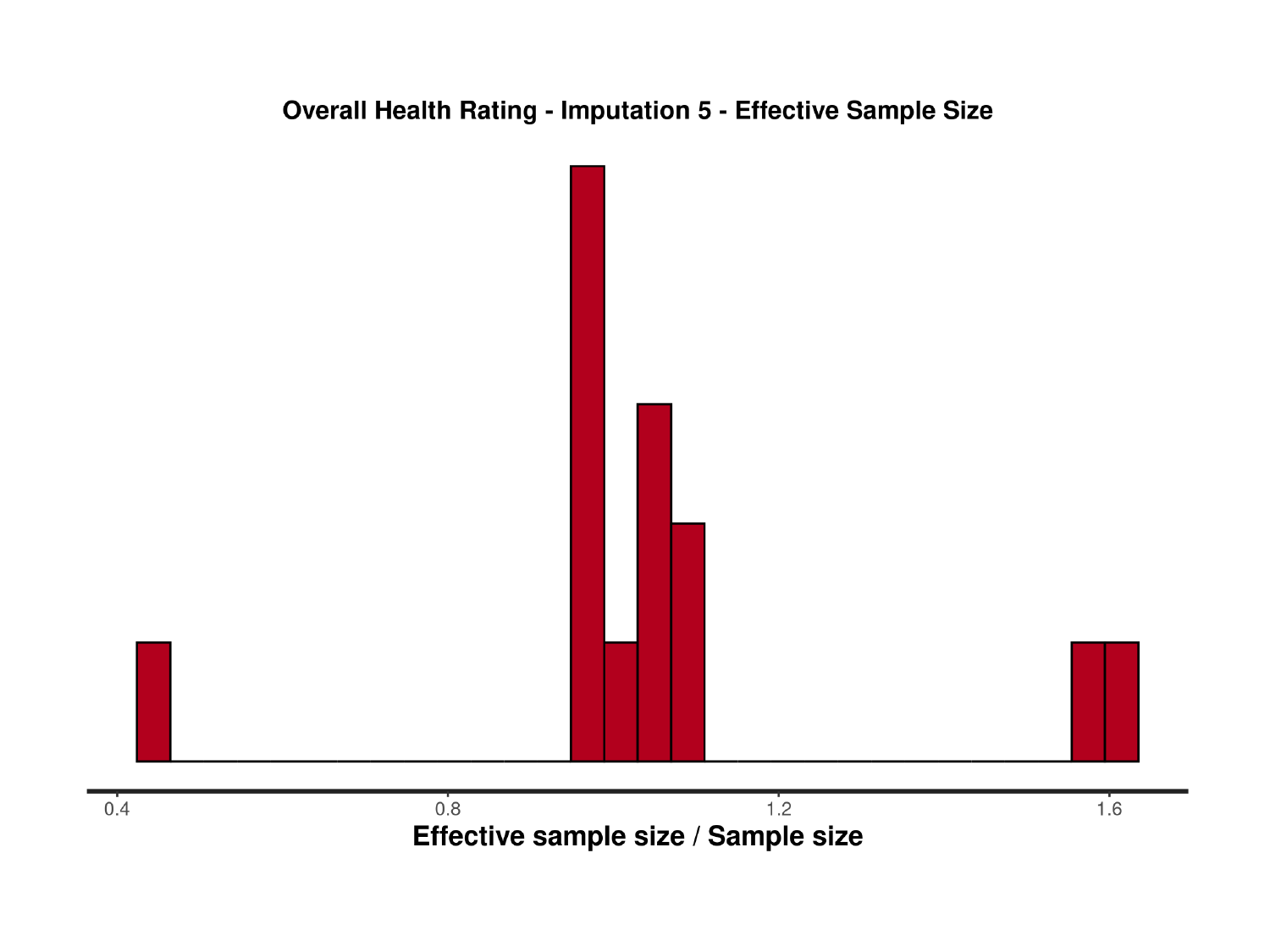

### Slide 92
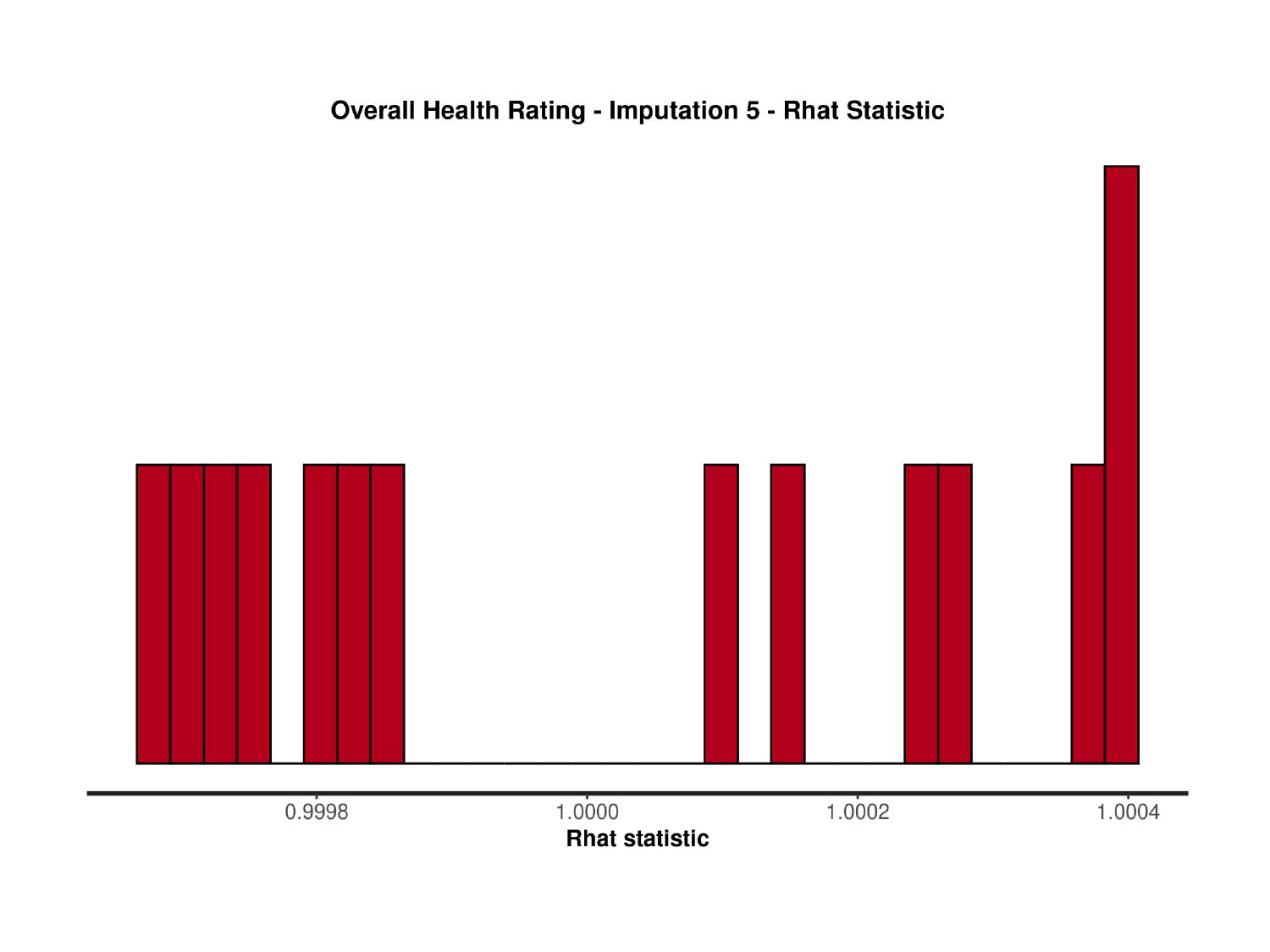

### Slide 93
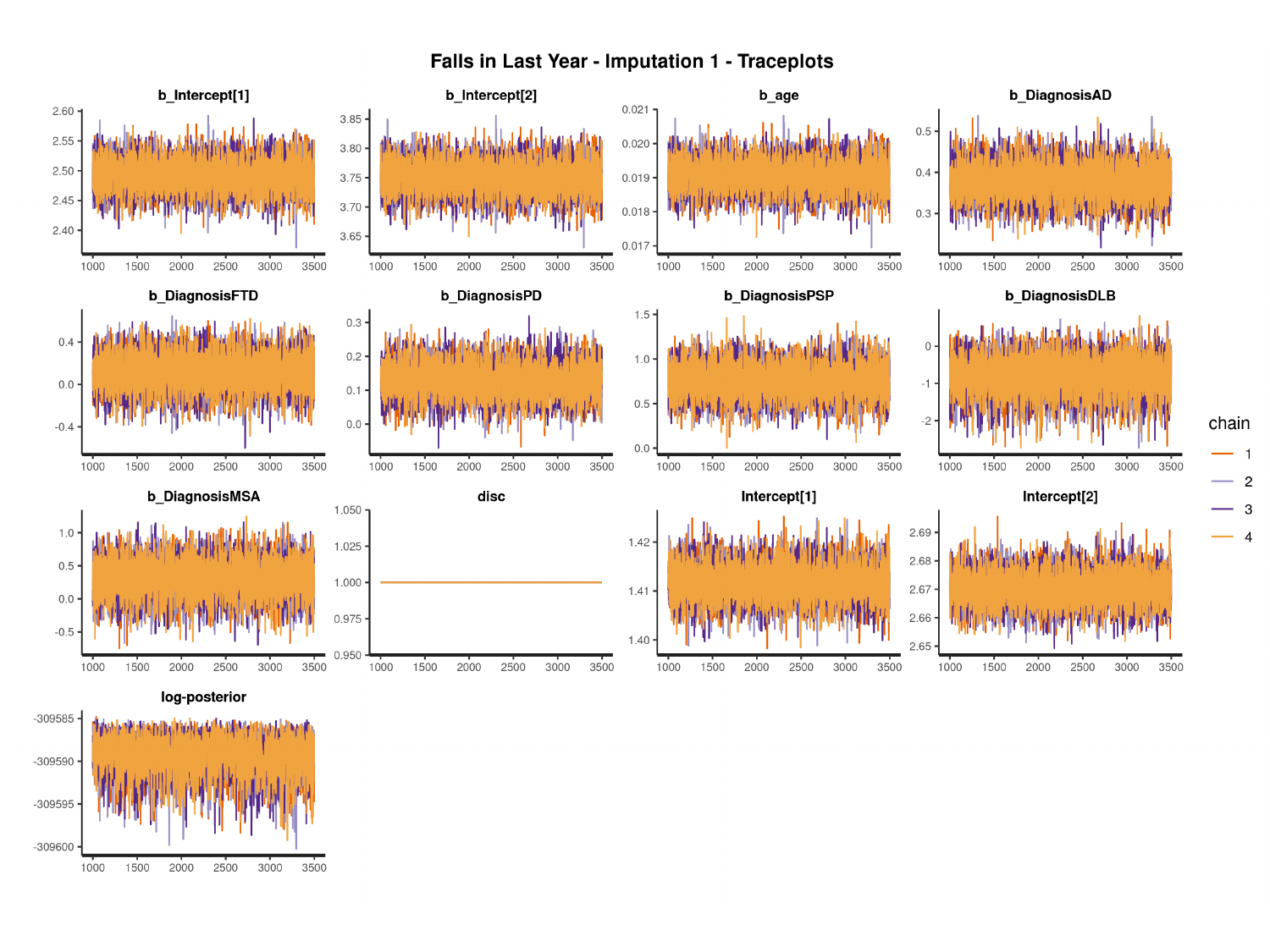

#

### Slide 94
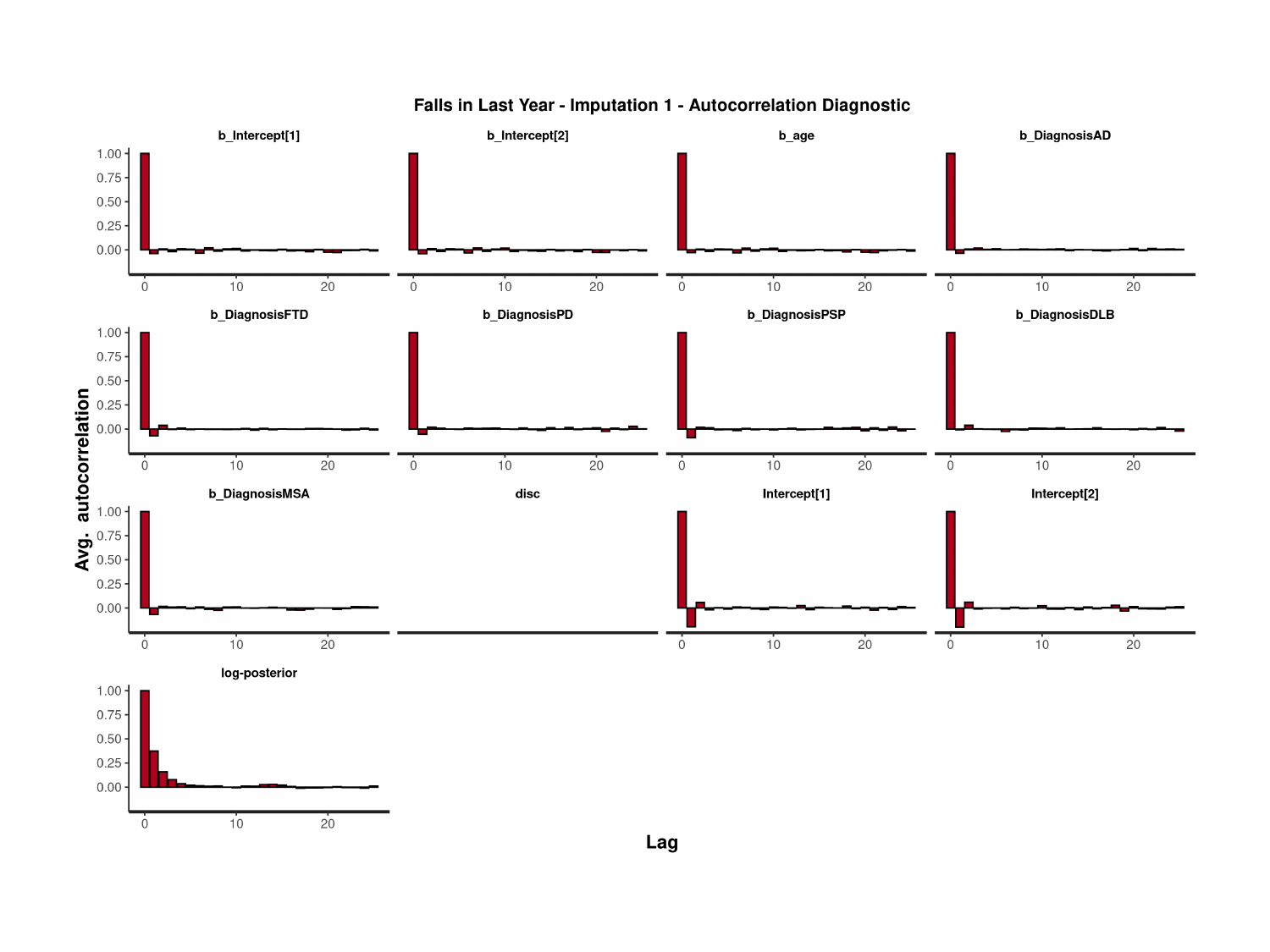

### Slide 95
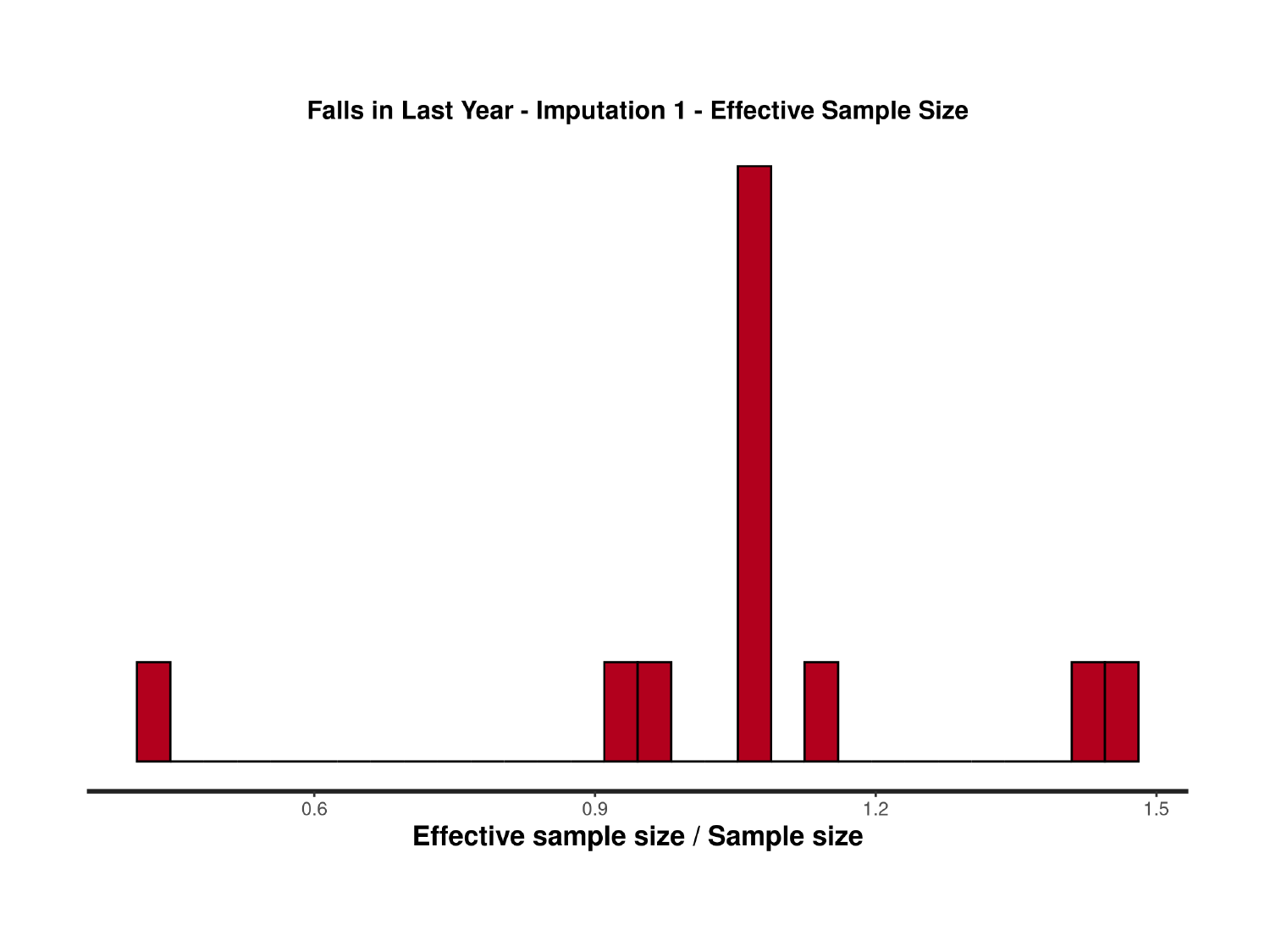

### Slide 96
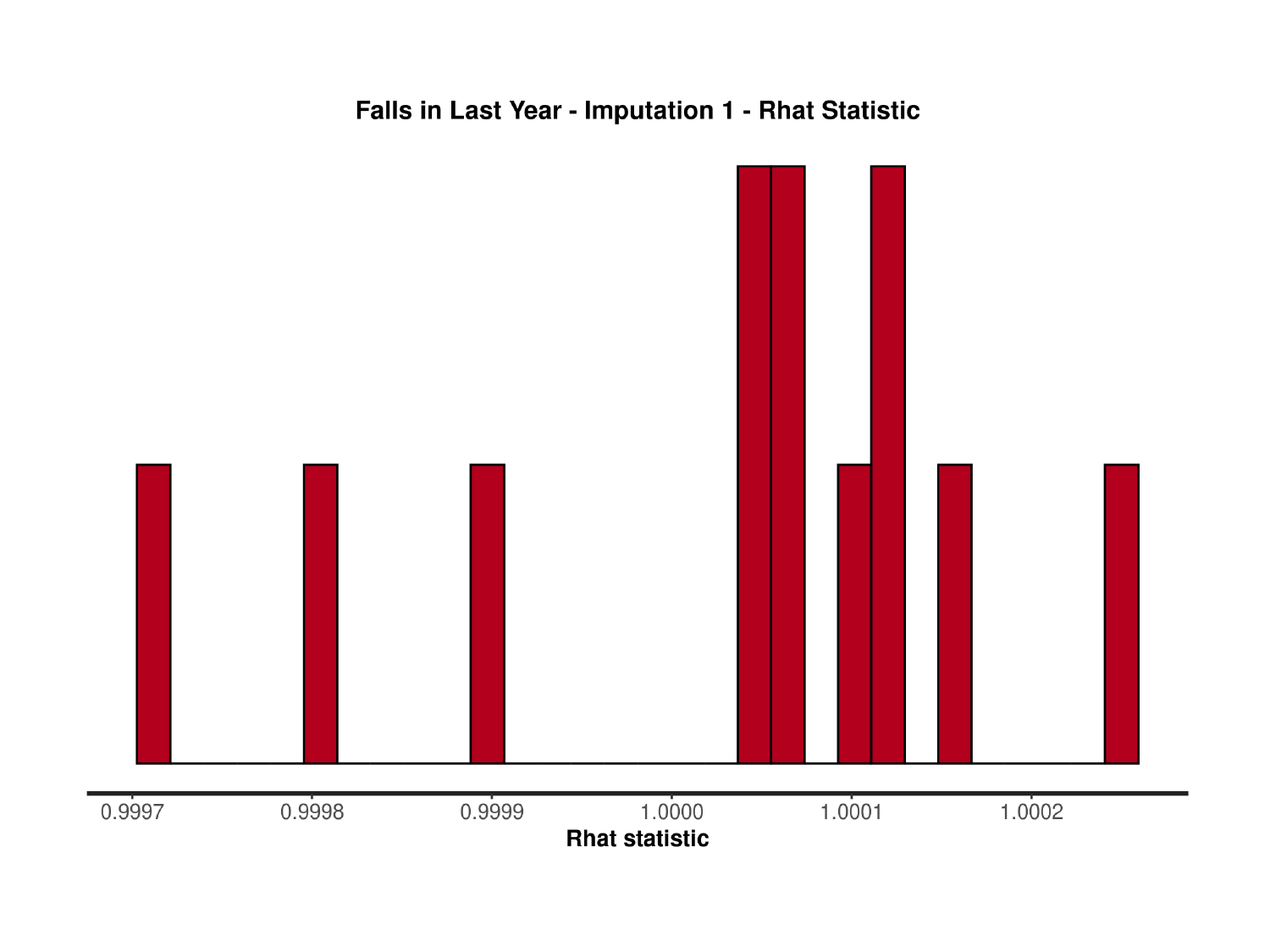

### Slide 97
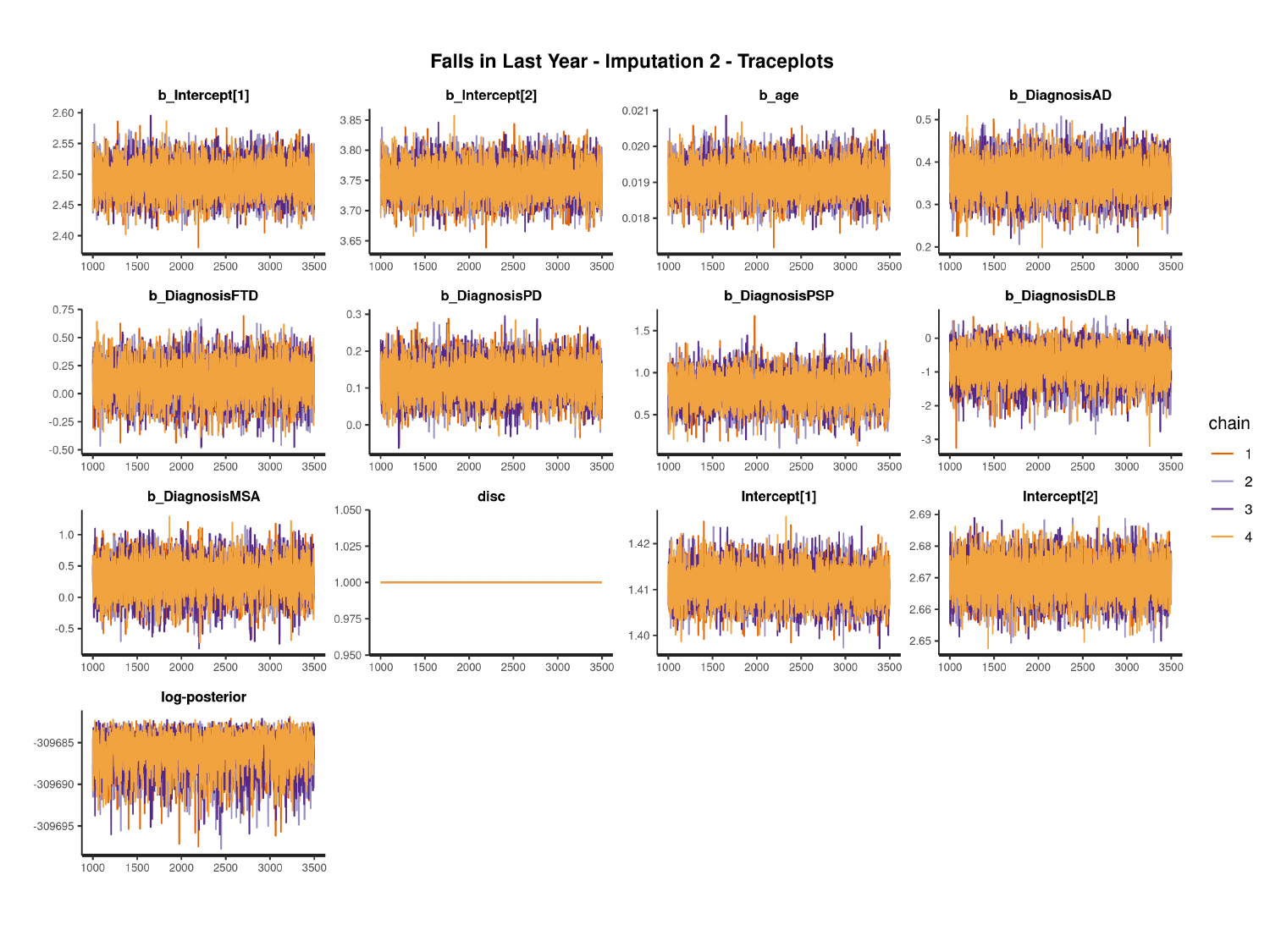

#

### Slide 98
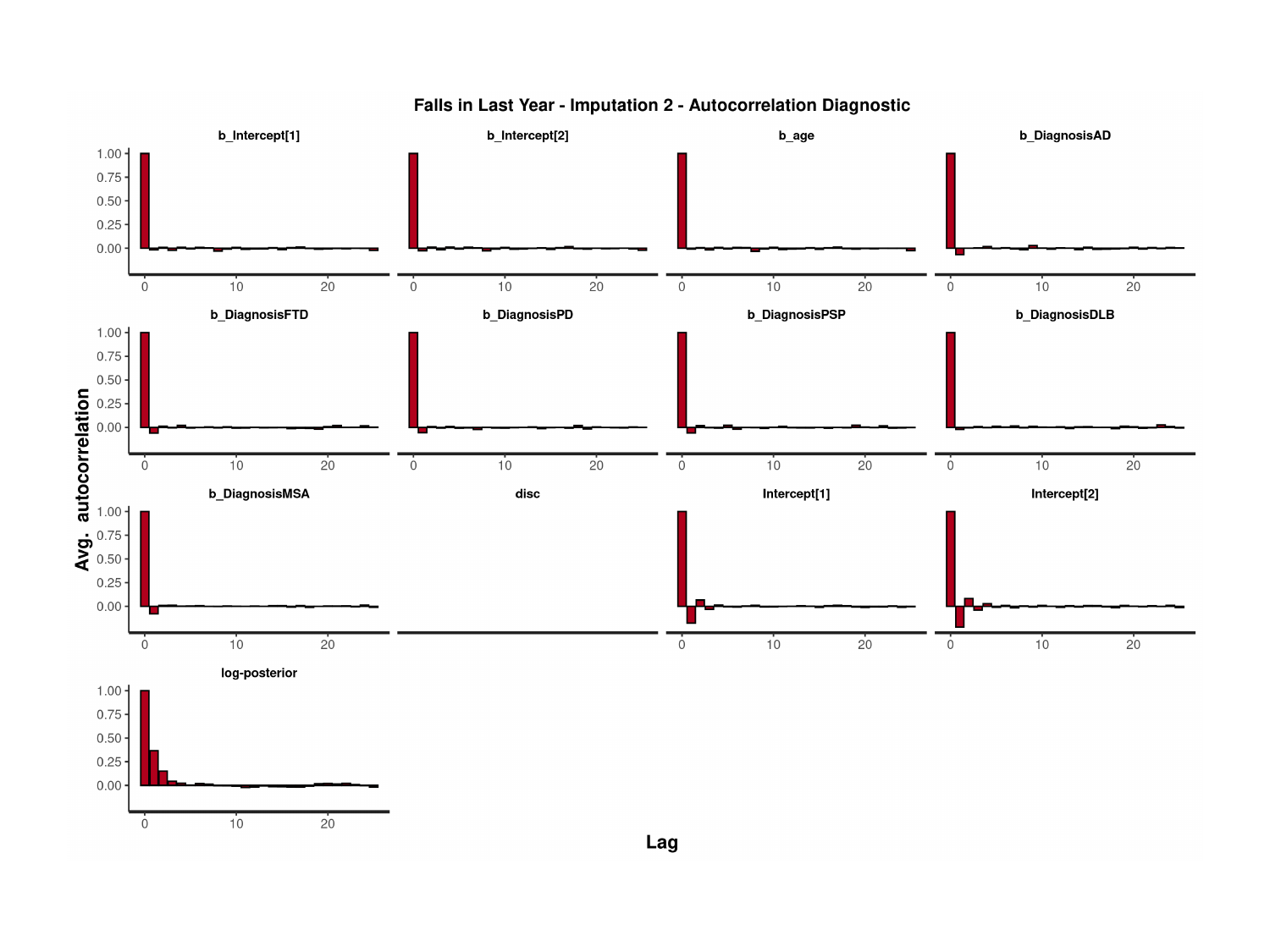

### Slide 99
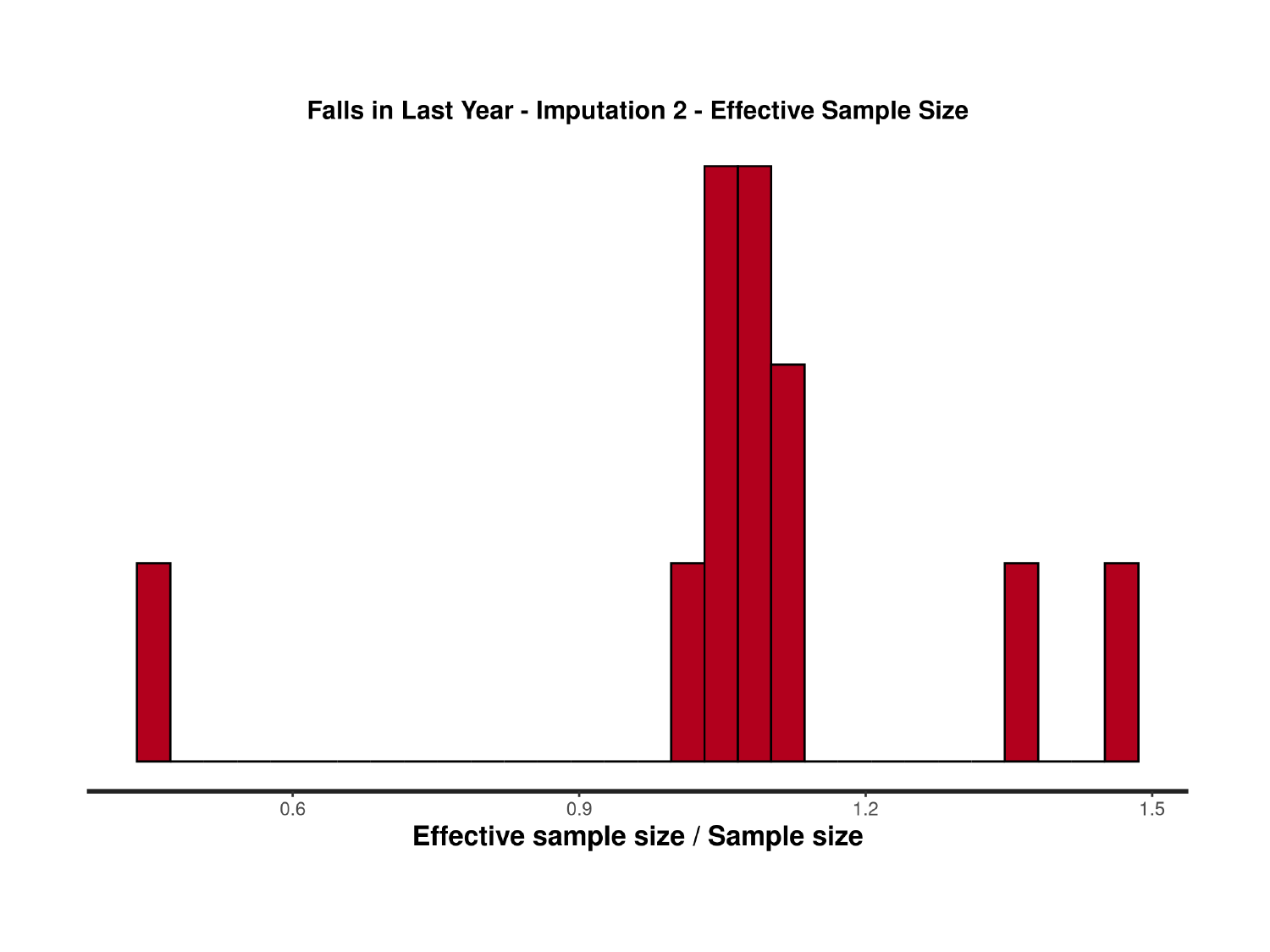

### Slide 100
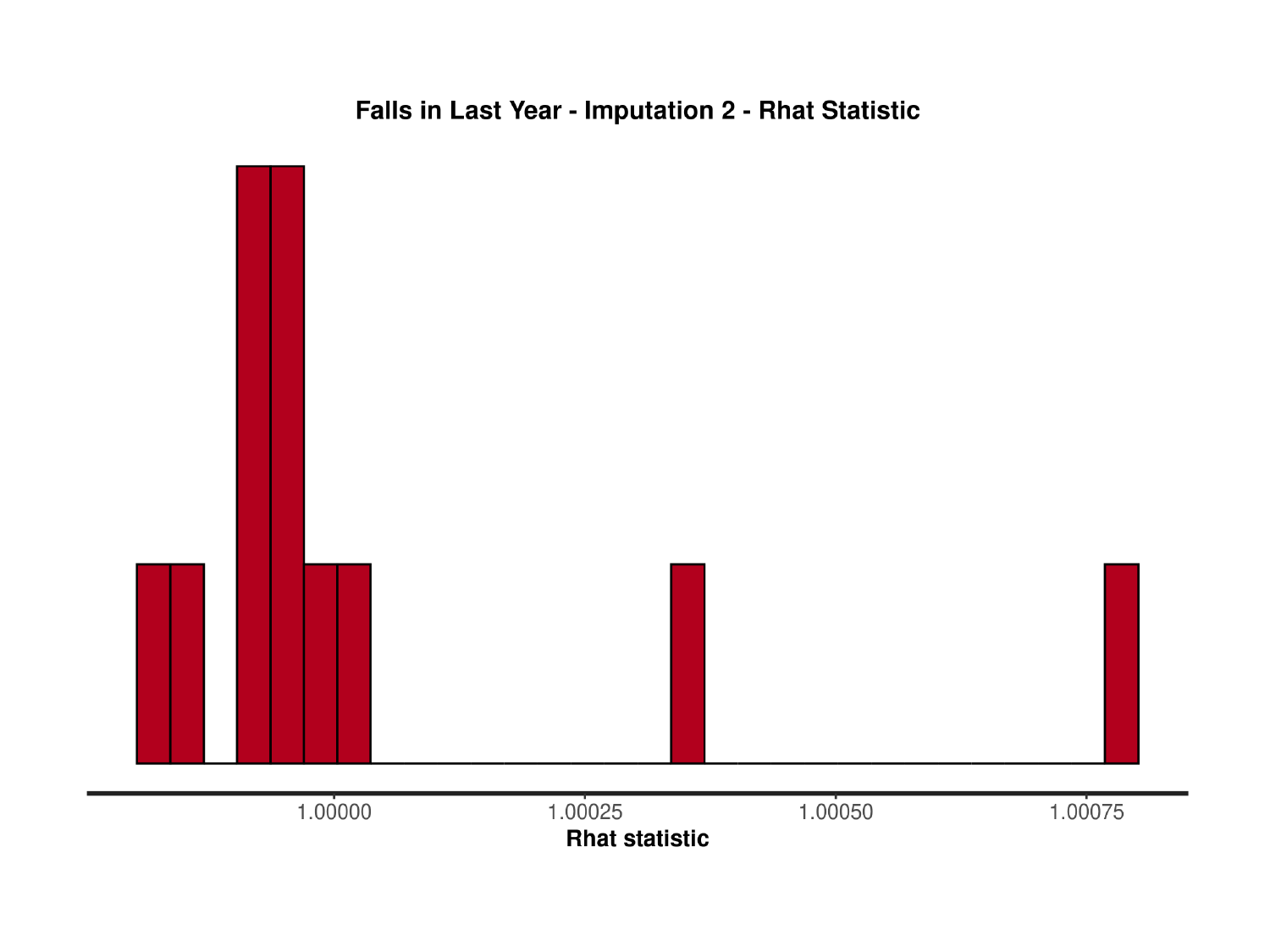

### Slide 101
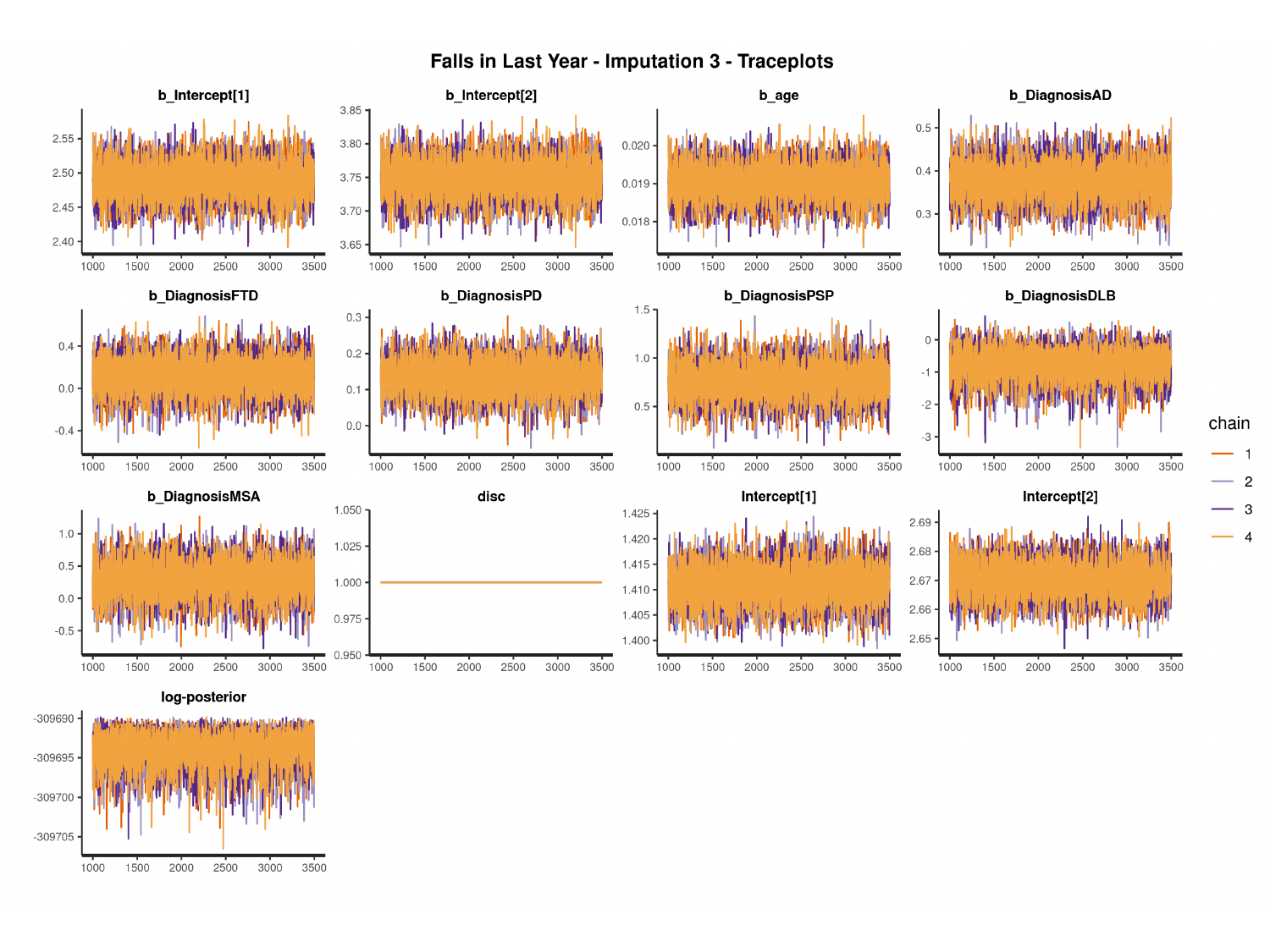

#

### Slide 102
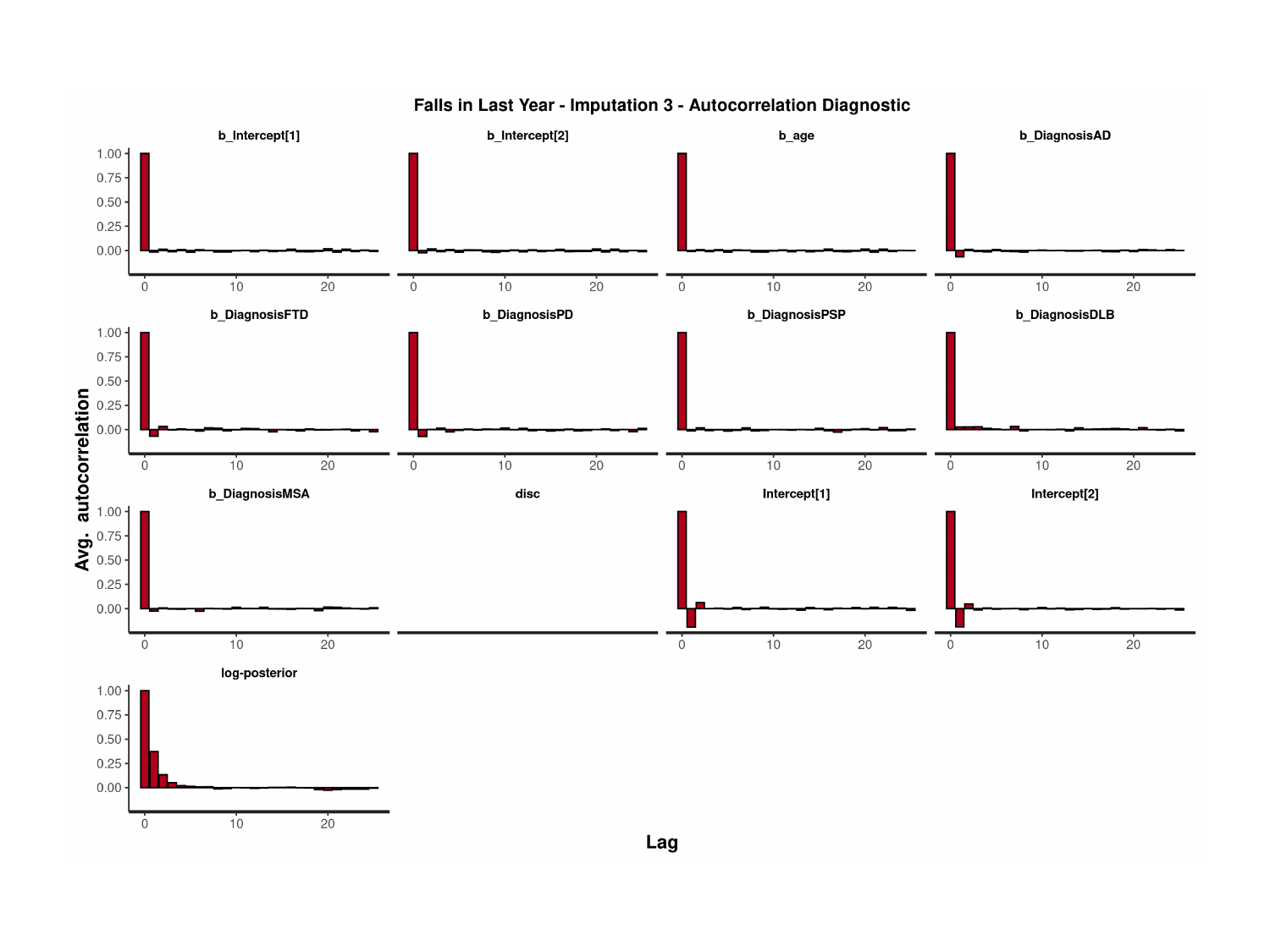

### Slide 103
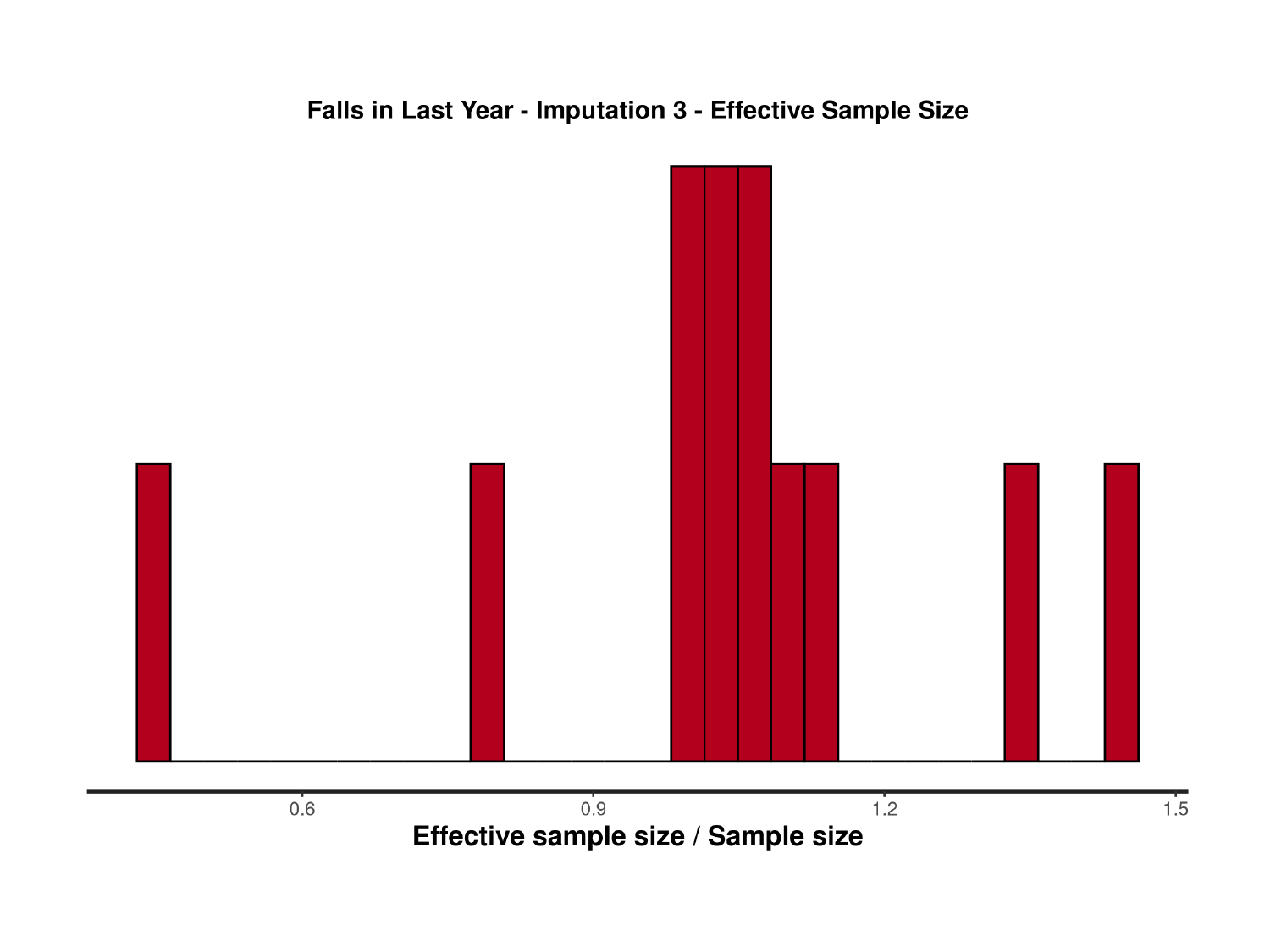

### Slide 104
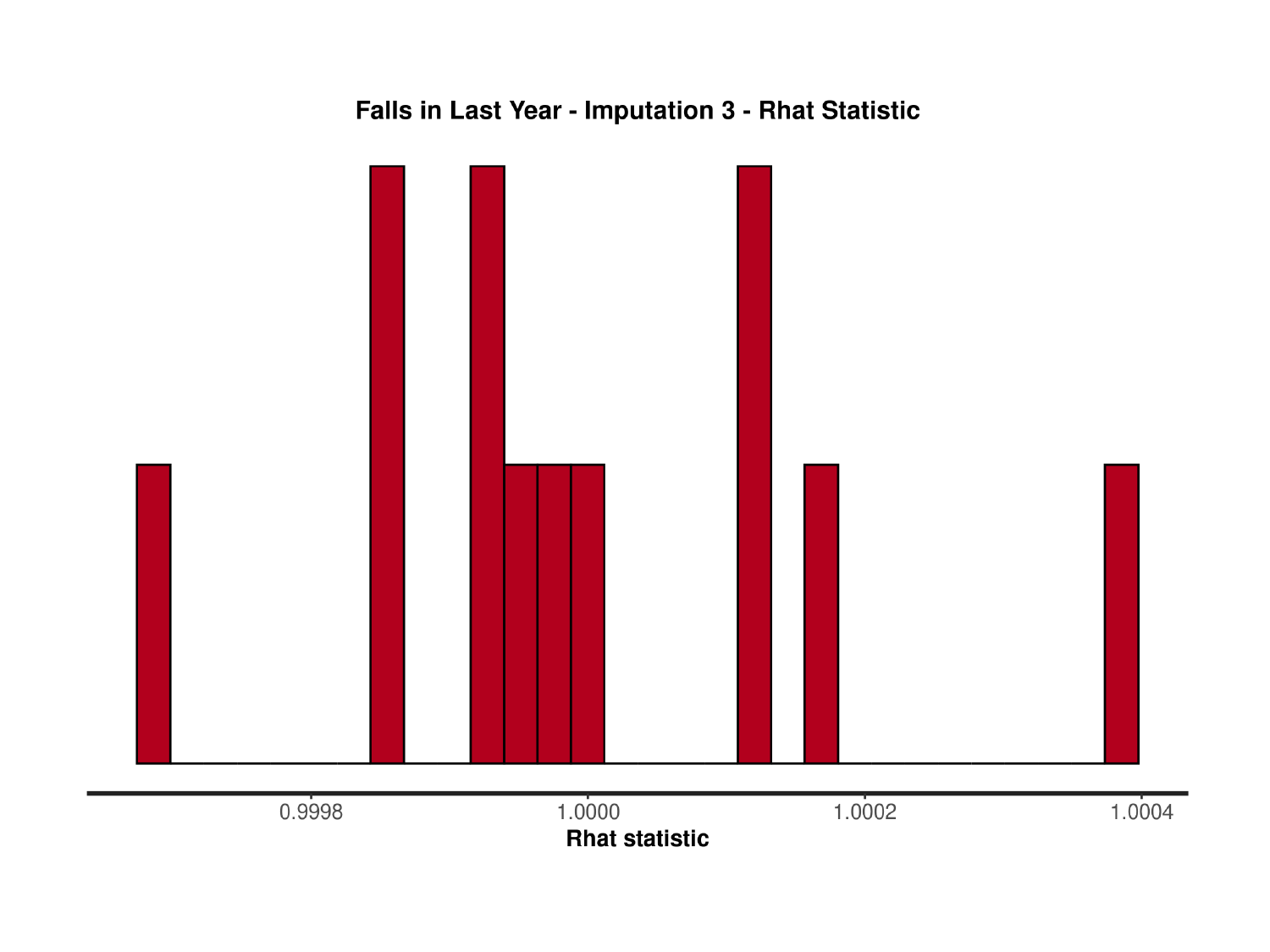

### Slide 105
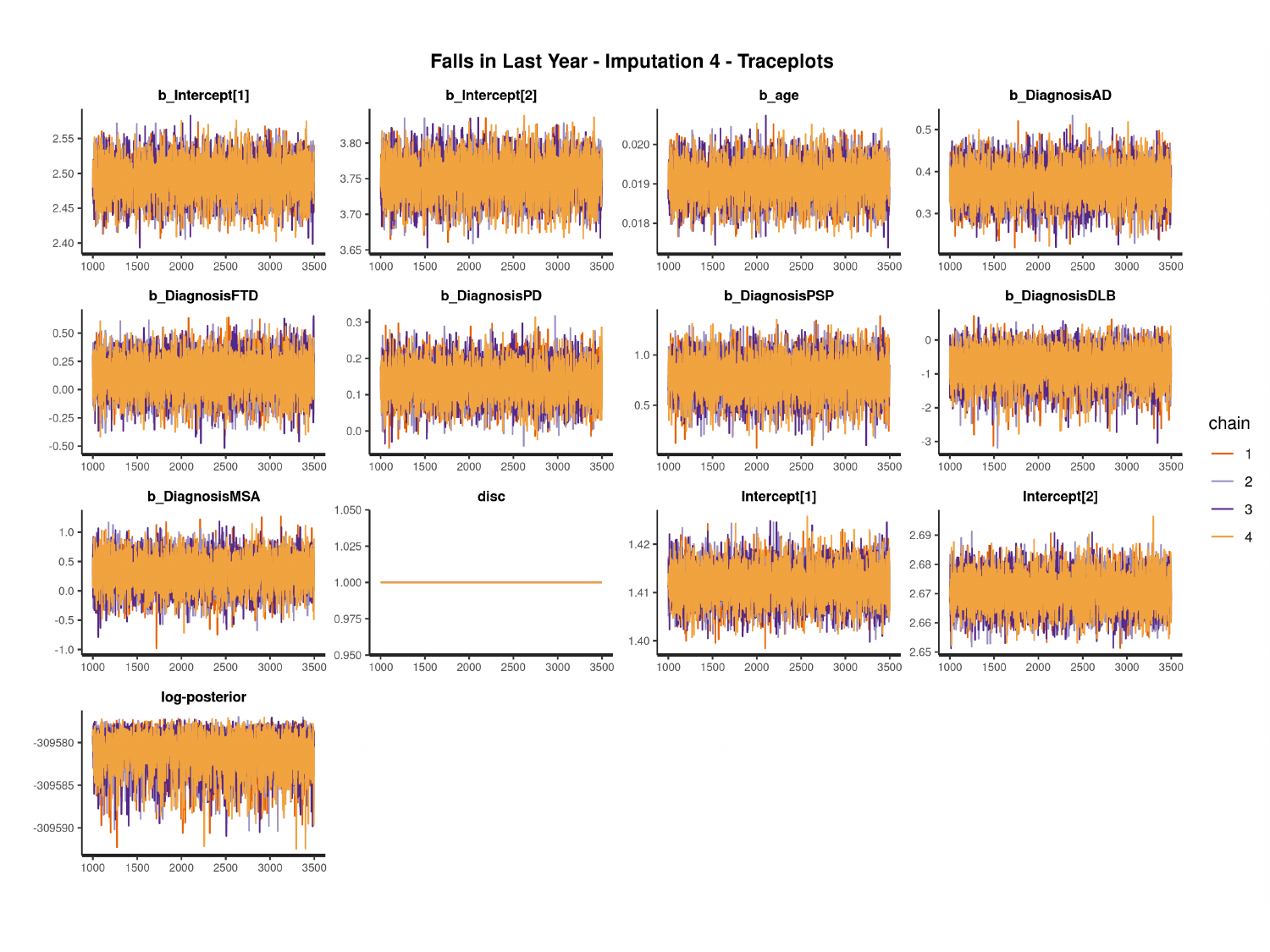

#

### Slide 106
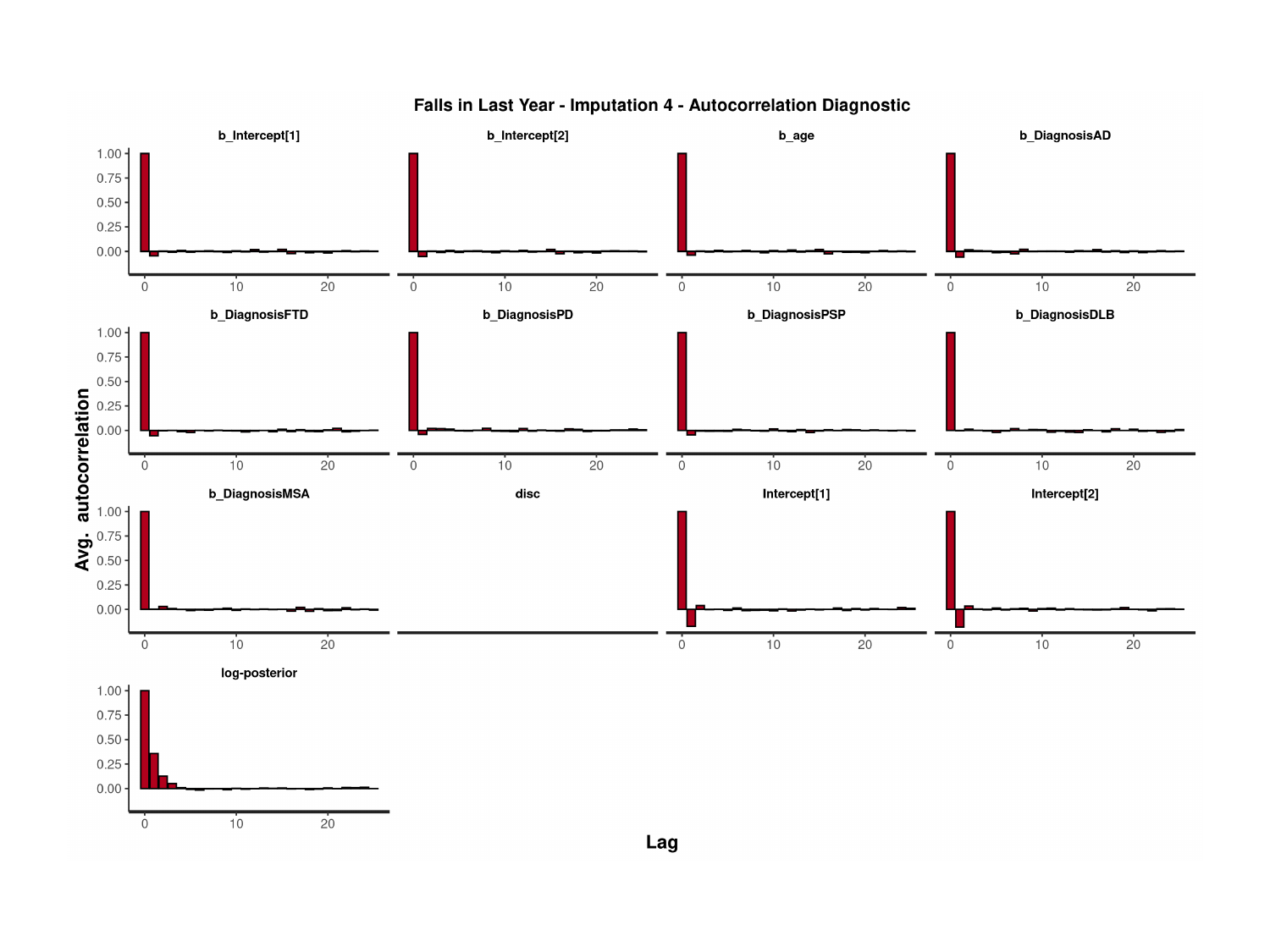

### Slide 107
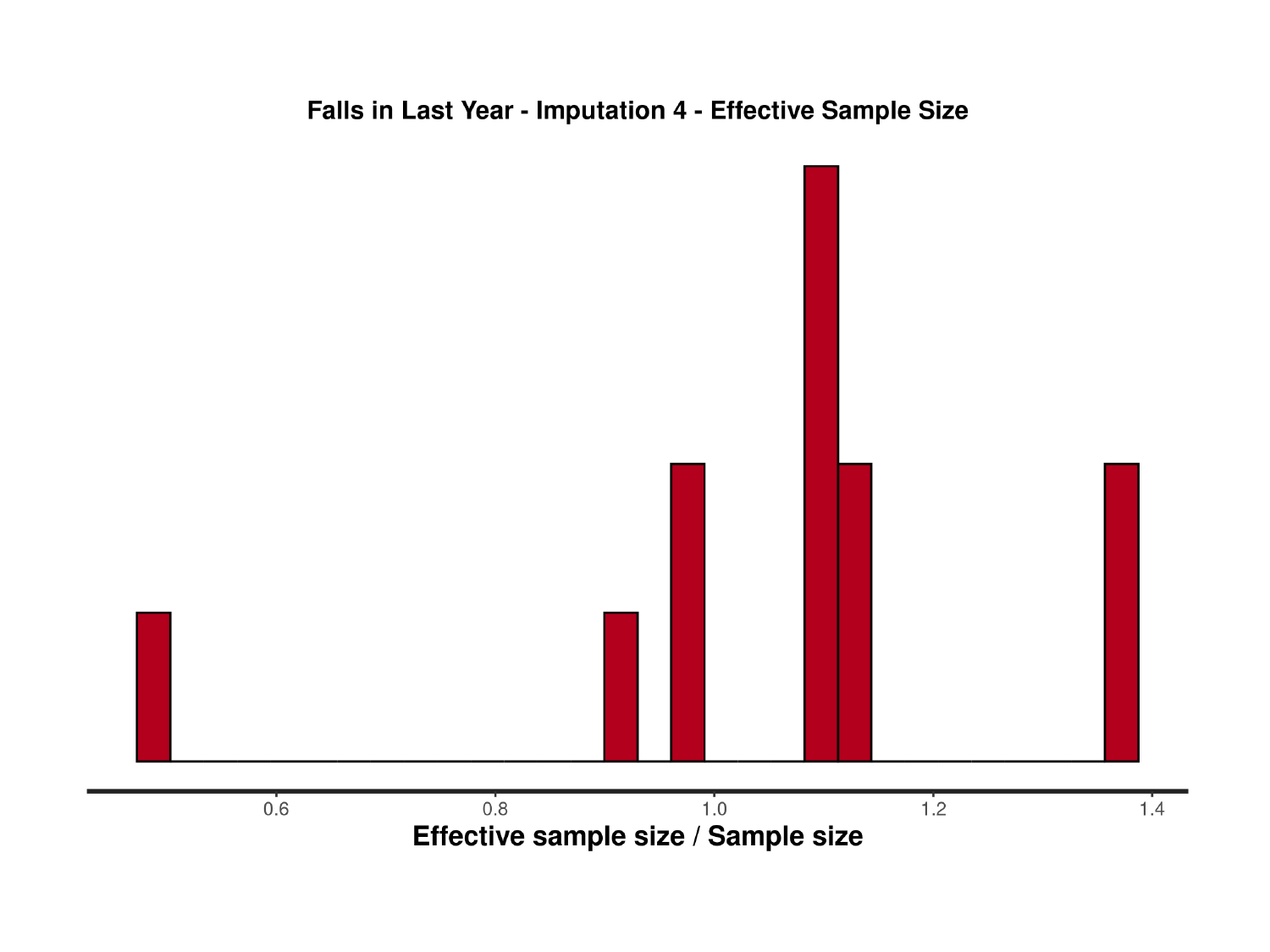

### Slide 108
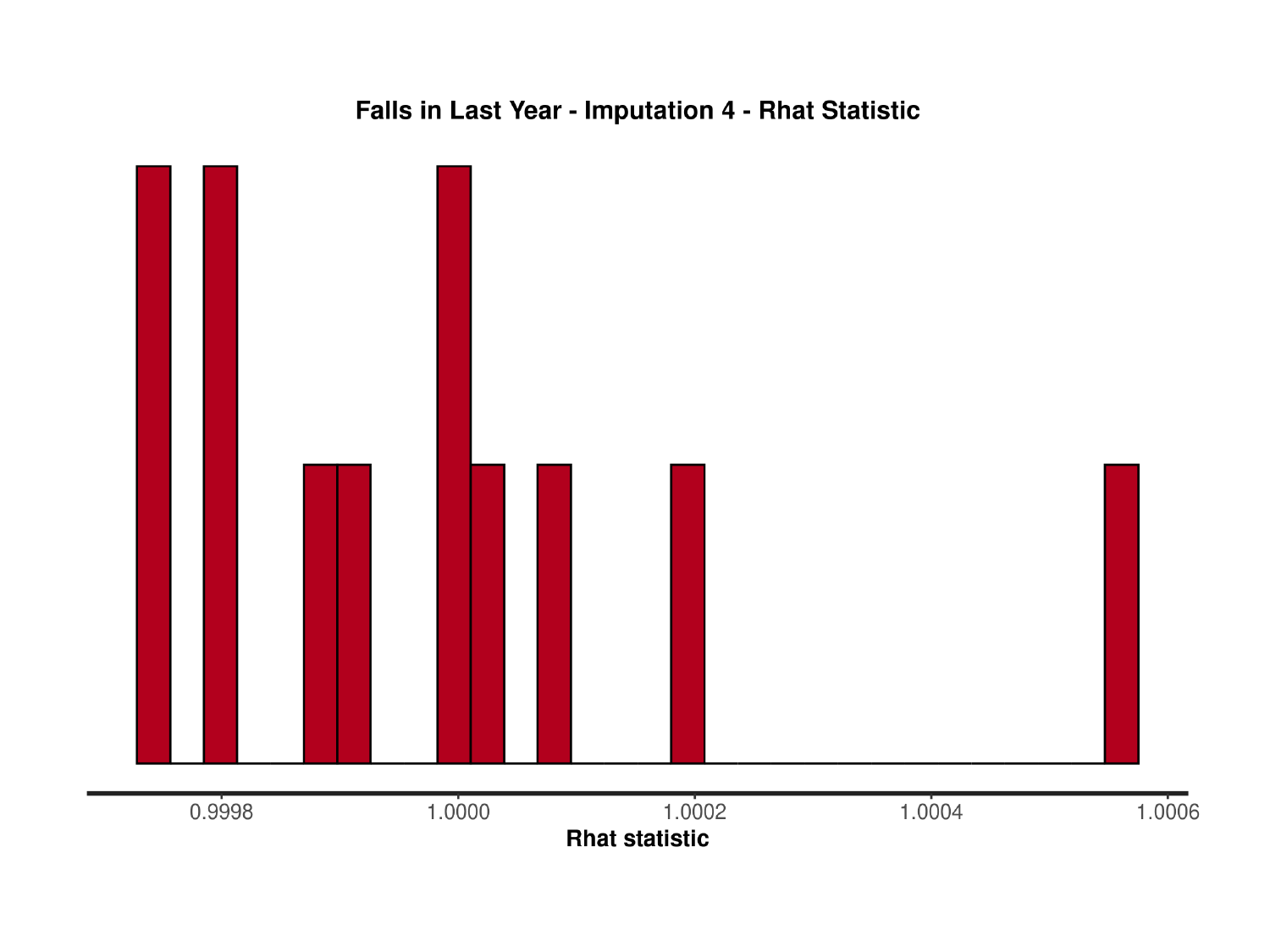

### Slide 109
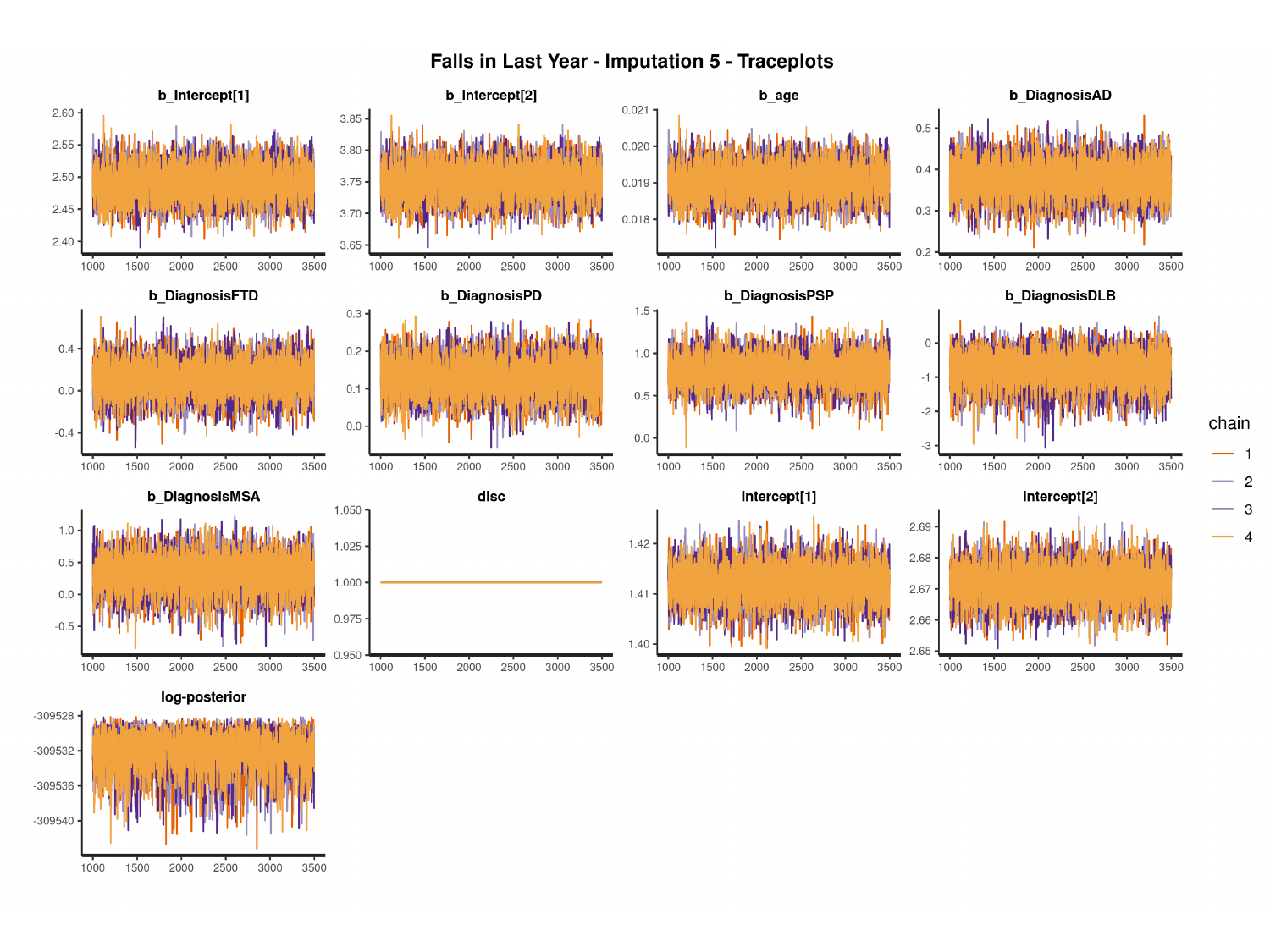

#

### Slide 110
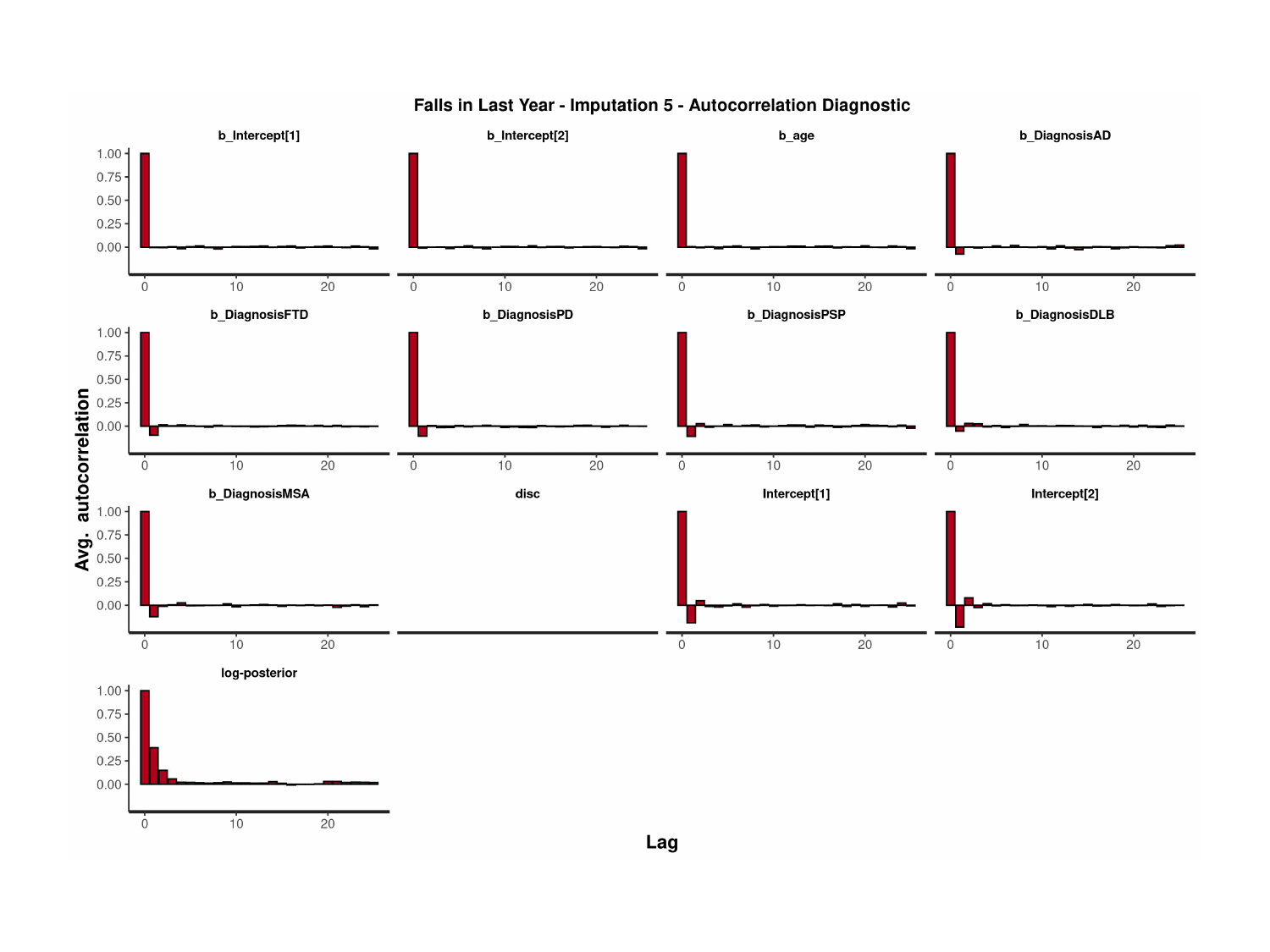

### Slide 111
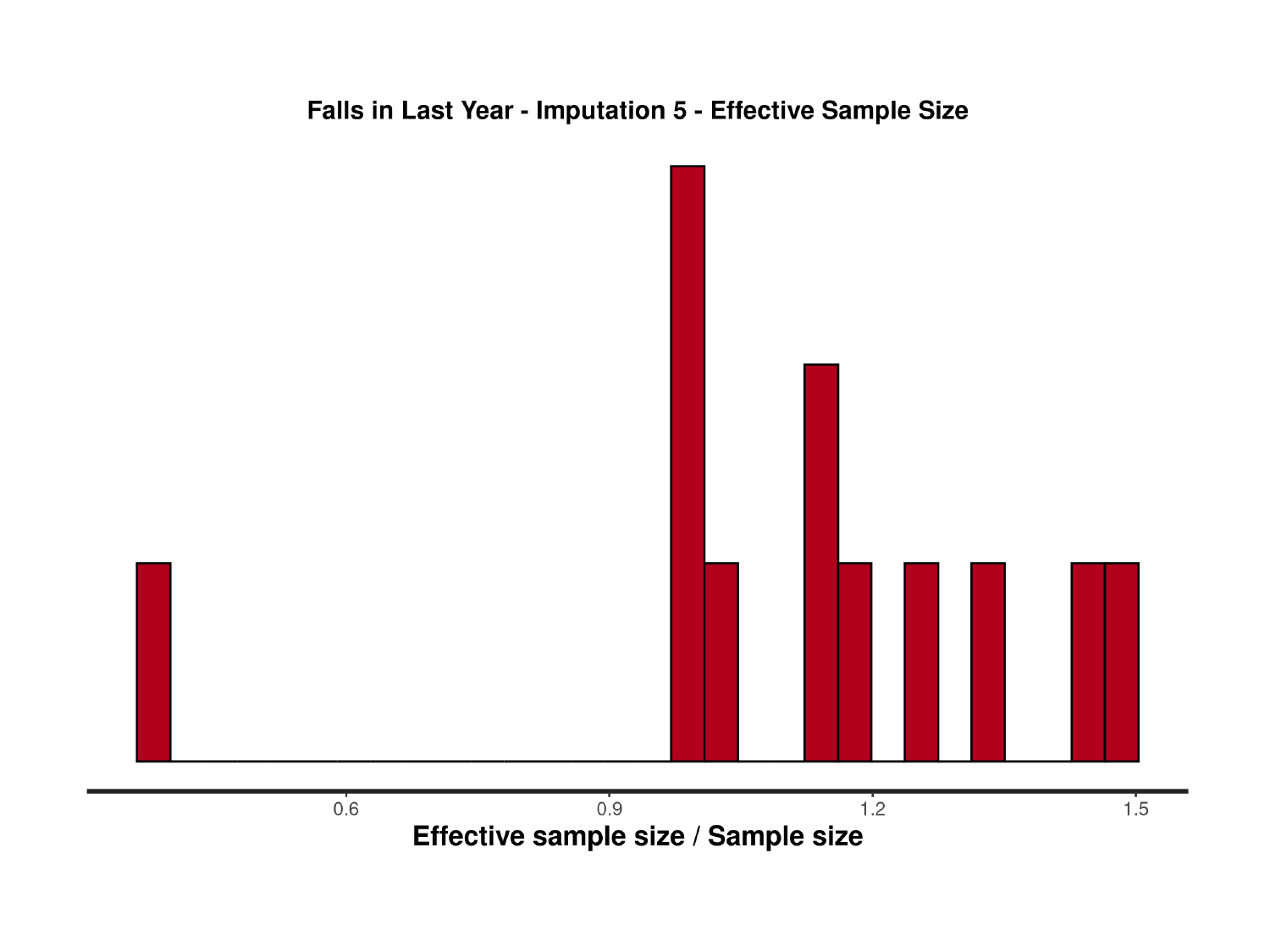

### Slide 112
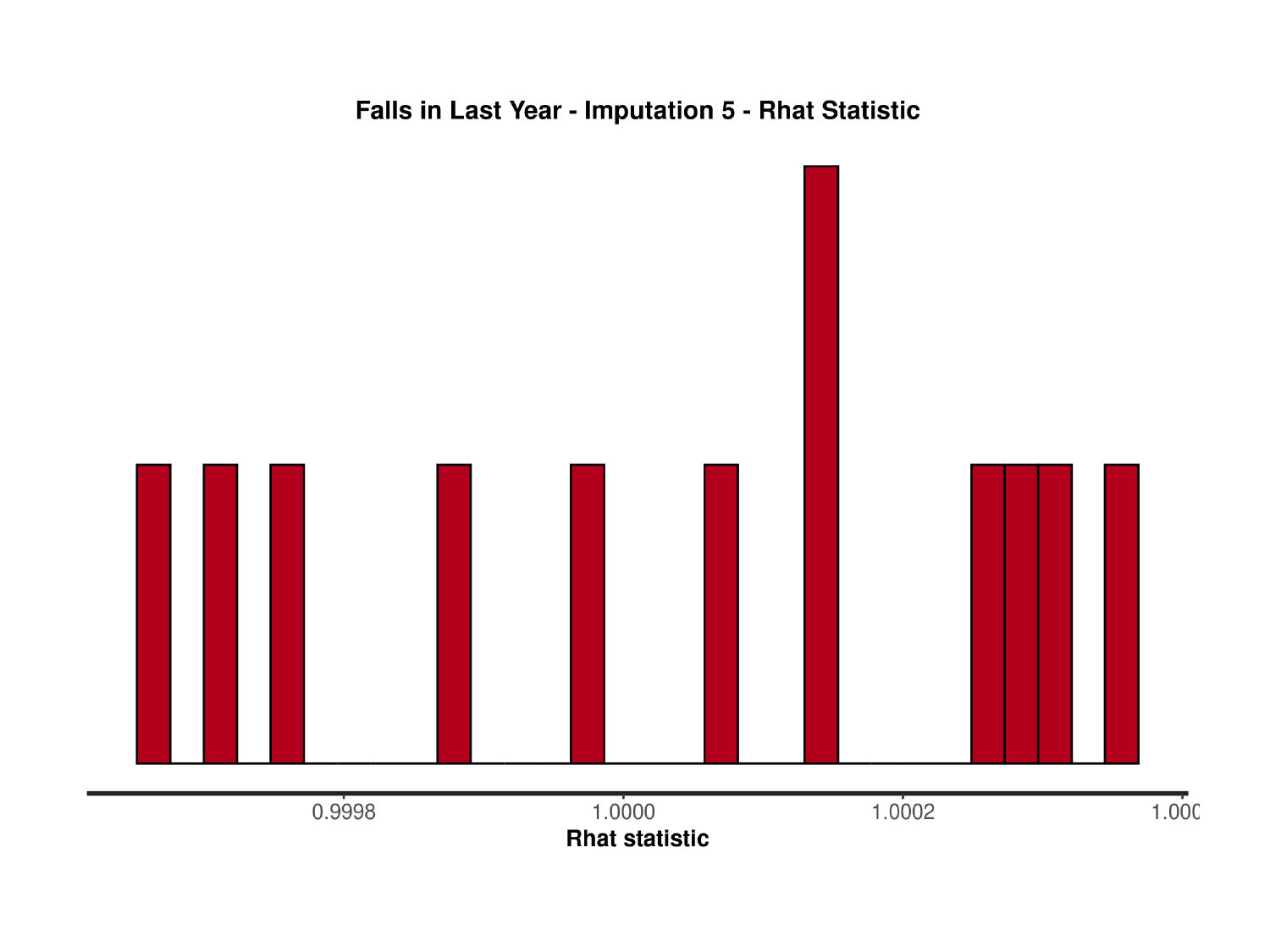

### Slide 113
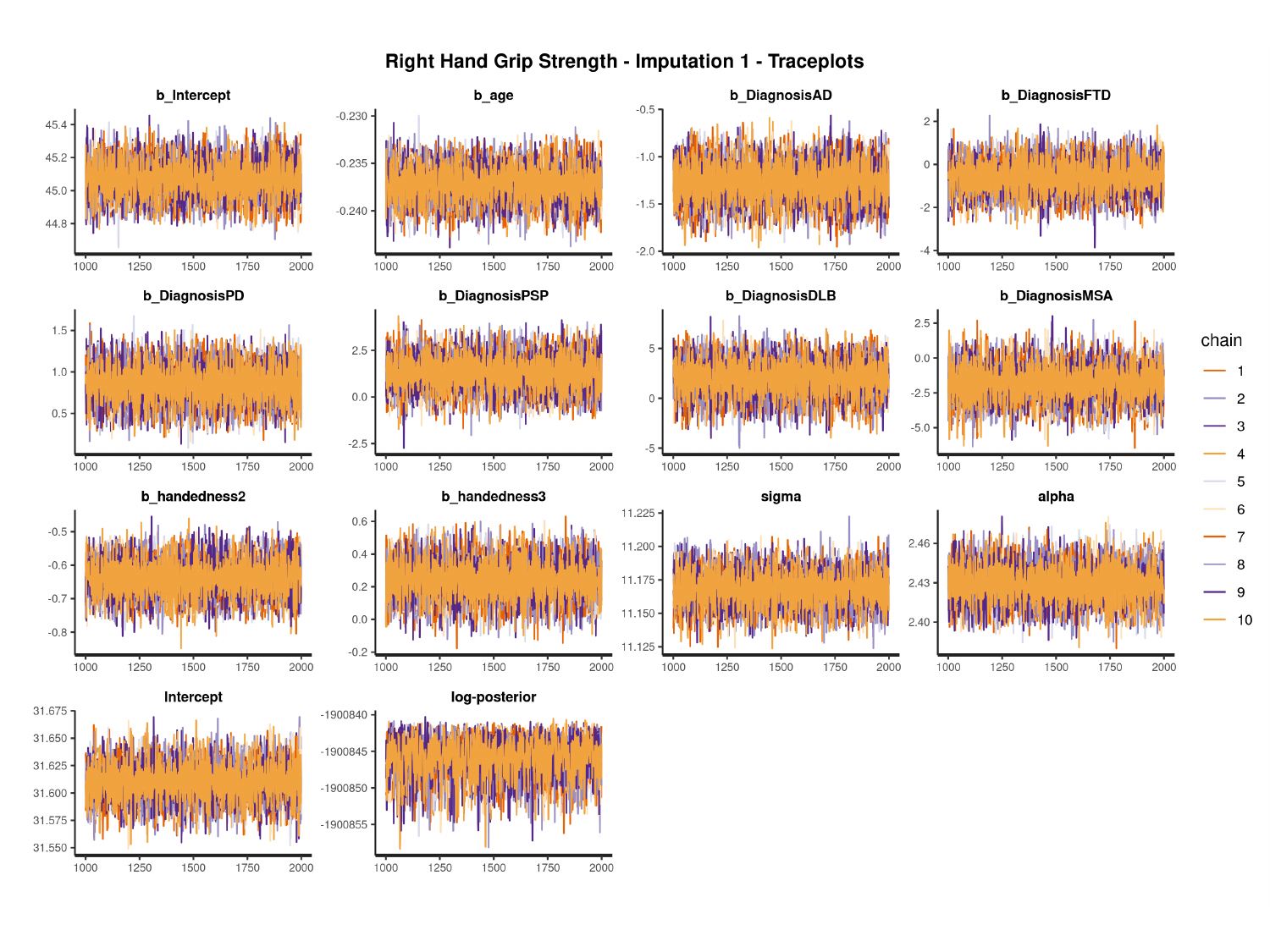

#

### Slide 114
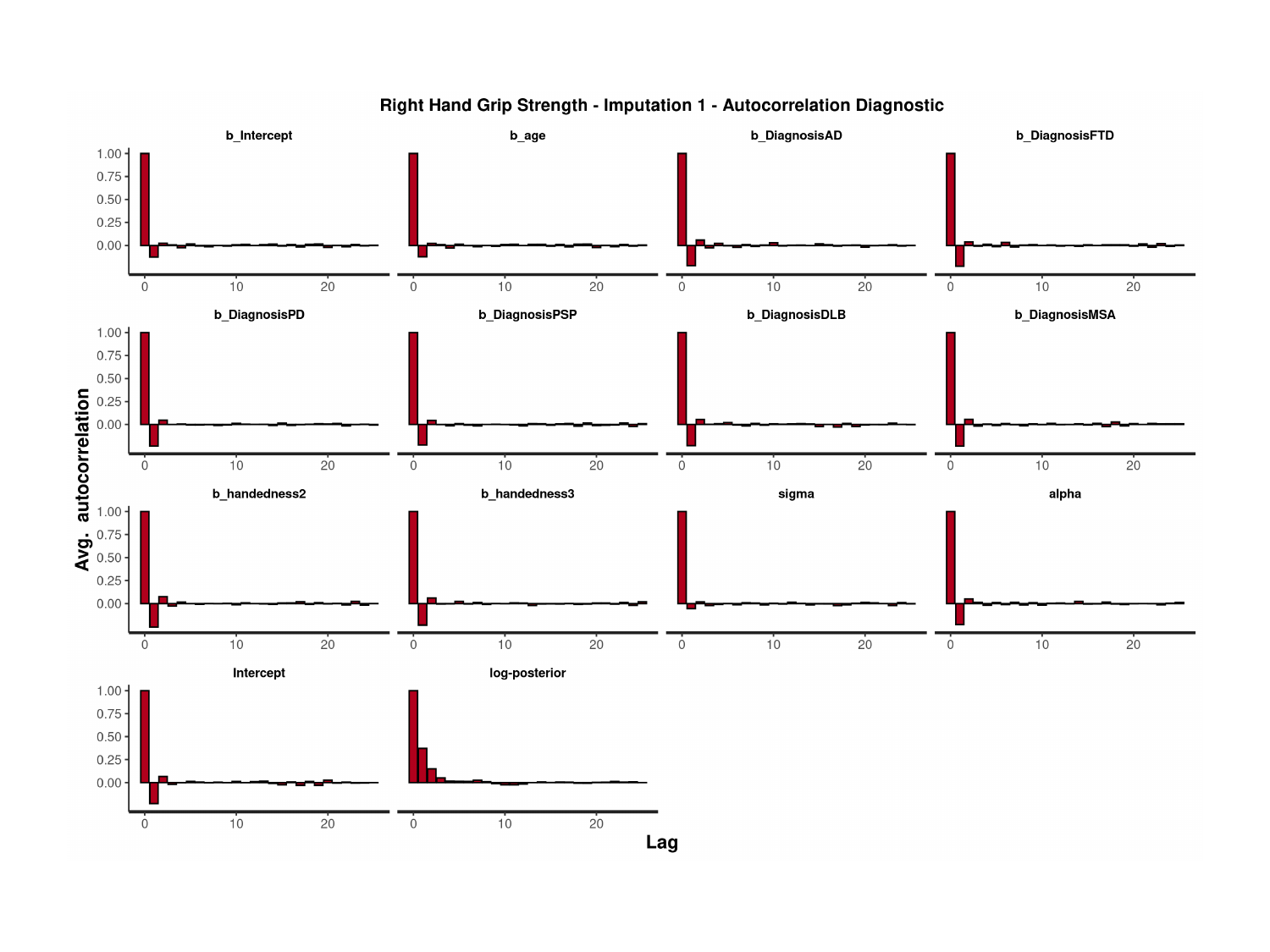

### Slide 115
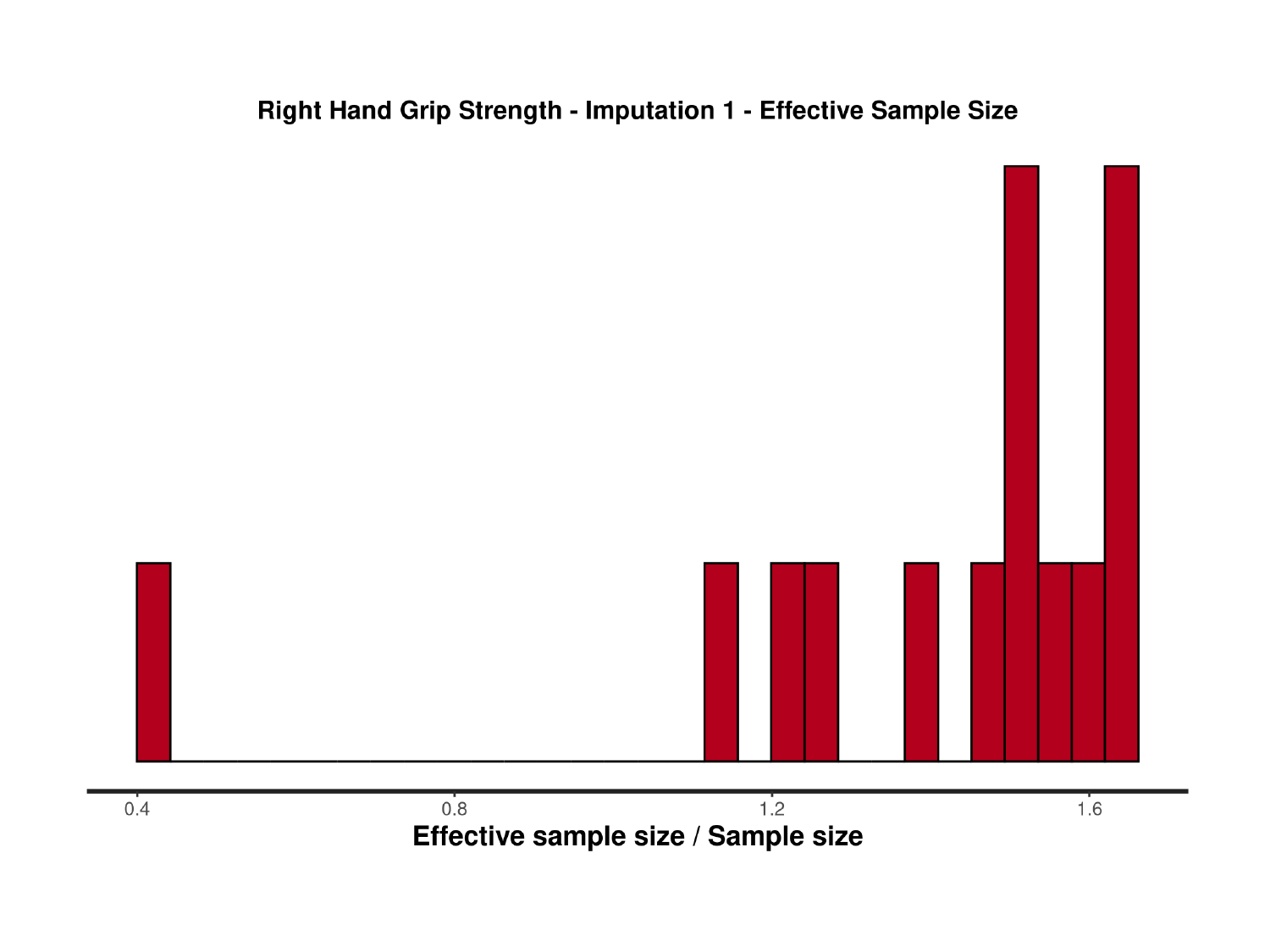

### Slide 116
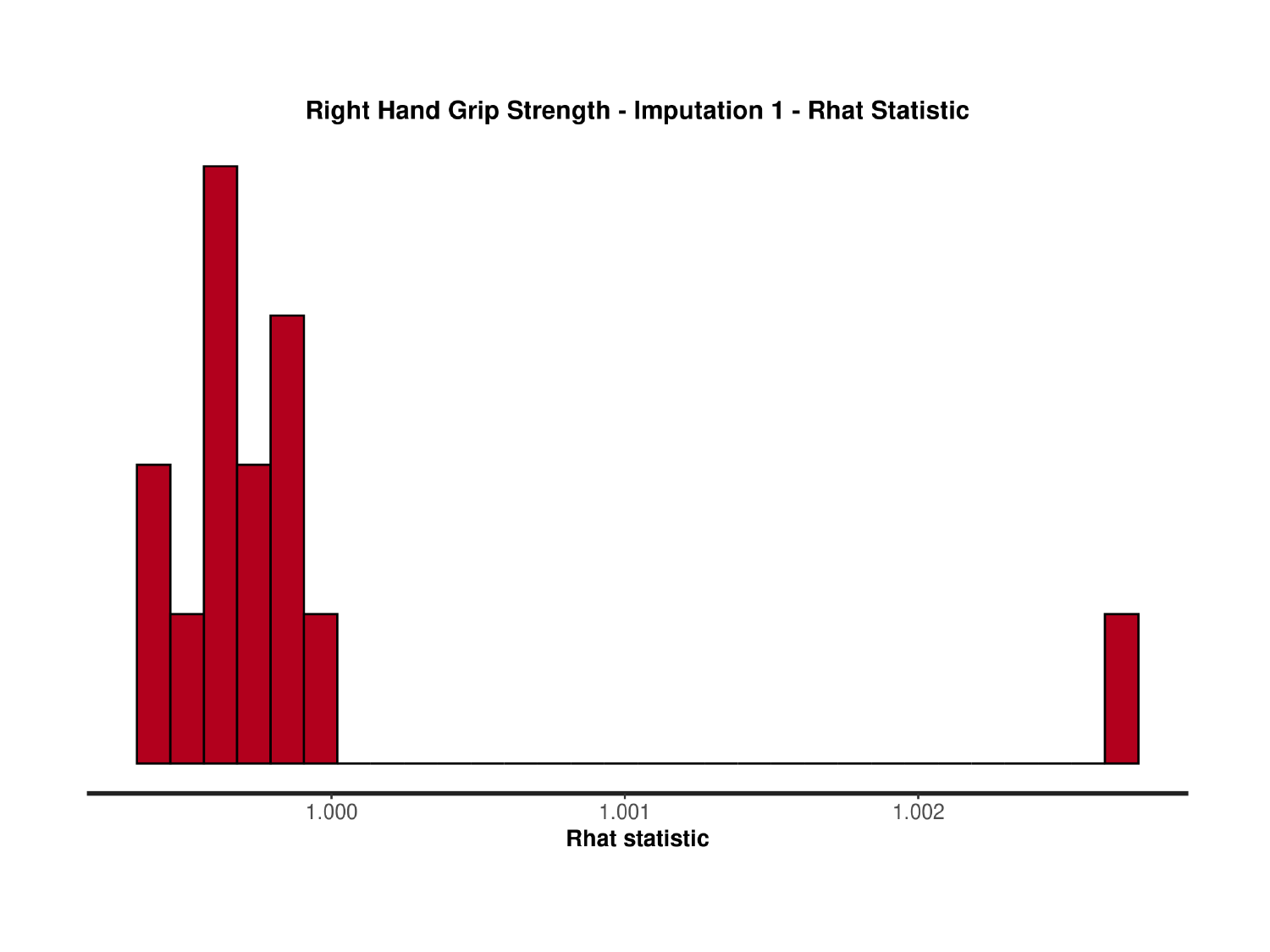

### Slide 117
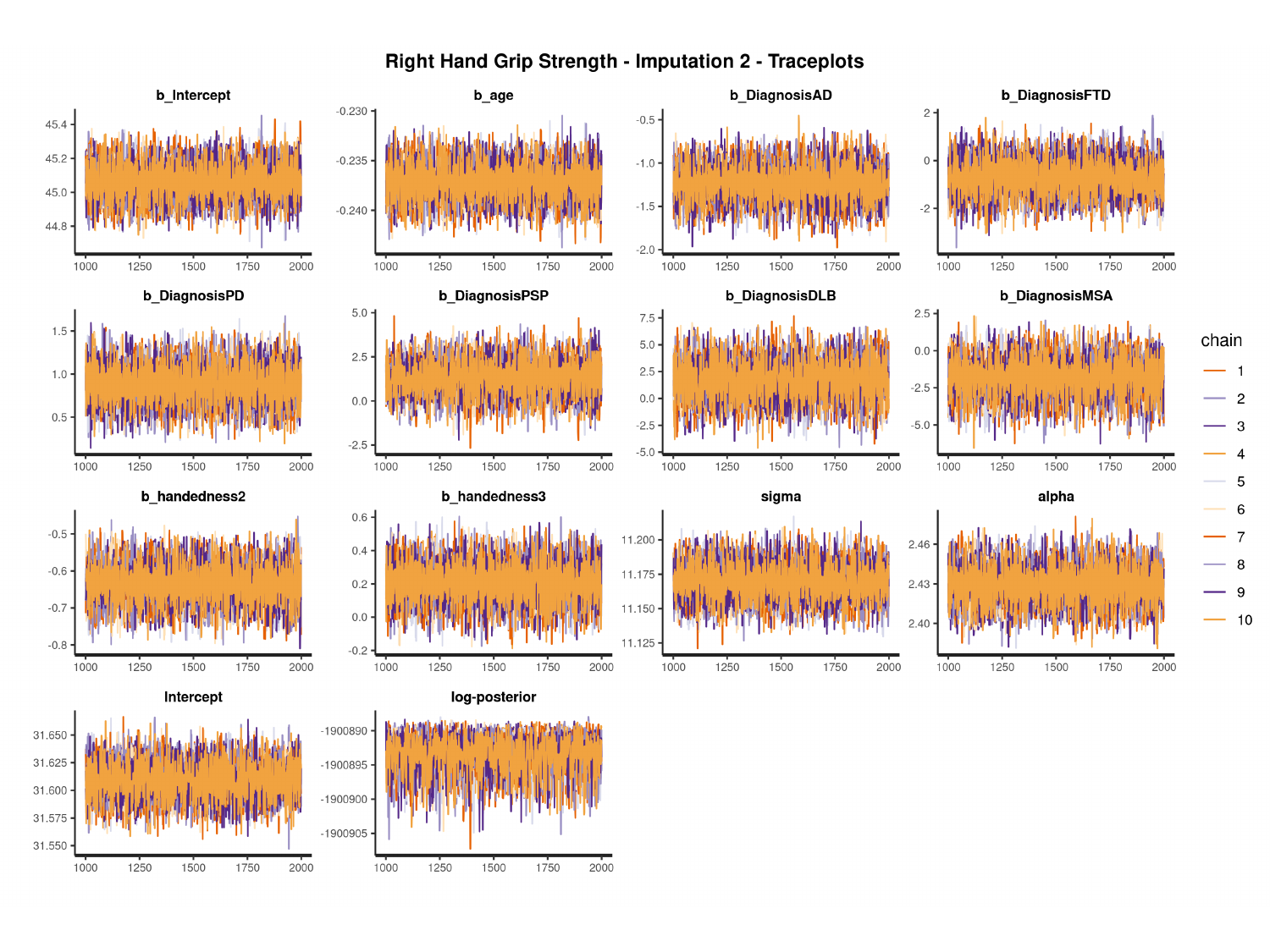

#

### Slide 118
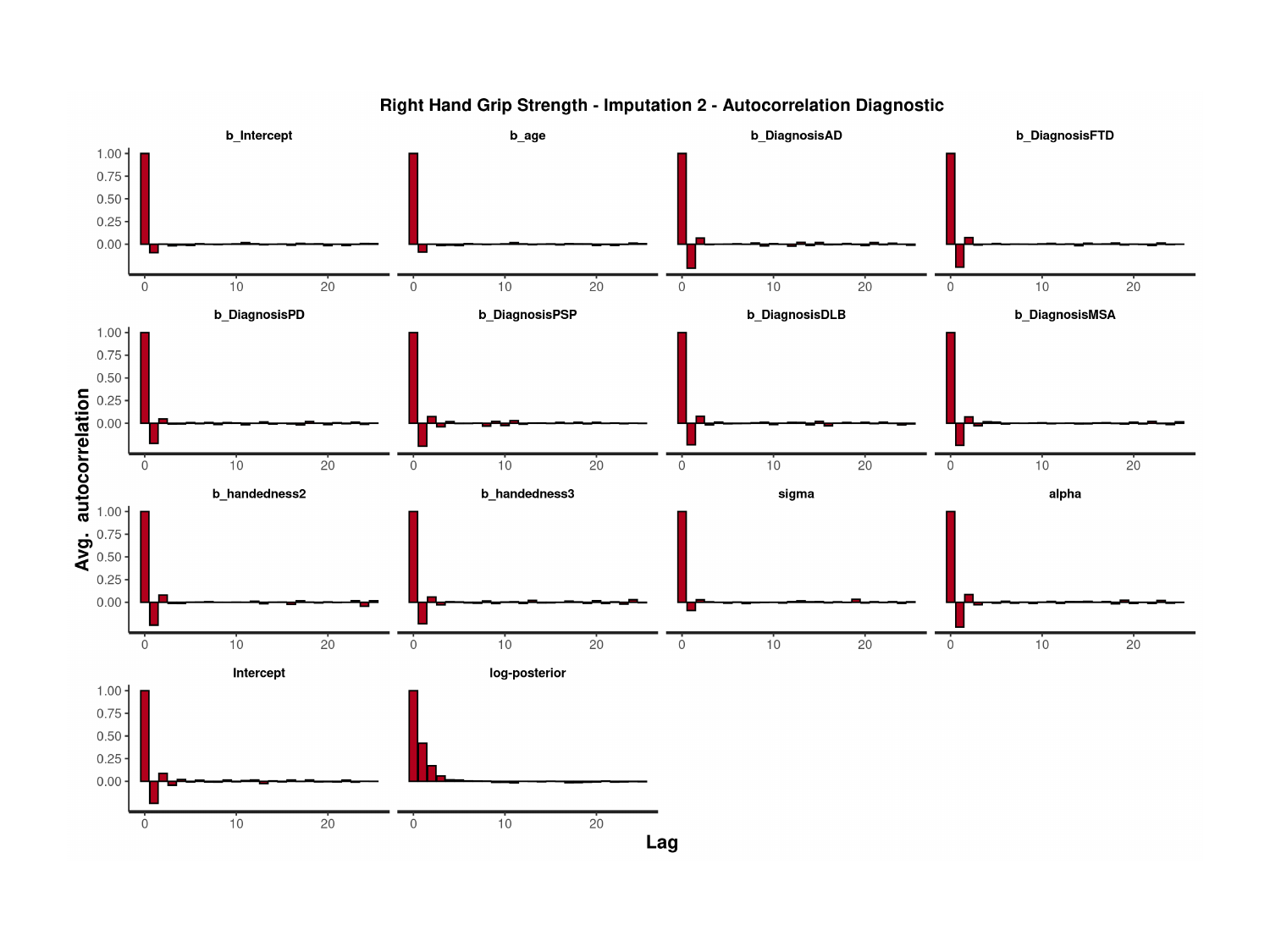

### Slide 119
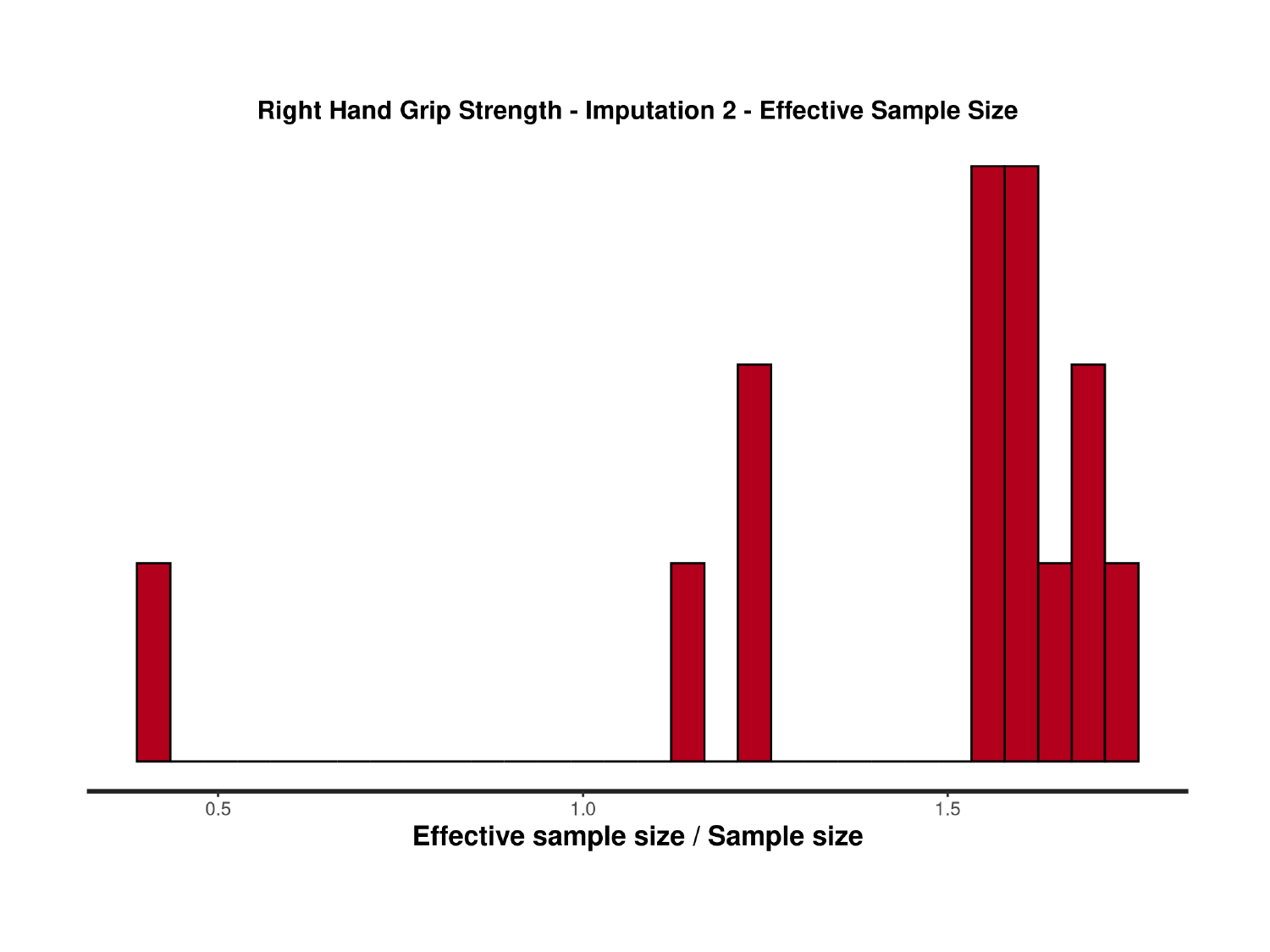

### Slide 120
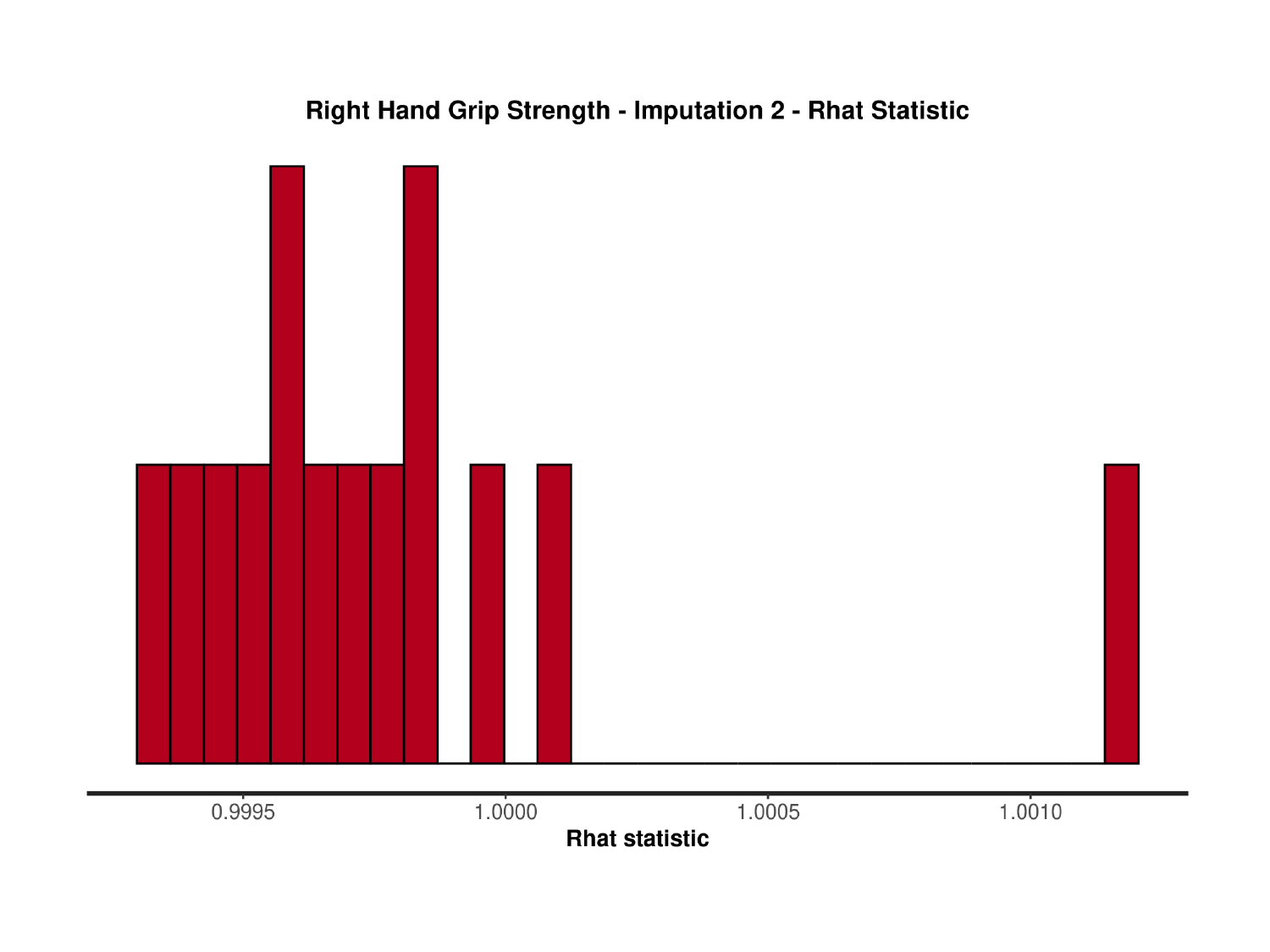

### Slide 121
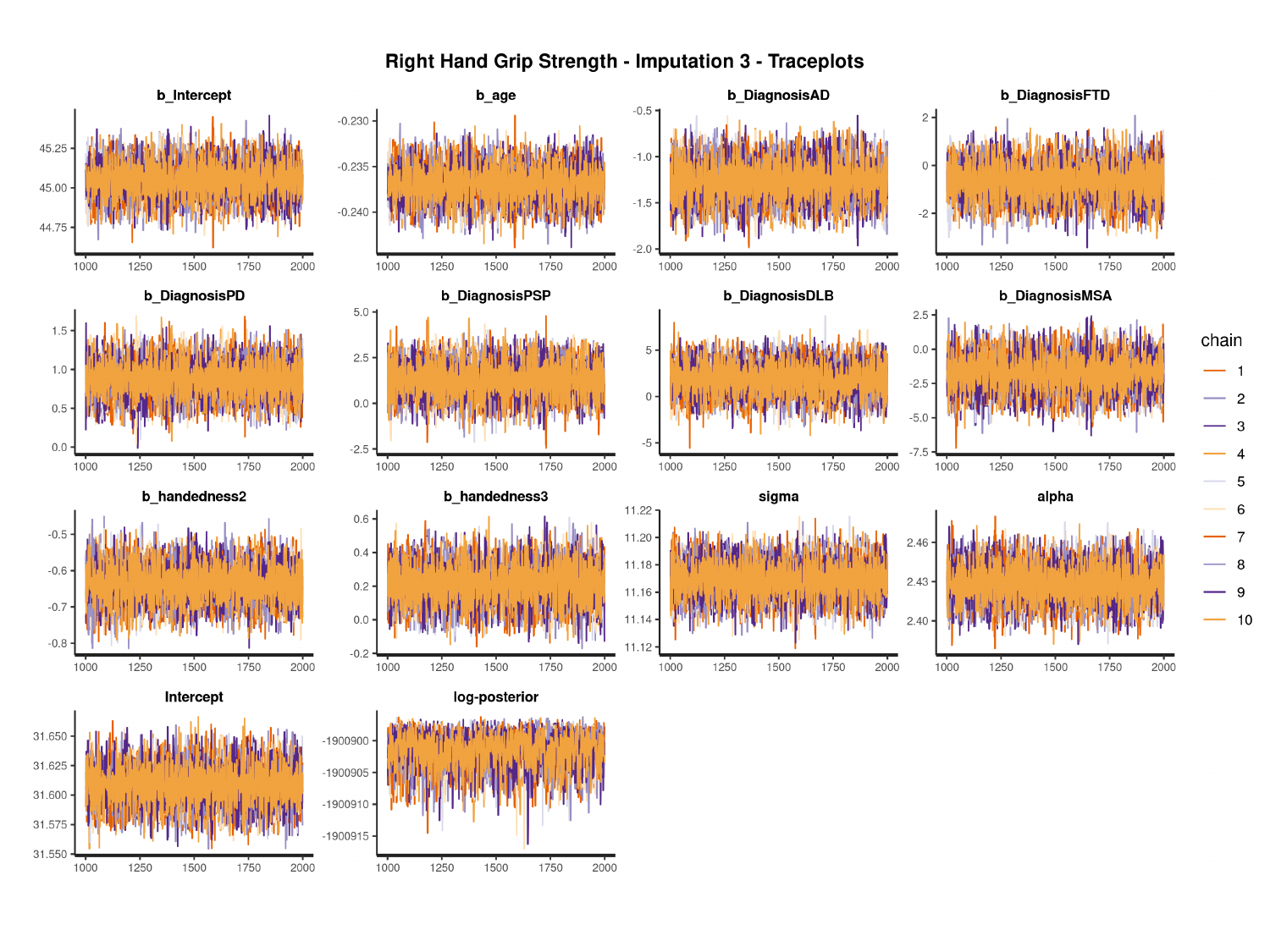

#

### Slide 122
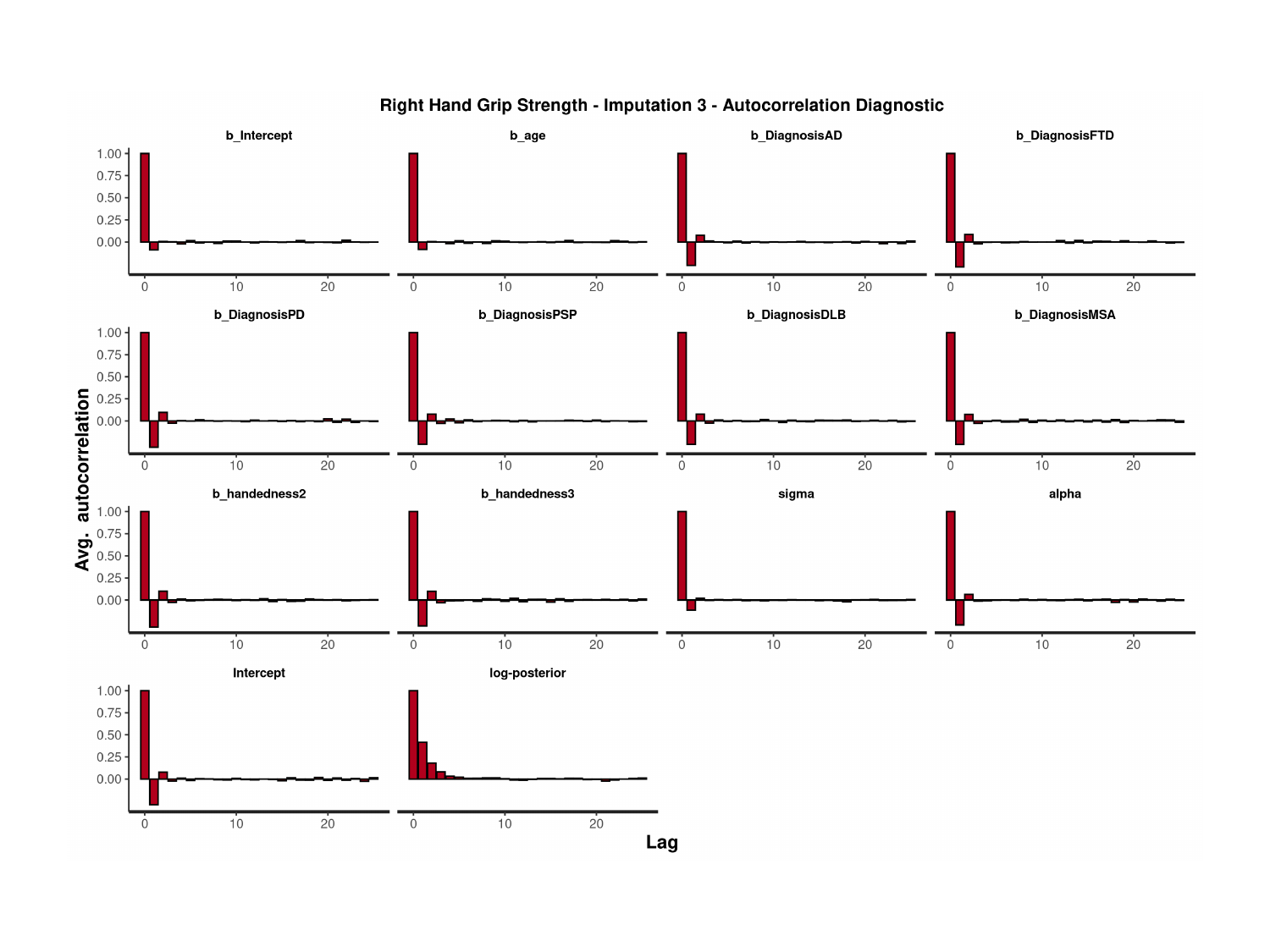

### Slide 123
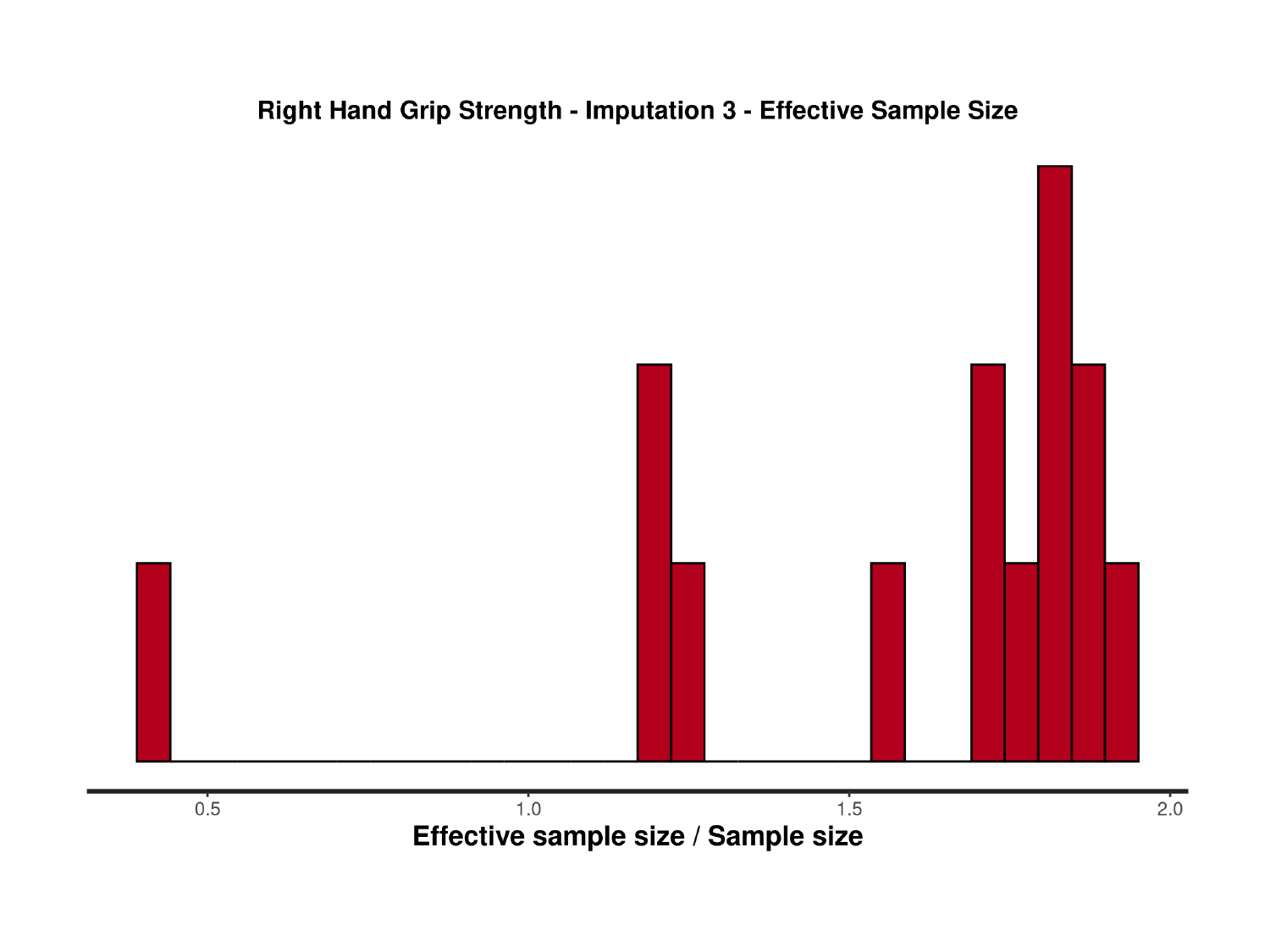

### Slide 124
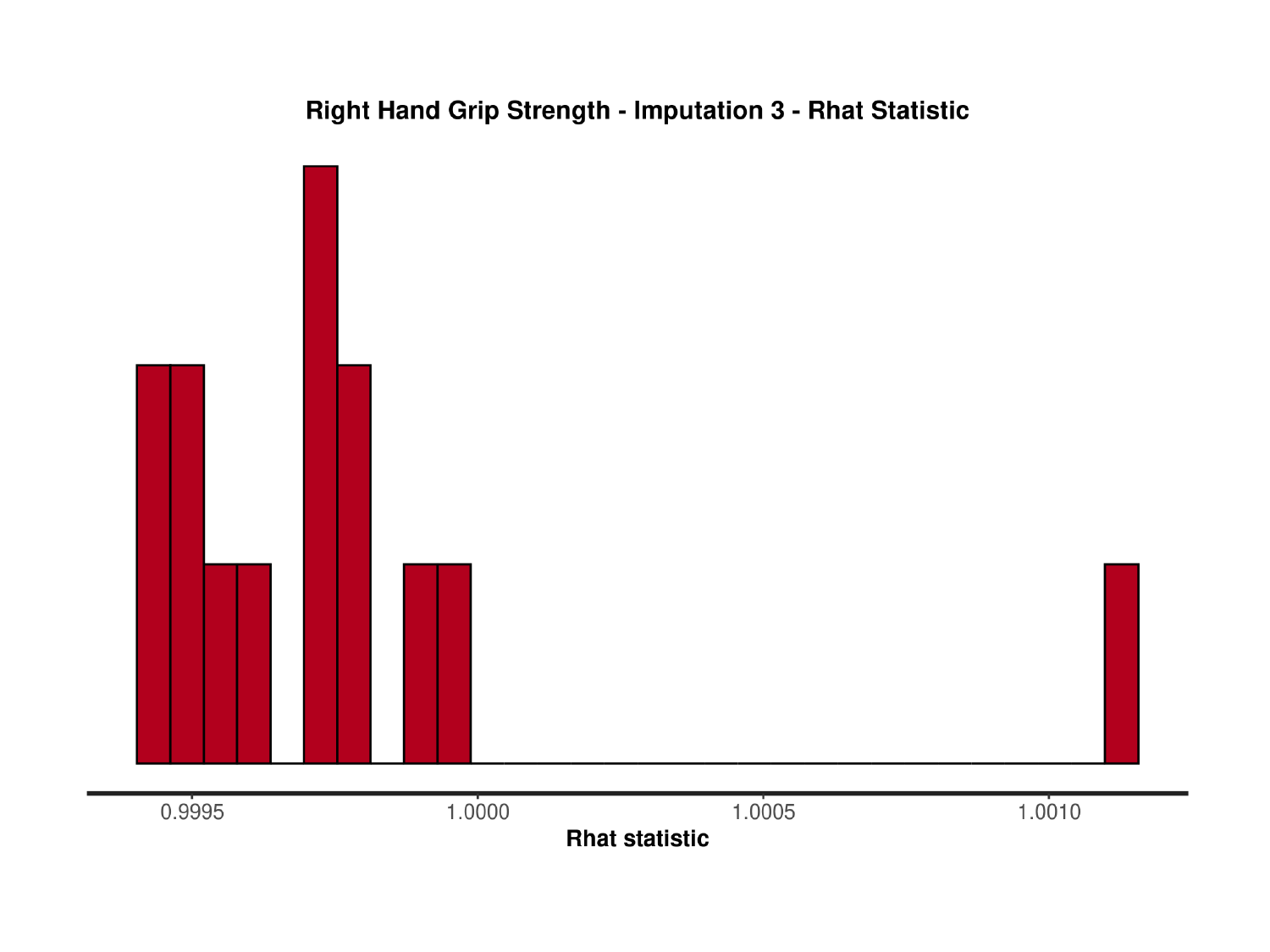

### Slide 125
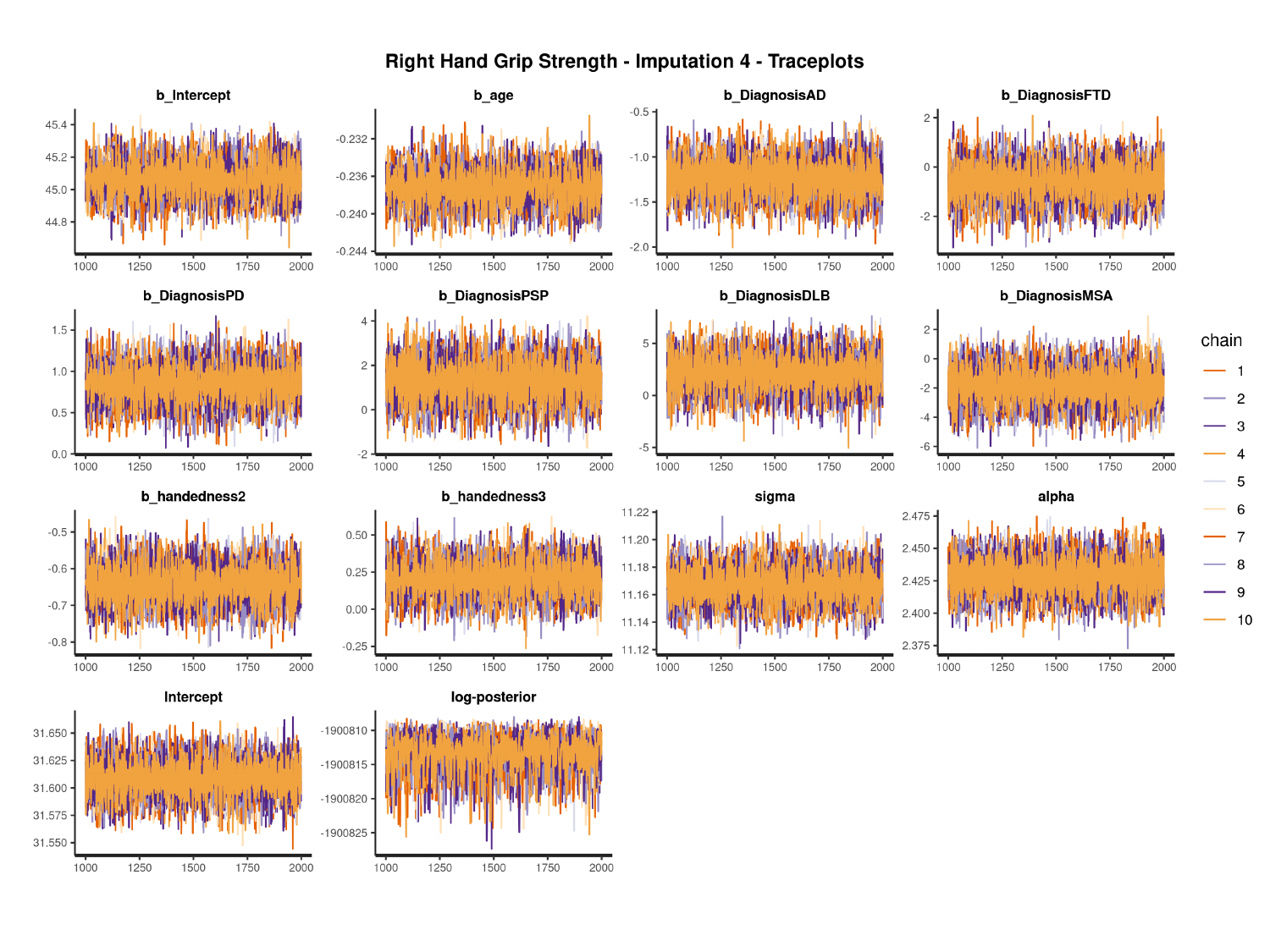

#

### Slide 126
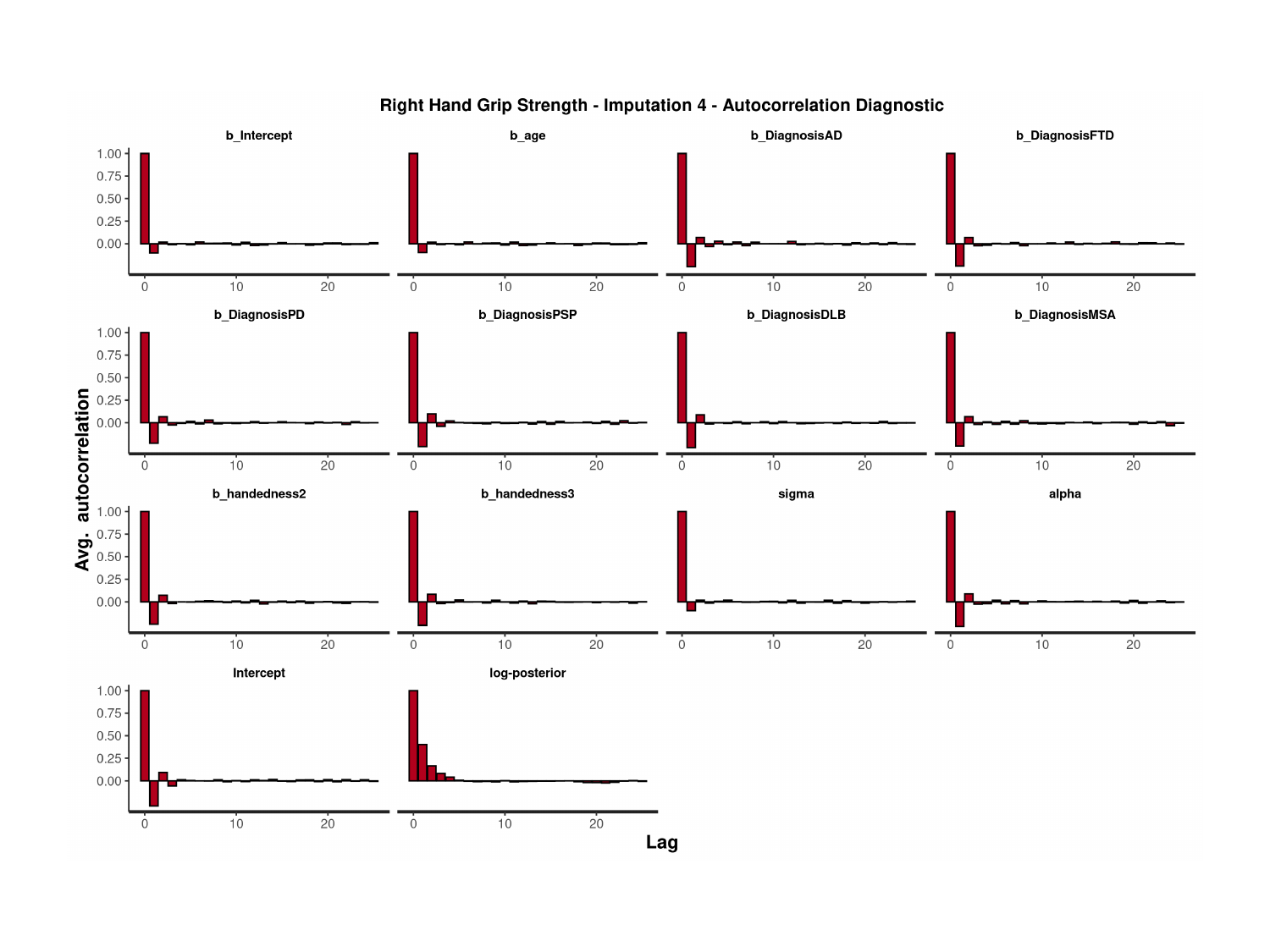

### Slide 127
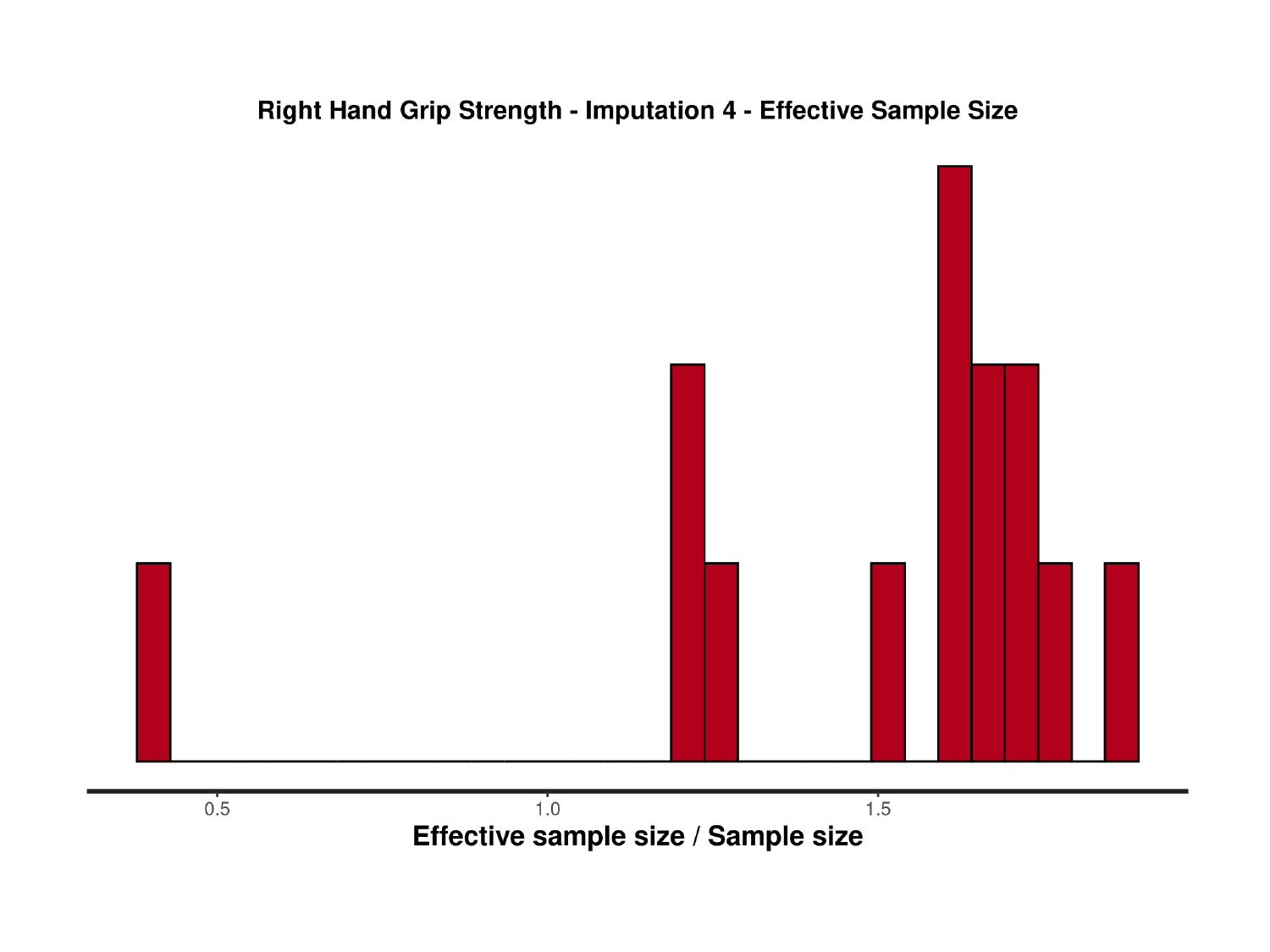

### Slide 128
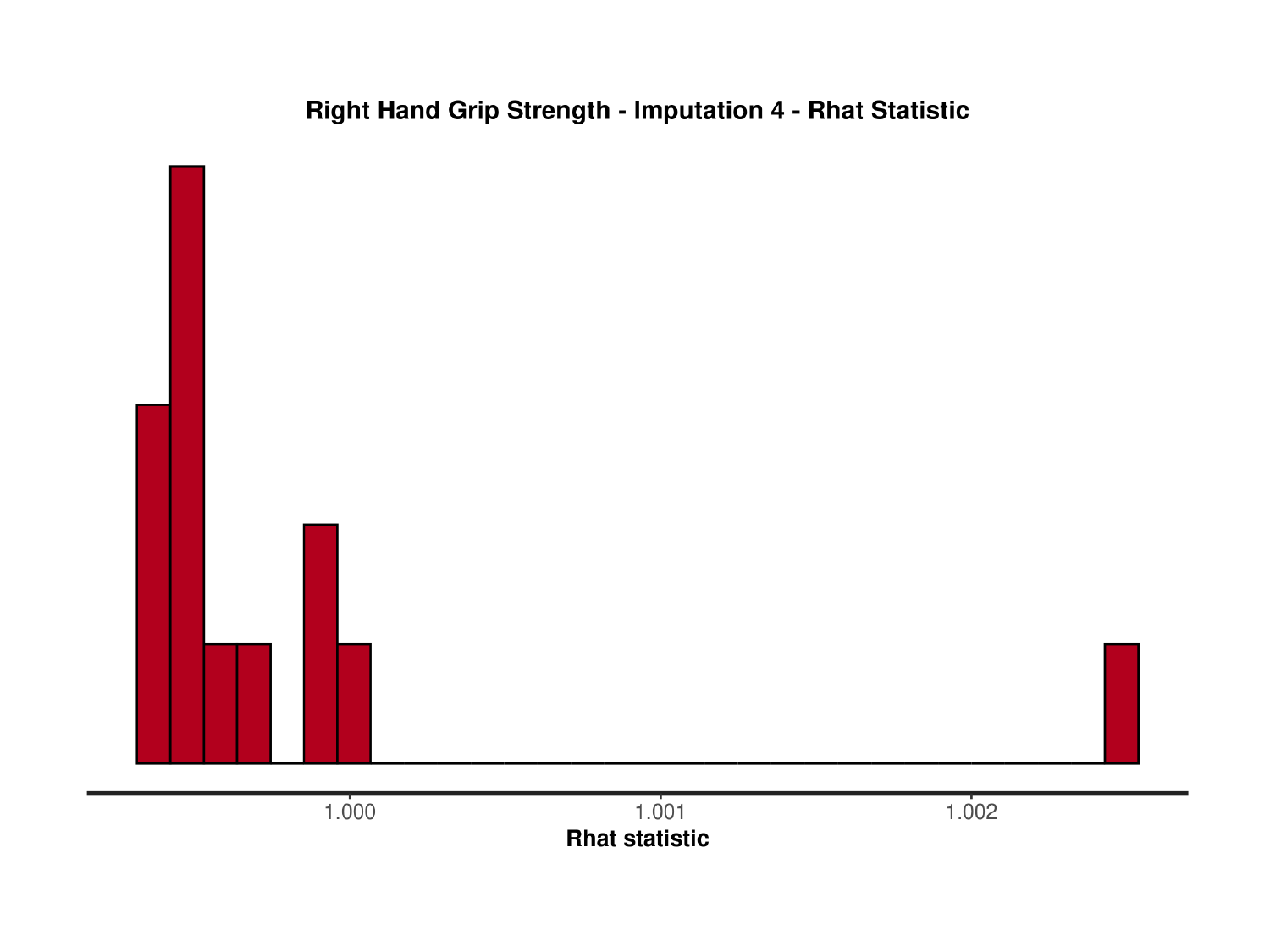

### Slide 129
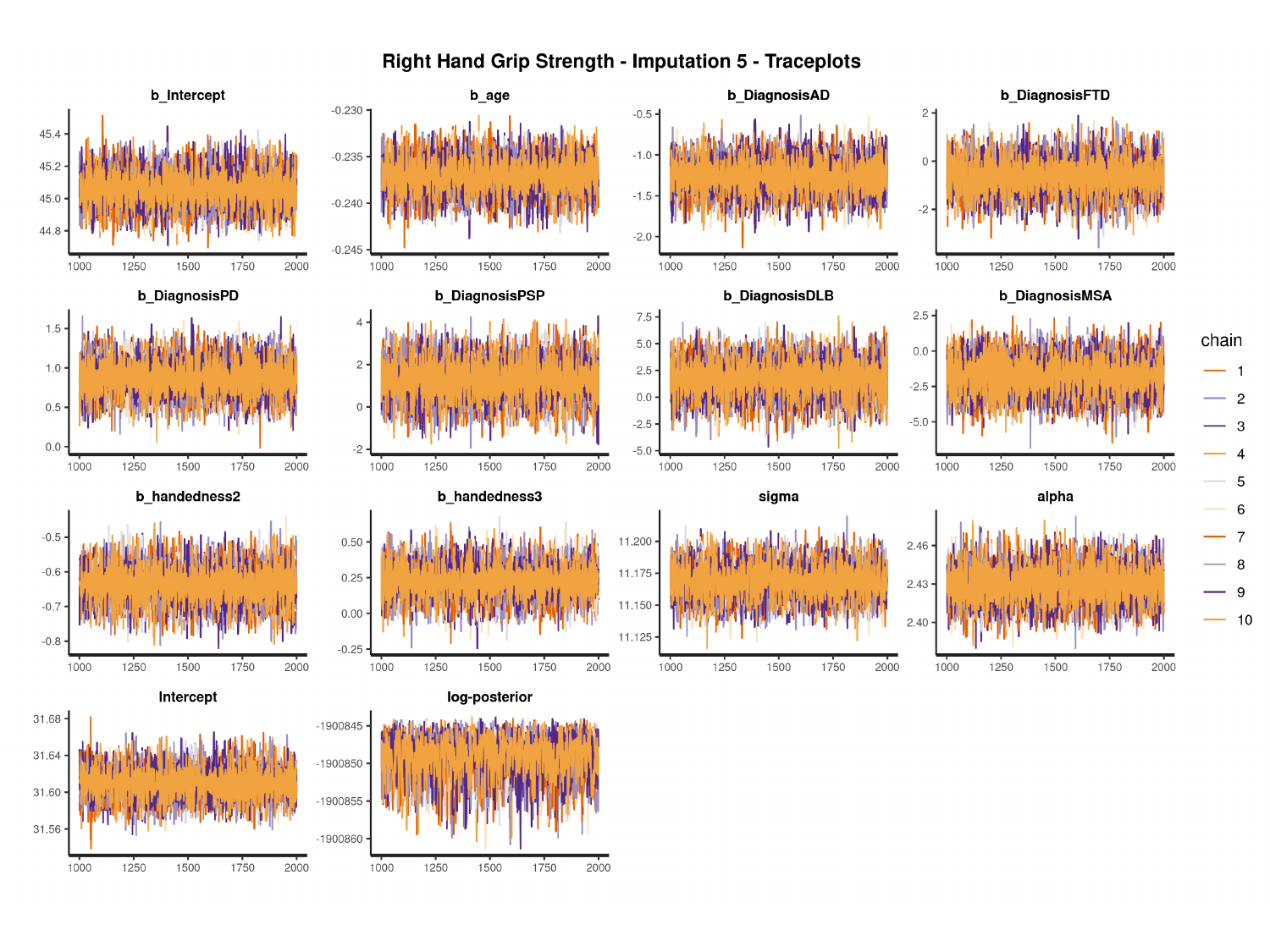

#

### Slide 130
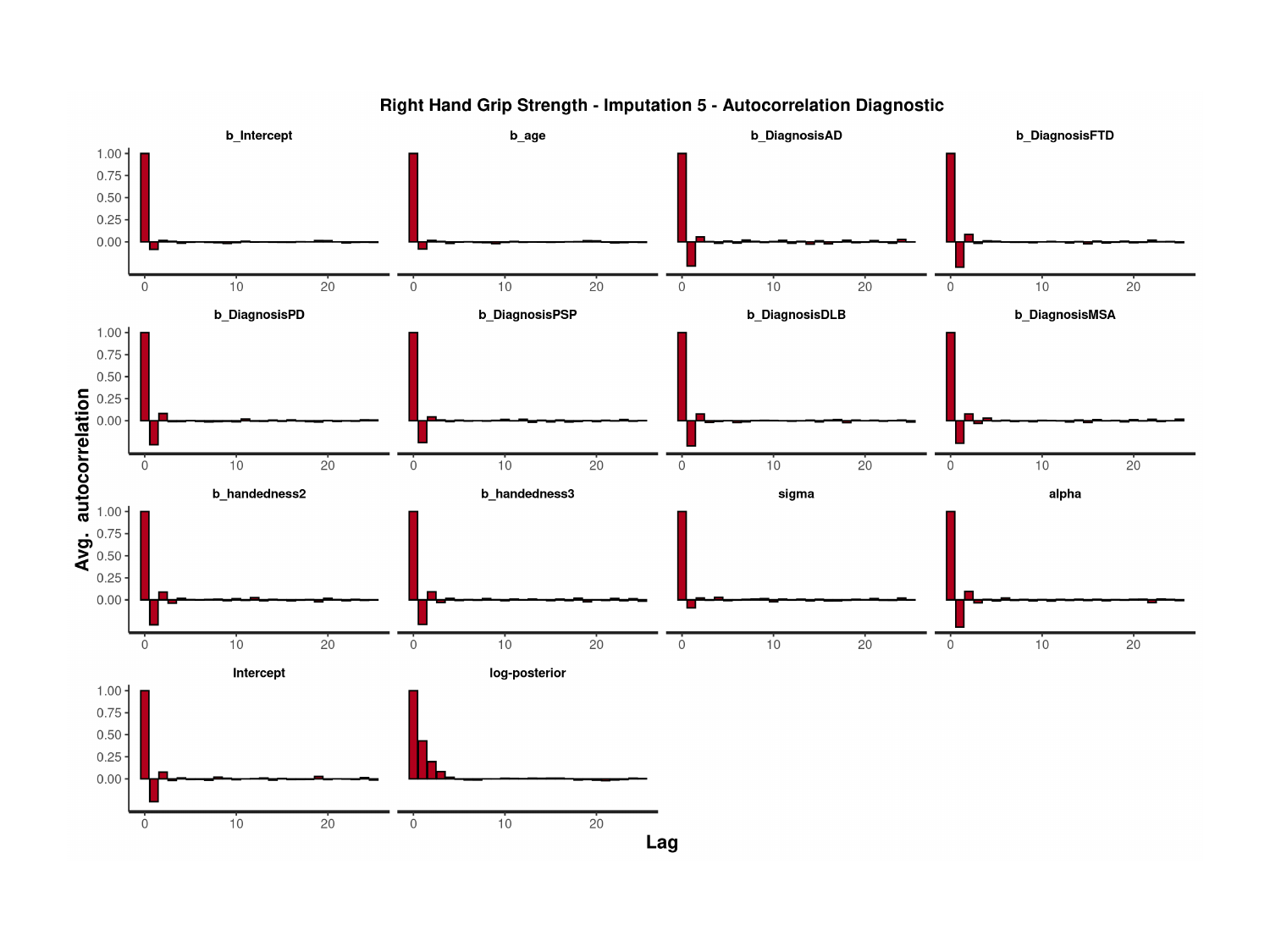

### Slide 131
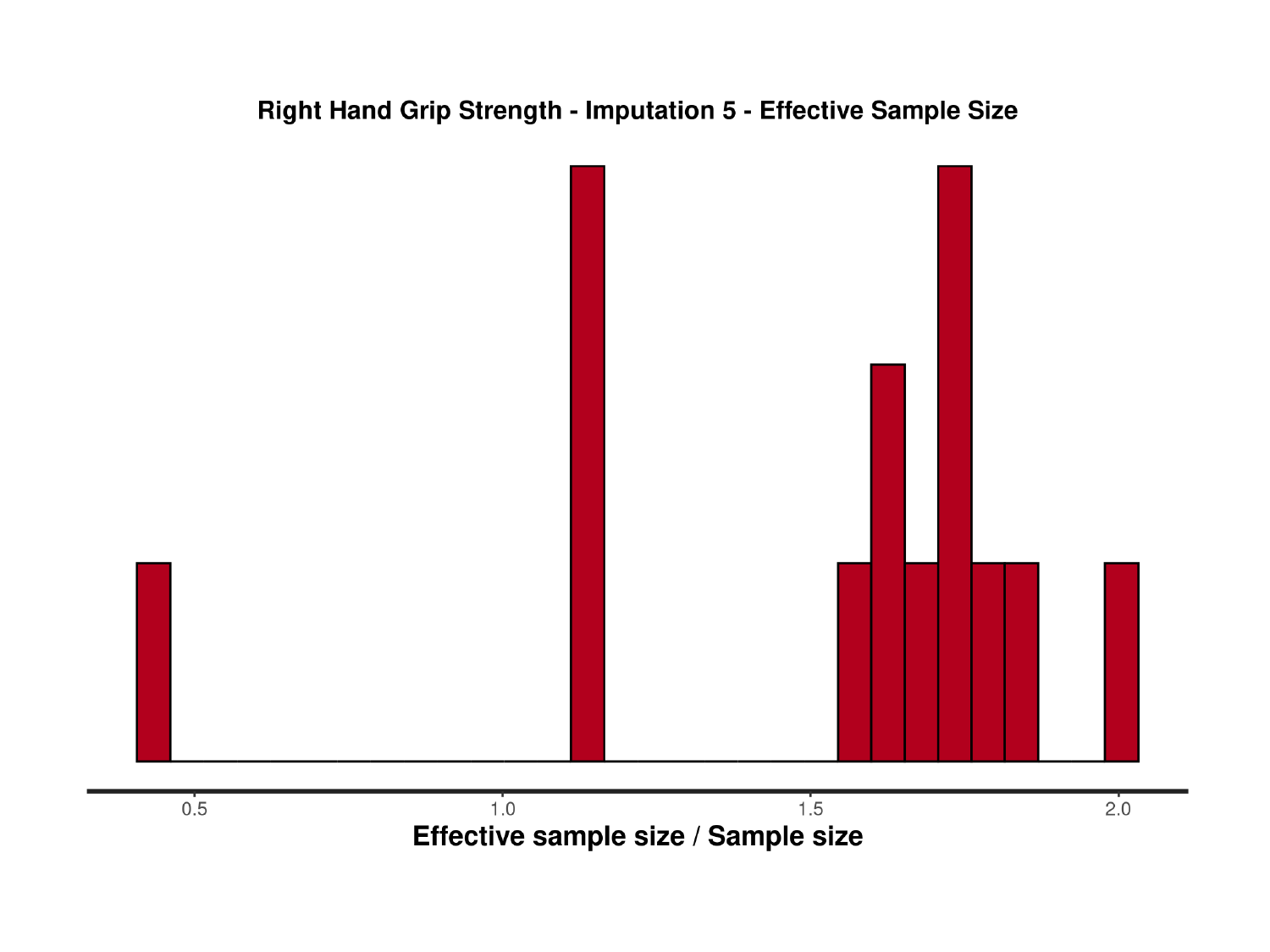

### Slide 132
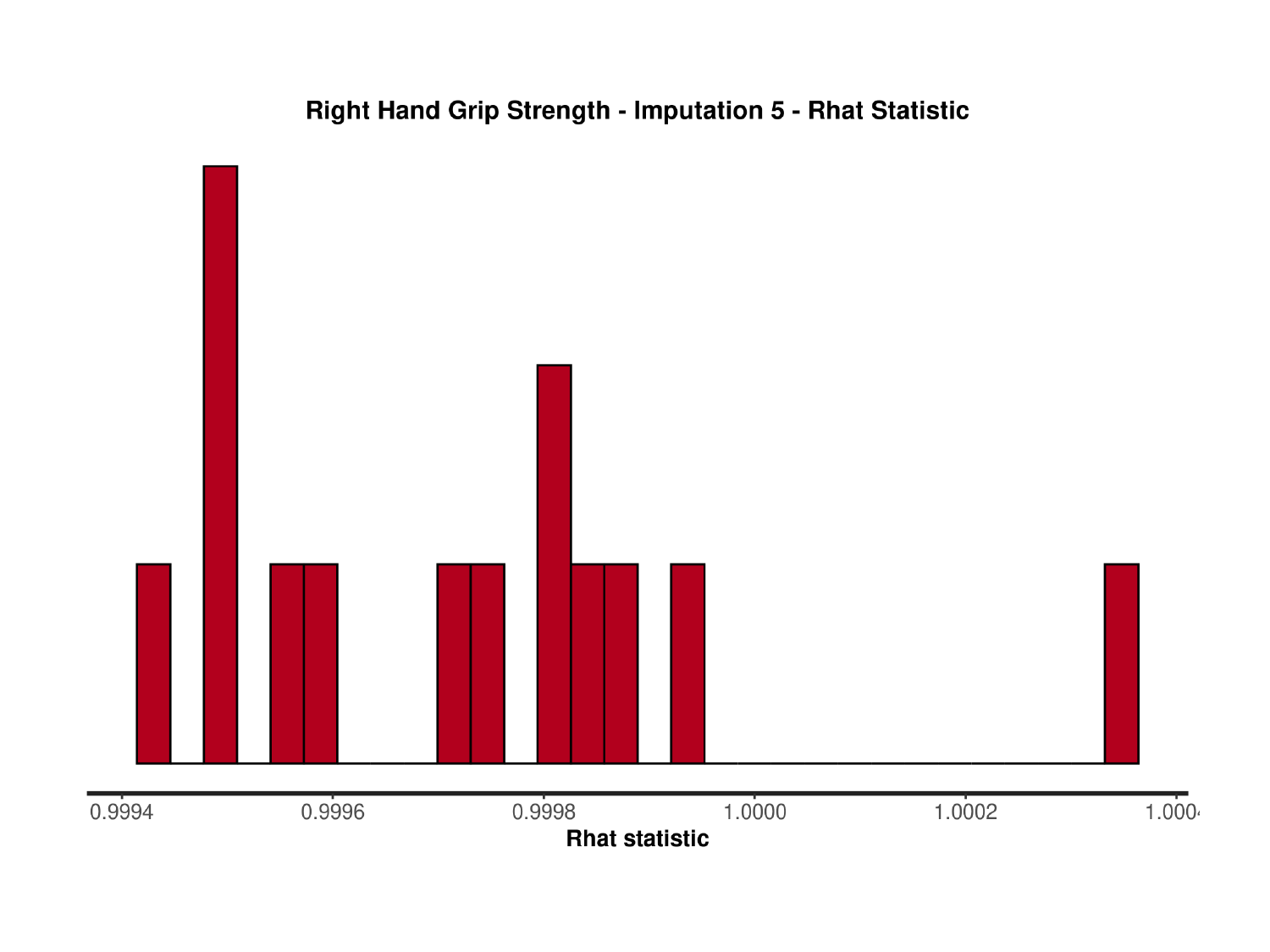

### Slide 133
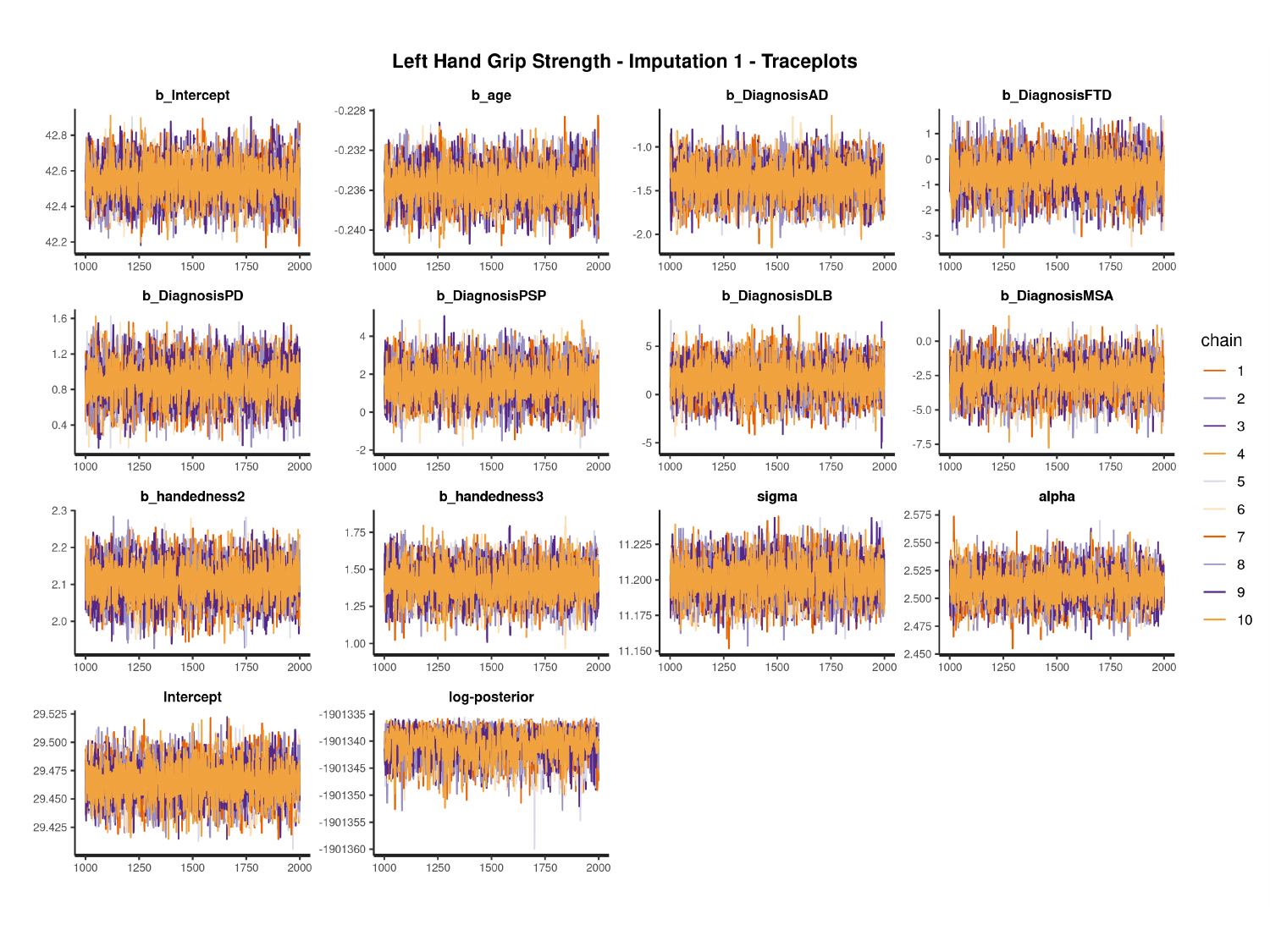

#

### Slide 134
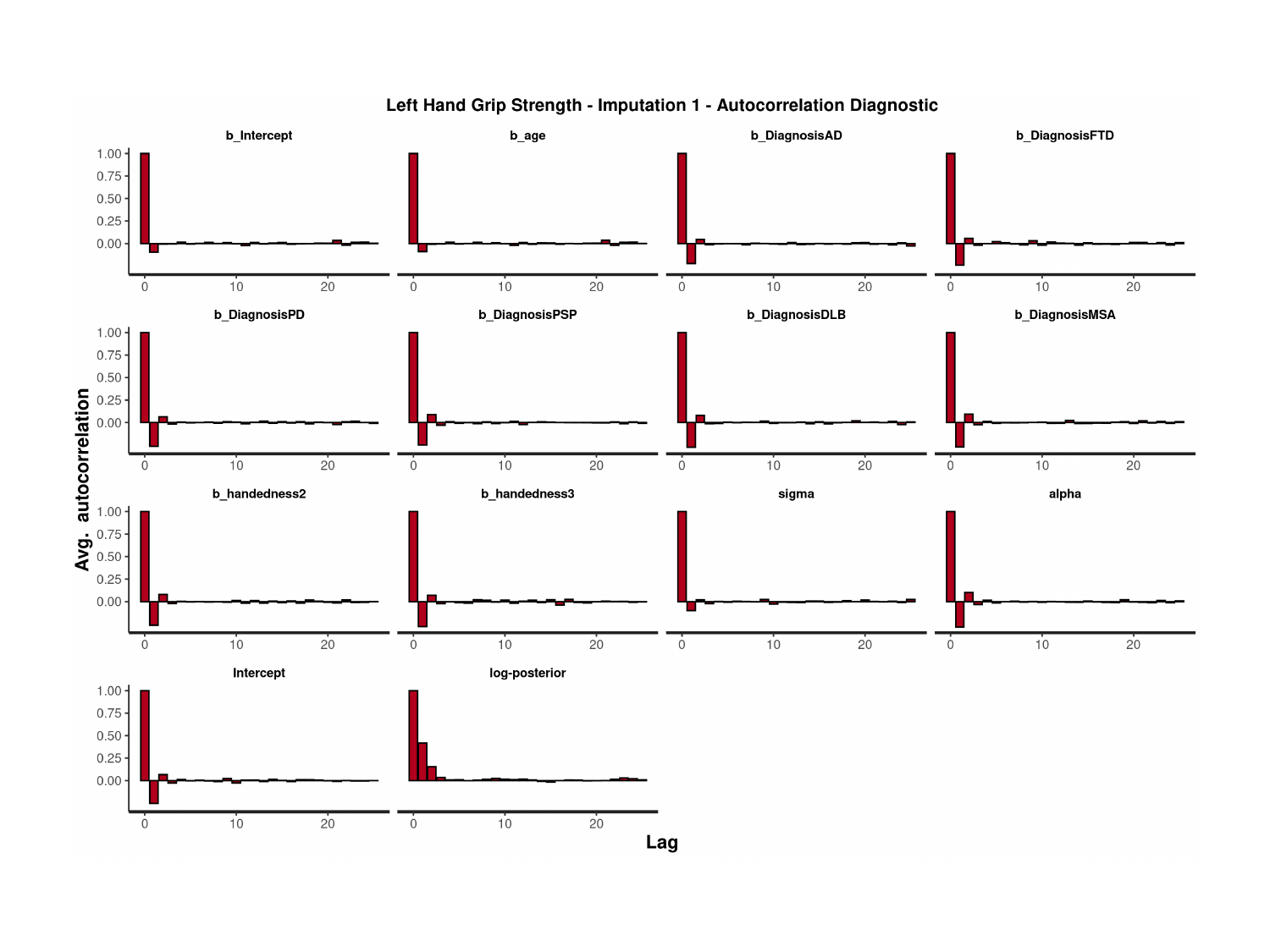

### Slide 135
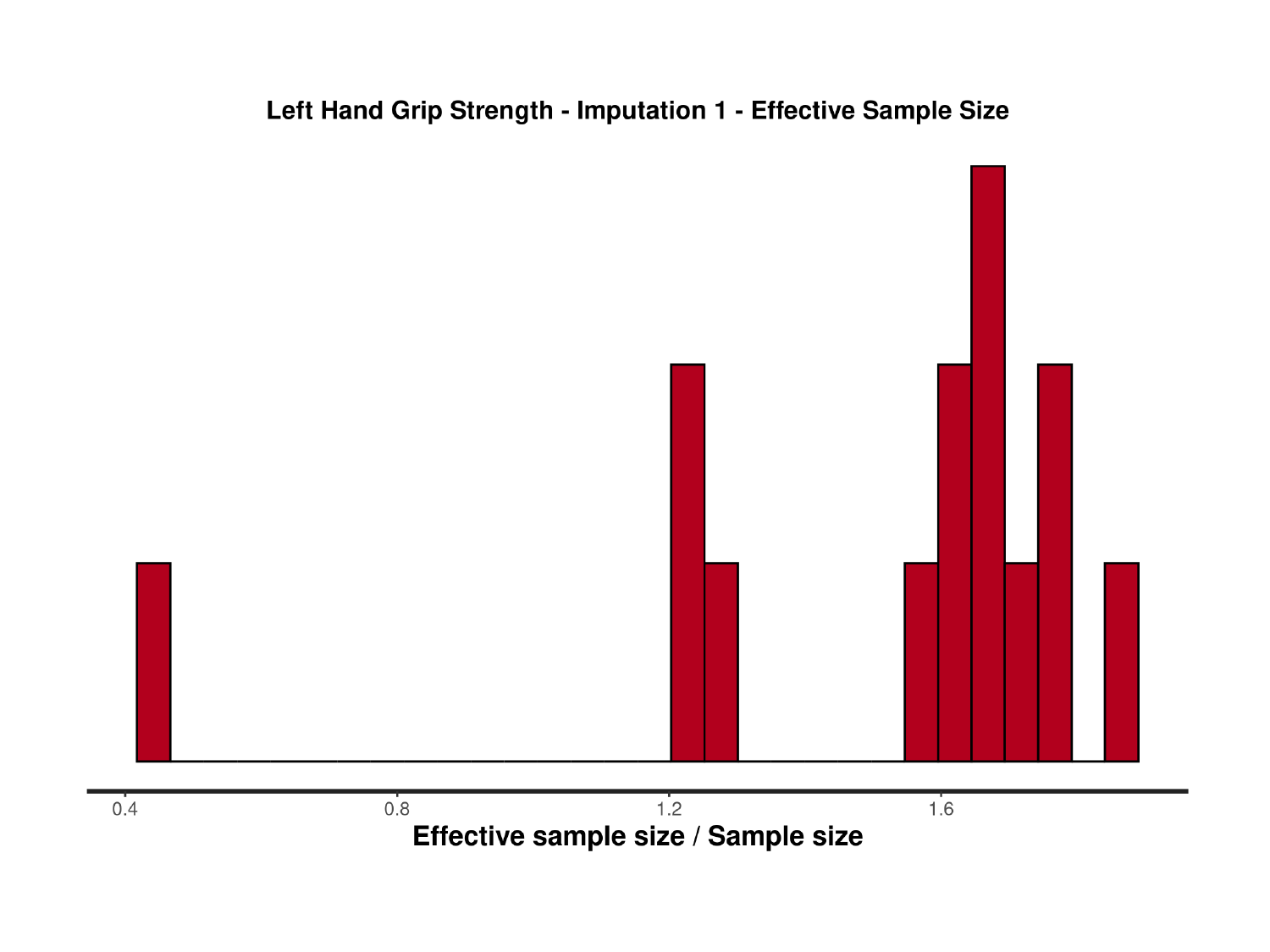

### Slide 136
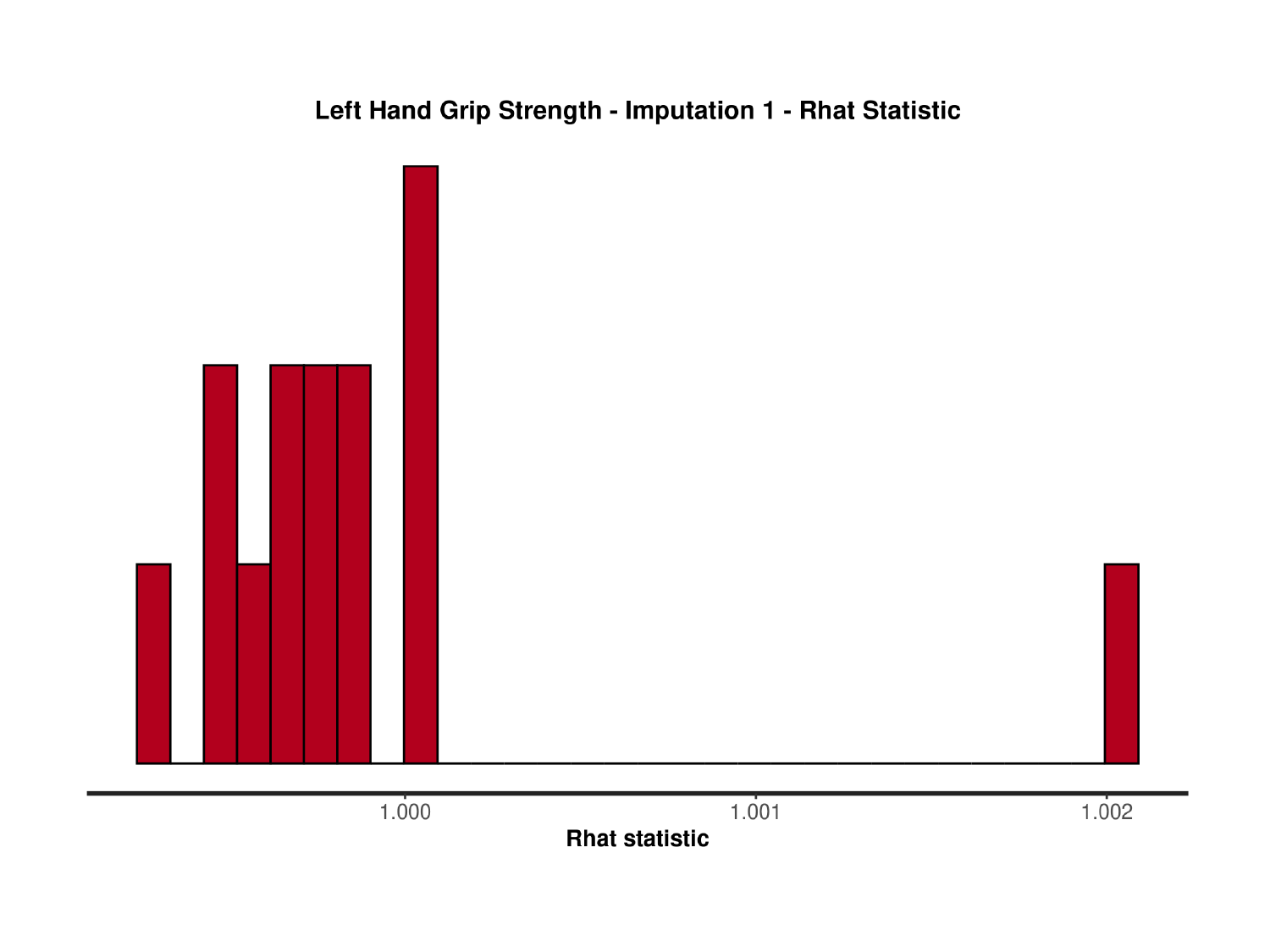

### Slide 137
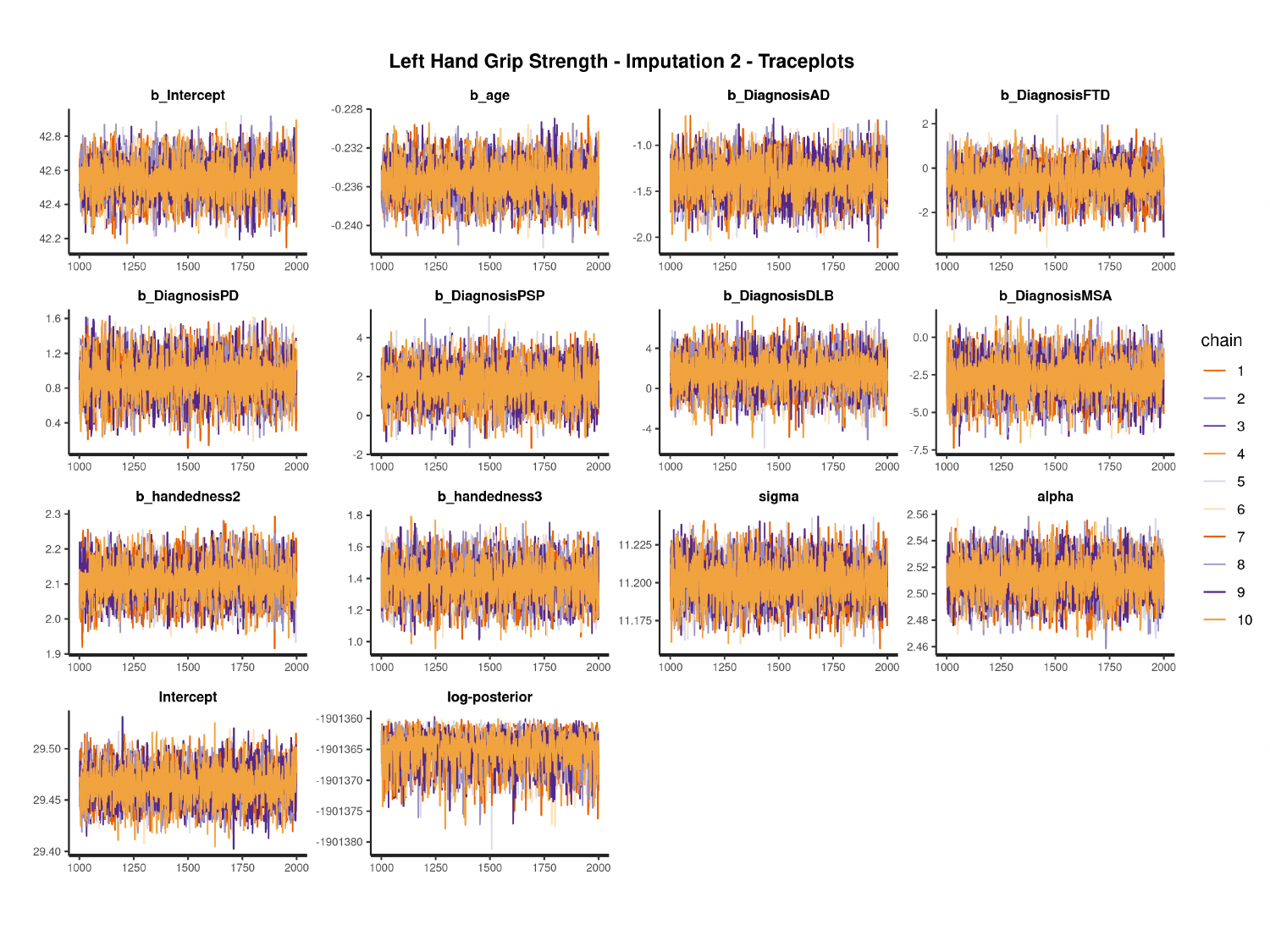

#

### Slide 138
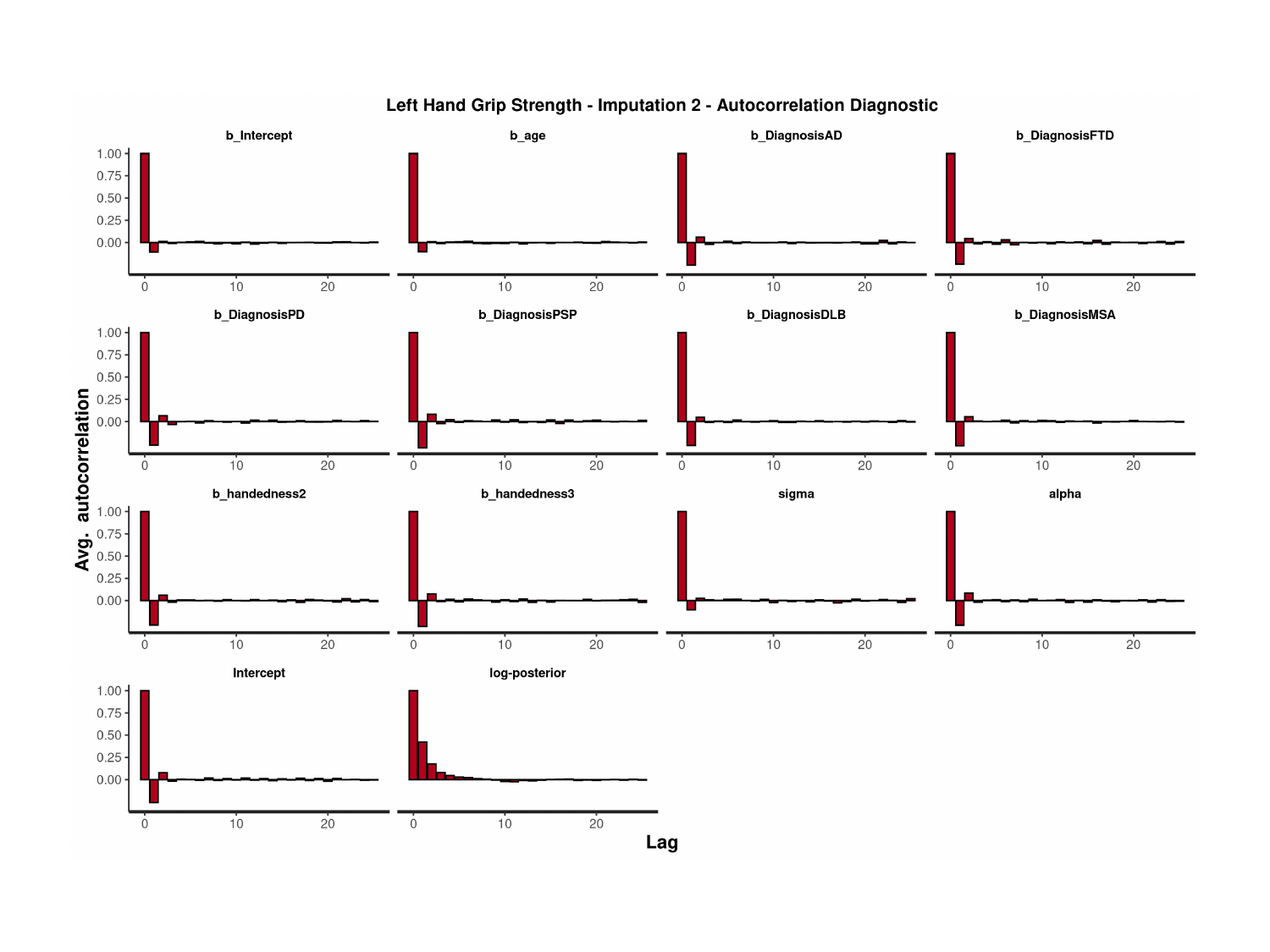

### Slide 139
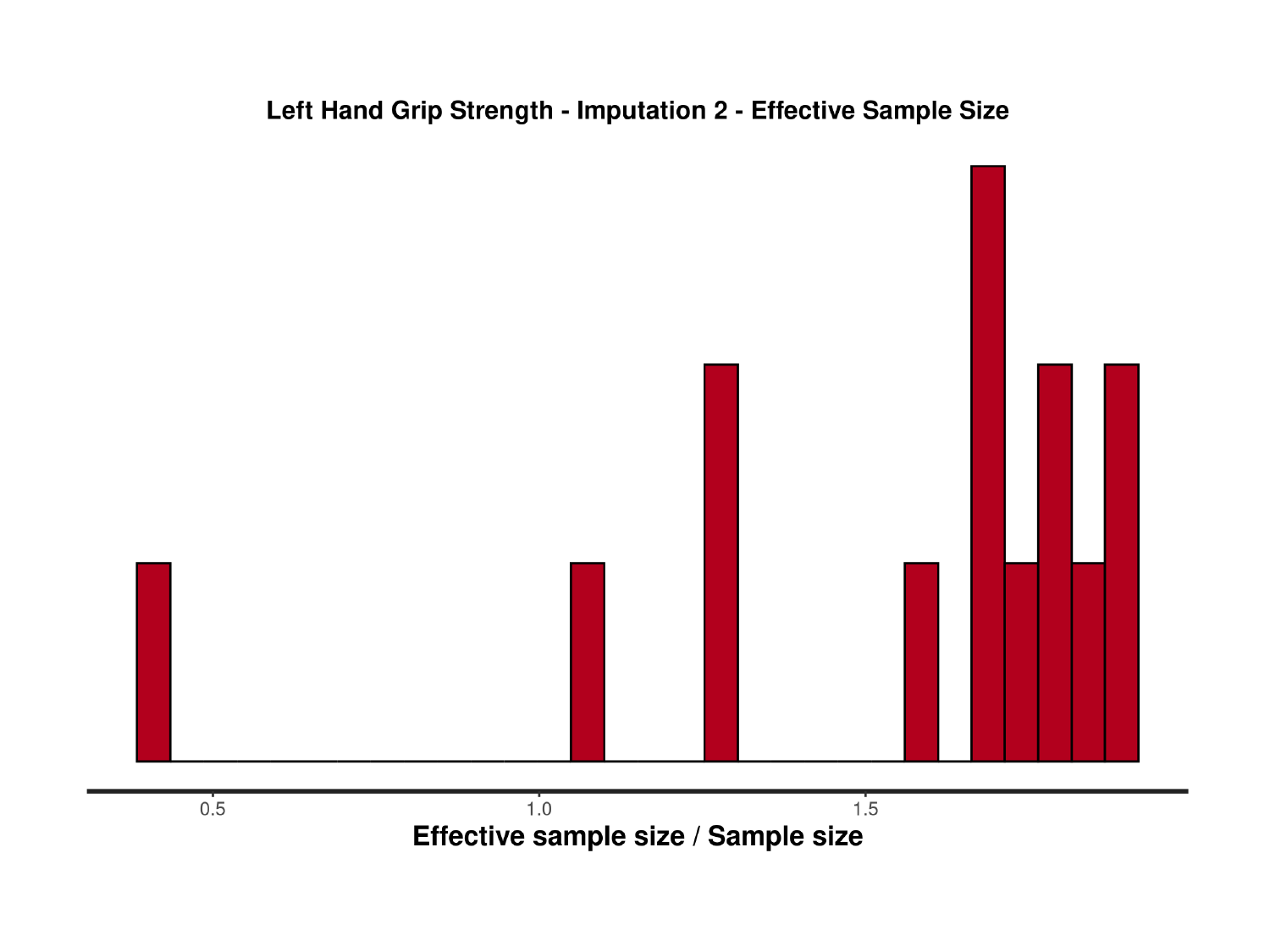

### Slide 140
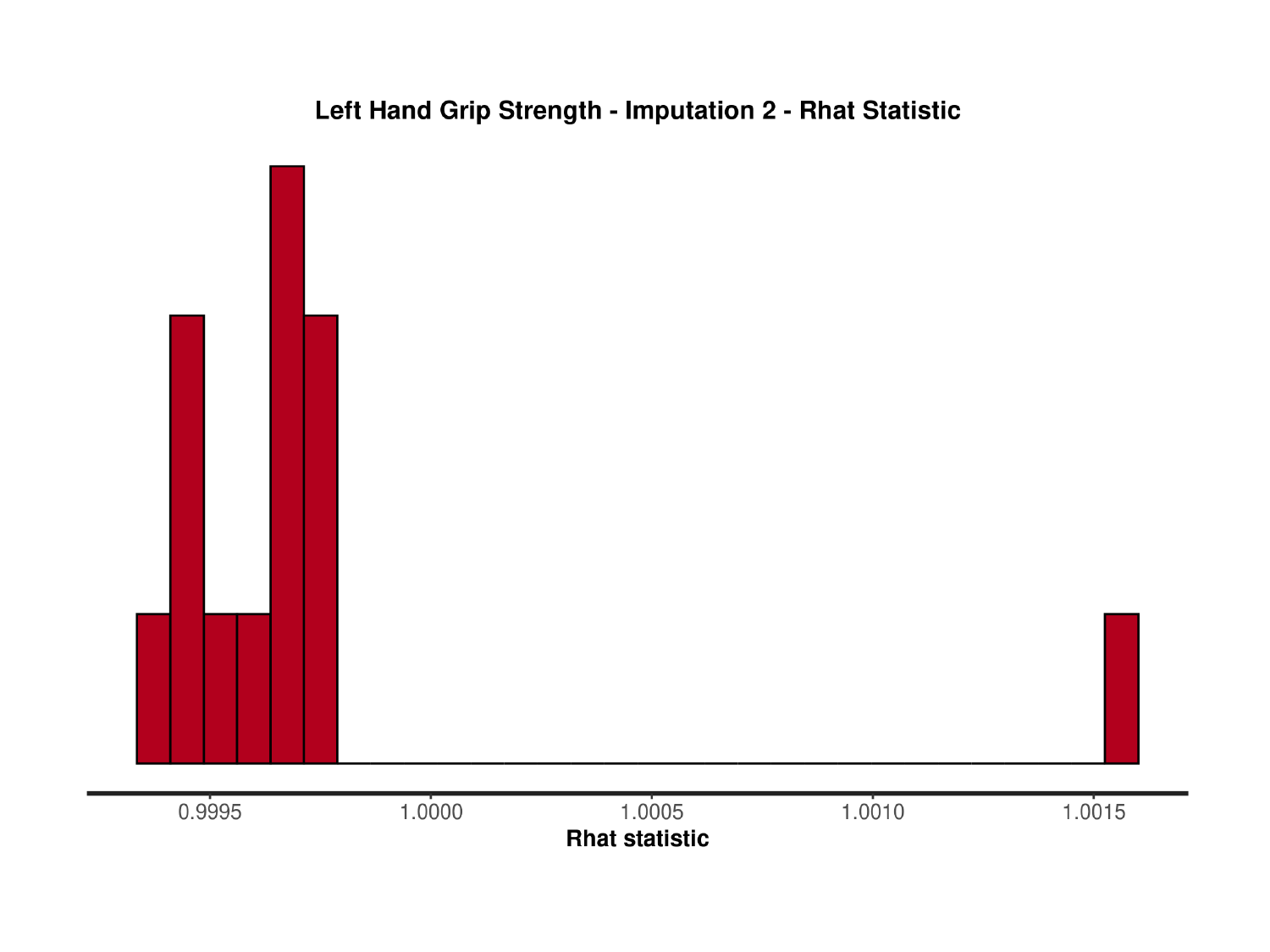

### Slide 141
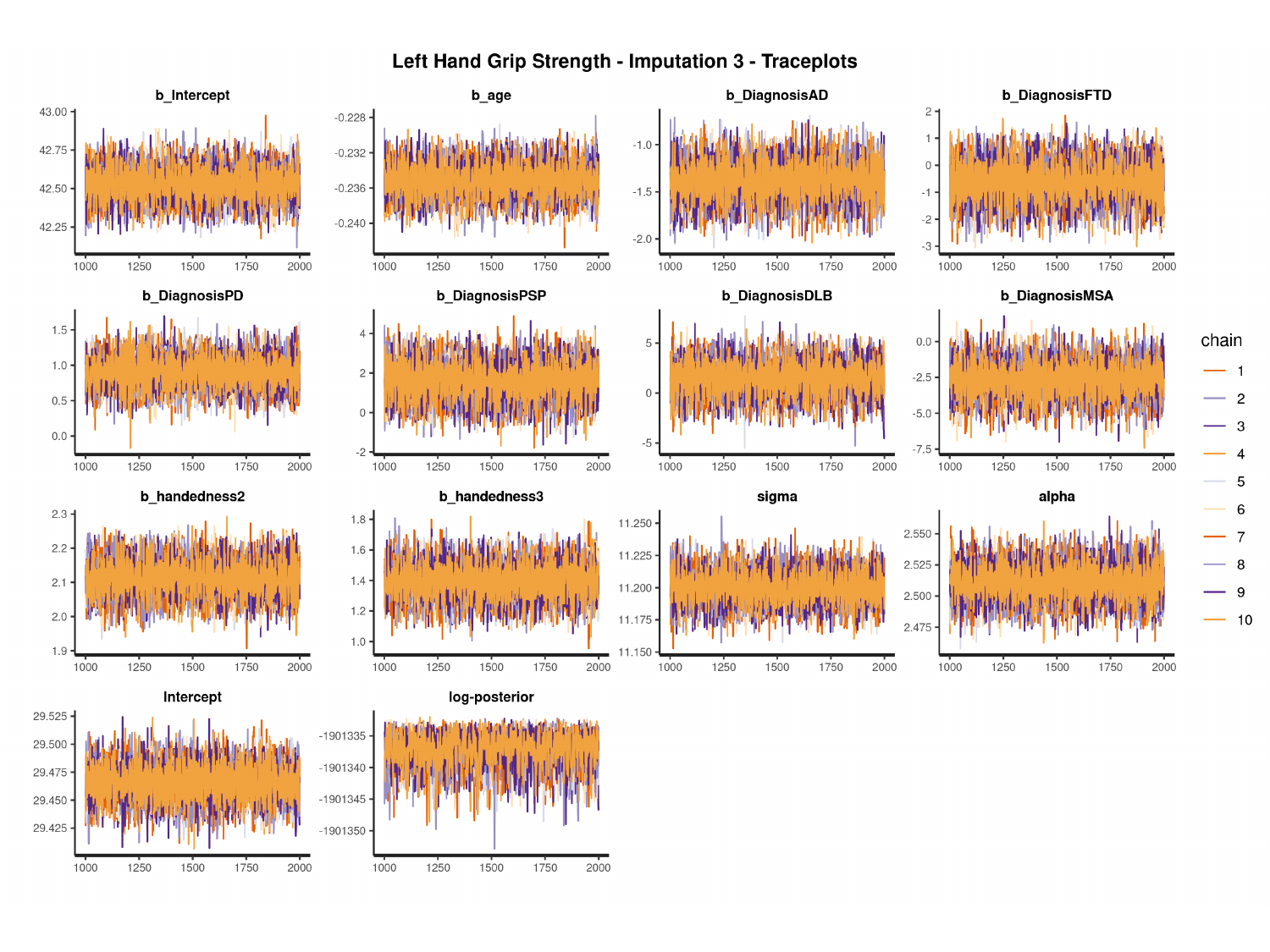

#

### Slide 142
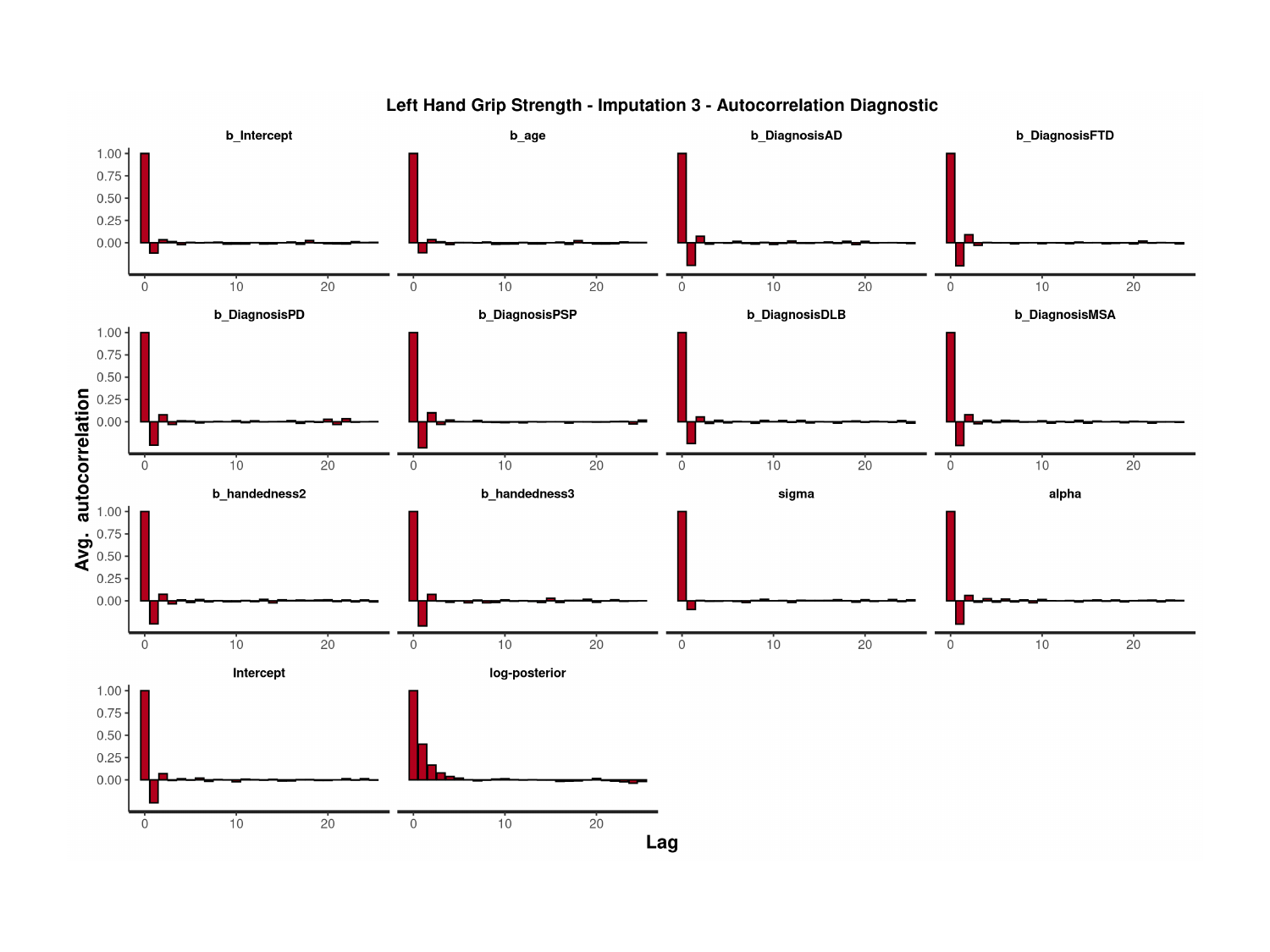

### Slide 143
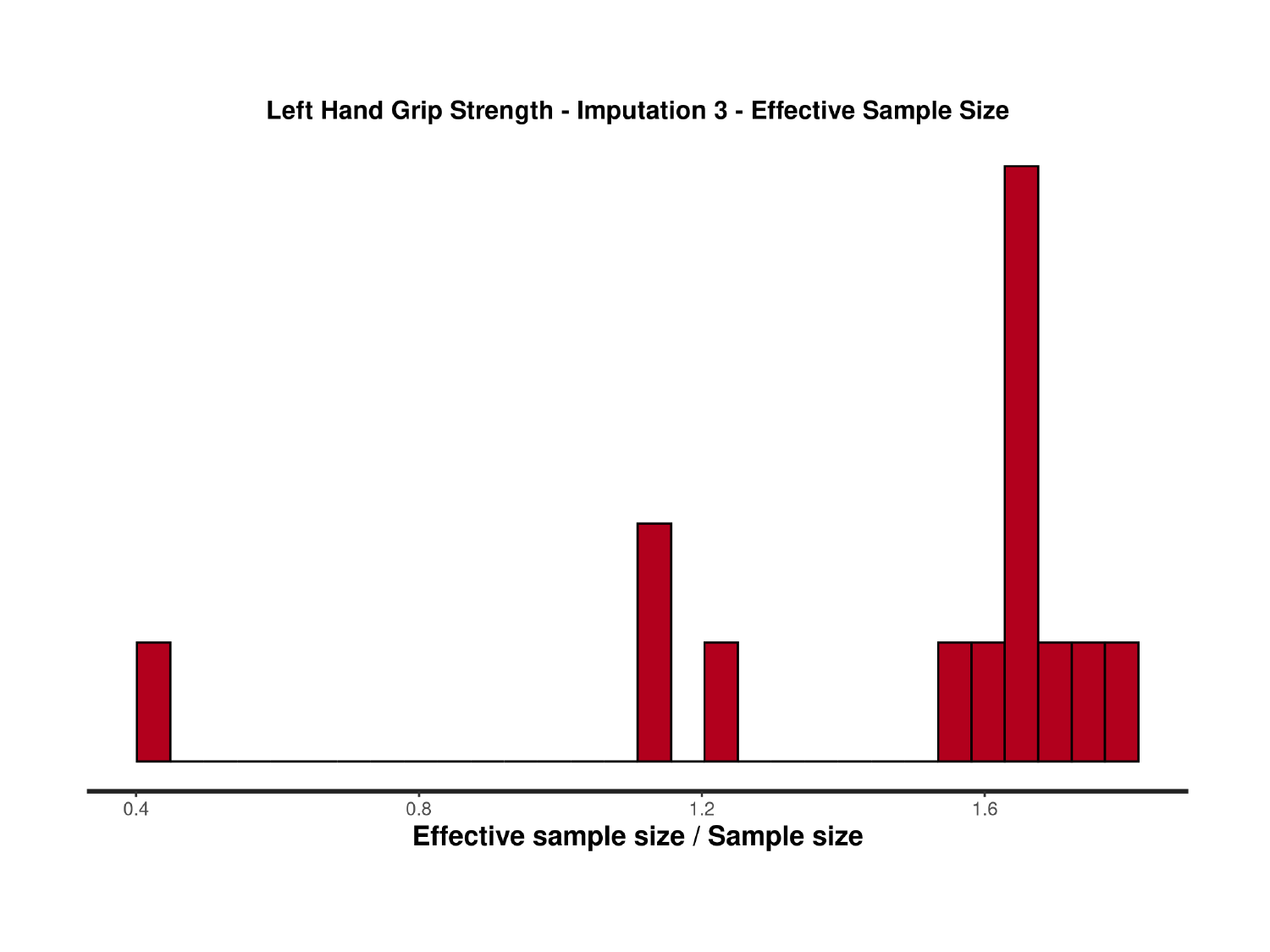

### Slide 144
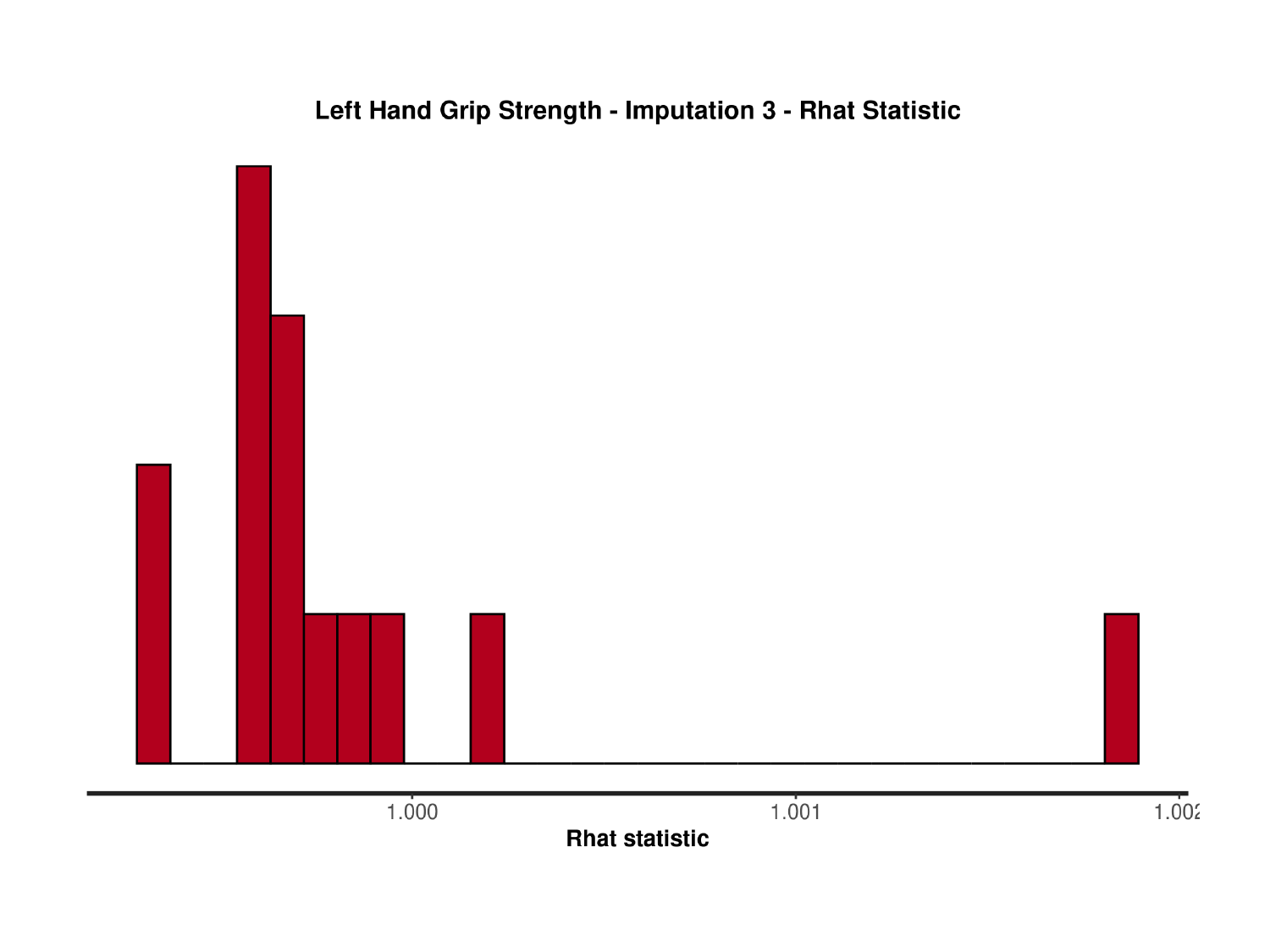

### Slide 145
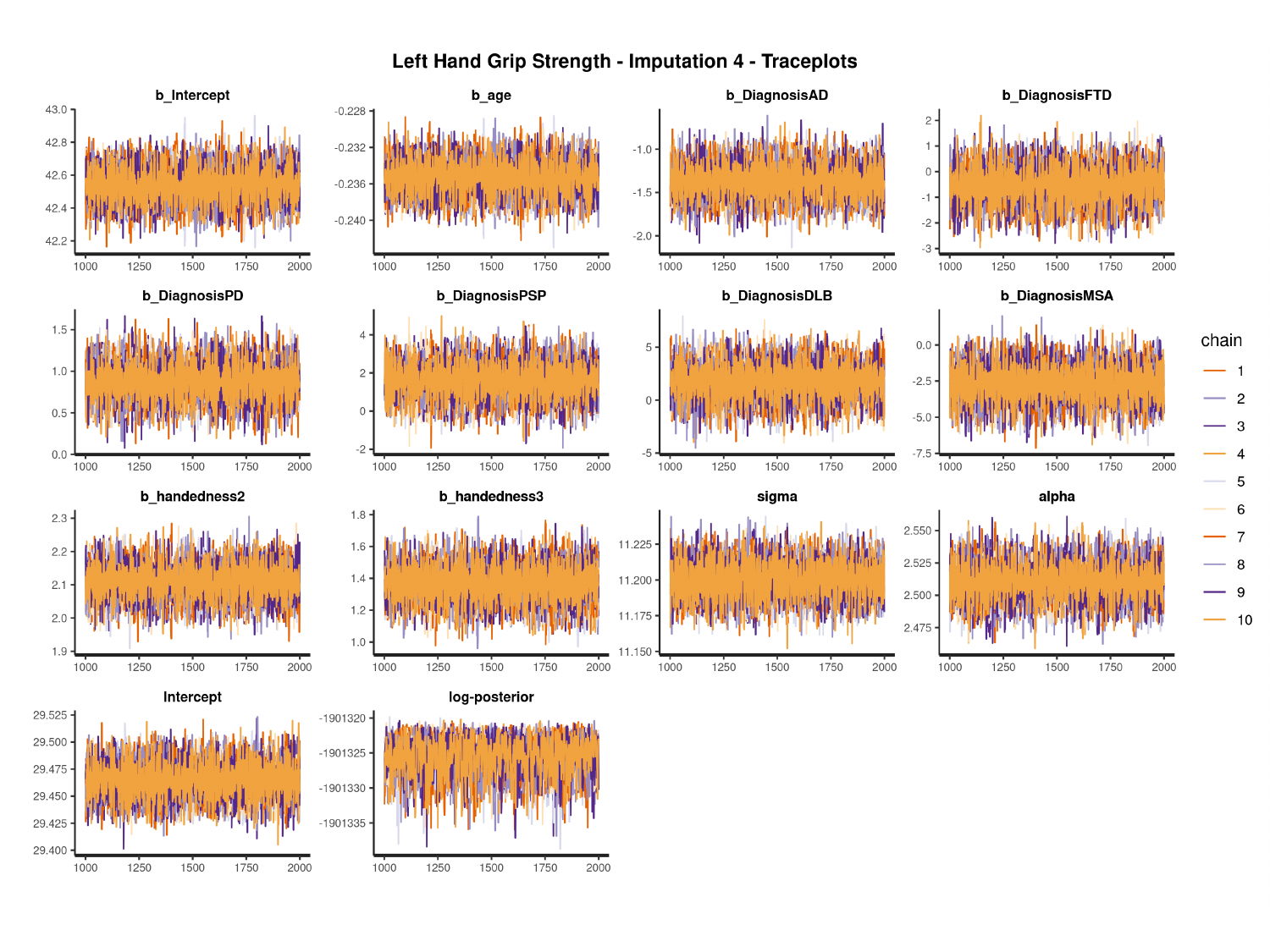

#

### Slide 146
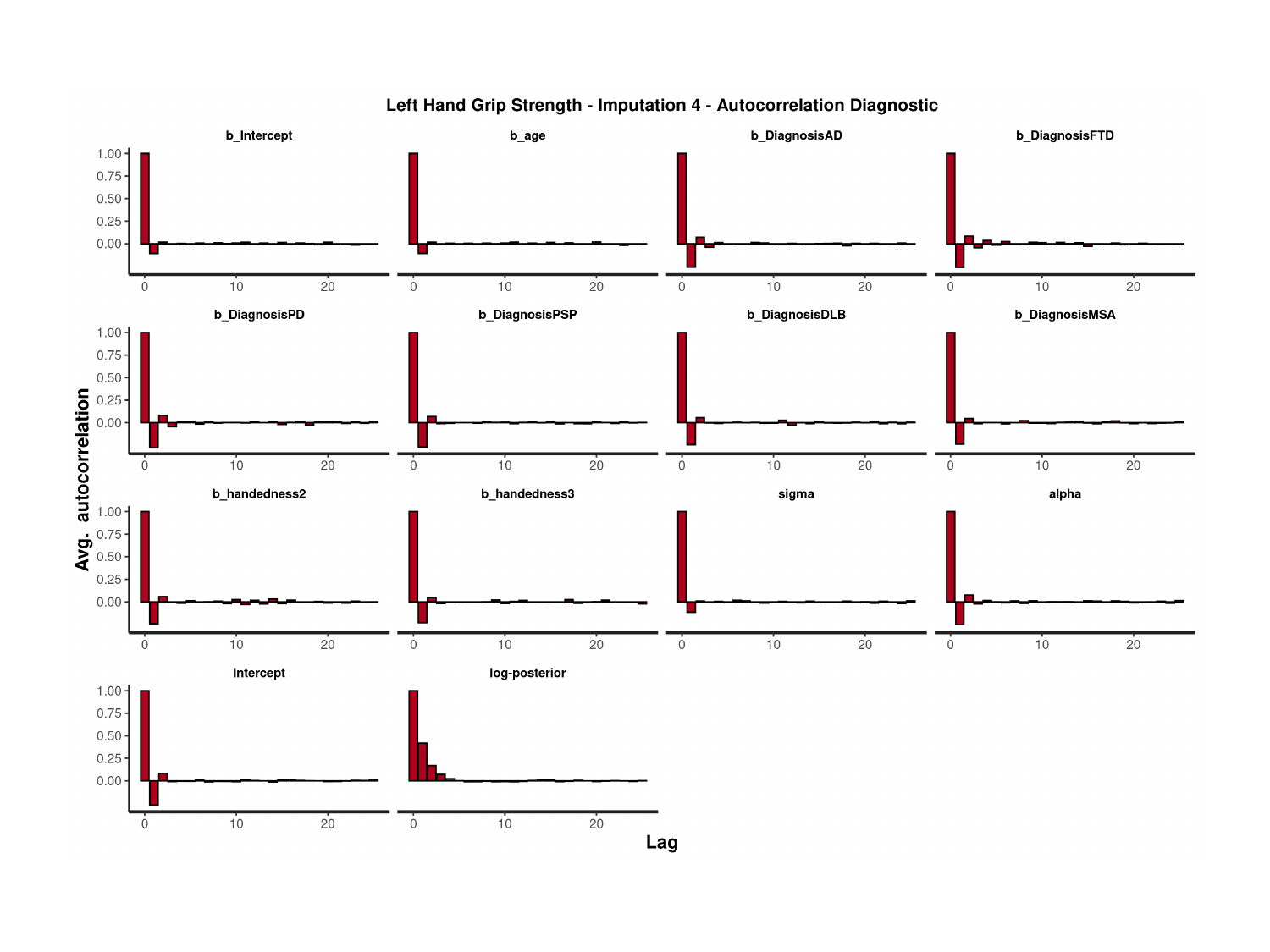

### Slide 147
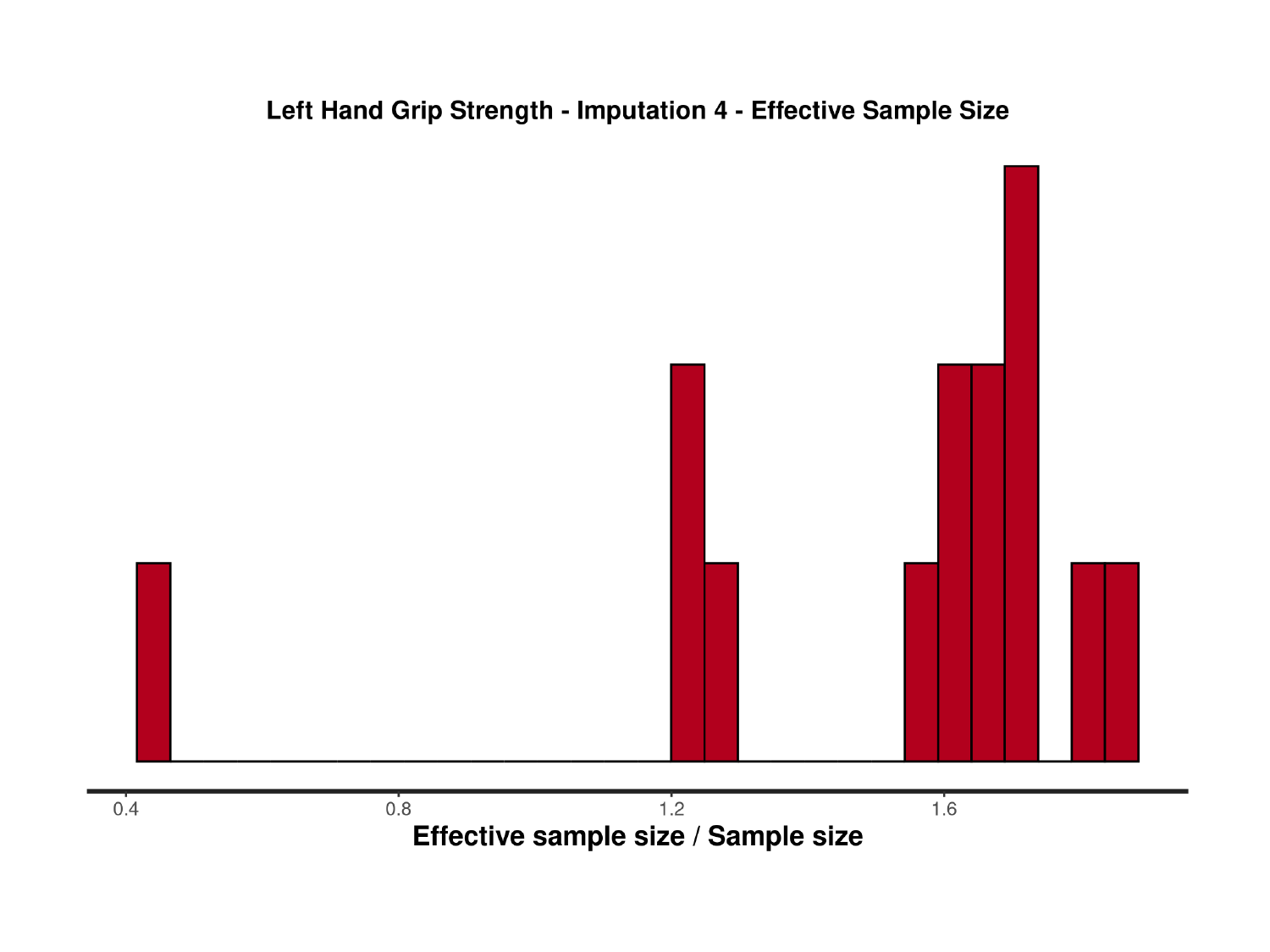

### Slide 148
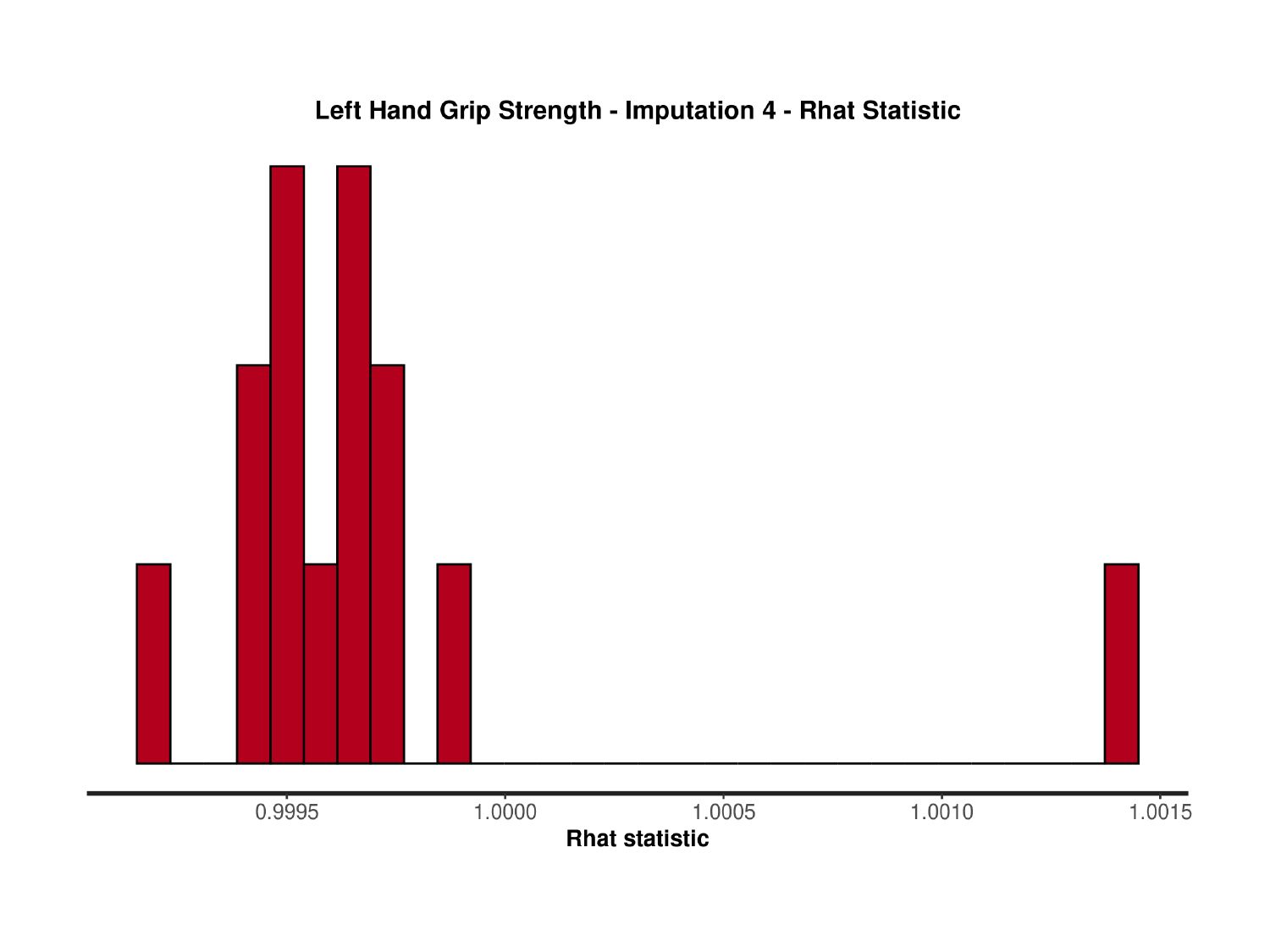

### Slide 149
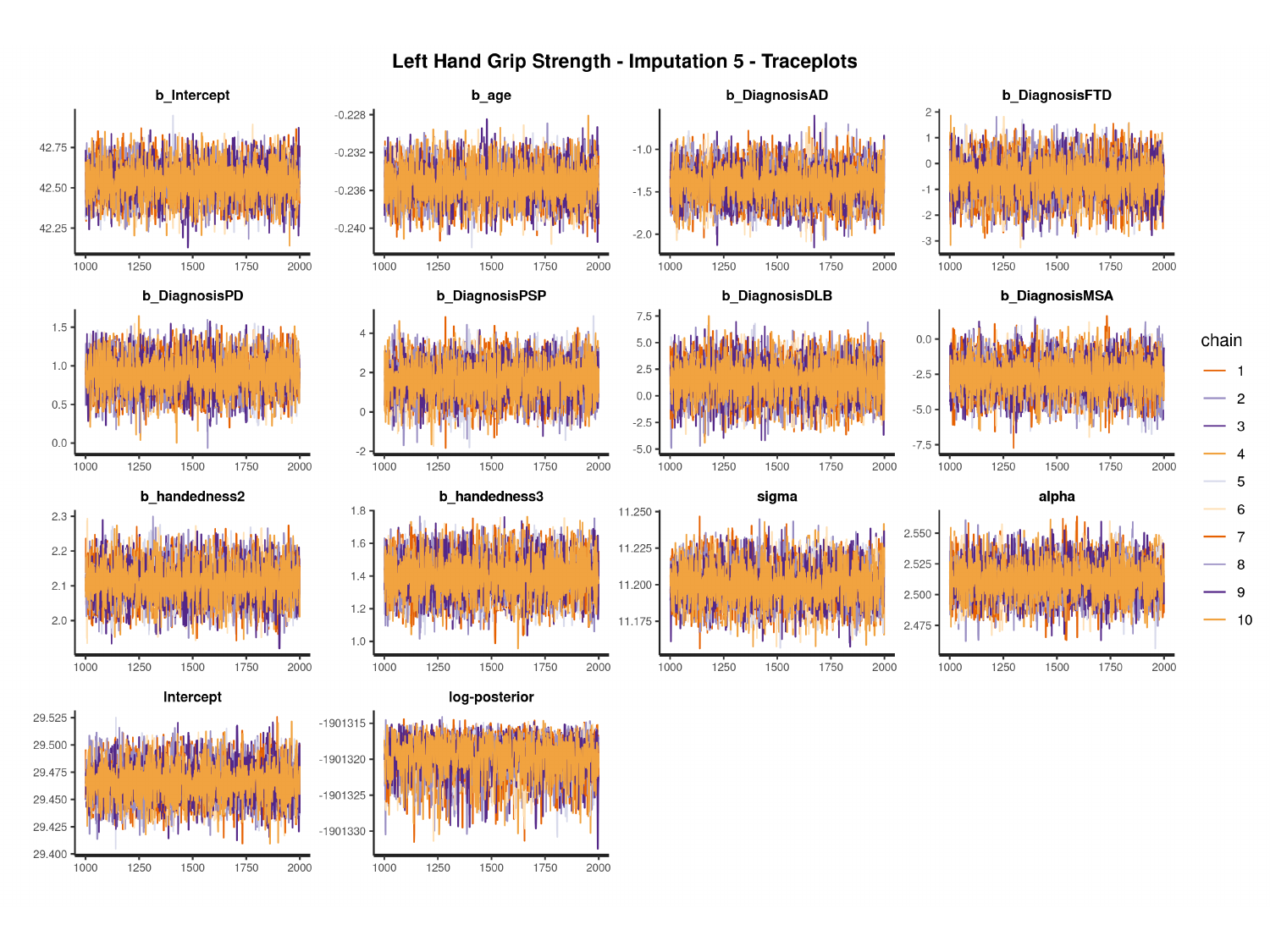

#

### Slide 150
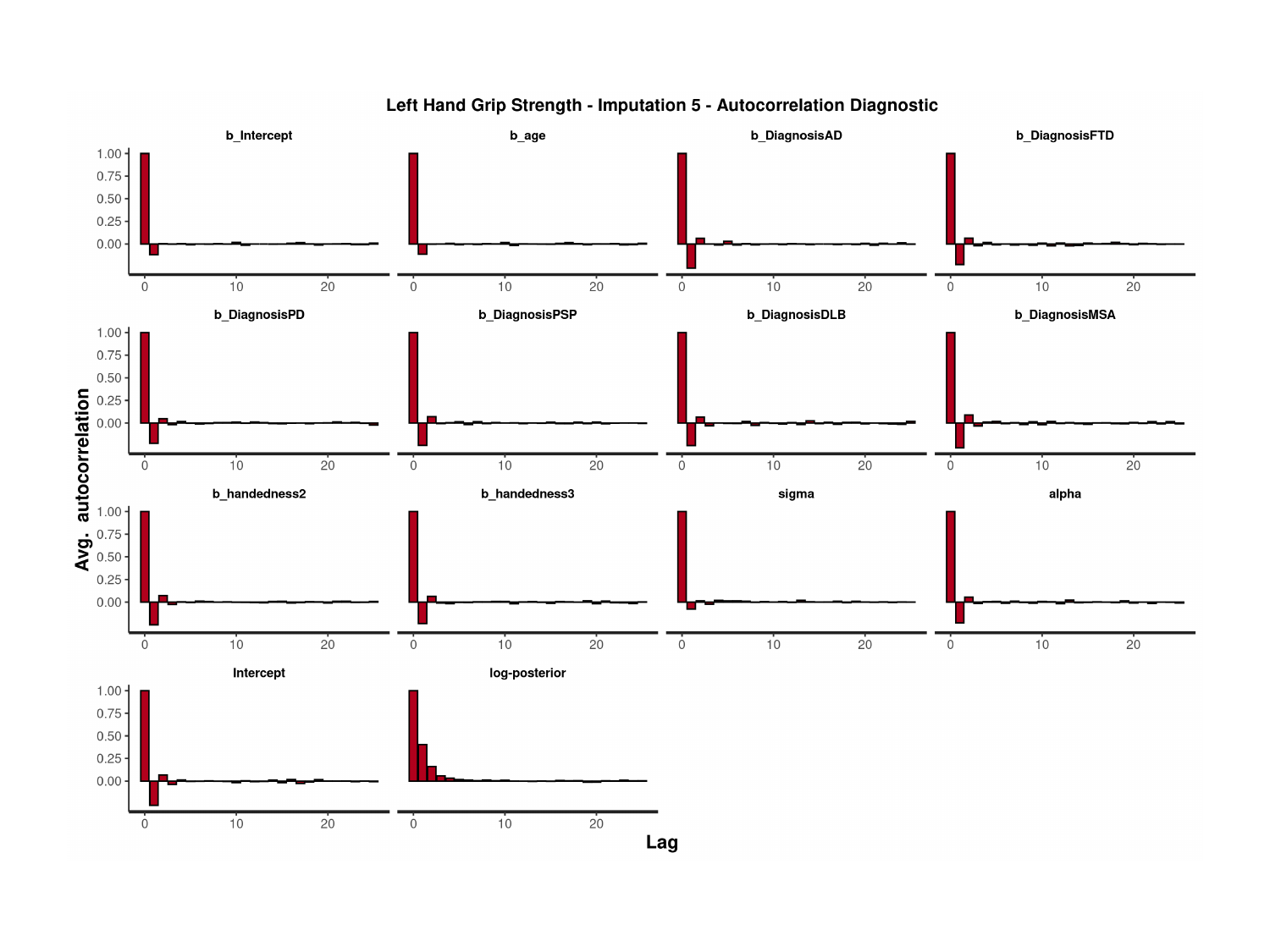

### Slide 151
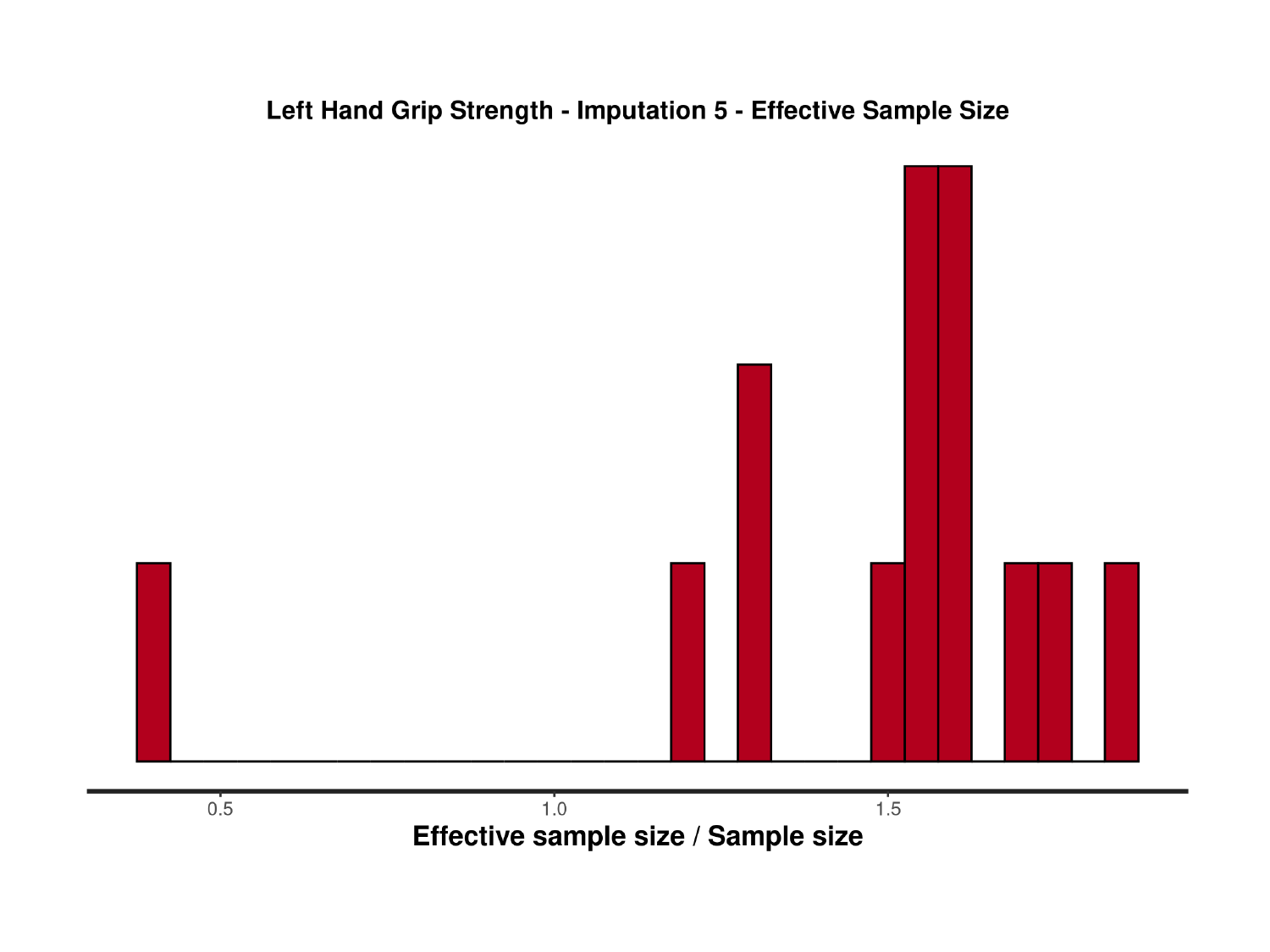

### Slide 152
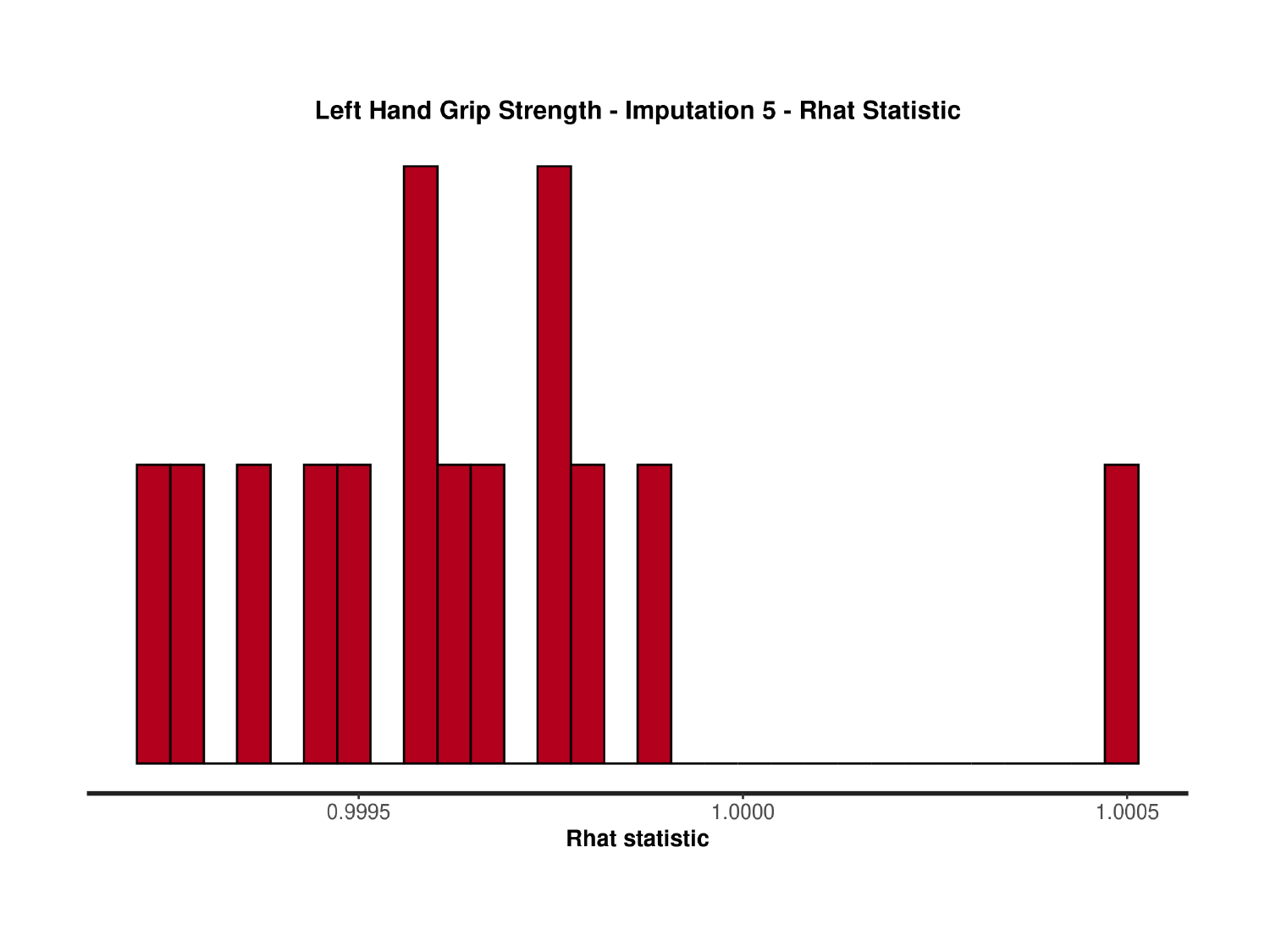

### Slide 153
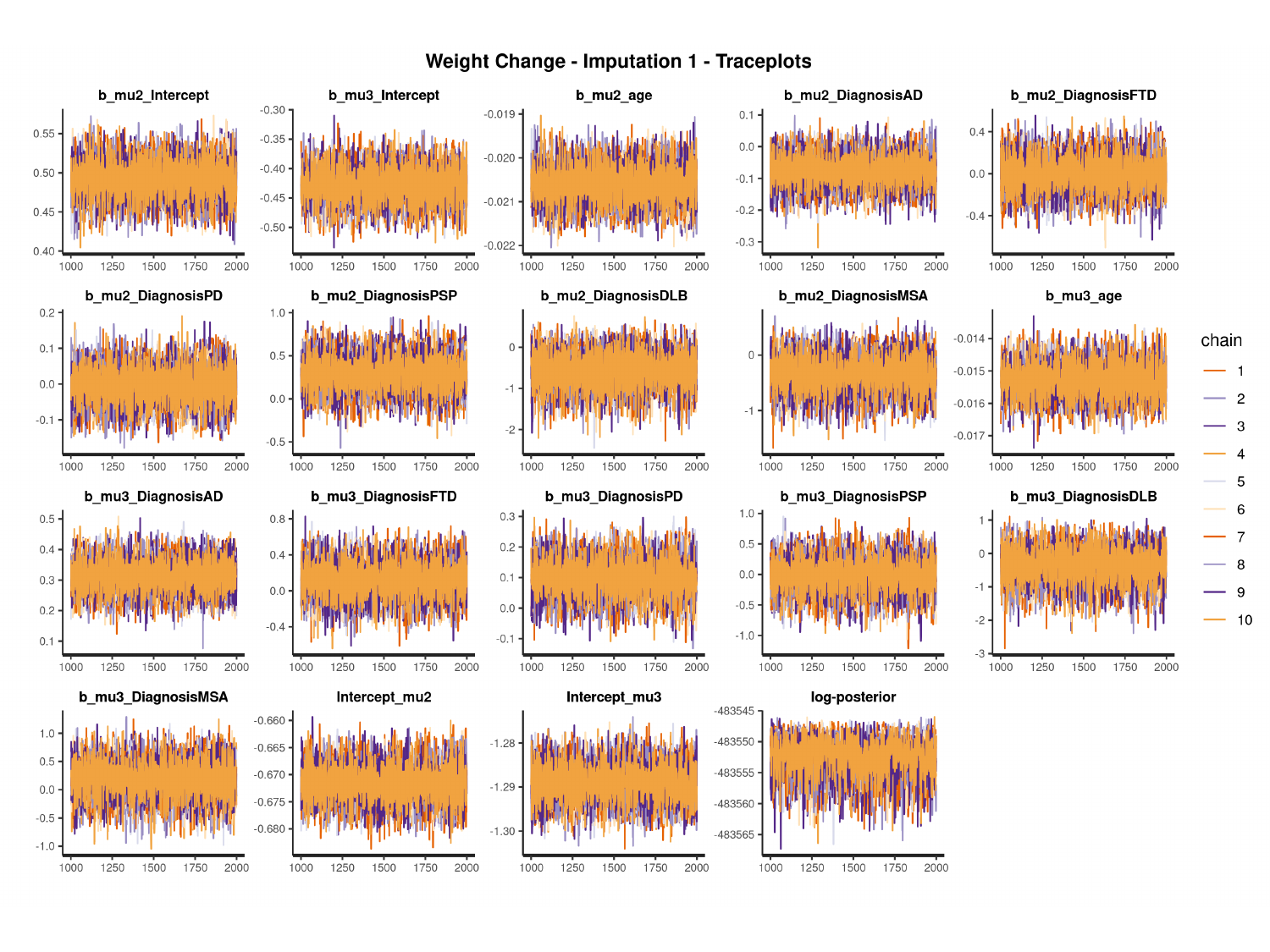

#

### Slide 154
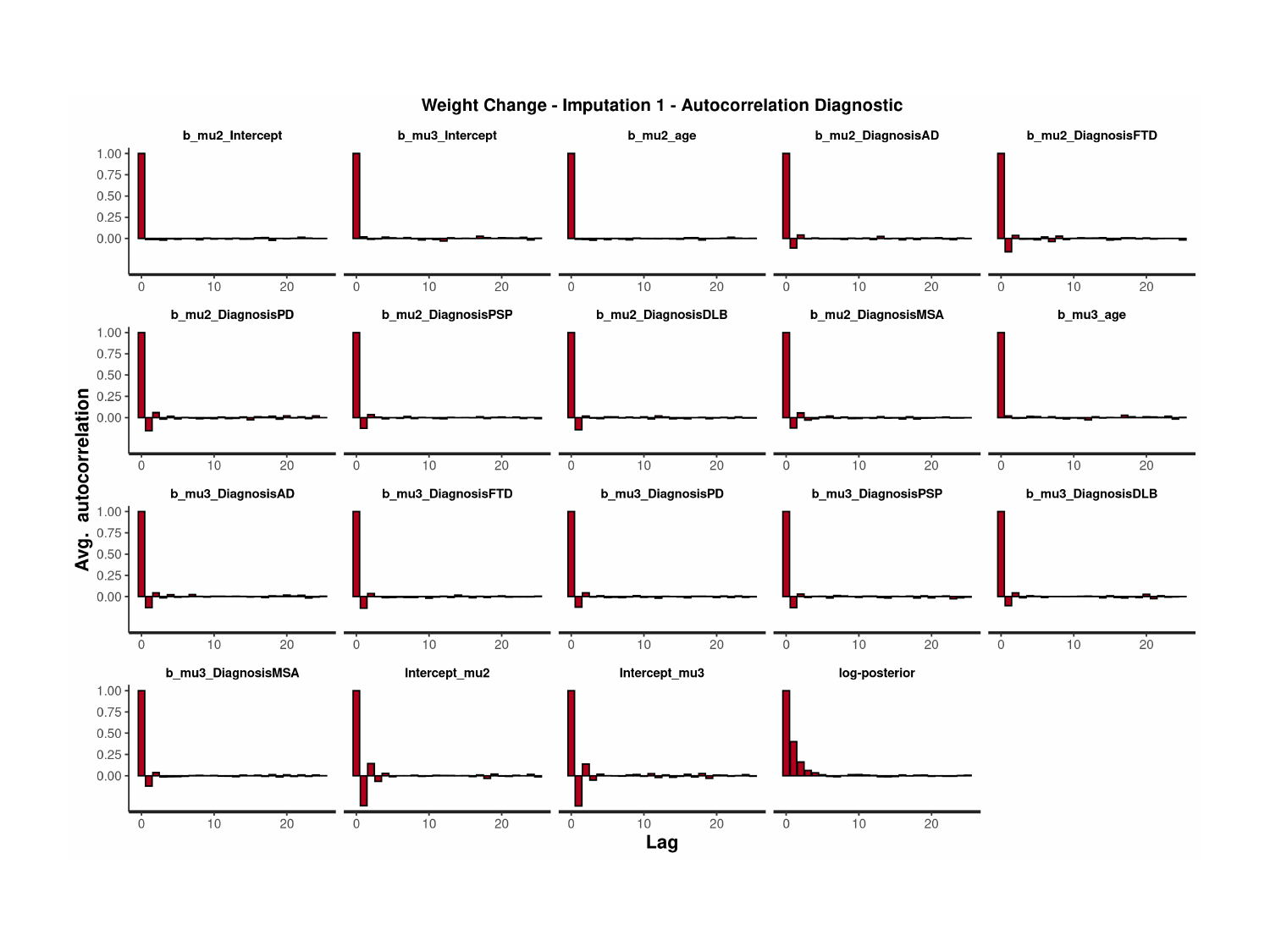

### Slide 155
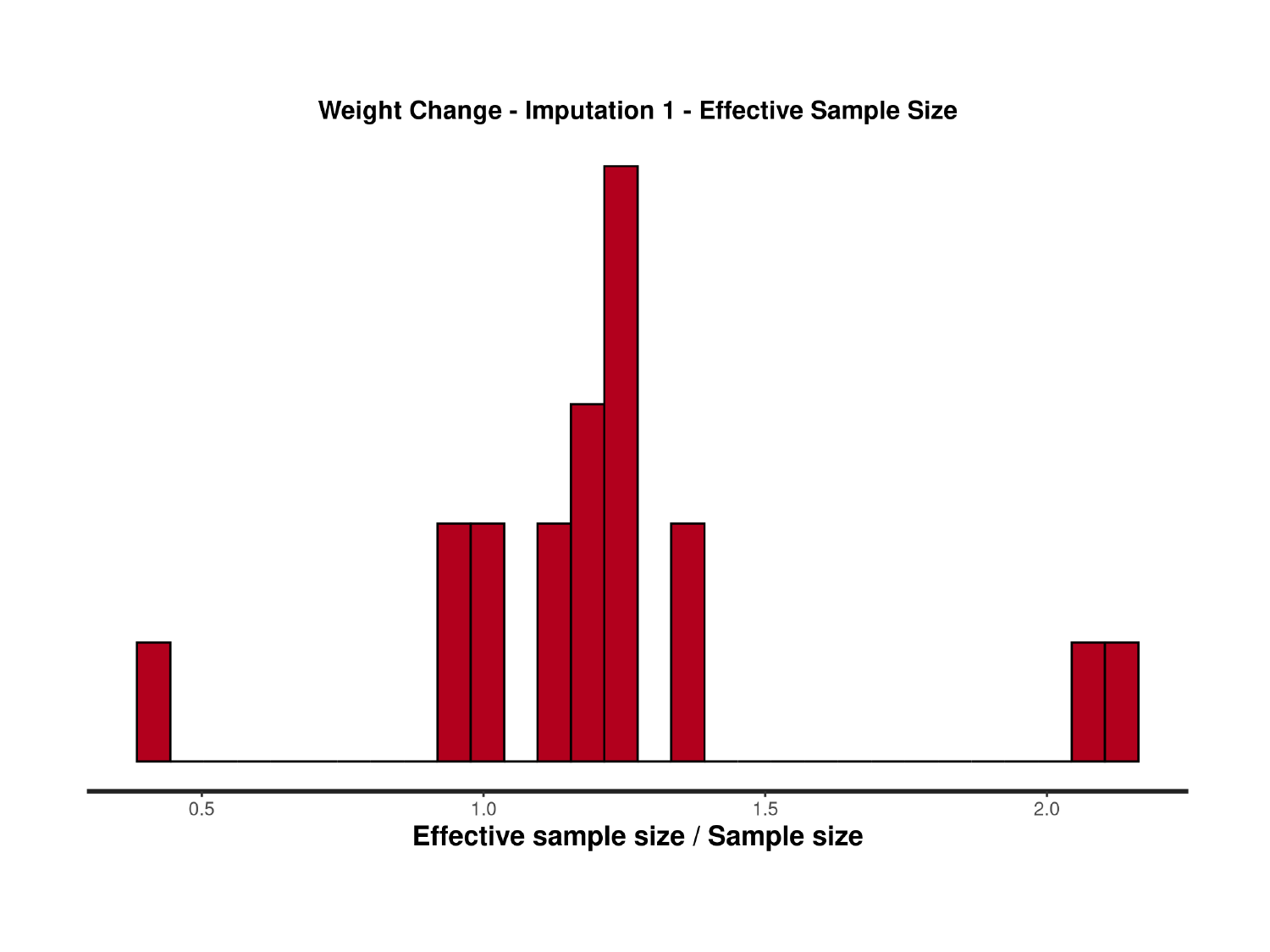

### Slide 156
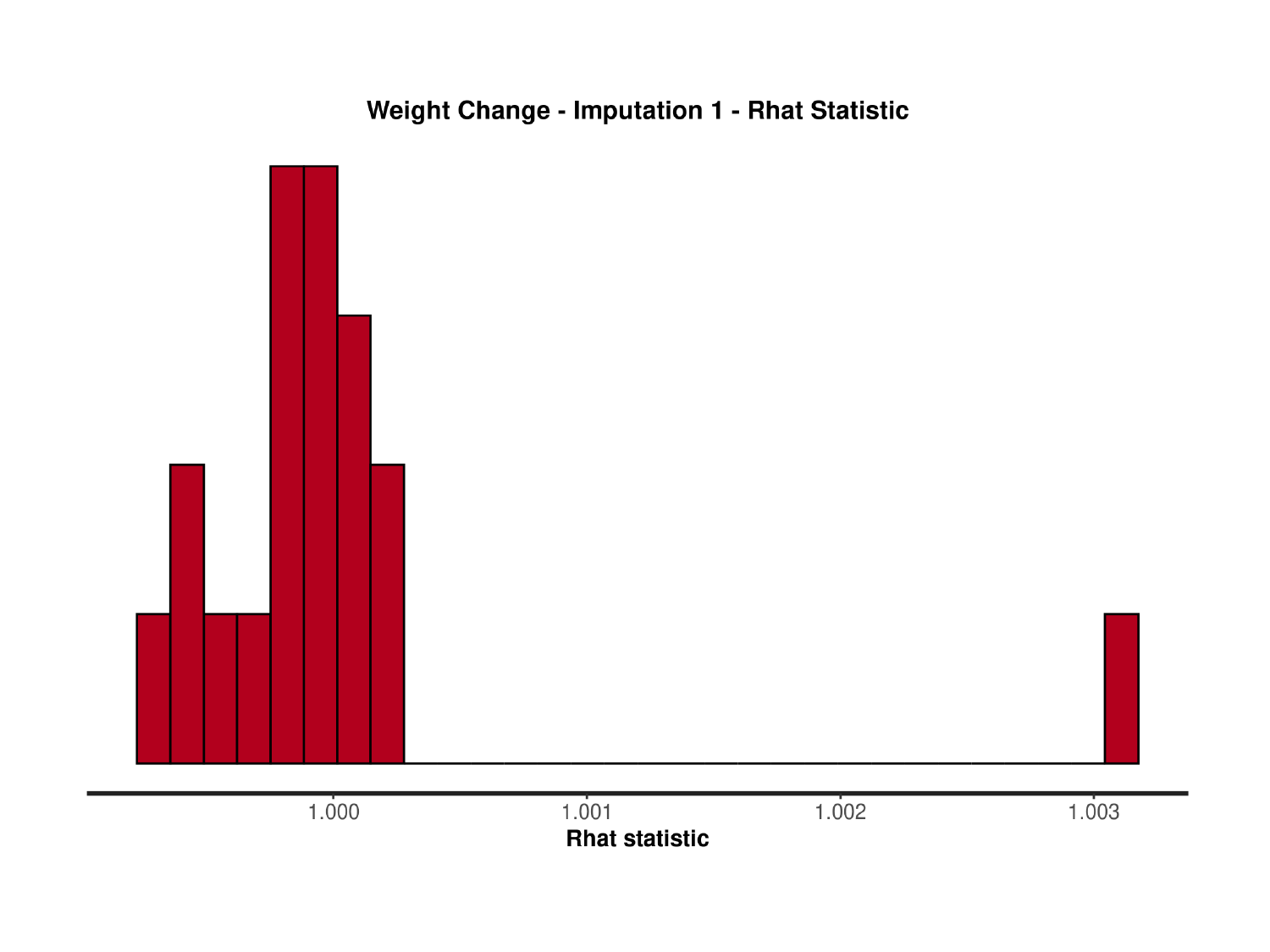

### Slide 157
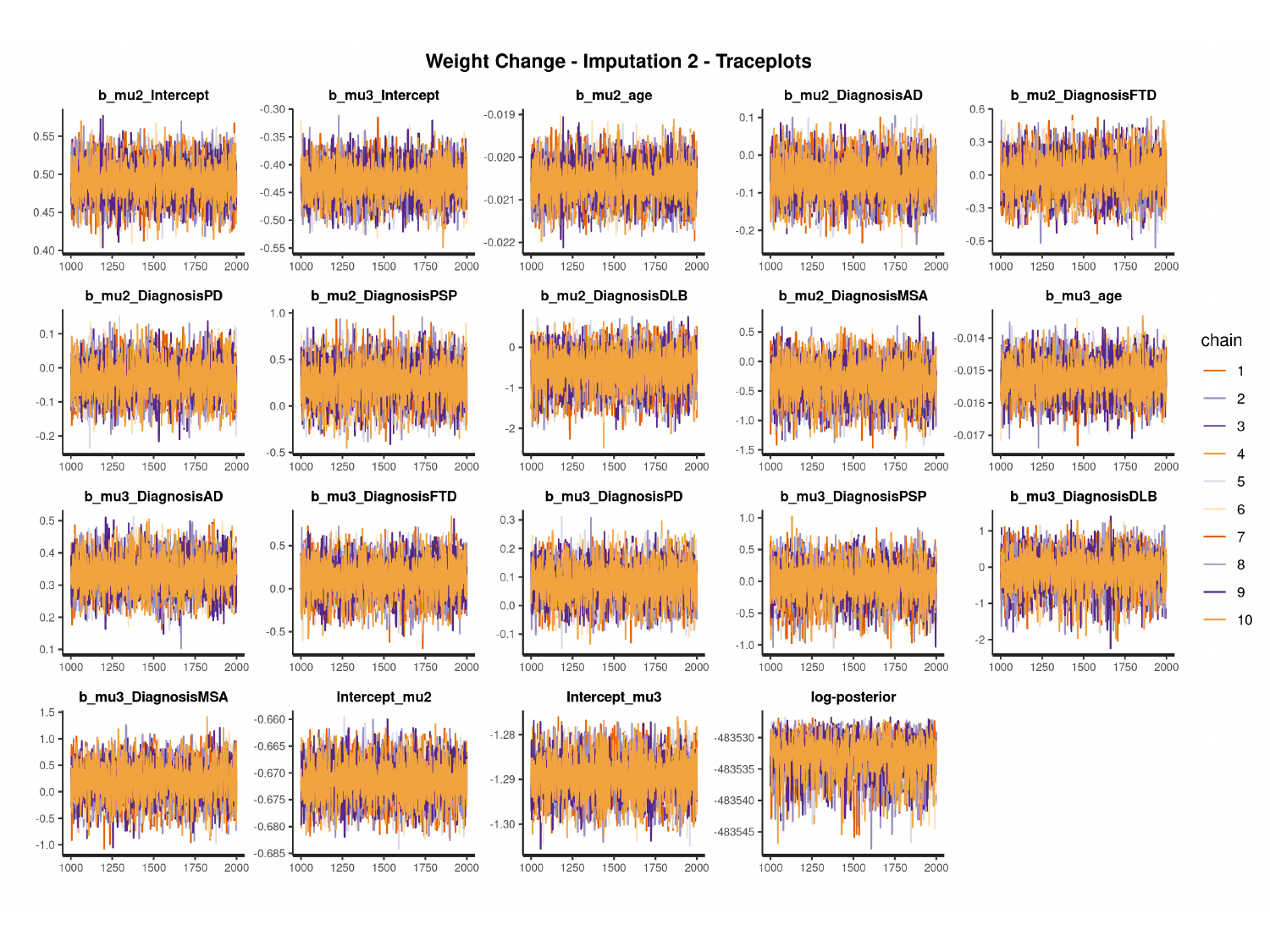

#

### Slide 158
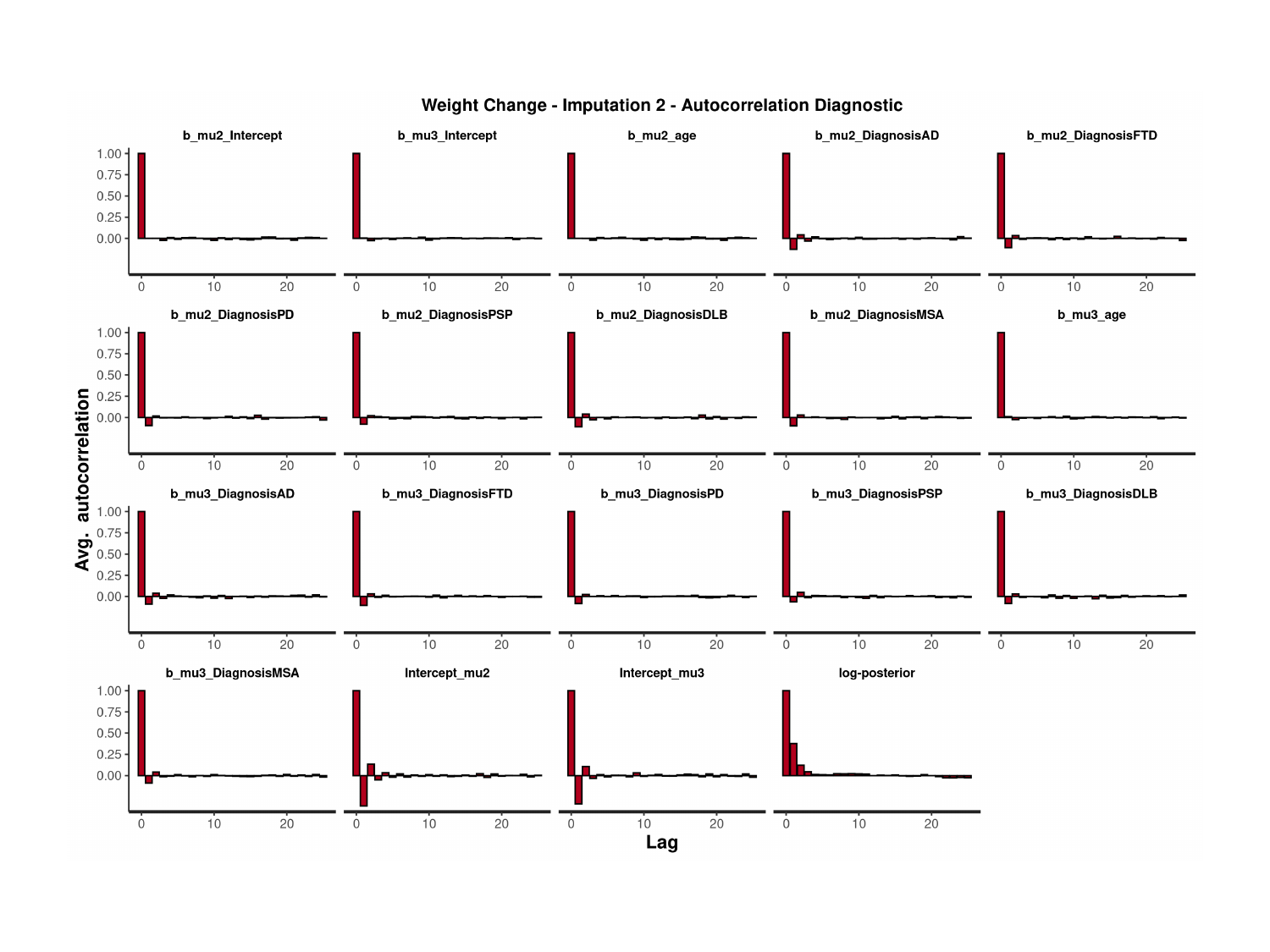

### Slide 159
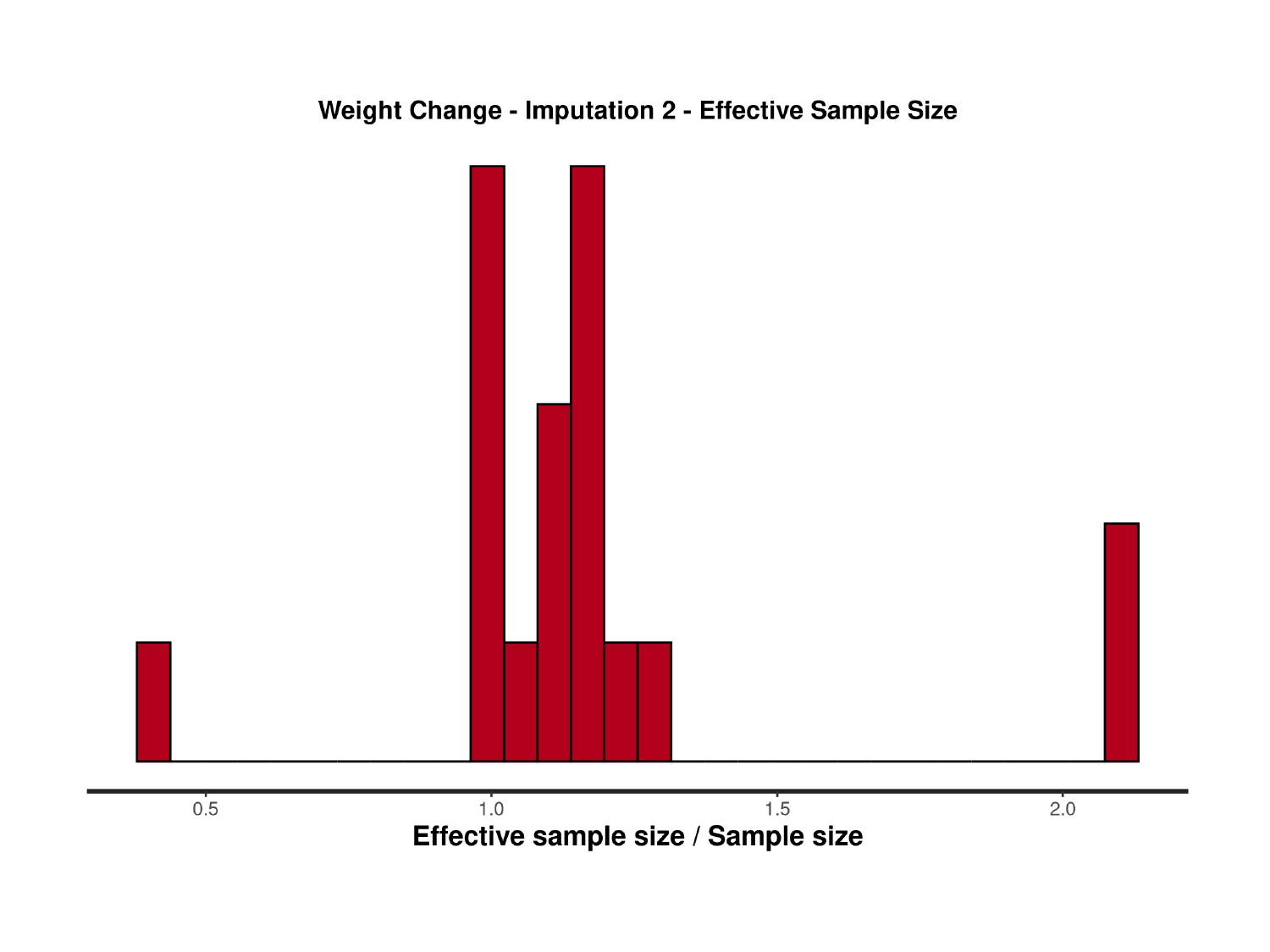

### Slide 160
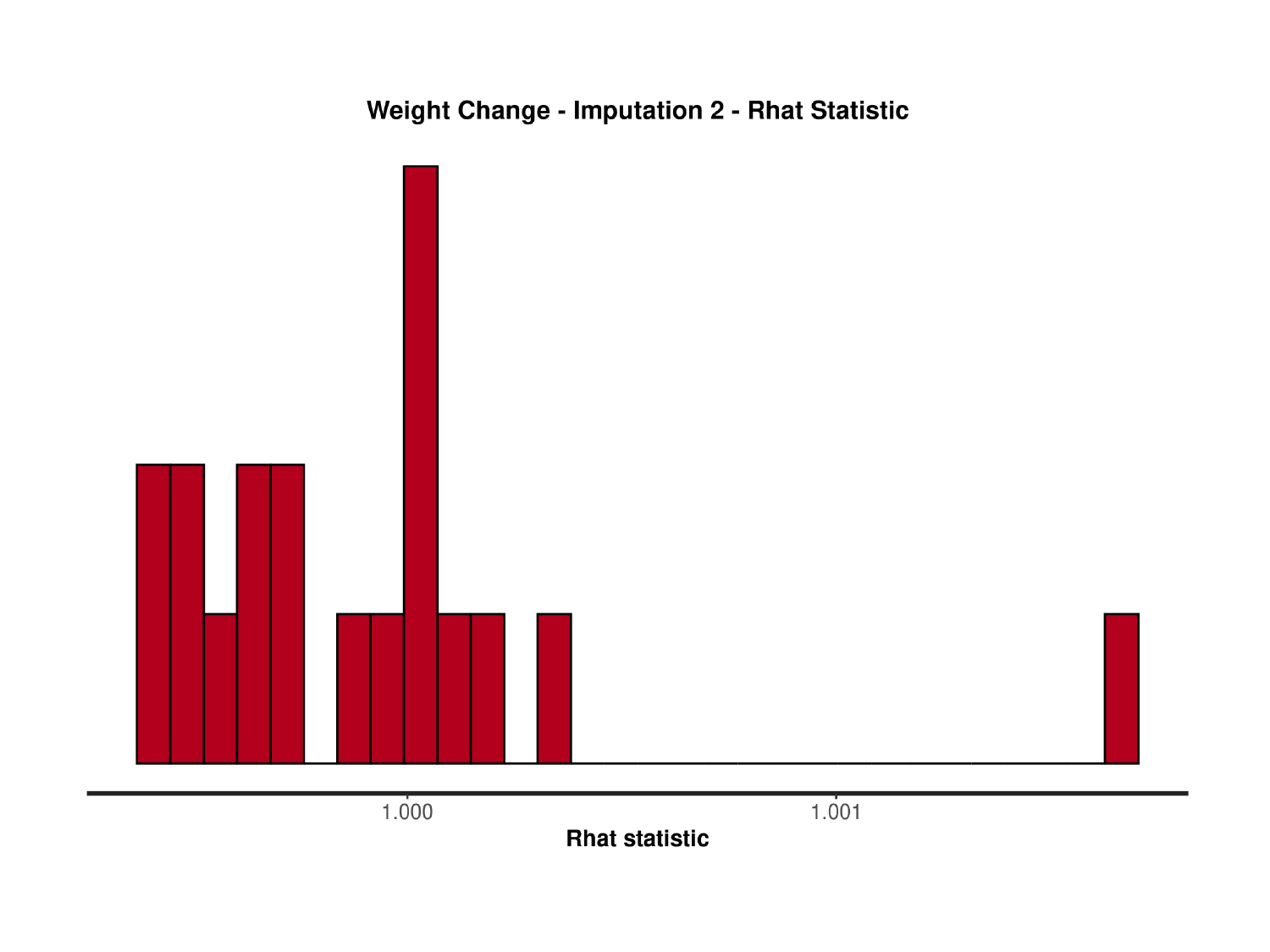

### Slide 161
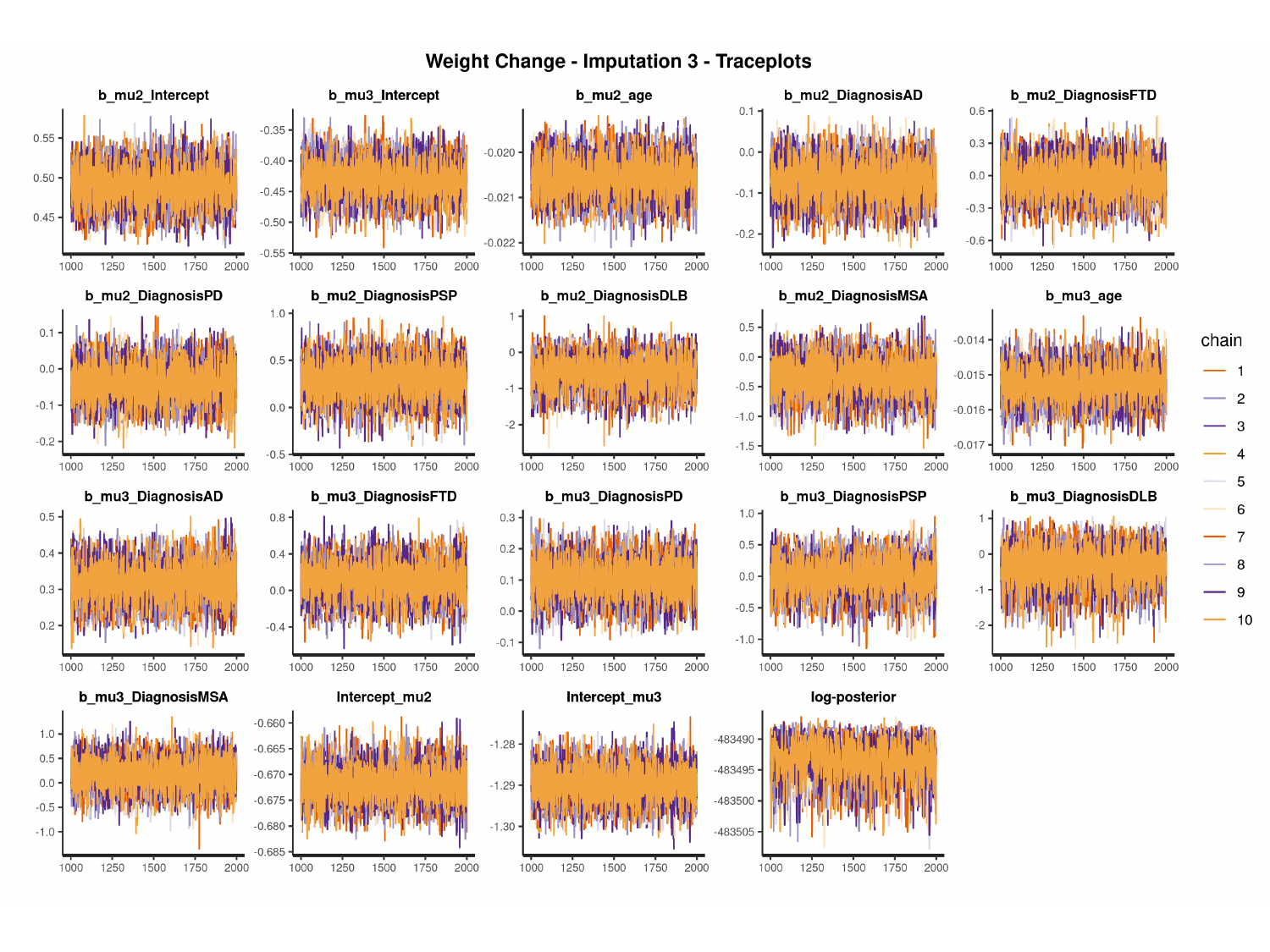

#

### Slide 162
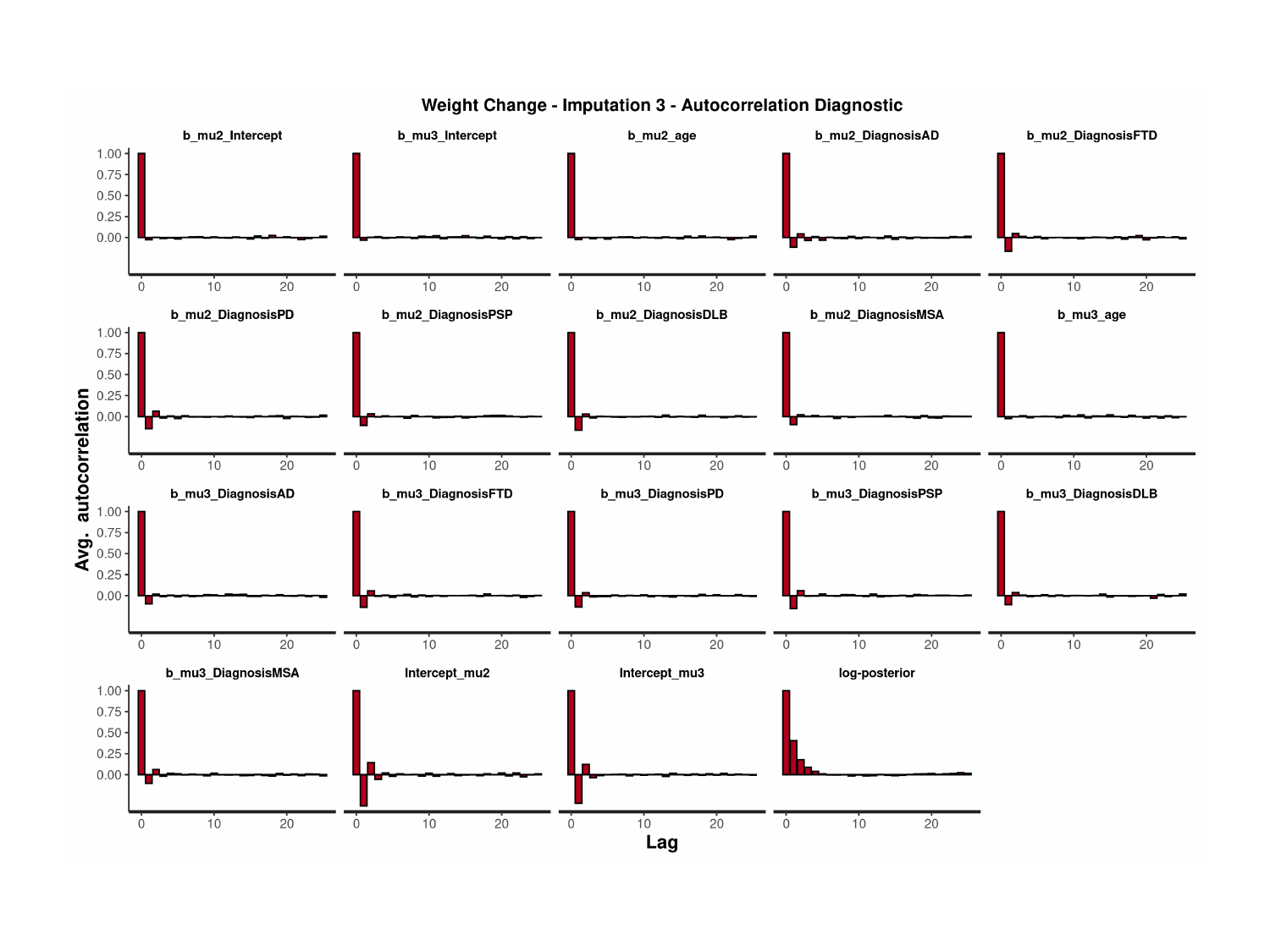

### Slide 163
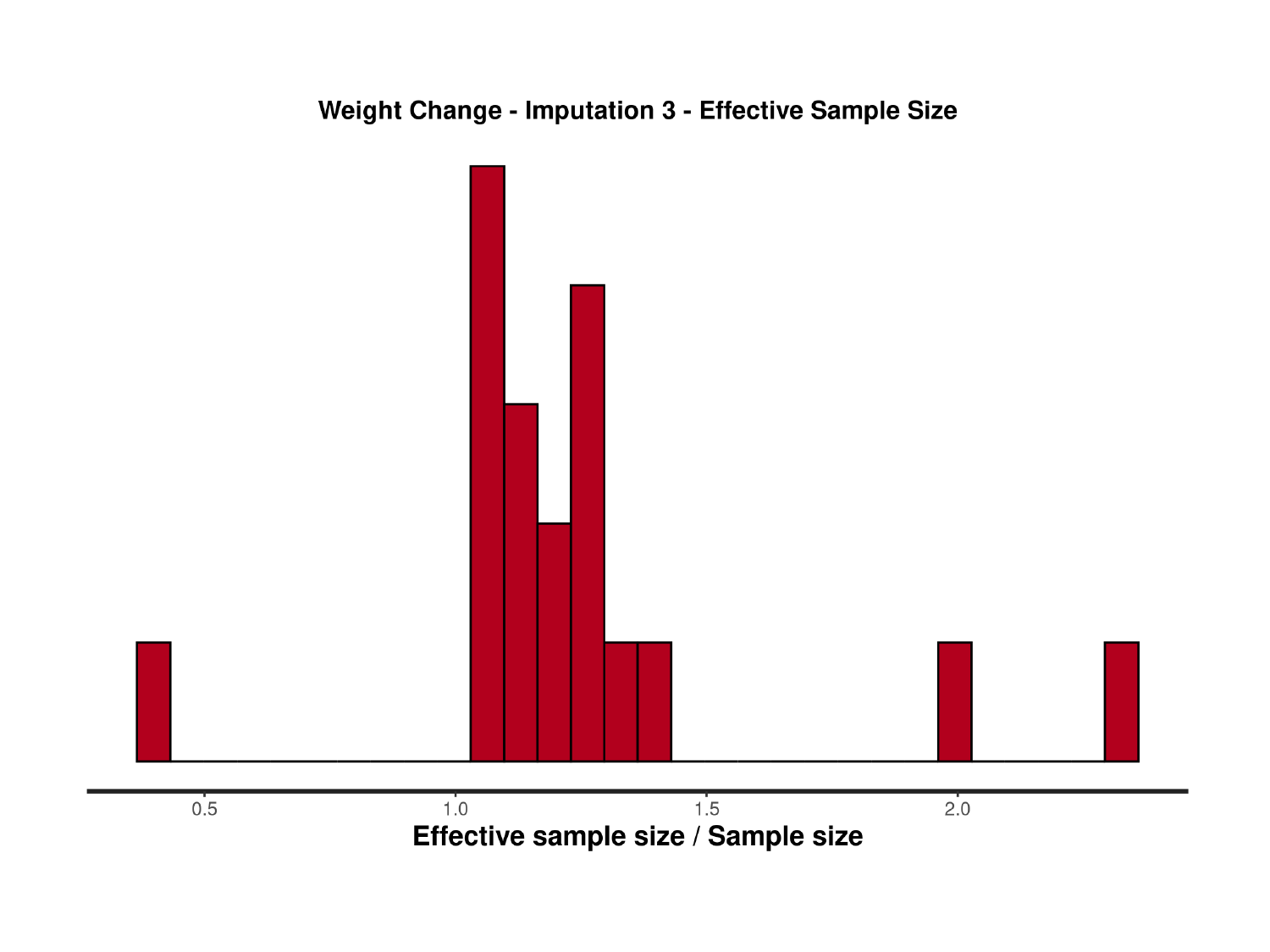

### Slide 164
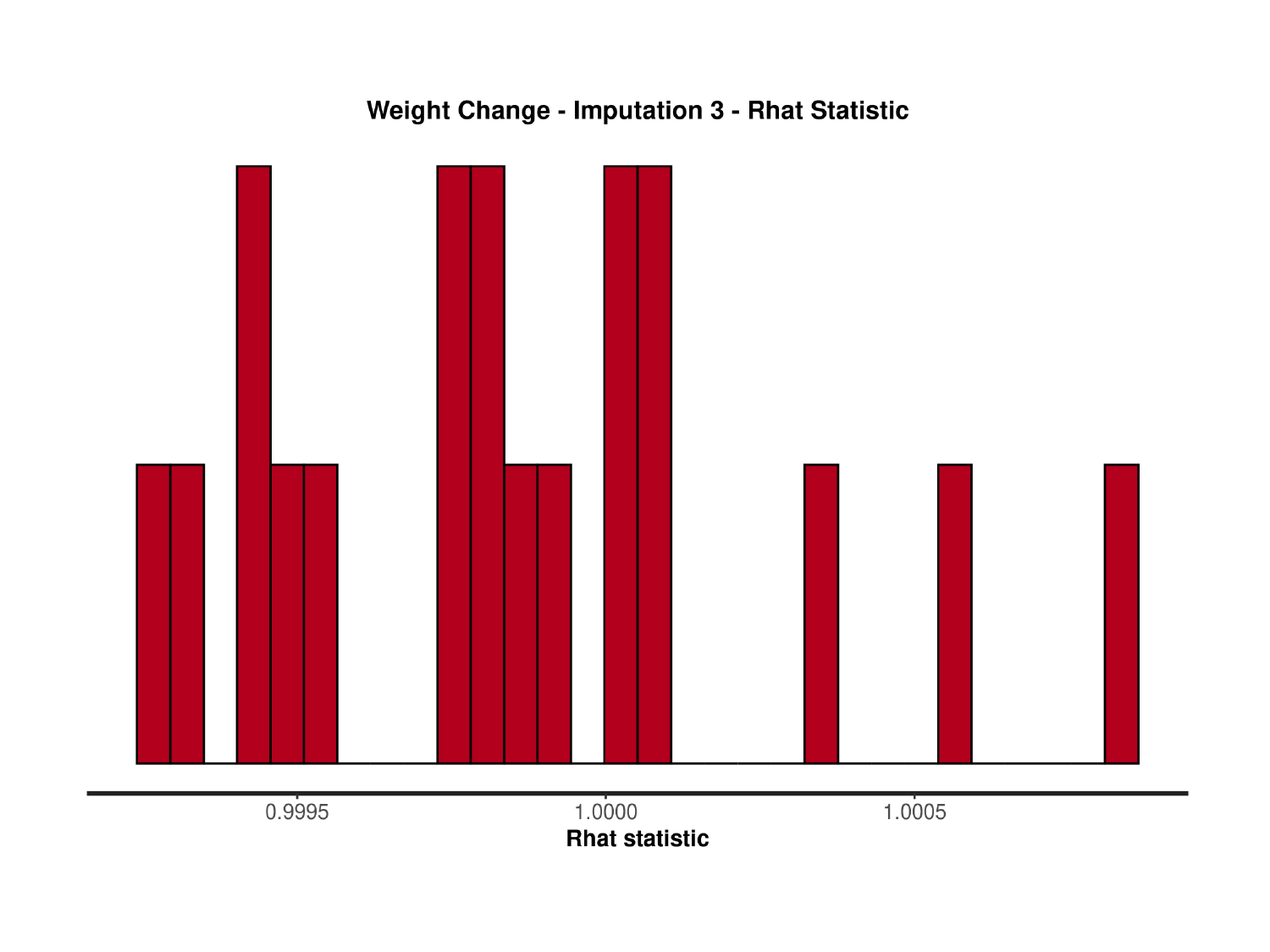

### Slide 165
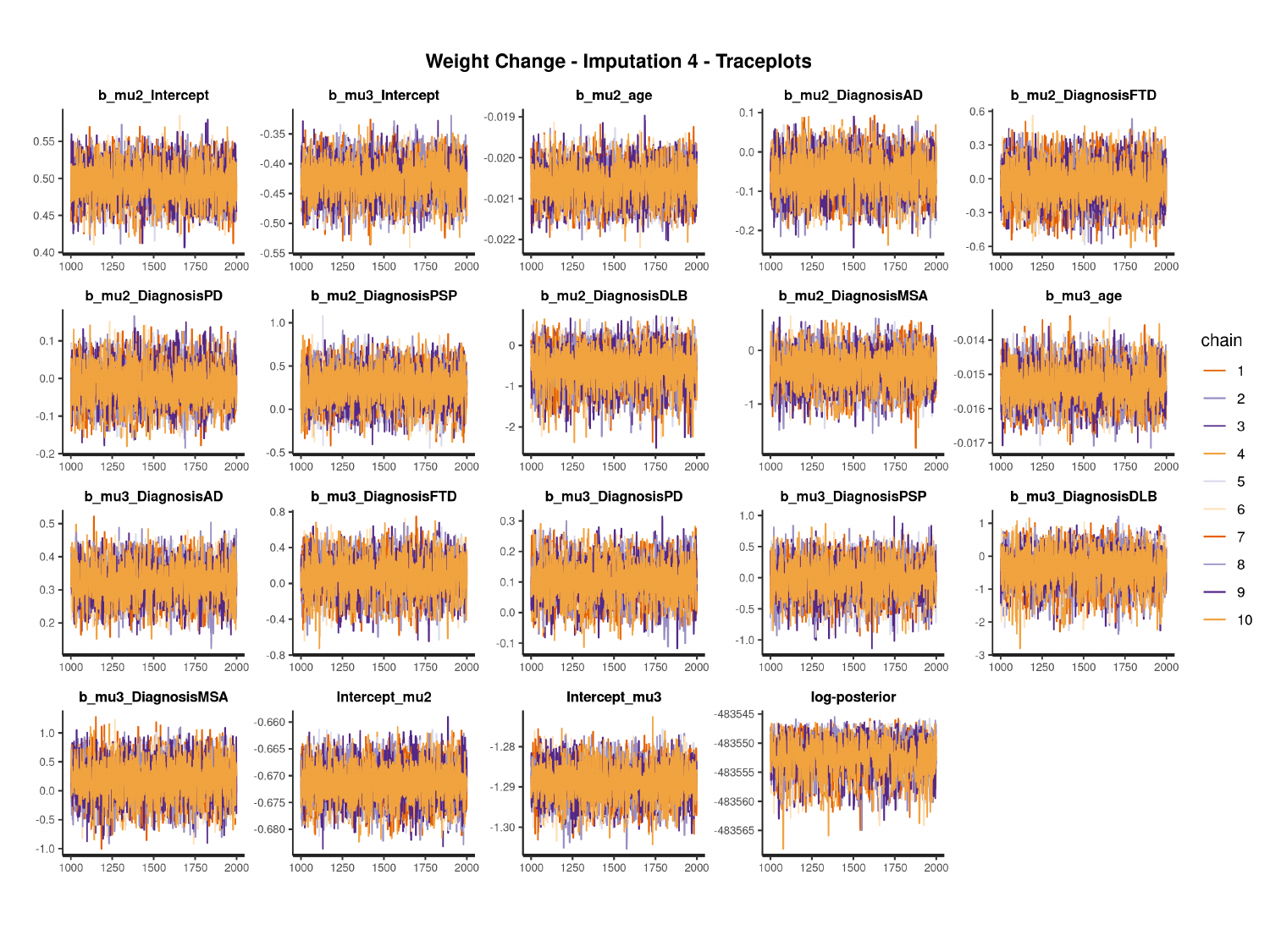

#

### Slide 166
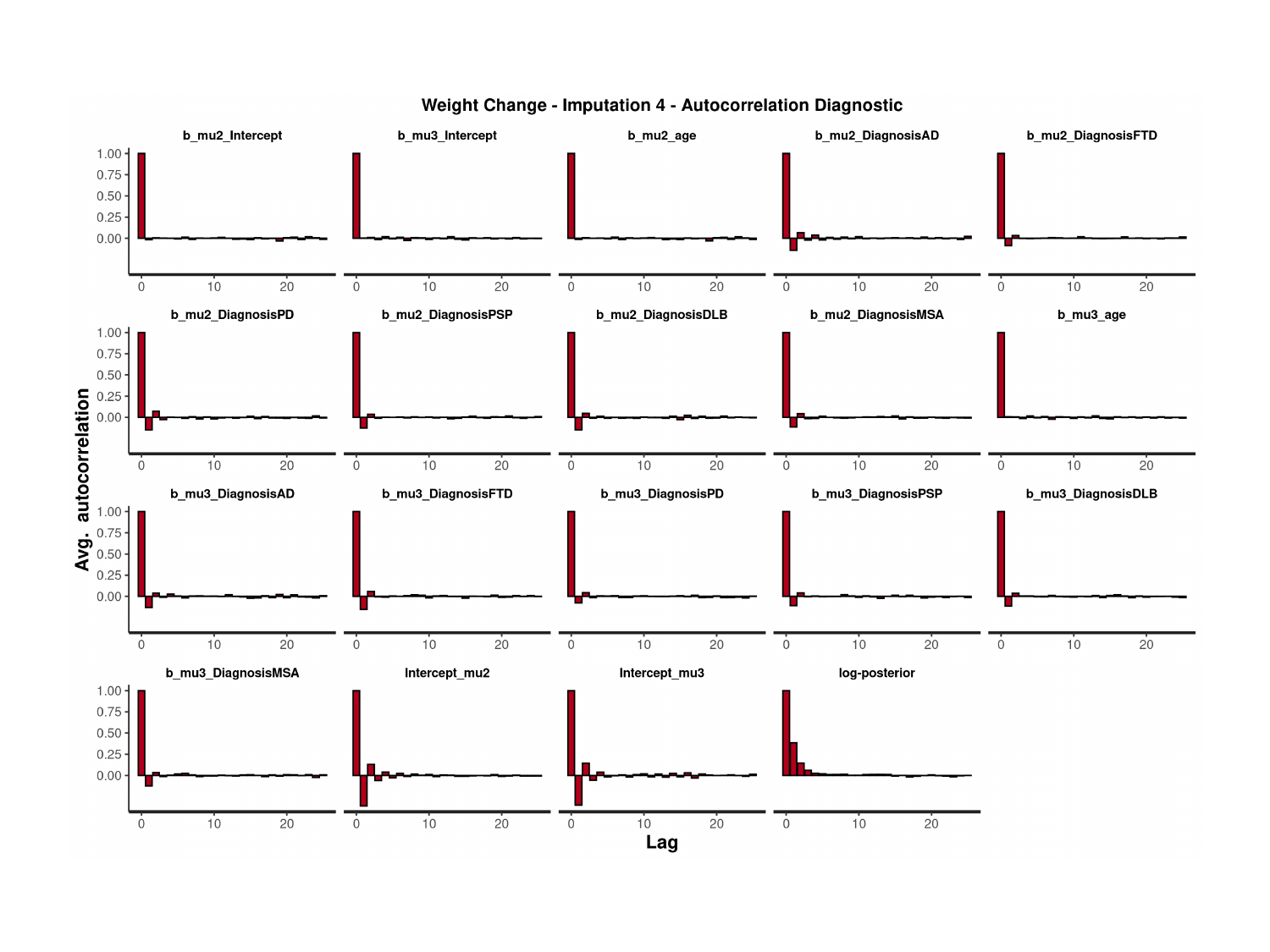

### Slide 167
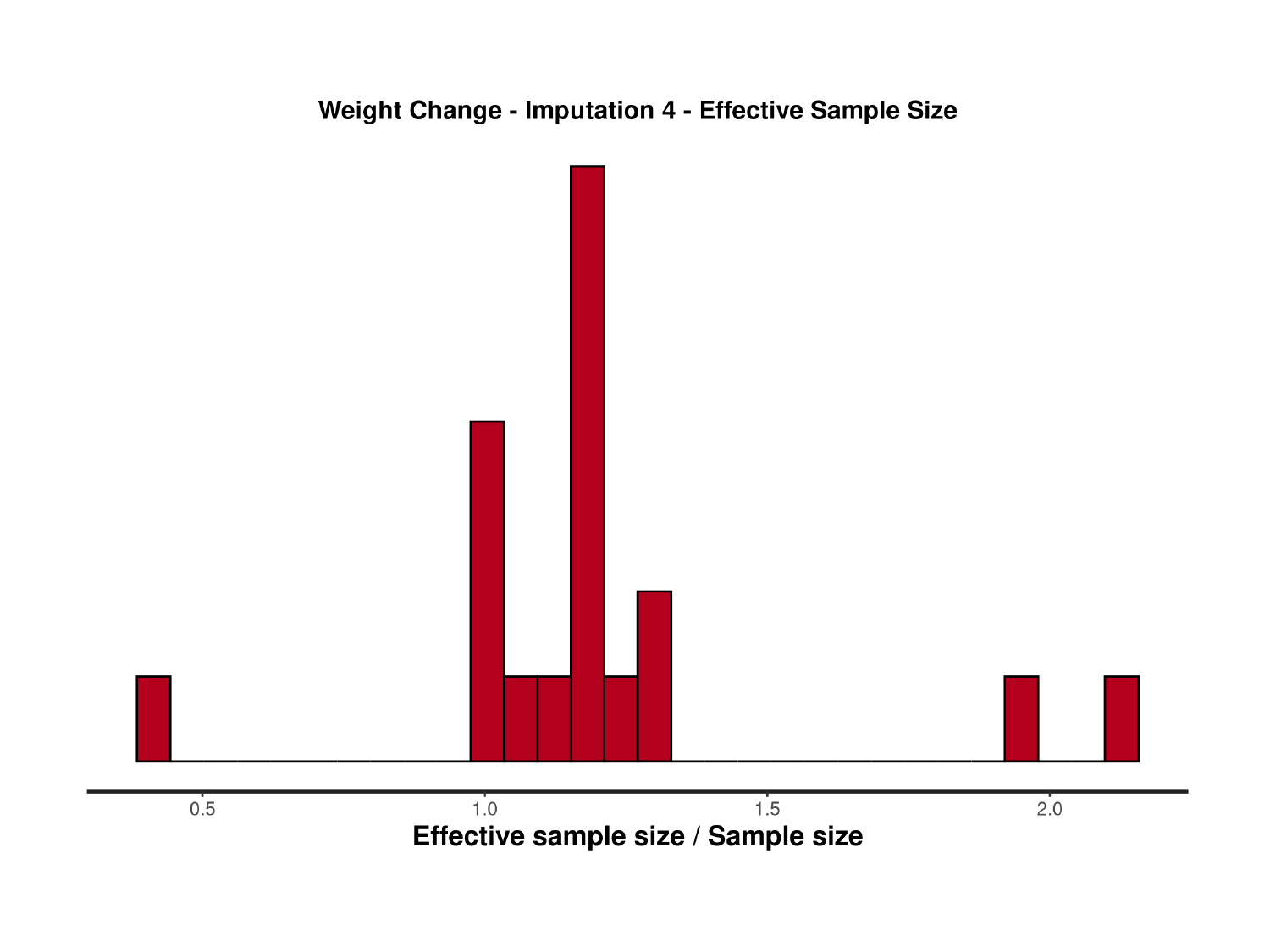

### Slide 168
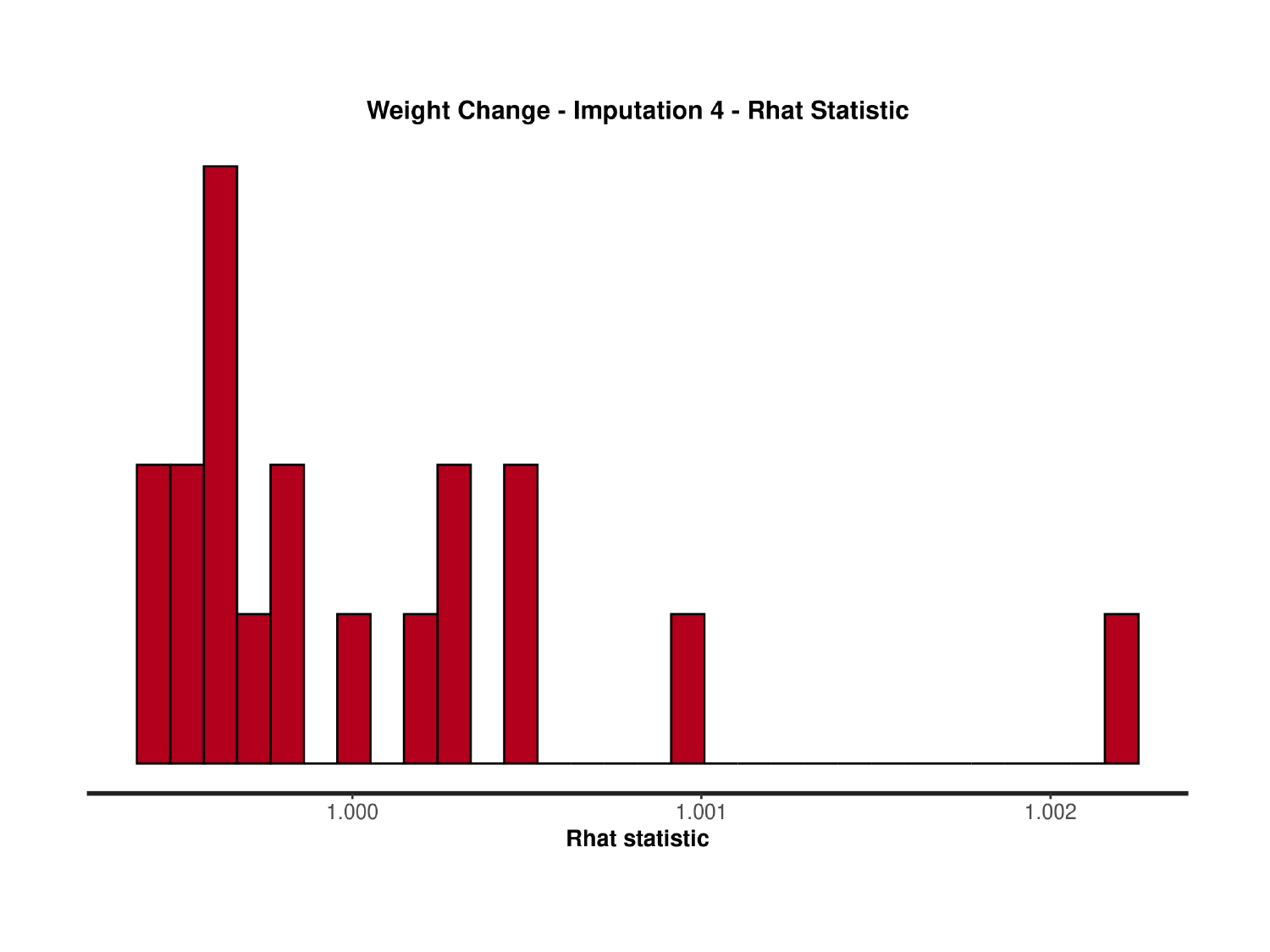

### Slide 169
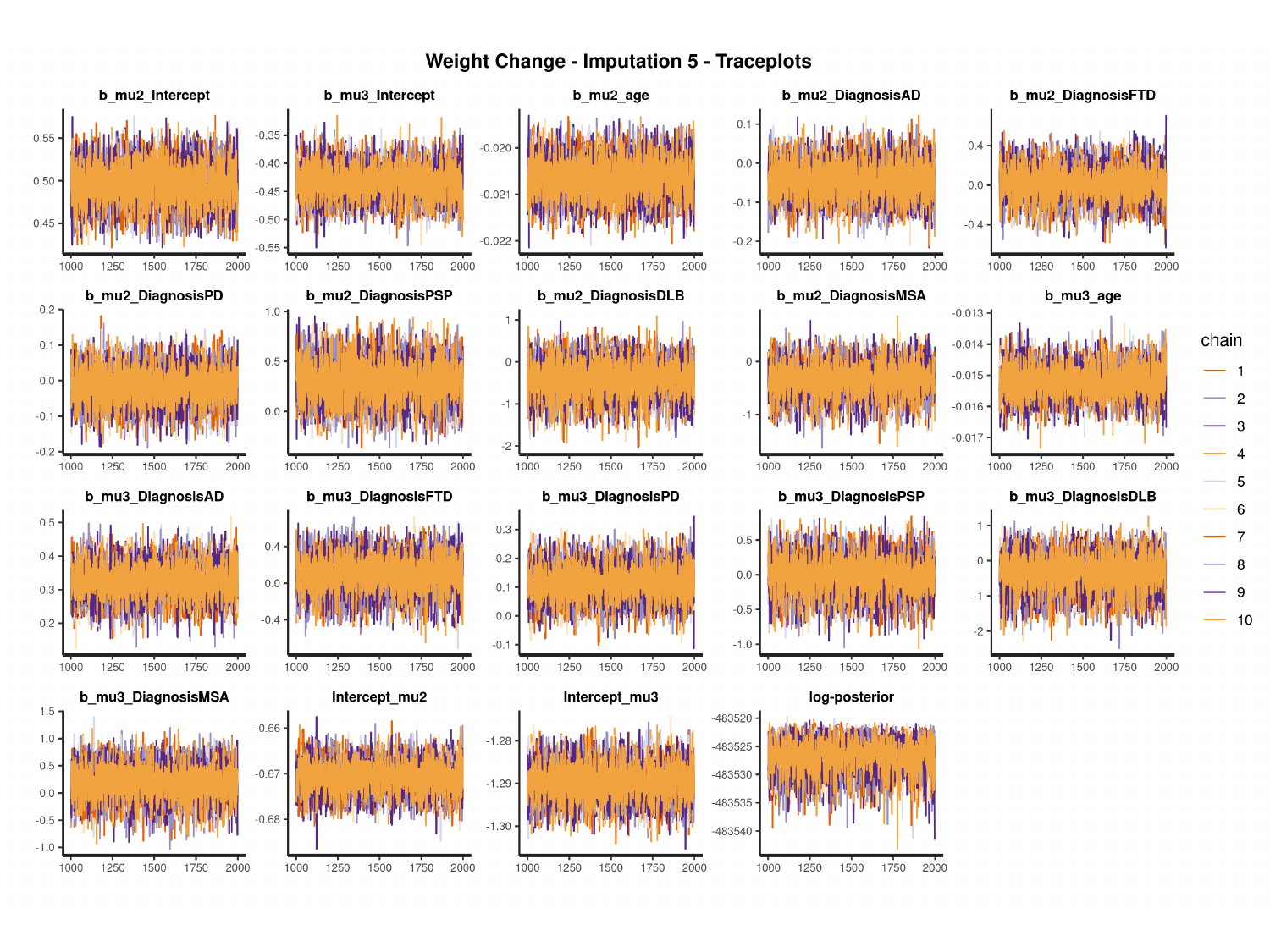

#

### Slide 170
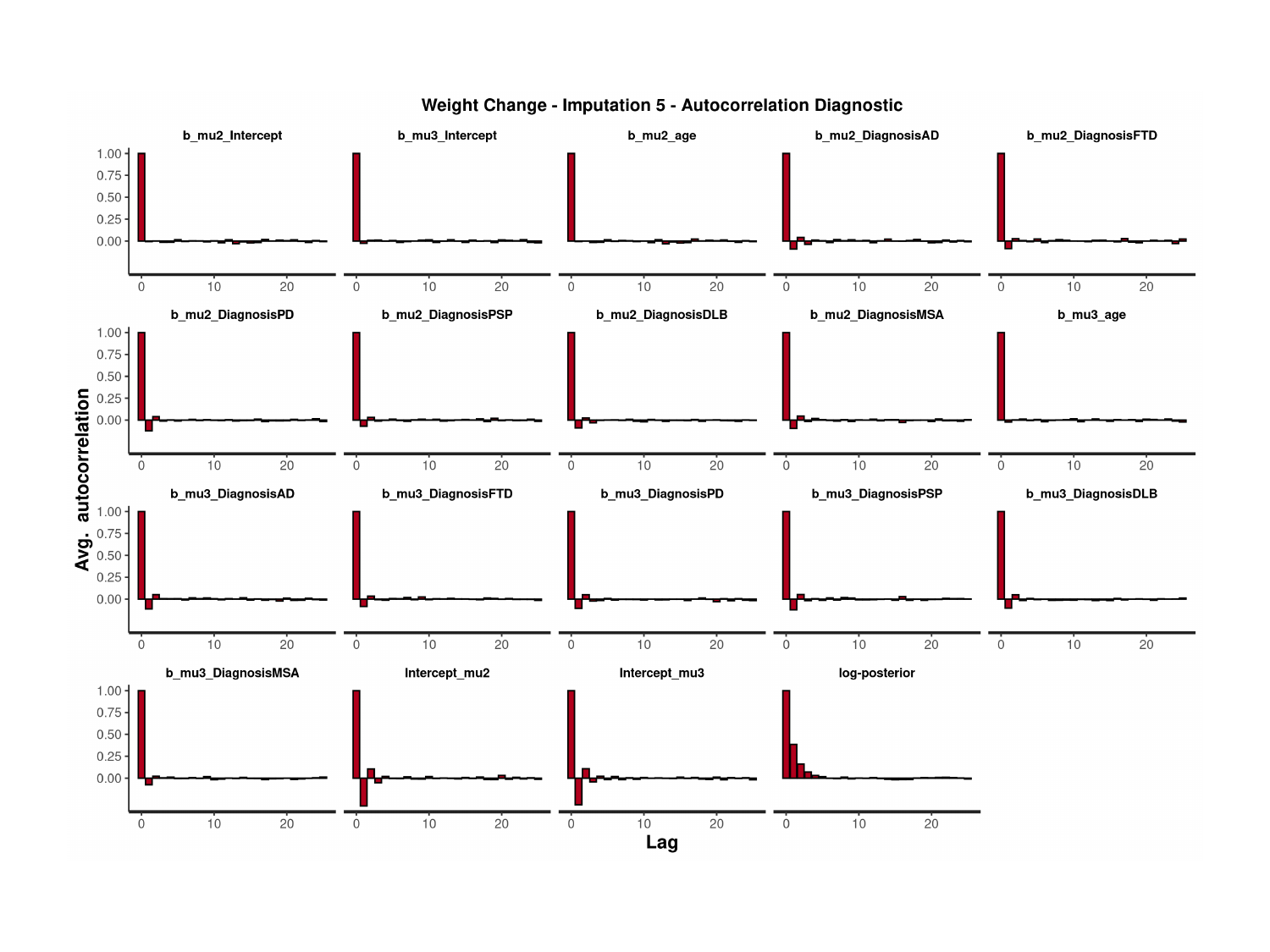

### Slide 171
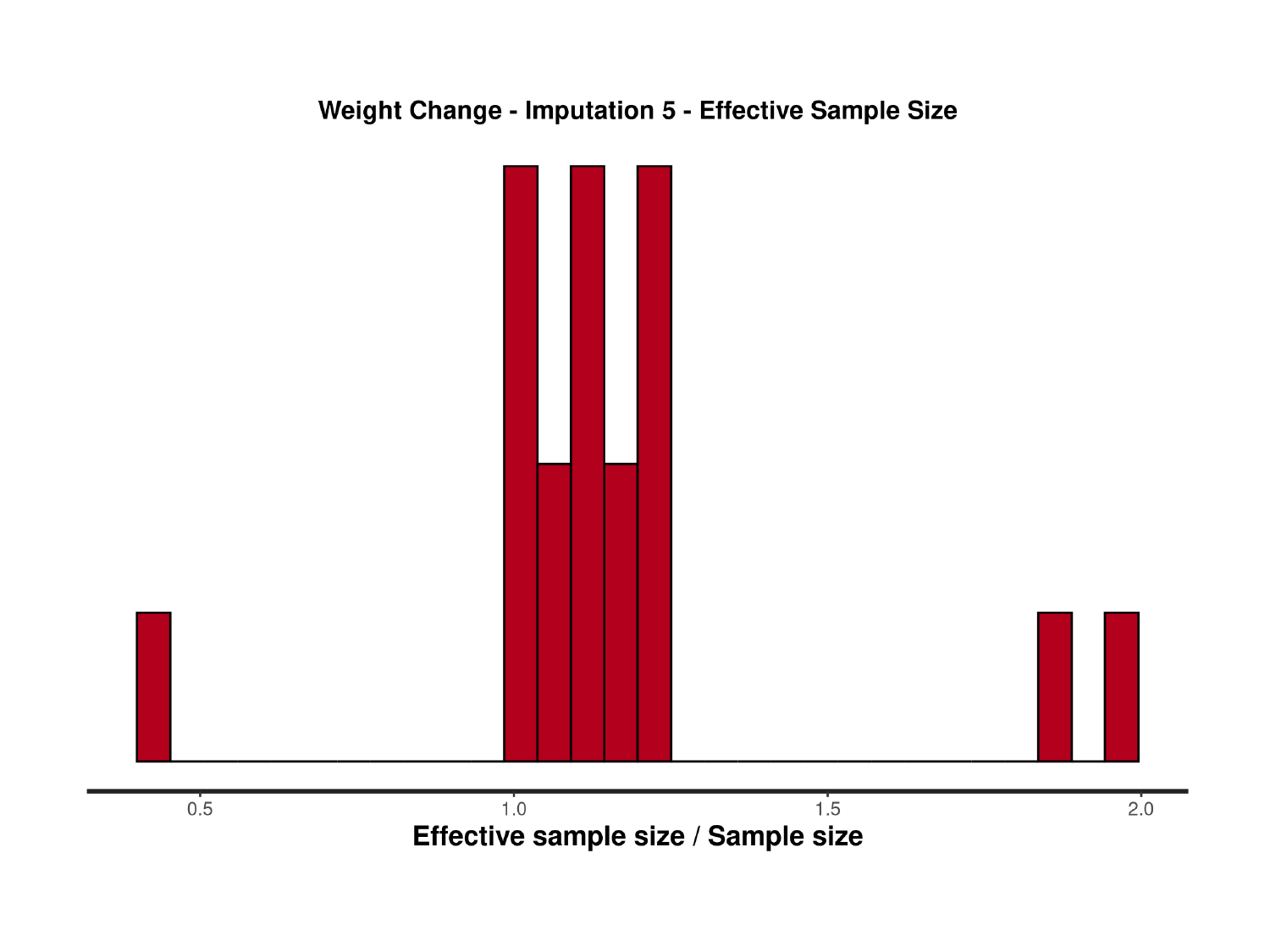

### Slide 172
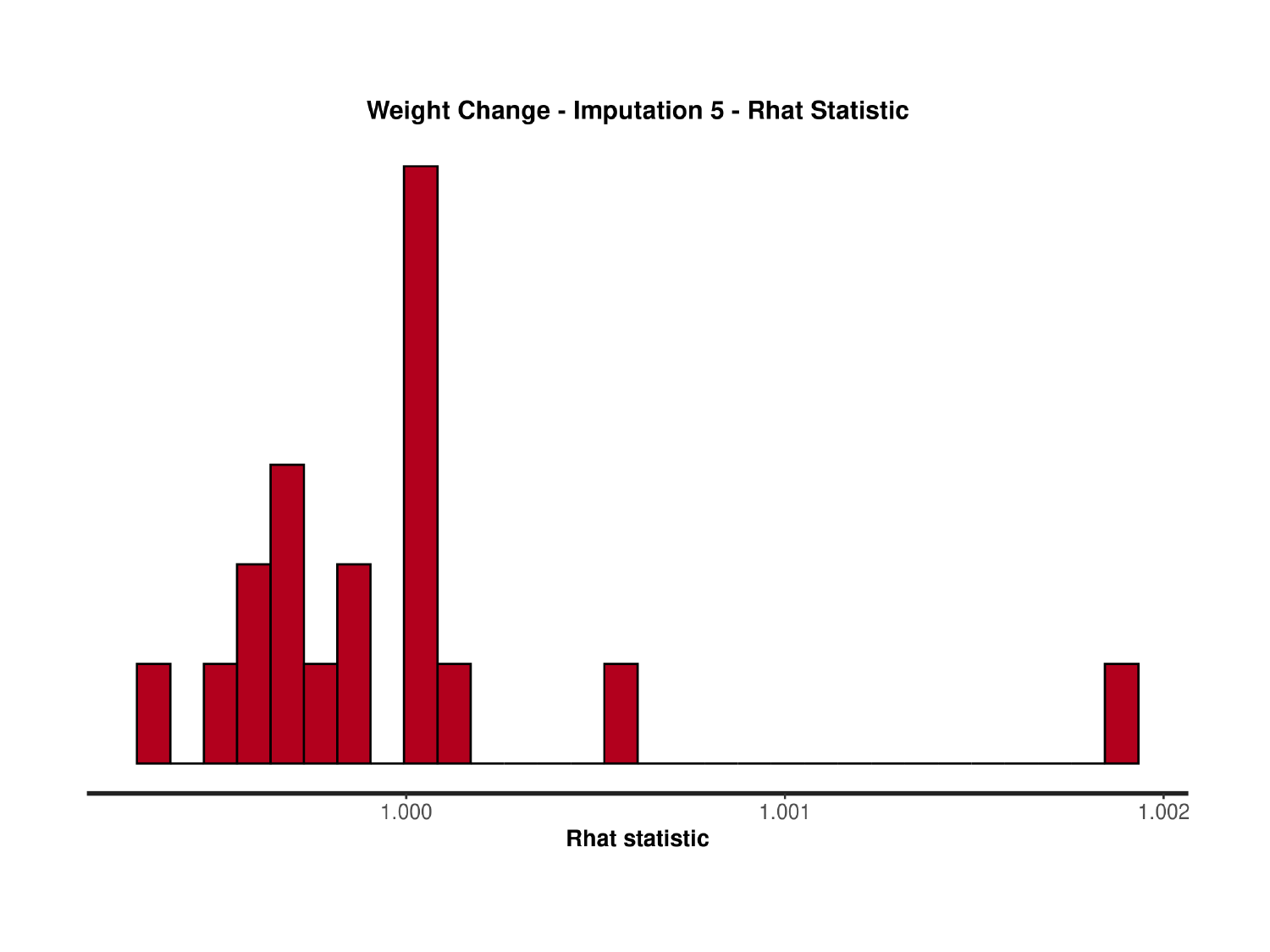
