## Supplementary material for "Pre-Diagnostic Cognitive and Functional Impairment in Multiple Sporadic Neurodegenerative Diseases": Posterior predictive checks for BRMS models

### Slide 1
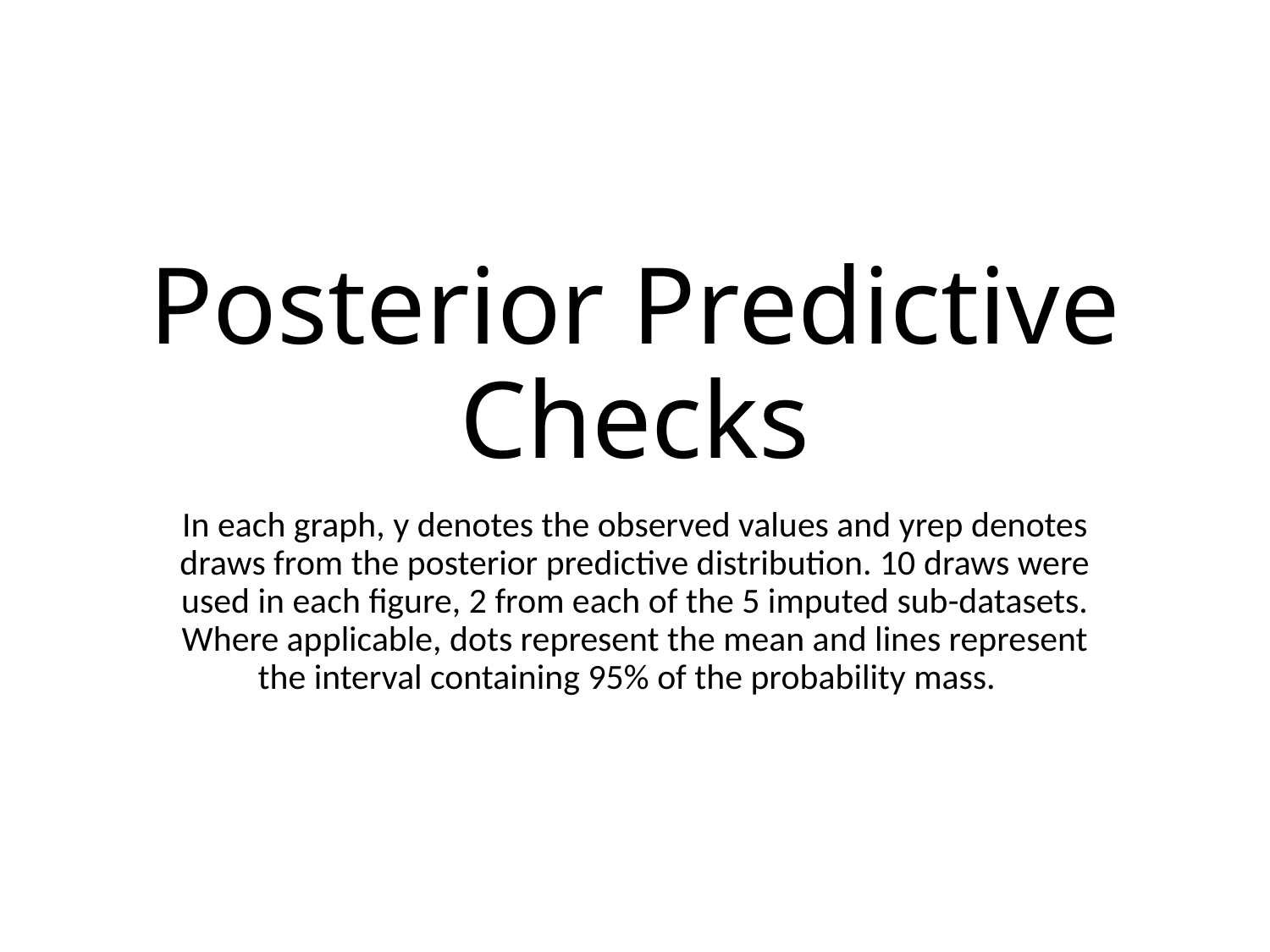

#### Posterior Predictive Checks
In each graph, y denotes the observed values and yrep denotes draws from the posterior predictive distribution. 10 draws were used in each figure, 2 from each of the 5 imputed sub-datasets. Where applicable, dots represent the mean and lines represent the interval containing 95% of the probability mass.

### Slide 2
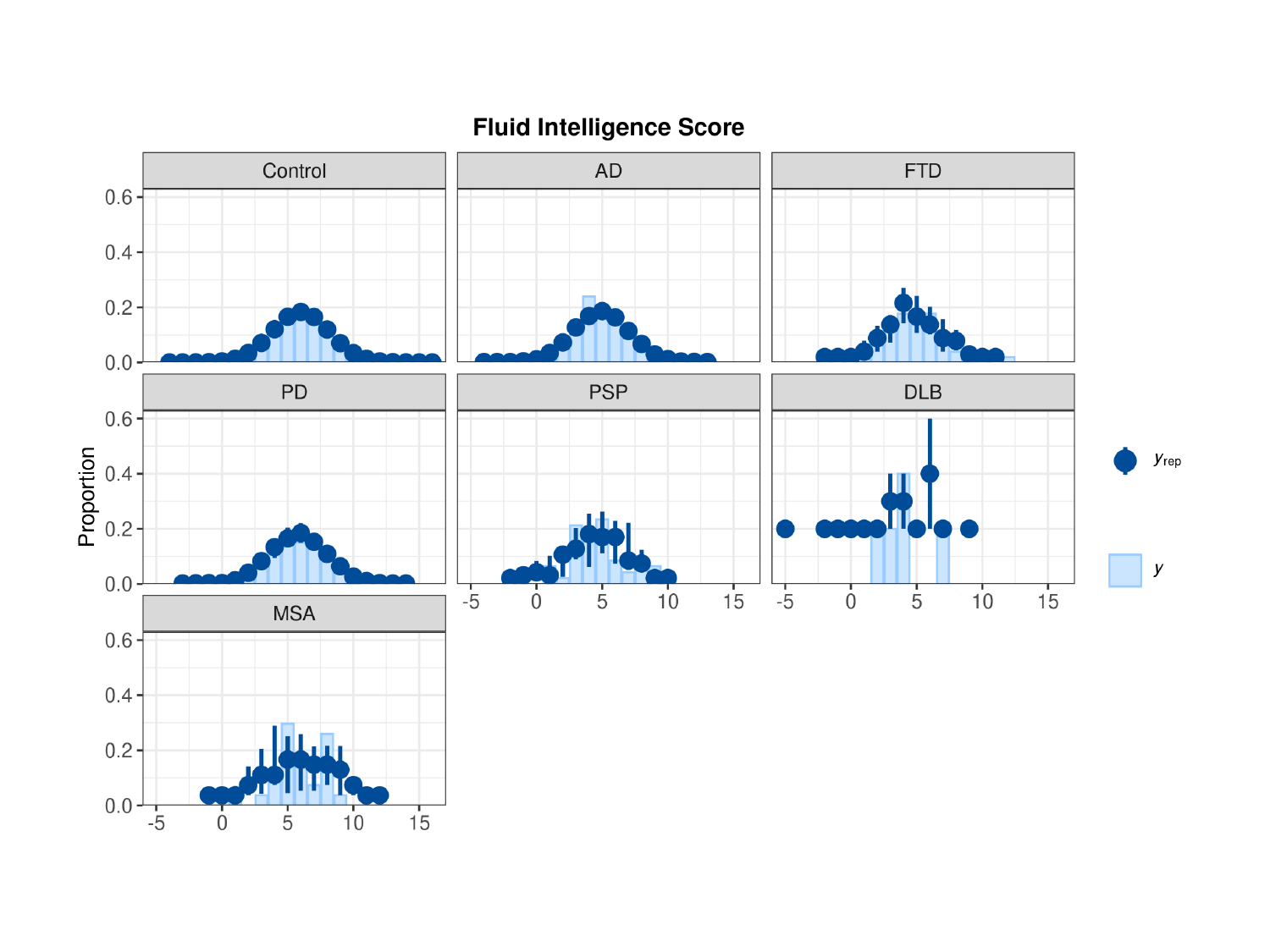

### Slide 3
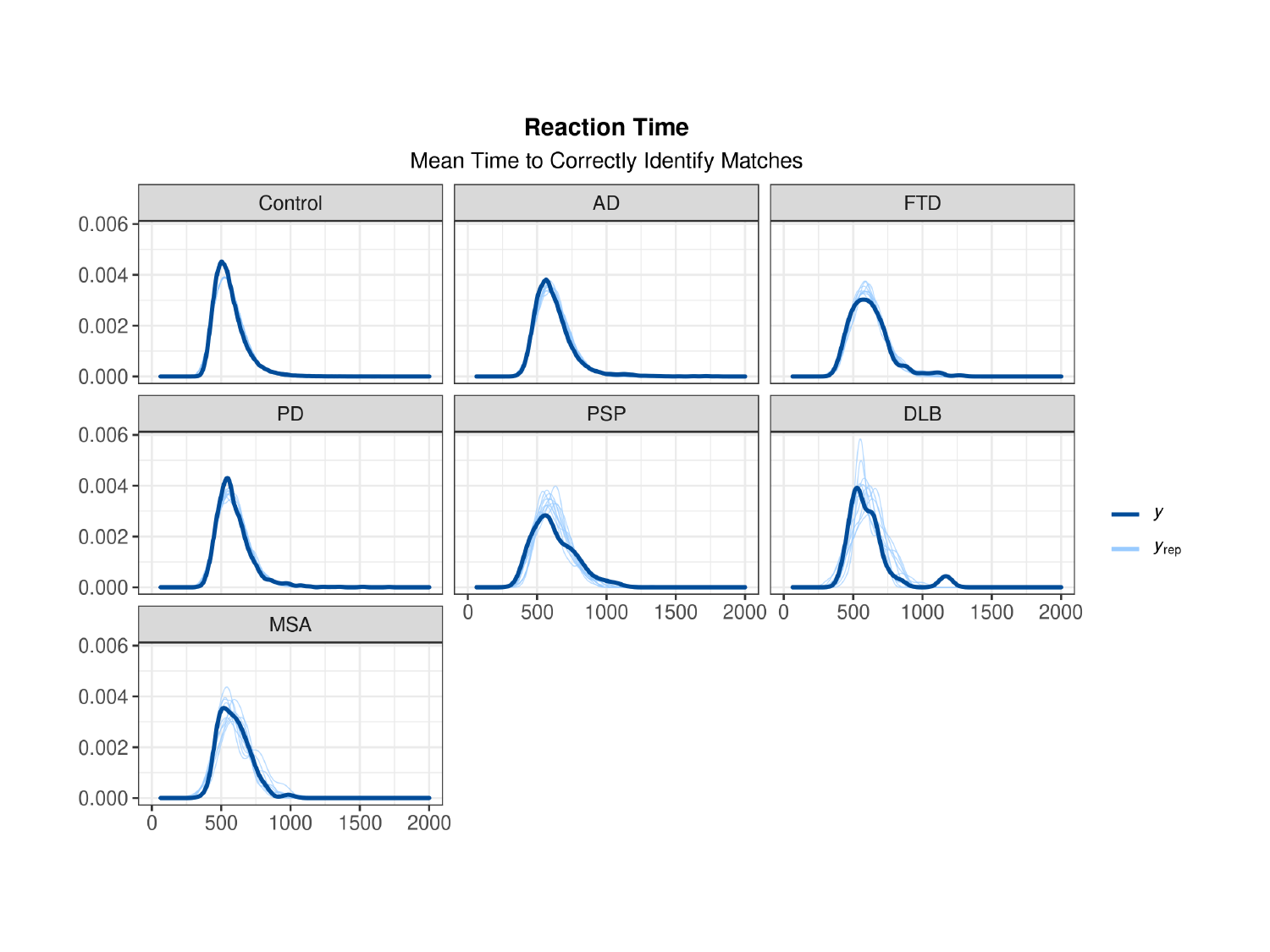

### Slide 4
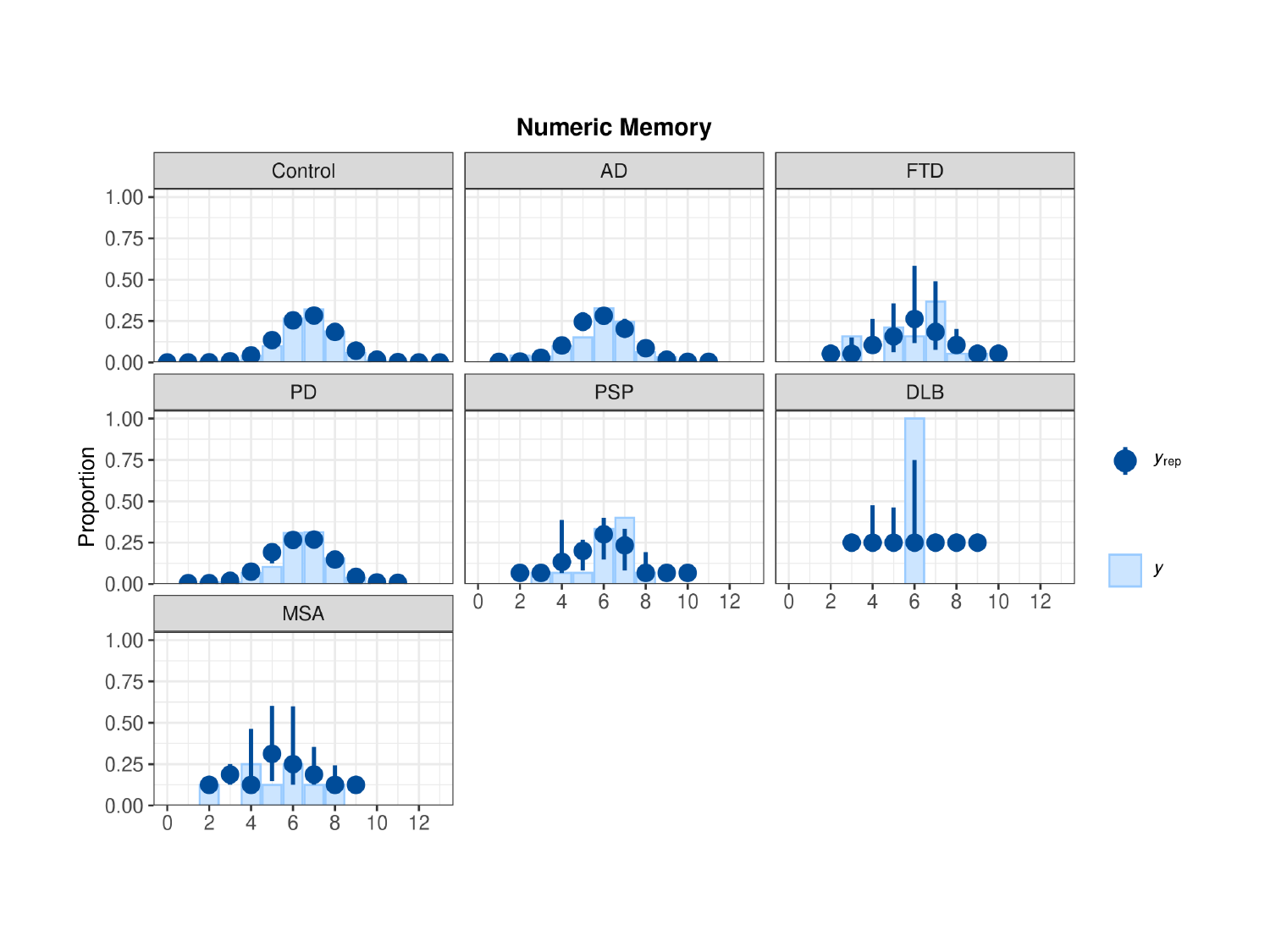

### Slide 5
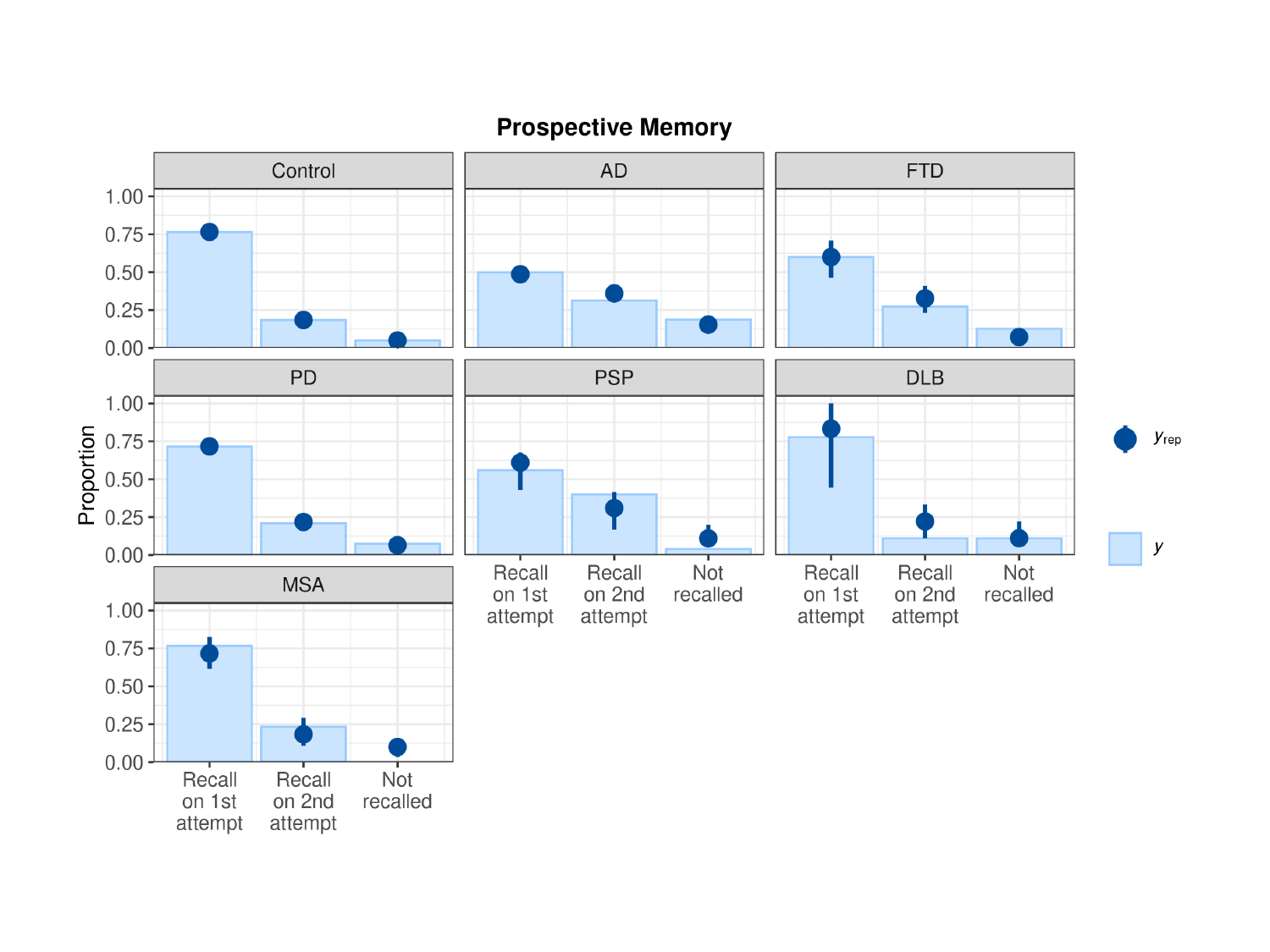

### Slide 6
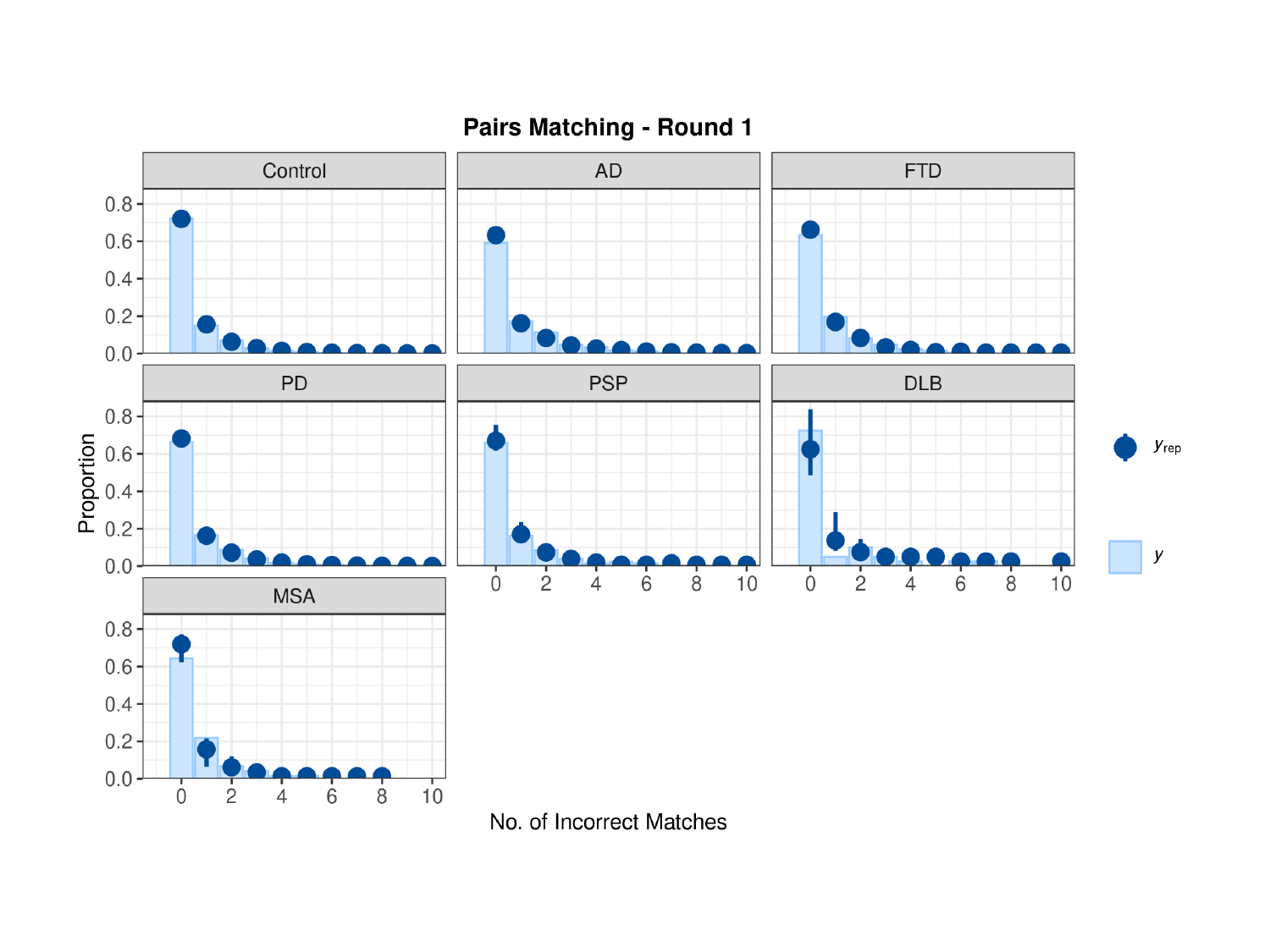

### Slide 7
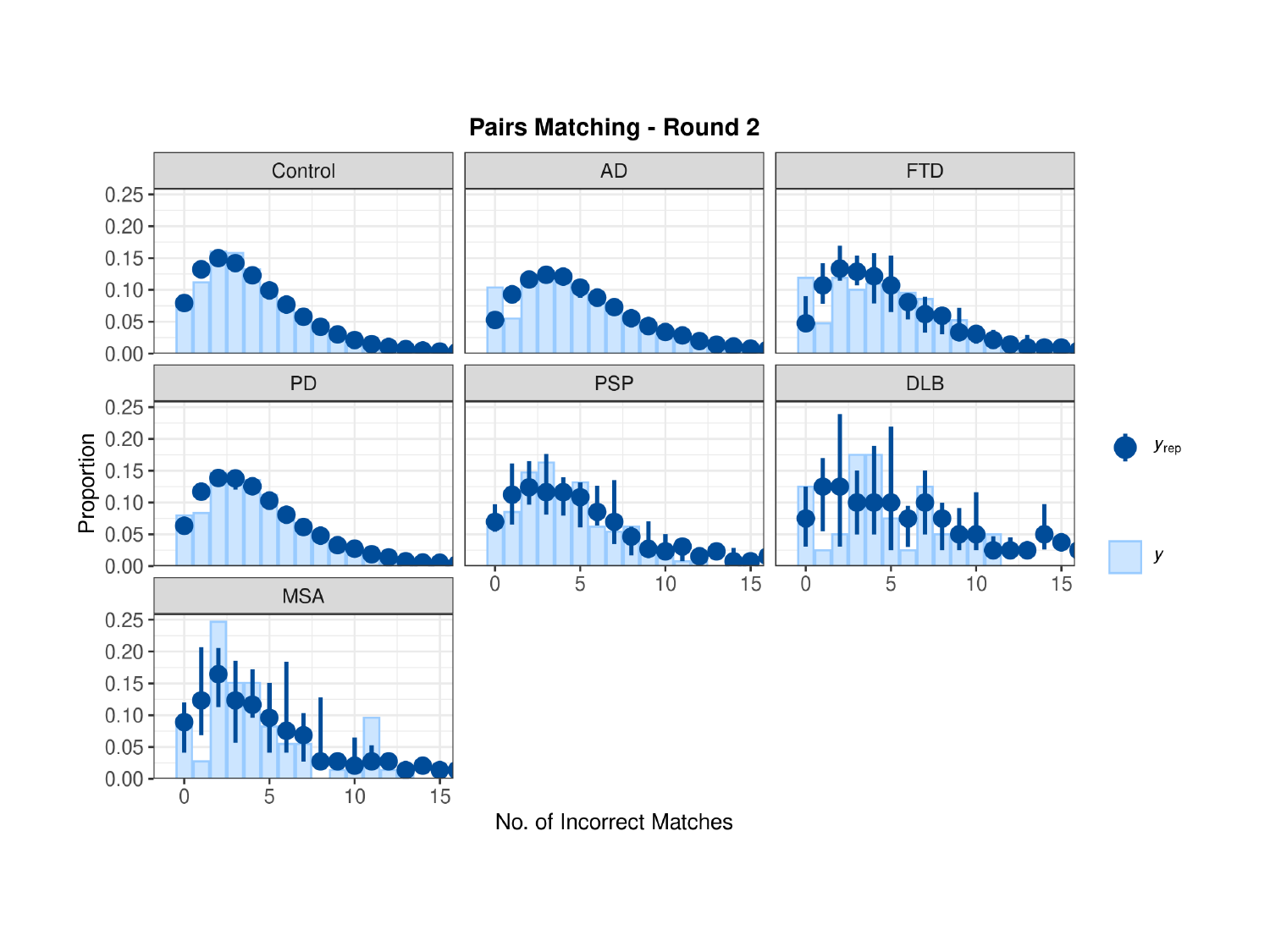

### Slide 8
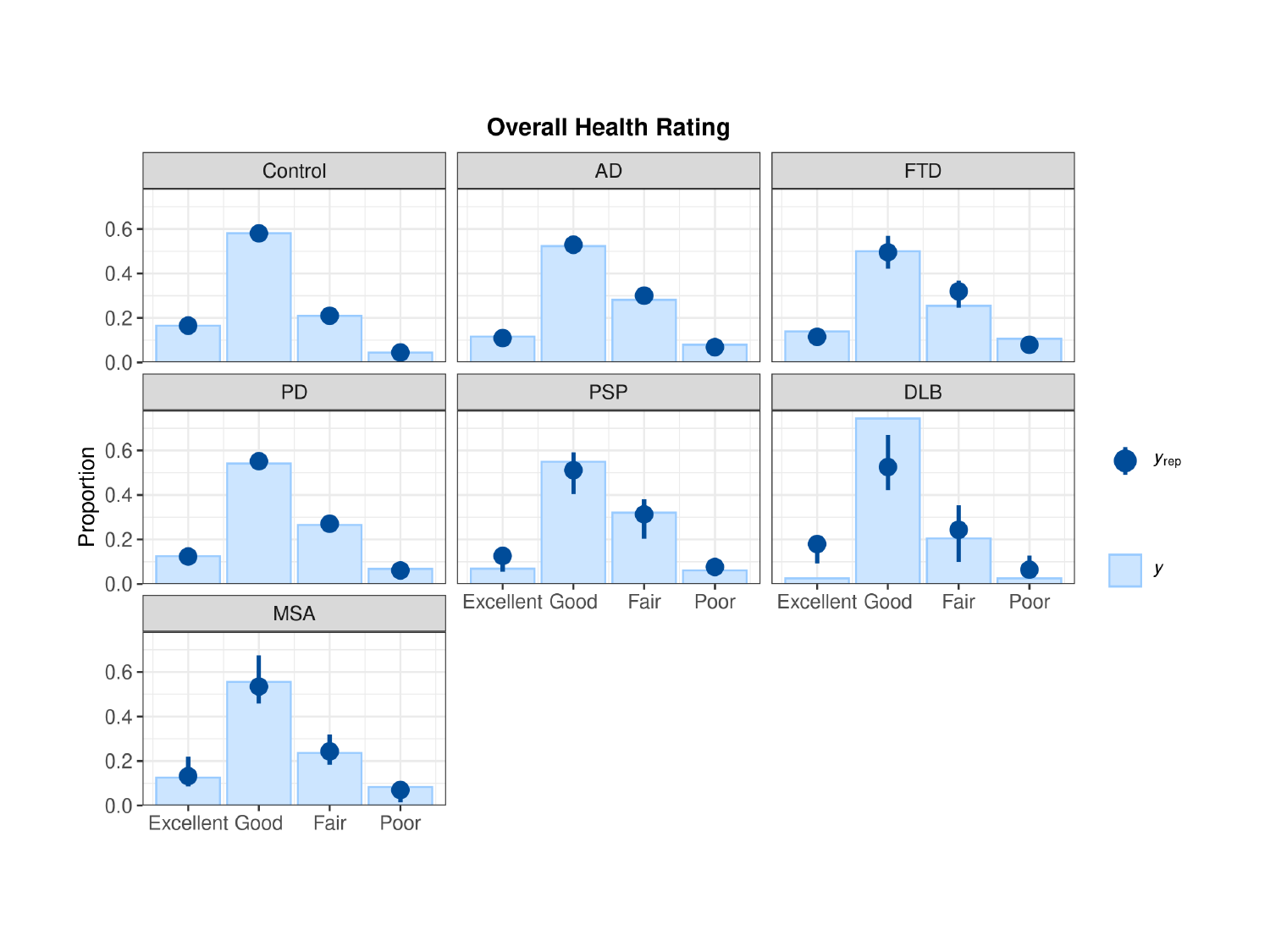

### Slide 9
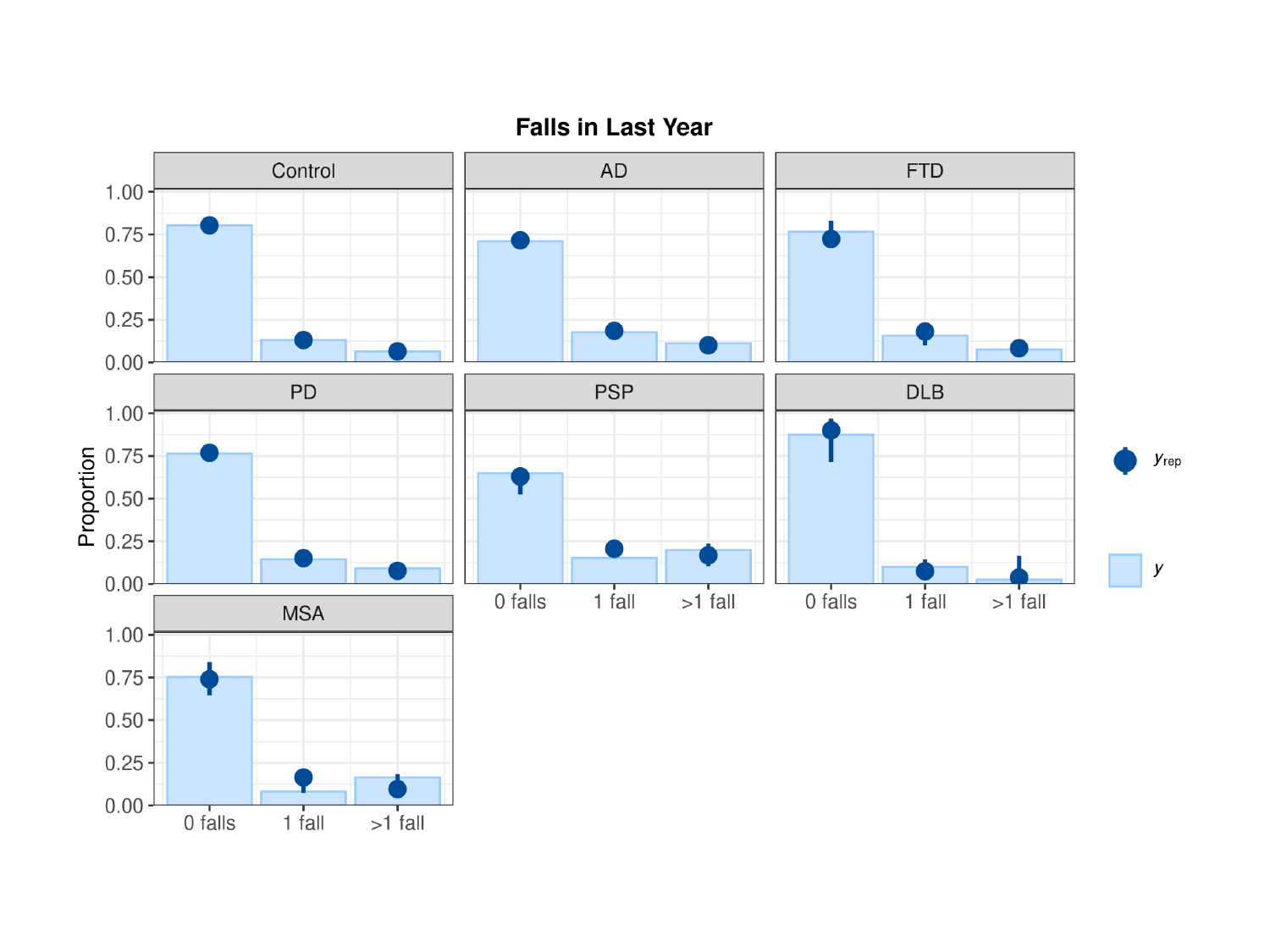

### Slide 10
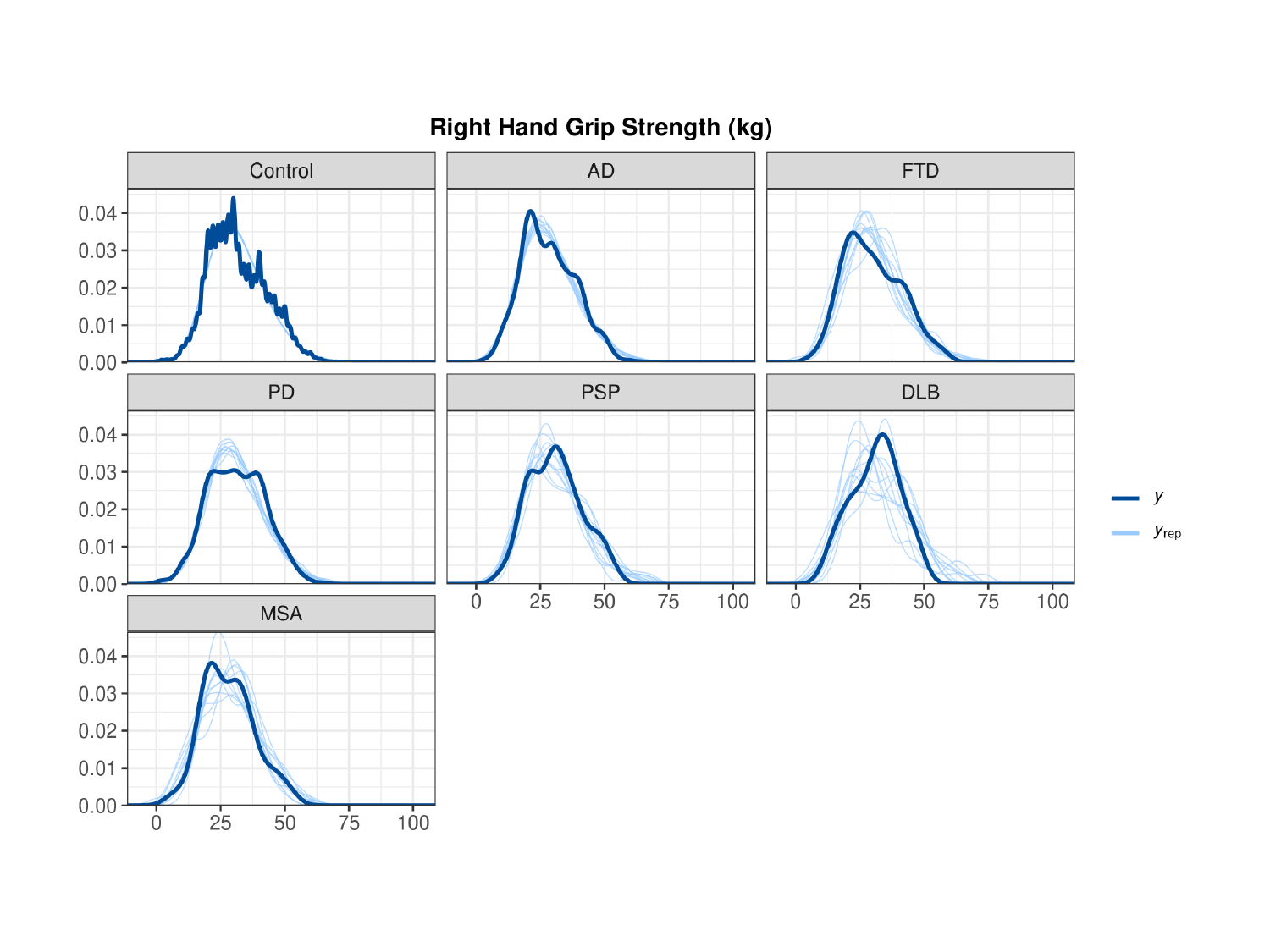

### Slide 11
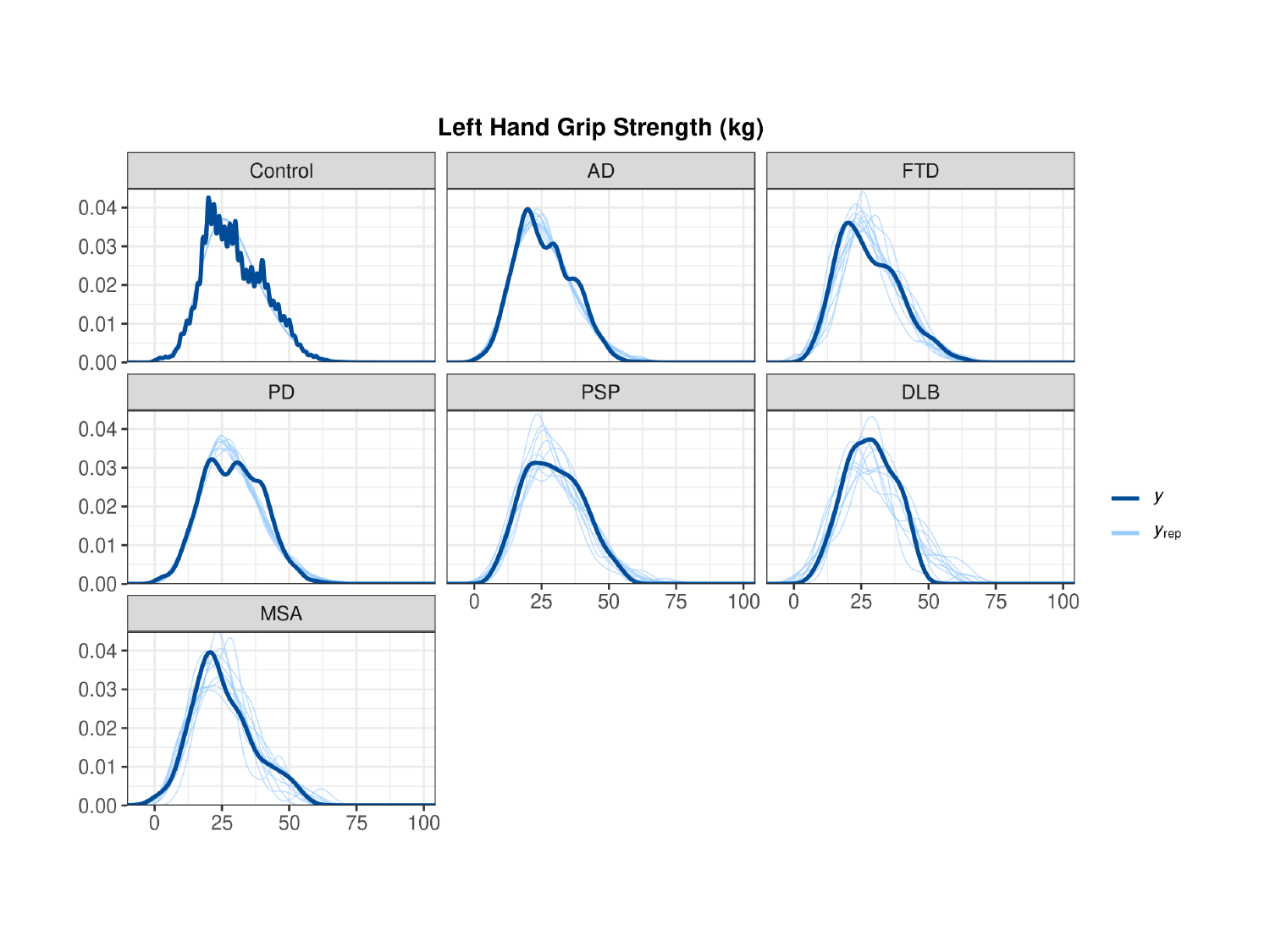

### Slide 12
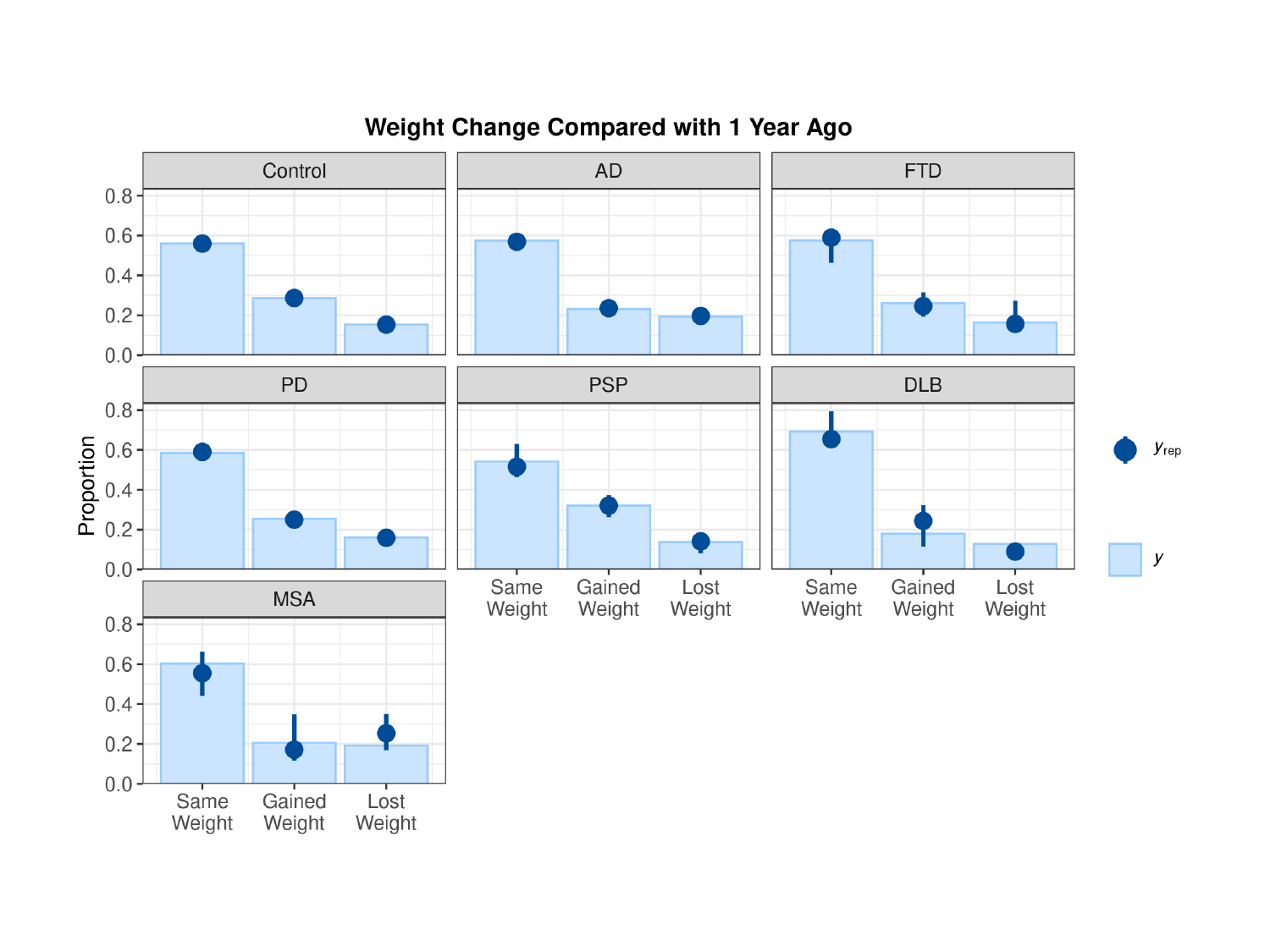
